# Compounding of rare pathogenic copy-number variants and polygenic background is consistent with assortative mating

**DOI:** 10.1101/2025.09.08.25335316

**Authors:** Caterina Cevallos, Chiara Auwerx, Robin Hofmeister, Théo Cavinato, Tabea Schoeler, Zoltán Kutalik, Alexandre Reymond

## Abstract

Copy-number variants (CNVs) are linked to a spectrum of outcomes and carriers of the same variant exhibit variable disease severity. We explored the impact of an individual’s polygenic score (PGS) on explaining these differences, focusing on 119 established CNV-trait associations involving 43 clinically-relevant phenotypes. We called CNVs among white British UK Biobank participants, then divided samples into a training set (n = 264,372) to derive independent PGS weights, and a CNV-carrier-enriched test set (n = 96,716). Assessing the individual, joint, and synergistic contribution of CNVs and PGSs, we identified a significant additive effect for 45 (38%) CNV-trait pairs, as well as two scale-dependent interactions between PGSs for gamma-glutamyltransferase levels and grip strength, and a 22q11.23 duplication encompassing multiple key genes in glutathione metabolism. A (spurious) *negative* correlation between an individual’s CNV carrier status and their PGS would be expected under selective participation-induced collider bias. Instead, we observed a widespread *positive* correlation, which could only be partially accounted for by linkage disequilibrium. Given a non-null inheritance rate for all 17 testable CNVs, we explored whether assortative mating could explain this positive CNV-PGS association. We found strong agreement between this correlation and the one predicted by assortment (r = 0.45, p = 2.0 *×* 10^-7^). Similar suggestive trends of positive correlation were observed between PGSs and genome-wide burden of CNVs or rare loss-of-function variants. Our results demonstrate that PGSs contribute to the variable expressivity of CNVs and rare variants, and improve the identification of individuals at higher risk of clinically relevant comorbidities. We also highlight pervasive assortative mating as a likely mechanism contributing to the compounding of genetic effects across mutational classes.

## Introduction

Copy-number variants (CNVs) are characterized by the deletion or duplication of DNA fragments ≥ 50 bp. In the late 2000s, genomic disorders were defined as a class of conditions caused by large recurrent CNVs, which represent among the strongest individual genetic risk factors for intellectual disability and developmental delay, psychiatric disorders, epilepsy, and congenital anomalies [1–6]. Yet, it was recognized early on that carriers of genomic disorder-associated CNVs express varying constellations of clinical features, implying their incomplete penetrance (i.e., not all carriers present all phenotypes linked with the genomic disorder) and variable expressivity (i.e., carriers show different degrees of phenotype severity) [7, 8]. This insight was strengthened by biobank studies showing that these CNVs are present in the general population – albeit at a lower frequency – and they associate with a broad range of traits [9–27], often paralleling findings from clinical cohorts – but with a lower penetrance and severity – while also revealing novel associations, showcasing the pleiotropy of these CNVs [28–32].

Phenotypic variability among CNV carriers can be explained through the liability threshold model, according to which, for each trait, an individual carries a liability that follows a normal distribution [33]. If that liability exceeds a threshold, the individual is considered a case. Provided that the impact of a CNV on disease liability is large enough to surpass this threshold, each carrier will be affected, and the CNV will be considered to be fully penetrant. Alternatively, the CNV might contribute to disease liability, but contributions of additional genetic, demographic, or environmental risk factors are required for the carrier to be affected, leading to incomplete penetrance and variable expressivity. Importantly, the impact of both the CNV and additional risk factors should be considered in a traitspecific manner, explaining phenotypic heterogeneity across carriers [32].

Early evidence of the contribution of additional genetic factors to disease liability stemmed from family-based studies [34–39]. Yet, often, detailed familial information is not available, making it is almost impossible to pinpoint the exact genetic factors contributing to increased liability, as the effect of rare highimpact variants, and the cumulative (but individually mild) effect of a large number of common variants captured by polygenic scores (PGSs) are indistinguishable [40]. The role of rare highimpact variants has been described through the two-hit model, wherein the CNV sensitizes the genome, but its expressivity is determined by additional rare variants [41]. For instance, clinically ascertained carriers of a genomic disorder-associated CNV experience more severe and diverse phenotypic consequences if they also carry a second large CNV [42] or a high burden of damaging singlenucleotide variants (SNVs) [43]. More recently, this model was extended to assess the contribution of the polygenic background to the expressivity of rare CNVs by demonstrating their additive contribution [27, 44–56]. For example, carriers of the schizophrenia-associated 22q11.2 deletion are more likely to be diagnosed with schizophrenia if they have a high PGS for the disease [51, 57], while carriers of the body mass index (BMI)-lowering 16p11.2 BP4-BP5 duplication are less likely to be underweight if they carry a high PGS for BMI [49]. Some of these studies further explored the interplay between CNVs and PGS by testing for interaction between the mutational classes or determining whether carriers of risk-increasing CNVs have a different PGS distribution than noncarriers, yielding variable conclusions. Trends for lower disease PGS among cases carrying disease-associated CNVs have been reported, and although a large study identified a significant interaction between schizophrenia PGS and schizophrenia-associated CNVs, others failed to robustly replicate CNV-PGS interactions [27, 44, 45, 47, 49, 50, 54]. Importantly, these studies primarily rely on case-control study designs for neuropsychiatric conditions.

Here, we sought to better understand the contribution of common SNVs to the expressivity of rare CNVs across a broad range of quantitative complex traits in the UK Biobank (UKBB). Starting from 119 CNV-trait associations involving 43 distinct phenotypes and 27 CNV regions [19], we first assessed how these associations are modified by an individual’s PGS. Next, to gain mechanistic insights regarding the co-occurrence of these two mutational classes in an individual, we explored three distinct theoretical models of the CNV-PGS relation (Fig. 1). First, participation bias may induce a (spurious) negative CNV-PGS correlation, as the UKBB exhibits a healthy cohort bias [58] (Fig. 1a). As such, carriers of deleterious CNVs who nevertheless take part in UKBB are expected to exhibit a PGS that counteracts the effect of the CNV, reducing expressivity and penetrance through genetic compensation. On the contrary, a positive CNV-PGS correlation is expected if the SNVs included in the PGS are in linkage disequilibrium with the CNV, *de facto* tagging the latter (Fig. 1b). An alternative mechanism explaining a positive CNV-PGS correlation is assortative mating, where the phenotypic similarity caused by one parent’s PGS and the other parent’s CNV results in a more likely coinheritance of the CNV and PGS in the offspring, pushing the phenotype in the same direction (Fig. 1c).

**Figure 1.**
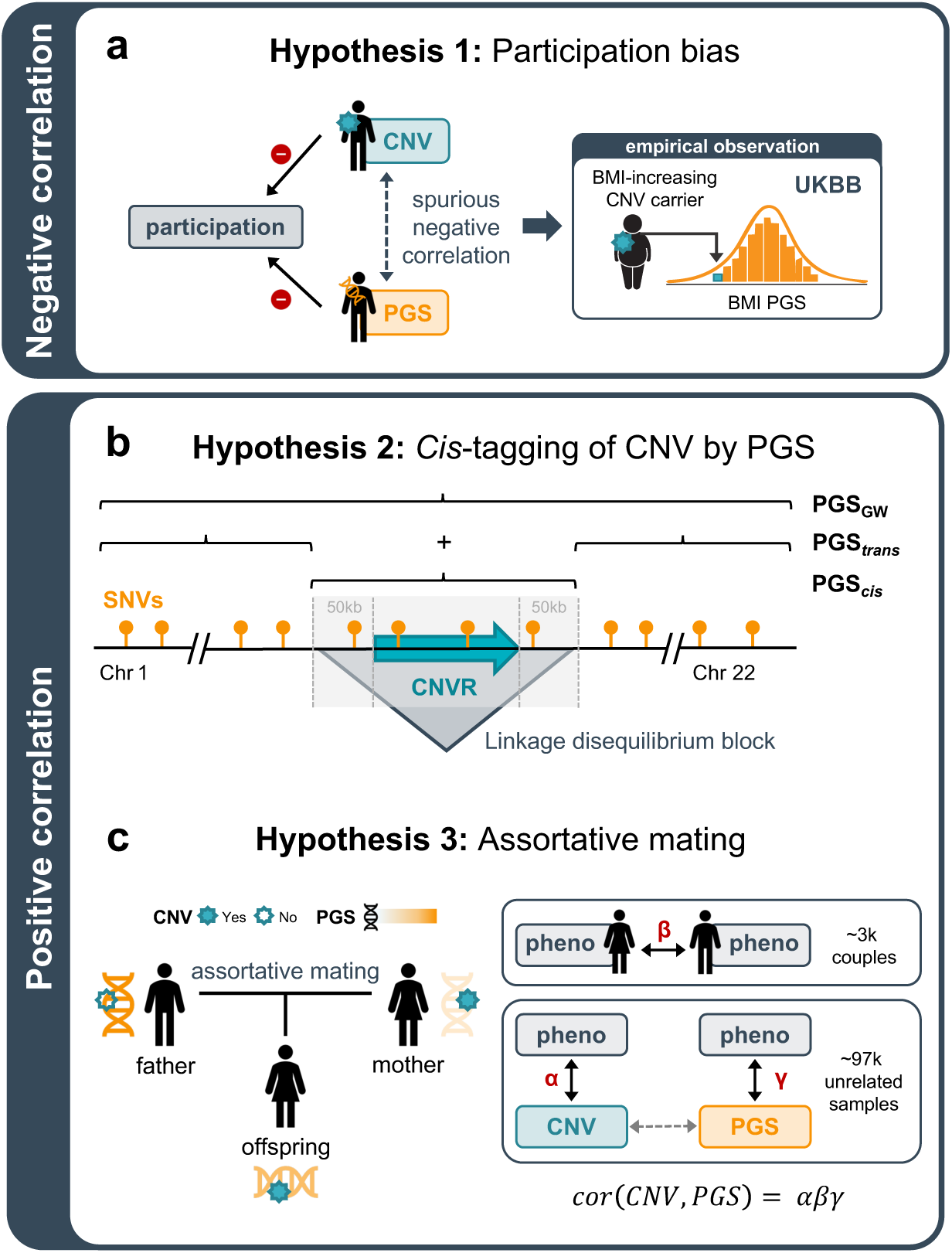
Mechanisms of CNV-PGS correlation. Schematic representation of three possible mechanisms explaining correlation between an individual’s CNV carrier status and polygenic score (PGS). **a,** Negative CNV-PGS correlation is expected in the presence of selective UK Biobank (UKBB) participation. Specifically, if both CNV carrier status and PGS predict participation in the same direction (e.g., decreasing or increasing participation probability), collider bias would result in a spurious negative CNV-PGS correlation among UKBB participants. As UKBB participants tend to be healthier than the general population [58], we expect both carriers of deleterious CNVs and individuals with a PGS that phenocopies the impact of the CNV to be less likely to participate. This leads to the observation of a lower PGS among carriers of a trait-increasing CNV compared to non-carriers, as illustrated here with a body mass index (BMI)-increasing CNV. **b,** A first mechanism explaining positive CNV-PGS correlation is tagging of the CNV by the PGS due to the CNV being in linkage disequilibrium with single-nucleotide variants (SNVs) included in the genome-wide PGS (PGSGW). This hypothesis can be explored by subseting PGSGW into PGS*cis*, which encompasses SNVs mapping to a padded CNV region (e.g., *±* 50 kb), and PGS*trans*, which encompasses all SNVs that do not map to that region. **c,** An alternative mechanism for positive CNV-PGS correlation is assortative mating, where two individuals exhibiting similar phenotypes due to different underlying genetic etiologies, e.g., a high PGS in the father and a trait-increasing CNV in the mother, produce offspring that carry both genetic risk factors. This hypothesis can be explored by assessing the fraction of empirically observed CNV-PGS correlation that can be explained by an indirect path mediated by assortative mating, derived from multiplying the CNV-trait correlation (*α*), the within-couple phenotypic correlation (*β*), and the PGS-trait correlation (*γ*).

## Results

### Study material overview

To assess the role of genetic background in shaping the expressivity of CNVs, we selected 119 genome-wide significant autosomal CNV-trait associations that we previously identified in the UKBB [19] (Supplementary Table 1). The 119 signals involved 43 quantitative phenotypes describing a broad range of physiological functions, including musculoskeletal (n = 5), metabolic (n = 11), neuropsychiatric (n = 2), cardiopulmonary (n = 4), hepatic, (n = 7), hematologic (n = 8), endocrine (n = 2), and renal (n = 4) measurements (Supplementary Table 2). Each signal involves a lead CNV-proxy probe whose copynumber most significantly associated with the trait through either of three association models: a mirror model, in which the deletion and duplication impact the phenotype in an opposite direction, and two models considering the effect of the deletion or duplication in isolation. White British UKBB participants carrying a CNV overlapping any of the lead probes were identified and CNVs were encoded according to the previously determined best association model [19]. We then divided samples into a training set (n = 264,372) that we used to derive genome-wide PGS (PGS_GW_) weights, and a CNV-carrier enriched test set of unrelated individuals (n = 96,716) for which PGS_GW_ were computed (see Methods; Supplementary Fig.1). The incremental phenotypic variance explained by PGS_GW_ compared to a baseline model (Δ*R*^2^) ranged between 2.3-24.6% (median = 11.6% [IQR = 7.9%]). Four traits had an Δ*R*^2^ over 20% – total bilirubin (24.6%), mean corpuscular hemoglobin (22.5%), platelet count (23.0%), and alkaline phosphatase (21.3%) – and four traits had a Δ*R*^2^ under 5% – waist-to-hip ratio (WHR; 4.0%), WHR adjusted for BMI (3.9%), neuroticism (2.9%), and grip strength (2.1%) (Supplementary Fig.2; Supplementary Table 2).

### PGS_GW_ and CNVs additively contribute to phenotypic variability

We jointly modeled the phenotypic impact of CNVs and PGS_GW_, finding that for all 119 CNV-trait pairs, both CNV and PGS_GW_ significantly contributed to phenotypic variability (p ≤ 0.05/119 = 4.2 × 10^-4^; Supplementary Table 3). We then estimated the effect of PGS_GW_ among carriers of trait-associated CNVs, detecting 45 (38%) Bonferroni-significant effects and an additional 30 nominally significant effects, distributed across all eight trait categories (Supplementary Fig. 3; Supplementary Table 4). The likelihood of detecting a significant PGS_GW_ effect strongly depended on the number of carriers assessed (p = 1.1 × 10^-6^) and the predictive performance of the PGS_GW_ (p = 1.1 × 10^-4^). As such, a single CNV-trait pair with less than 100 carriers had a Bonferroni-significant effect of PGS_GW_ on the adjusted phenotype. As 55 (46%) CNV-trait pairs have less than 100 CNV carriers, we expect that larger cohorts of CNV carriers will reinforce evidence of the additive contribution of CNVs and PGSs to phenotypic variability. We next illustrate the clinical relevance of these findings through a series of examples (Fig. 2; Extended Data 1). 16p11.2 BP4-BP5 CNVs exhibit genuine pleiotropy directly affecting multiple physiological systems, with highly heterogeneous phenotypic presentation [31, 32]. We found a nominally significant contribution of PGS_GW_ to the variable expressivity of 14 (58%) out of 24 phenotypes associated to CNVs in the region. Example phenotypes include height (*β*_DEL_ = -8.2 cm; p_DEL_ = 1.0 × 10^-31^; among deletion carriers: *β*_PGS_ = 8.7 cm; p_PGS_ = 1.3 × 10^-10^; Fig. 2a), BMI (*β*_DEL_ = 6.2 kg/m^2^; p_DEL_ = 8.4 × 10^-35^; among deletion carriers: *β*_PGS_ = 4.8 kg/m^2^; p_PGS_ = 0.044; Fig. 2b), or platelet count (*β*_mirror_ = -38.8 10^-9^ cells/L; p_mirror_ = 2.5 × 10^-21^; among CNV carriers *β*_PGS_ = 42.4 × 10^-9^ cells/L; p_PGS_ = 6.7 × 10^-5^; Fig. 2c). Of note, we previously showed that the effect of the CNV on platelet count was independent of the carrier’s BMI and height [31]. Here, we propose that the CNV effect on platelet count might be exacerbated by the carrier’s PGS_GW-platelet count_, positioning the polygenic background as a key determinant of the phenotype constellation expressed by a given 16p11.2 BP4-BP5 CNV carrier.

**Figure 2.**
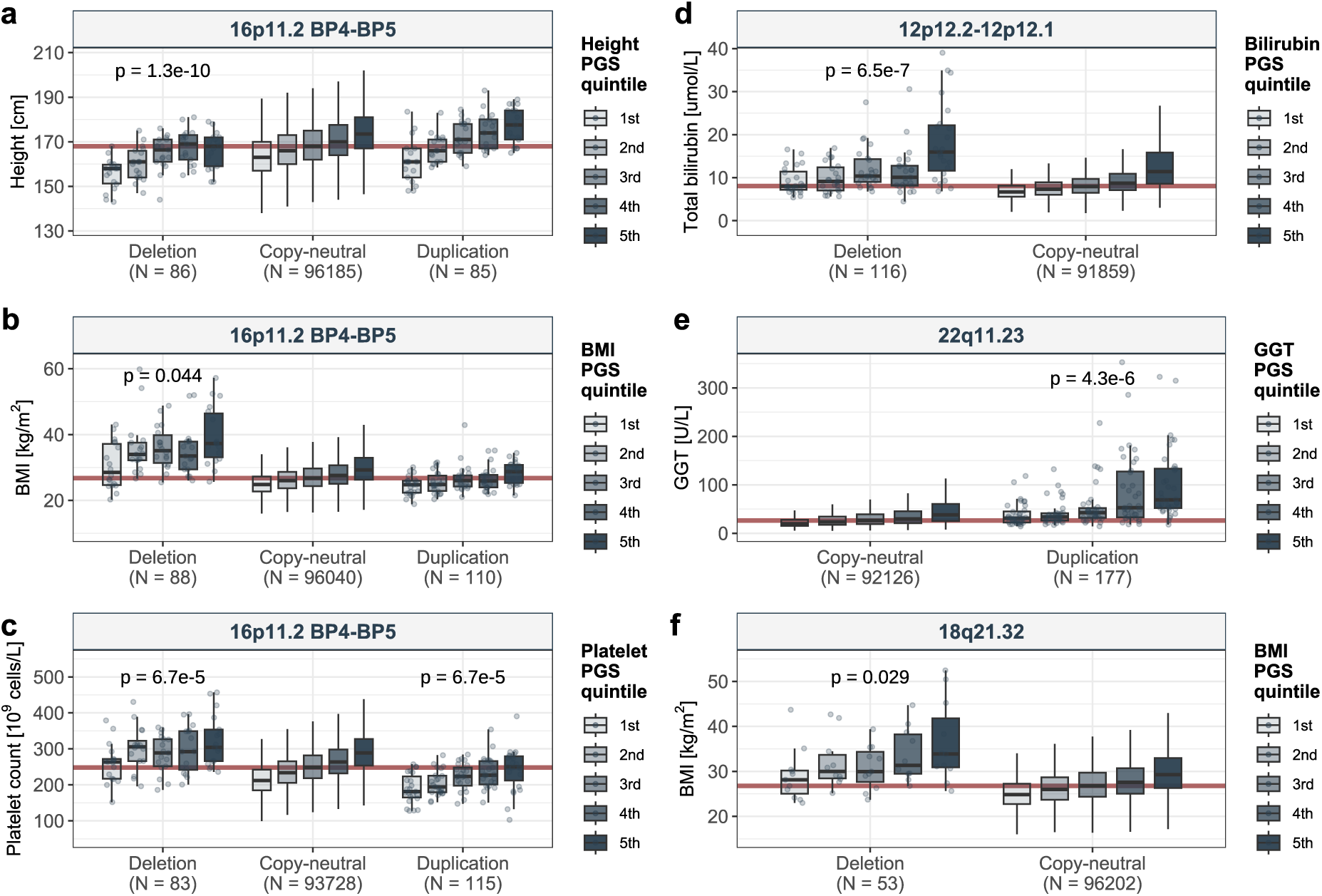
Additive contribution of PGSGW and CNV to phenotypic variability. Boxplots showing the phenotypic distribution (y-axis) of individuals according to their genome-wide polygenic score (PGSGW) quintile and CNV carrier status (x-axis) for **a,** height and 16p11.2 BP4-BP5, **b,** body mass index (BMI) and 16p11.2 BP4-BP5, **c,** platelet count and 16p11.2 BP4-BP5, **d,** total bilirubin levels and 12p12.2-12p12.1, **e,** gammaglutamyl transferase (GGT) and 22q11.23, and **f,** BMI and 18q21.32. Individual data points are shown for CNV carriers as grey dots. The red line represents the median trait value among copy-neutral individuals. Significance of the PGSGW’s effect on phenotypes among carriers of the trait-associated CNV is shown above the assessed CNV genotype groups, whose sample size is indicated as N. Only CNV genotype categories with *≥* 15 individuals are represented.

Laboratory measurements can reach extreme values in individuals carrying CNVs in genes linked to the biomarker’s transport or metabolism, if they concurrently have a high PGS_GW_. This is the case for total bilirubin levels among carriers of a 12p12.2-12p12.1 deletion with a high PGS_GW-bilrubin_ (*β*_DEL_ = 3.0 umol/L; p_DEL_ = 5.9 x 10^-15^; among deletion carriers: *β*_PGS_ = 7.1 umol/L; p_PGS_ = 6.5 x 10^-7^; Fig. 2d). The CNV region encompasses *SLCO1B1* and *SLCO1B3*, two genes encoding hepatic bilirubin transporters [59] whose mutational load has been described to form allelic series for bilirubin levels [18, 19]. Another example relates to gammaglutamyltransferase (GGT) levels among carriers of a 22q11.23 duplication with high PGS_GW-GGT_ (*β*_DUP_ = 34.7 U/L; p_DUP_ = 1.9 x 10^-31^; among duplication carriers: *β*_PGS_ = 62.0 U/L; p_PGS_ = 4.3 x 10^-6^; Fig. 2e). The duplicated region harbors multiple genes involved in glutathione metabolism, including gammaglutamyltransferase 1 (*GGT1*) and 5 (*GGT5*). As such, a substantially higher fraction of duplication carriers presents abnormally elevated (> 50 U/L) GGT levels, compared to copy-neutral individuals (37% vs 17%; p = 8.3 x 10^-15^), with a prevalence of 71% among duplication carriers in the top PGS_GW-GGT_ quintile. These observations indicate that integrating CNV and PGS information could guide the interpretation of clinical test results by identifying individuals with a genetic predisposition to increased biomarker levels.

Finally, despite showing robust trends and strong literature support, some PGS_GW_ effects fail to reach significance, often because of small sample size and/or low PGS_GW_ predictive performance. For instance, PGS_GW-BMI_ nominally contributes to adiposity among the 53 carriers of a BMI-associated 18q21.32 deletion (*β*_DEL_ = 4.2 kg/m^2^; p_DEL_ = 1.6 x 10^-10^; among deletion carriers: *β*_PGS_ = 8.4 kg/m^2^; p_PGS_ = 0.029; Fig. 2f). This intergenic deletion is upstream of *MC4R*, a gene involved in energy homeostasis and appetite regulation, which has been associated with monogenic forms of obesity [60]. Similarly, the extent of muscle wasting among carriers of the 17p12 duplication associated with the degenerative neuropathy of Charcot-Marie-Tooth type 1A [61, 62] is nominally reduced in duplication carriers with a protective PGS_GW-grip strength_ (*β*_DUP_ = -10.1 kg; p_DUP_ = 3.8 x 10^-48^; among duplication carriers: *β*_PGS_ = 13.4 kg; p_PGS_ = 6.3 x 10^-3^). Hence, PGS_GW_ can help identify CNV carriers that are at the highest risk of developing severe CNV expression and would benefit the most from lifestyle and/or therapeutic interventions. Together, these examples illustrate how the polygenic background contributes to phenotypic heterogeneity and variable expressivity of pathogenic CNVs across a broad spectrum of traits.

### CNV-PGS_GW_ interactions are rare

After establishing the additive role of CNVs and PGS_GW_, we explored whether the effect of PGS_GW_ depended on an individual’s CNV carrier status. While we did not detect any CNV-PGS_GW_ interaction surviving Bonferroni correction (p ≤ 0.05/119 = 4.2 × 10^-4^) using adjusted phenotypes, two Bonferroni-significant interactions were identified when analyzing raw phenotypic values (Supplementary Table 5). These two scale-dependent interactions both mapped to the previously described 22q11.23 region (Fig. 2e) and revealed a stronger contribution of the polygenic background to increased GGT levels (*β_interaction_* = 35.2 U/L; p = 2.0 × 10^-5^) and decreased grip strength (*β_interaction_* = -12.0 kg; p = 1.4 × 10^-4^) among CNV carriers. Importantly, we confirm that these interactions are not artifacts of poor genotype quality, as both remain Bonferroni-significant when using a PGS*_trans_* that excludes SNVs within ± 250 kb of the CNV region (p_GGT_ = 3.4 × 10^-5^; p_grip strength_ = 1.4 × 10^-4^). In a second time, we repeated the analysis substituting CNV carrier status for an individual’s genome-wide deletion or duplication burden, defined as the number of genes disrupted by deletions or duplications, respectively. We did not detect any Bonferroni-significant interactions on either the normalized or raw phenotypic scale (p ≤ 0.05/(2 × 43) = 5.8 × 10^-4^) (Supplementary Table 6).

Failure to detect interactions might stem from limited power. Indeed, within the empirically observed range of CNV frequencies (5 × 10^-5^ to 5 × 10^-3^) and interaction effect sizes on the adjusted phenotype scale (≤ 0.012), our power to detect an interaction is virtually null (Supplementary Fig. 4). On the raw scale, we observe larger effect sizes, with half of the CNV-trait pairs having an interaction term with an effect size ≥ 1 (Supplementary Table 5). This improves detection power, possibly explaining why significant interactions were detected on that scale. We conclude that while we detected two scale-dependent interactions wherein the impact of the polygenic background was exacerbated among CNV carriers, there is, in most cases, no evidence to support that the polygenic background impacts complex traits differently among CNV carriers.

### Carriers of trait-increasing CNVs have higher PGS

Given the healthy cohort bias of UKBB, we hypothesized that CNV carriers have a polygenic background that counteracts the CNV’s effect, resulting in carriers having a different PGS_GW_ distribution than copy-neutral individuals (Fig. 1a). Strikingly, 90 (76%) CNV-trait pairs had a CNV effect on PGS_GW_ whose direction *aligned* with the CNV effect on the adjusted phenotype (p < 1 × 10^-3^, based on 1,000 permutations; Fig. 3a; Supplementary Fig. 5; Extended Data 2; Supplementary Table 7). Focusing on 11 CNV-trait pairs with a nominally significant CNV effect on PGS_GW_, ten were directionally concordant, indicating that CNV carriers tend to have a PGS_GW_ that exacerbates the CNV’s phenotypic impact.

**Figure 3.**
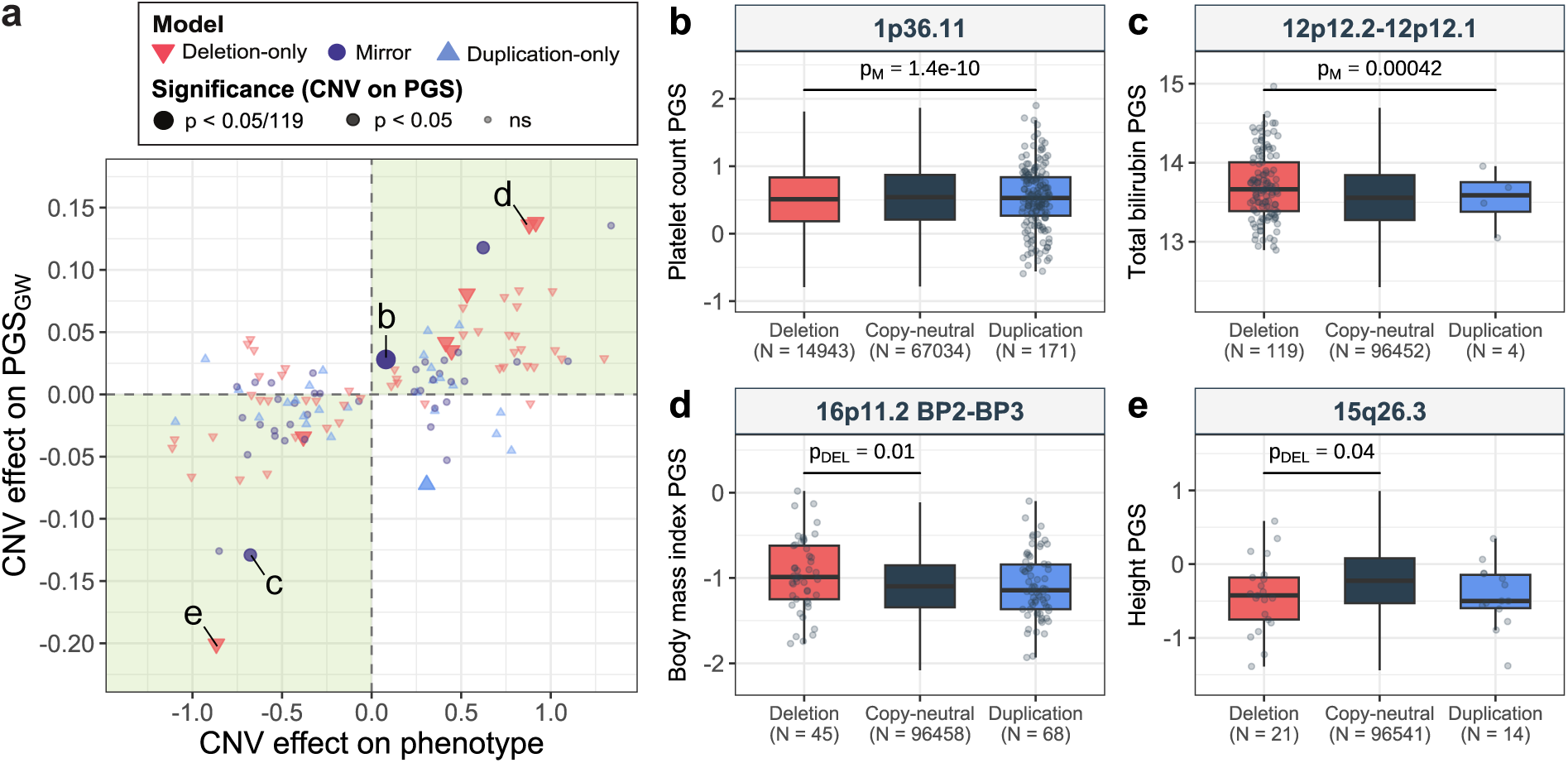
CNV carriers harbor a PGSGW that reinforces the effect of the CNV. **a,** Effect of CNV carrier status on genome-wide PGS (PGSGW) (y-axis) against the effect on adjusted phenotypes (x-axis) for 119 CNV-trait pairs. Shape and color indicate the assessed model, while size and transparency reflect the significance of the CNV effect on the PGSGW, with “ns” indicating p *>* 0.05. Green areas encompass CNV-trait pairs where carriers of a trait-associated CNV have a PGSGW that is directionally concordant with the phenotypic effect of the CNV. CNV-trait pairs depicted in **b-e** are labeled. **b-e,** Boxplots representing the distribution of PGSGW (y-axis) for **b,** platelet count, **c,** total bilirubin, **d,** body mass index (BMI), and **e,** height according to CNV carrier status at **b,** 1p36.11, **c,** 12p12.2-12p12.1, **d,** 16p11.2 BP2-BP3, and **e,** 15q26.3 (x-axis). The number of individuals is indicated as N. Individual data points are shown for CNV genotype categories with N *≤* 1,000. Significance of the CNV effect on PGSGW encoded according to the mirror (M) or deletion-only (DEL) model is displayed.

The copy-number of the 1p36.11 region encompassing the *RHD* gene – which encodes the Rhesus (Rh) factor antigen key in determining the Rh blood group – showed a Bonferroni-significant positive effect (p ≤ 0.05/119 = 4.2 × 10^-4^) on both platelet count and PGS_GW-platelet count_ (p = 1.4 x 10^-10^; Fig. 3b). Other nominally significant examples include the association between the 12p12.2-12p12.1 copy-number and both decreased levels of total bilirubin and lower PGS_GW-bilirubin_ (p = 4.205 × 10^-4^ > 0.05/119; Fig. 3c). Similarly, higher PGS_GW-BMI_ was observed among carriers of the BMI-increasing 16p11.2 BP2-BP3 deletion (p = 0.012; Fig. 3d), which encompasses the obesity-linked gene *SH2B1* [30, 63]. Finally, lower PGS_GW-height_ was observed among carriers of a 15q26.3 partial deletion of *ADAMTS17* (p = 0.042; Fig. 3e). *ADAMTS17* is linked to autosomal recessive Weill-Marchesani syndrome, which is characterized by short stature [64]. Despite individual effects being weak, the observed data pattern appears conserved across a broad spectrum of CNV-trait pairs. Importantly, it is incompatible with strong selective participation bias, which would induce a negative (spurious) correlation between CNV and PGS_GW_ (Fig. 1a).

### *Cis*-tagging only partially explains the positive correlation between CNV-PGS_GW_

A first hypothesis explaining the positive CNV-PGS_GW_ correlation is tagging of the CNV by PGS_GW_, so that the presence of the CNV is positively correlated through linkage with SNVs included in PGS_GW_ (Fig. 1b). To test this, we decomposed PGS_GW_ into a PGS*_cis_*based solely on SNVs mapping to the CNV region ± 50 kb, ± 100 kb, ± 250 kb, and the full chromosome and conversely, a PGS*_trans_* excluding SNVs that mapped to these intervals. We then calculate the Pearson correlation between these partial PGSs and the CNV carrier status (Supplementary Table 8). Focusing on ten CNV-trait pairs with a nominally significant effect of the CNV carrier status on PGS_GW_ whose direction aligned with the CNV’s effect on the phenotype, the association between the 1p36.11 copy-number and increased platelet count stood out by showing a strong, Bonferroni-significant correlation between CNV status and PGS_cis_, indicating that the CNV-PGS_GW_ was primarily driven by local linkage disequilibrium (Fig. 4). Notably, this was the only signal for which the phenotypic impact of the CNV was significantly diminished upon conditioning on PGS_GW_ (Supplementary Fig. 6; Supplementary Table 3). With a frequency of 3.8%, the *RHD* deletion is the only common CNV in our study. The region was highlighted by the Human Pangenome Reference Consortium for its extreme complexity, with a frequency of 17% for the haplotype carrying the full *RHD* deletion [65], suggesting that our CNV calls underestimate the true deletion frequency. Still, the fact that this locus is the only one with robust evidence of tagging aligns with such a mechanism relying on the CNV being present on (a) specific haplotype(s) – as opposed to appearing recurrently *de novo* – allowing the formation of a correlation pattern between the CNV and SNVs.

**Figure 4.**
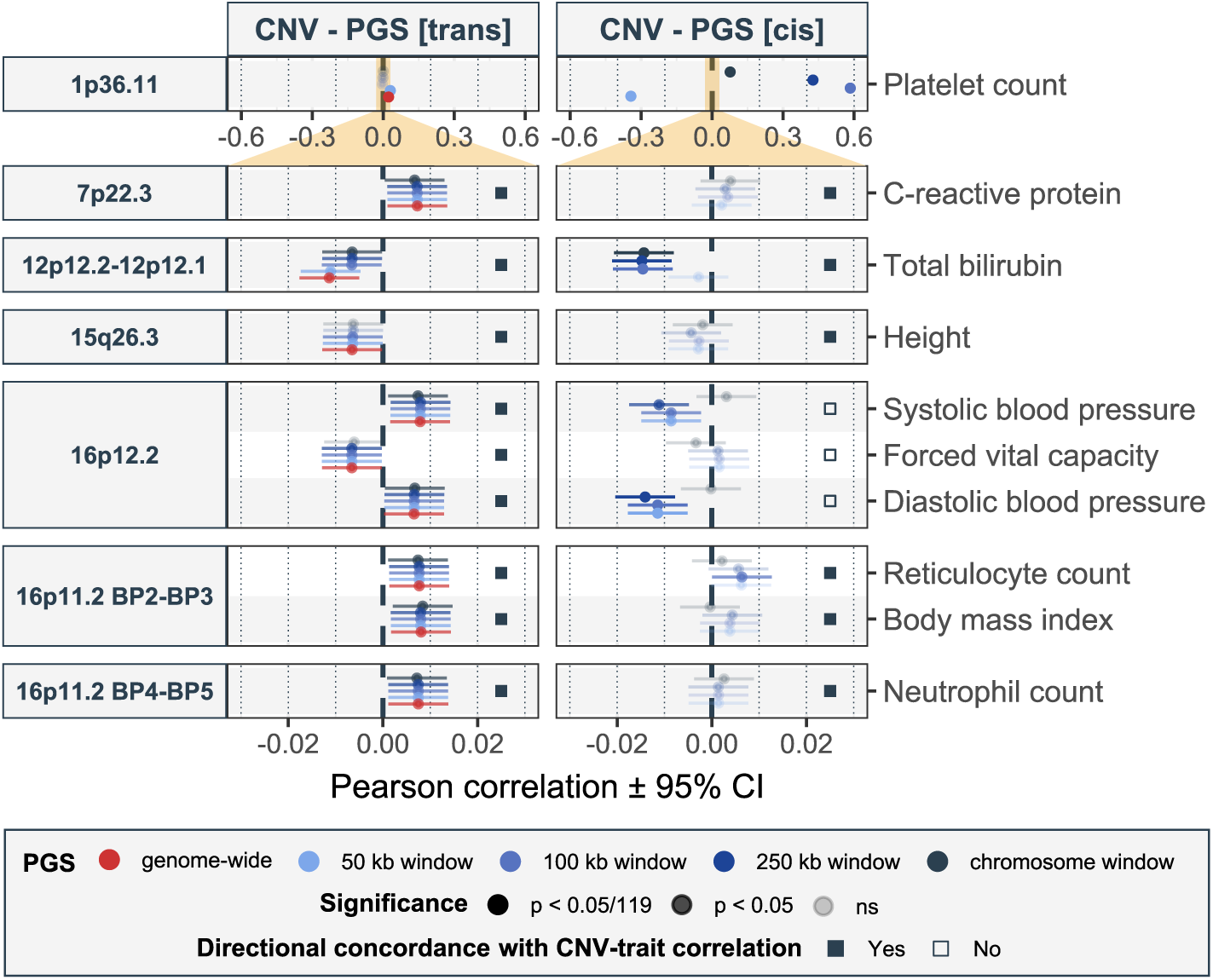
*Cis*-tagging only partially explains the positive correlation between CNV-PGSGW. Pearson correlation with 95% confidence interval (CI; x-axis) between polygenic scores (PGSs; y-axis, right) and CNV carrier status (y-axis, left) for ten CNV-trait pairs with a nominally significant effect of the CNV carrier status on PGSGW whose direction aligned with the CNV’s effect on adjusted phenotypes. The left panel shows correlation with the genome-wide PGSGW (red) and various PGS*trans* (blue shades), while the right panel shows correlations with various PGS*cis*(blue shades). Transparency reflects the significance level, with “ns” indicating p *>* 0.05. The small grey square indicates whether the correlation between the CNV carrier status and PGS*trans*-250kb (left panel) or PGS*cis*-250kb (right panel) are directionally concordant with the CNV’s effect on the phenotype. We use a different x-axis scale for the 1p36.11 CNV region for better visualization (orange area).

We next turned to the remaining nine signals (Fig. 4), of which three showed at least a nominally significant CNV-PGS*_cis_* correlation. Interestingly, 12p12.2-12p12.1 CNVs seem to be tagged by bilirubin-associated SNVs located 50-100 kb from the CNV region, as indicated by the lack of a significant CNV-PGS*_cis_*_-50kb_ correlation and a drop in PGS*_trans_* correlation excluding variants within the 50-100 kb range. The 12p12.2-12p12.1 *cis* region contains other genes from the *SLCO* family with overlapping functions in bilirubin transport [66], so that variation in that locus represents a strong determinant of bilirubin levels. Yet, PGS*_trans_*_-chr12_ remained correlated at nominal significance with the CNV carrier status, indicating that linkage disequilibrium cannot fully account for the CNV-PGS_GW_ correlation. The two other pairs with a nominally significant CNV-PGS*_cis_*correlation involve 16p11.2 deletions with systolic and diastolic blood pressure, for which we observe local tagging within the ± 250 kb region. Interestingly, for these pairs, the direction of correlation between the CNV and PGS*_cis_*opposes the direction of correlation with PGS*_trans_*, which itself aligns with the deletion’s impact on blood pressure. Hence, local *cis*-tagging masks part of the CNV-PGS_GW_ correlation, indicating that the latter is not driven by *cis*-tagging.

We conclude that among these ten CNV-trait pairs, one is fully accounted for by *cis*-tagging, three show mixed evidence of *cis*-tagging in combination with another mechanism driving CNV-PGS_GW_ correlation, and six show no evidence for *cis*-tagging. We further note that 20 out of 29 CNV-trait pairs with a nominally significant CNV-PGS*_cis_*but no significant CNV-PGS_GW_ correlation have opposing CNV-PGS*_cis_* and CNV-PGS_trans_ correlations (Supplementary Fig. 7). This creates a scenario wherein local and genome-wide background oppose each other, making it less likely for the CNV-PGS_GW_ correlation to reach significance.

### Assortative mating exacerbates the phenotypic impact of rare CNVs

Next, we explored assortative mating as an alternative mechanism to explain the remainder of the positive CNV-PGS_GW_ correlation. Under assortative mating, phenotypic similarity caused by one parent’s high PGS_GW_ and the other parent’s riskincreasing CNV results in coinheritance of these genetic risk factors in the offspring (Fig. 1c). First, we examined correlations between adjusted phenotypes among the 3,396 UKBB couples present in our test set. A total of 33 (77%) same-trait comparisons showed significant positive correlations, with vitamin C (*r* = 0.48, p = 1.1 × 10^-165^), BMI (*r* = 0.26, p = 7.8 × 10^-53^), body fat mass (*r* = 0.25, p = 2.4 × 10^-49^), weight (*r* = 0.23, p = 9.2 × 10^-43^) and height (*r* = 0.21, p = 8.0 × 10 ^-36^) showing the strongest correlation (Supplementary Fig. 8; Supplementary Table 9). Comparing these estimates to those obtained from 51,664 couples present in the full UKBB data set yielded highly consistent estimates (*r* = 0.98, p = 6.6 × 10^-32^; Supplementary Fig. 9a; Supplementary Table 10). Similarly, using a stricter couples’ definition that identified 1,213 couples within our test set yielded highly correlated estimates (*r* = 0.97, p = 8.5 × 10^-27^; see Methods; Supplementary Fig. 9b; Supplementary Table 10). This confirms widespread phenotypic assortment across diverse traits in our sample, which could channel correlation between CNVs and PGS_GW_.

Next, we leveraged within-couple phenotypic correlation to estimate the fraction of the total CNV-PGS*_trans_*_-250kb_ correlation that can be explained through a path mediated by assortative mating. The latter is defined as the product of the CNV-phenotype correlation (*α*), the within-couple phenotypic correlation (*β*), and the PGS*_trans_*_-250kb_-phenotype correlation (*γ*) (Fig. 1c). We decided to use PGS*_trans_*_-250kb_, which mimics PGS_GW_ while ensuring that our estimation of assortative mating is not conflated by linkage disequilibrium. Indirect CNV-PGS correlations estimated via the assortative mating path significantly correlated with total CNV-PGS*_trans_*_-250kb_ correlation (*r* = 0.45, p = 2.0 × 10^-7^; Fig. 5a; Supplementary Table 11), and 50,000 permutations of *α* (p < 2 × 10^-5^), *β* (p = 4.4 × 10^-4^), or *γ* (p < 2 × 10^-5^), showed it was highly unlikely to obtain a total-indirect CNV-PGS*_trans_*_-250kb_ correlation larger than 0.45 (Fig. 5b). As expected, the total CNV-PGS*_trans_*_-250kb_ correlation negatively correlated with the indirect path expected under selective participation bias, discarding this competing scenario as the driving force behind our results.

**Figure 5.**
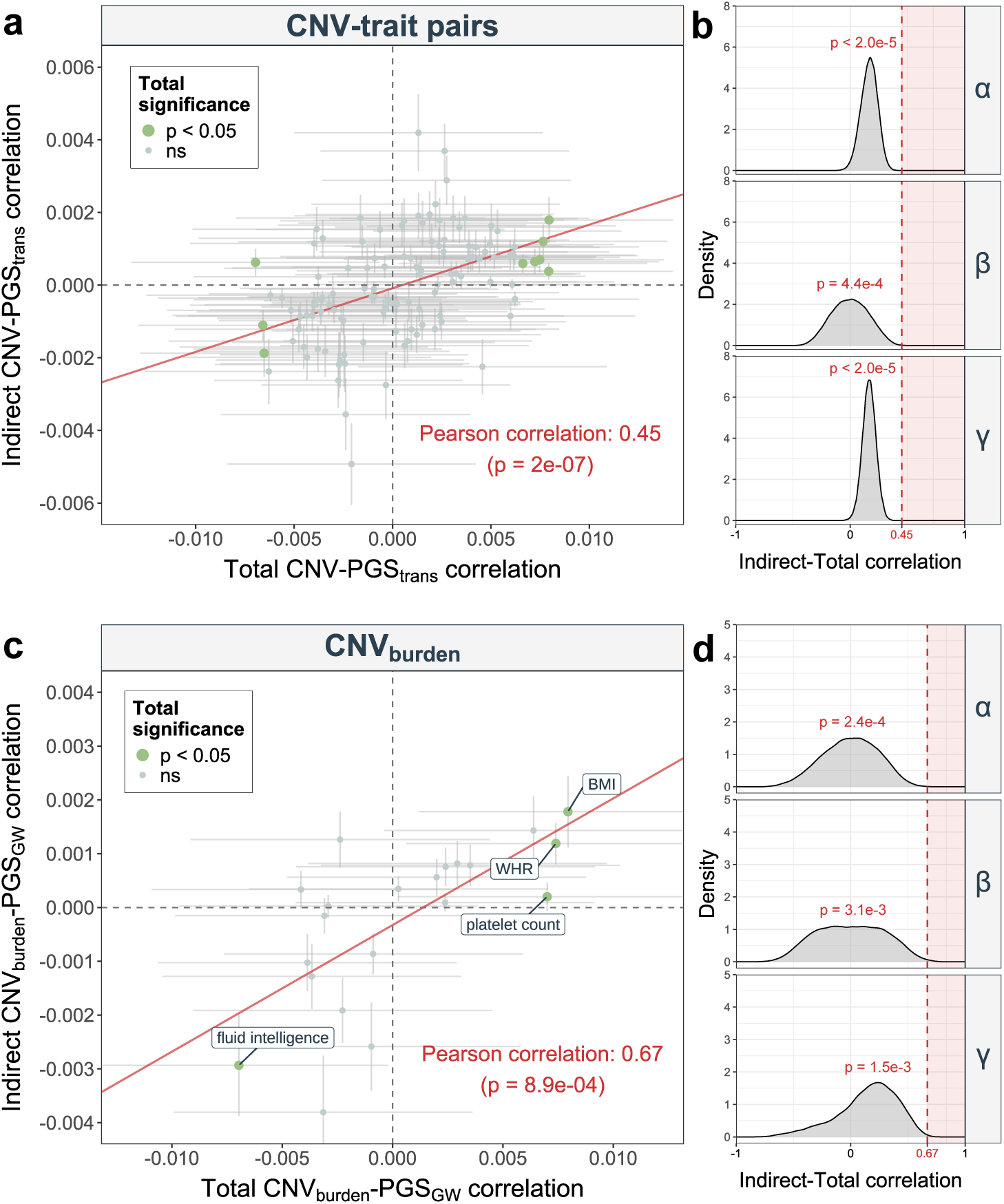
Assortative mating partially explains the CNV-PGS correlation. **a,** Correlation between total (x-axis) and indirect (y-axis; through assortative mating) CNV-PGS*trans*-250kb correlation for 119 CNV-trait pairs. Error bars represent 95% confidence intervals. Size and color reflect the significance level of the total CNV-PGS*trans*-250kb correlation, with “ns” indicating p *>* 0.05. The red line represents the fitted linear regression of the indirect CNV-PGS*trans*-250kb correlation against the total CNV-PGS*trans*-250kb correlation. **b,** Density plots displaying the distribution of the total versus indirect CNV-PGS*trans*-250kb correlations obtained from 50,000 permutations of *α* (top), *β* (middle), and *γ* (bottom) across the 119 CNV-trait pairs. The empirical correlation and associated p-value are denoted in red. Values more extreme than the observed correlation are shaded in red. **c,** Correlation between total (x-axis) and indirect (y-axis; through assortative mating) CNV burden-PGSGW correlation for 21 traits significantly (p *≤* 0.05/43 = 1.2 *×* 10^-3^) associated with the CNV burden and **d,** associated permutation tests’ results, following the same legend description as in (**a**) and (**b**), respectively, with nominal-significant effects labeled. BMI = body mass index; WHR = waist-to-hip ratio.

To ensure the robustness of our findings, we performed a series of sensitivity analyses. Repeating the analysis excluding couples used to calculate *β* to estimate *α* and *γ* produced similar results (*r* = 0.42; p = 2.2 × 10^-6^; Supplementary Fig. 10a; Supplementary Table 12), as did adjusting phenotypes for potential interactions between sex and other covariates (*r* = 0.46, p = 1.3 × 10^-7^; Supplementary Fig. 10b; Supplementary Table 13) and for coordinates of residence (*r* = 0.45, p = 2.1 × 10^-7^; Supplementary Fig. 10c; Supplementary Table 14). To further ensure that these results are not driven by overestimation of *α* due to Winner’s curse, we repeated the analysis based on 98 CNV-trait pairs that survive a stricter multiple-testing correction threshold for the CNV-trait association strength (see Methods). Despite the lower number of tested pairs (-18%) reducing our power to detect a signal through the assortative mating path, the correlation remained unchanged (*r* = 0.46, p = 2.0 × 10^-6^; Supplementary Fig. 10d; Supplementary Table 15). From a theoretical perspective, we show that analytically derived boundaries for *r*(*CNV, PGS_GW_*) are wide and thus not expected to correlate with the product of *α* × *β* × *γ*, indicating that our results are not due to mere mathematical constraint of correlated variables (Supplementary Note 1). We also show – analytically and through simulations – that under assortment, the correlation between CNV carrier status and PGS_GW_ is approximated by *r*(*CNV, PGS_GW_*) = *α* × *β* × *γ* (Supplementary Notes 2 and 3). Together, these analyses indicate that our finding is unlikely to be driven by artifacts and suggest that a substantial fraction of CNV-PGS_GW_ correlation might be explained by assortative mating.

### Rare CNVs are often inherited

Assortative mating can only account for CNV-PGS_GW_ correlation if at least a substantial proportion of CNVs are inherited. Given that many assessed CNVs are recurrent and are expected to have a high *de novo* rate, we sought confirmation of this prerequisite by estimating inheritance rates across the 27 studied CNV regions. Seven deletions and ten duplications met our criteria to assess inheritance rate (see Methods). All 17 testable CNVs exhibited a non-zero inheritance rate, with a pooled inheritance rate of 23% [95%-CI: 21-25%] and 79% [95%-CI: 73-84%], for deletions and duplications, respectively (Supplementary Table 16). Despite the low number of relatives in UKBB hampering the precision of our estimates, these numbers indicate that the assessed CNVs can be inherited in the general population, supporting our proposed assortative mating model.

### Widespread positive correlation across mutational classes

Finally, we tested whether our proposed mechanism of genetic compounding through assortative mating could be generalized beyond focal CNV-trait pairs. First, we estimated the association of the CNV burden with PGS_GW_ for 21 traits impacted by the CNV burden (p ≤ 0.05/43 = 1.2 × 10^-3^; Supplementary Table 17). While no effect survived multiple testing correction (p ≤ 0.05/21 = 2.4 × 10^-3^), we observed nominally significant effects of the CNV burden on the PGS_GW_ for BMI (p = 0.021), waist-to-hip ratio (p = 0.032), platelet count (p = 0.043), and fluid intelligence (p = 0.044). In line with earlier findings, 18 out of 21 effects were directionally concordant with the effect of the CNV burden on the adjusted phenotypes – including the four nominally significant effects described above – suggesting that carriers of trait-associated CNVs are more prone to carry a phenotype-exacerbating PGS_GW_. We tested whether this pattern could be accounted for by assortative mating by substituting the locus-specific CNV carrier status by the CNV burden in our assortative mating framework (Fig. 1c). We observed a significant correlation between the total CNV burden-PGS_GW_ correlation and the indirect path mediated by assortative mating (*r* = 0.67, p = 8.9 × 10^-4^) (Fig. 5c; Supplementary Table 18). Permutation of *α* (p = 2.4 × 10^-4^), *β* (p = 3.1 × 10^-3^), and *γ* (p = 1.5 × 10^-3^) supported the significance of our results (Fig. 5d).

Second, we examined if the association between PGS and CNVs extended to variants of other mutational classes. A total of 15 traits were significantly modulated by the autosomal predicted loss-of-function (pLoF) SNV burden (p ≤ 0.05/43 = 1.2 × 10^-3^; Supplementary Table 17). For these, we estimated the pLoF burden’s impact on PGS_GW_ and identified a single nominally significant effect on PGS_GW-BMI_ (p = 0.027) whose effect size was directionally concordant with the pLoF’s burden effect on adjusted BMI (Supplementary Table 17). Finally, we observed a small but significant correlation between the pLoF and CNV burden (*r* = 0.005; p = 3.9 × 10^-3^). We note that the rarity of CNVs in our dataset makes it unlikely to pick up any strong correlation. Indeed, simulating the CNV and pLoF burdens based on a negative binomial and normal distributions, respectively (Supplementary Fig. 11a), we show that we have _∼_75% power to detect a correlation of 0.005 (Supplementary Fig. 11b), yet tend to underestimate the correlation’s magnitude (Supplementary Fig. 11b-c). Although evidence remains limited and larger studies will be required to confirm these trends, current data align with accumulation of genetic variants with coordinated phenotypic effects across mutational classes as a widespread phenomenon within the general population.

## Discussion

In this study, we explored the interplay between rare deleterious CNVs and common genetic variants, demonstrating that CNV expressivity is influenced by the carrier’s polygenic background through additive effects. We also observed a mild but widespread positive CNV-PGS correlation, most of which could not be accounted for by linkage disequilibrium or participation bias. We propose assortative mating as a mechanism that could explain this relationship, supported by a non-null inheritance rate for a substantial fraction of the studied CNVs. A similar trend of positive correlation was observed between CNV or pLoF burdens and PGS, suggesting a pervasive mechanism in which the impact of rare deleterious variants is compounded by common variants through assortative mating.

There has been a surge in interest towards better conceptual understanding of how common and rare variants jointly influence complex traits. Studies have demonstrated that polygenic background contributes to the variable expressivity of rare “monogenic” variants, allowing improved risk estimation [49, 67, 68]. Targeted studies focusing on specific phenotypes such as autism [27, 46, 53], schizophrenia [27, 47, 51, 57], intellectual disability and developmental delay [48, 55, 56, 69, 70], or obesity [71, 72] showed that overall, rare and common variants act additively. Our results reinforce this body of evidence by extending these findings to a more diverse set of quantitative traits, supporting that PGS_GW_ can stratify CNV carriers to identify those who are at increased risk of developing CNV-associated comorbidities. Indeed, CNV carriers with a PGS_GW_ counteracting the effect of the CNV exhibited phenotype values within the range of copy-neutral individuals, while carriers with a phenotype-exacerbating PGS_GW_ tended to exhibit phenotypes outside the normal range. This pertains to the relative contribution of CNVs versus PGS, the latter being reported to have a broader impact on socio-economic outcomes than pathogenic CNVs [73]. We argue that while PGSs might be more relevant at the population scale, the presence of a CNV has important implications at the individual level and future studies should explore how to best integrate CNV and PGS to optimize relevance in terms of personalized medicine.

Focusing on more complex CNV-PGS relations, we tested for non-additive effects on two different scales and identified two robust, scale-dependent interactions wherein CNV and polygenic background exacerbate each other’s phenotypic impact. The population distribution of GGT levels – and to a lesser extent of grip strength – is skewed and non-normal, possibly explaining why the interactions become apparent only on the raw phenotypic scale. To date, only weak CNV-PGS interactions have been reported [27, 44, 45, 49, 54, 67, 68, 71, 72], so that these effects represent among the first robust evidence of epistasis between rare CNVs and polygenic background. Yet, further research will be required to disentangle scale from interaction effects [74], and assess whether these findings are of clinical relevance, i.e., whether phenotypic differences among individuals that are outside the normal phenotypic range have clinical relevance. It is also worth noting that we might miss other true interactions due to lack of power or because the interaction only becomes visible with phenotypic transformations that were not tested. Indeed, theoretical work predicts that disease risk variants might exhibit stronger phenotypic consequences in highrisk polygenic background due to the nonlinear relationship between disease liability and disease probability [75]. This emphasizes the importance of the choice of the link function for binary traits and the choice of the scale/transformation for continuous outcomes, as well as the key role of ascertainment to accurately characterize rare variant pathogenicity. We conclude that while we could demonstrate interactions for specific CNV-trait pairs, the majority of assessed pairs do not show evidence of deviation from additivity at current sample sizes.

While the effect of PGS_GW_ on the phenotype appeared mainly independent of CNV status, the CNV status correlated positively with an individual’s PGS_GW_, paralleling a recent report showing that UKBB carriers of CNVs causing neurodevelopmental conditions have lower PGS_GW_ for educational attainment [55]. We found this pattern to be widespread across diverse phenotypes and, after excluding that it was fully driven by linkage disequilibrium, identified assortative mating as a likely pervasive contributor, although we note that it only partially explains the observed CNV-PGS_GW_ correlation. While the imperfect agreement between the CNV-PGS_GW_ correlation and our predictor of it is – at least in part – due to regression dilution bias (as our predictor is a product of three noisy estimates), other factors that we did not explore might also play a role. For instance, while we aimed to account for population stratification by restricting our analyses to a single genetic ancestry and controlling for 40 SNV-derived principal components as well as coordinates of residence, we cannot exclude that part of the positive CNV-PGS_GW_ is driven by residual population stratification. Still, CNV-PGS_GW_ correlation induced by population stratification is not expected to be proportional to assortative mating; while it might account for part of the CNV-PGS_GW_ correlation that is *not* explained by assortative mating, it is unlikely to drive our observations.

Positive phenotypic assortment is well-established for neuropsychiatric phenotypes, anthropometric features, and lifestyle factors [39, 76–79], with recent biobank studies reporting partner phenotypic correlation among less conspicuous traits, e.g., blood measurements [76]. Studies of the downstream consequences of assortative mating on genomic architecture have emphasized how ignoring its impact can bias estimation of key genetic metrics [80–89], yet, the mechanisms through which it acts and the full scale of its implications remain underappreciated. Given assortative mating on a given trait, if a rare CNV contributes to that trait and is inherited – which we demonstrate is a plausible assumption – we expect compounding genetic factors in subsequent generations until the number of accumulated variants starts affecting fitness. This induces a positive correlation between CNV (or other rare variants) and PGS_GW_, analogous to what was observed for PGSs between different chromosomes [81]. Previous work has shown that over generations, assortative mating correlates with co-occurrence of psychiatric conditions between parents and offspring [79] and leads to genetic anticipation, wherein disease liability increases over generations among families carrying rare pLoF variants with variable expressivity [39]. Another study proposed parental assortment to explain the correlation between PGS and rare variant burden associated with cognitive and educational outcomes both between partners and within individuals [90]. Our data support this mechanism and suggest that it operates across a broader spectrum of variants and phenotypes.

Our assortative mating framework is targeted toward detecting direct assortment, which induces a correlation in genetic variants directly associated with the trait on which assortment takes place. This scenario is conceivable for traits such as height or BMI, but seems improbable for traits such as bilirubin levels. Yet, these phenotypes can act as biomarkers, correlating to visible traits on which assortment might occur. Such complex mechanisms are evidenced by both direct same-trait and indirect crosstrait assortment having been demonstrated in the UKBB [76]. Acknowledging that mate selection is likely to be based on a large combination of traits, a partner-choice index has recently been proposed [91]. An interesting related consideration is that most CNVs assessed in this study are pleiotropic. As such, polygenic liability for various traits might determine the presence and/or severity of a given phenotype, explaining phenotypic heterogeneity across carriers of the same CNV. One key question, however, is whether PGSs for different CNV-associated traits are independent. Persistent cross-trait assortative mating was shown to induce spurious genetic correlations [84], highlighting the importance of factoring in both pleiotropy and polygenicity [92] – an extension that goes beyond the scope of this study.

Another consideration is sample ascertainment, as cohorts in which healthy participants are over-(e.g., volunteer samples [58, 93, 94]) or under- (e.g., case-control studies [95, 96] or electronic health record-linked biobanks with higher disease burden [97, 98]) represented may exhibit a negative CNV-PGS correlation (Fig. 1a), as documented in previous clinical studies [44, 45, 47, 50]. In UKBB, we observed a weak, yet consistent, positive CNV-PGS correlation. While healthy volunteer bias in UKBB could have attenuated this association, it did not outweigh the positive correlation induced by assortative mating. We conclude that while selective participation in volunteer biobanks may alter the magnitude of observed effects and should therefore be acknowledged, correct inference with respect to directionality of effects remains largely possible [58, 99]. Case-control studies, in contrast, are subject to extreme participant selection, possibly canceling out any positive CNV-PGS correlation induced by assortative mating. This suggests that genetic findings obtained from population biobanks are likely to be more robust against selection bias than those obtained from clinical studies, highlighting the usefulness of the former when interrogating the role of rare and common variants in disease susceptibility.

Finally, our study should be interpreted in light of some limitations. The examined CNVs were detected from genotyping microarrays and span large genomic regions, with the corollary that they have strong phenotypic consequences and are rarer. While these characteristics were purposefully selected for in our study, low CNV frequency limits the power of our analyses and influences its outcomes; we expect that investigating CNVs with higher frequencies and smaller effect sizes would identify more cases in which *cis*-tagging represents a dominant mechanism. Indeed, SNV associations have been found to tag stronger associations with smaller structural variants [22, 100–102]. Accordingly, the only common CNV in our analyses – the *RHD* deletion – was subject to *cis*-tagging. Recent efforts in producing large genome sequencing datasets coupled with the development of methods allowing robust detection of structural variants across the size and allele frequency spectrum will allow exploration of the interplay between different mutation classes at a scale [100, 103]. Furthermore, because PGS portability decreases with genetic distance [104], we restricted our study to a single genetic ancestry group with a sufficiently large sample size to explore the interplay between PGS and rare CNVs. This stresses the importance of continued efforts to develop large and diverse biobanks with rich phenotypic data to enable replication studies and extend the proposed framework to a broader set of phenotypes, including binary disease outcomes. Larger sample sizes and further methodological development will also enable better estimation of CNV inheritance rates, allowing incorporation of the latter in our assortative mating frame-work. Improved collection and integration of genetic and environmental factors will likely reveal a more complex and nuanced picture of the mechanisms shaping variable expressivity. Importantly, because participation bias and assortative mating have opposing effects on the CNV-PGS correlation, we likely underestimate the role of assortative mating. This is exacerbated by our observation that local linkage disequilibrium can oppose the global CNV-PGS_GW_ correlation. Finally, our assortative mating model ignores potentially pleiotropic CNV and PGS effects acting on (further) off-target traits under assortment, an aspect that could be further explored in future studies.

In conclusion, our study supports the notion that rare deleterious CNVs and common polygenic back-grounds additively shape diverse phenotypes and identifies assortative mating as a candidate mechanism promoting the co-occurrence of risk-associated common and rare genetic contributions.

## Materials and methods

### Study material

#### Software

Analyses were run on the UK Biobank Research Analysis Platform using Python v3.11.5 and R v4.4.0.

#### Cohort description & sample selection

This study is based on volunteer participants from UKBB, a population cohort encompassing _∼_500,000 individuals (54% females) from the UK general population aged 40-69 years at recruitment (2006-2010). Genetic data from genotype microarray (UKBB field identifier #22418) and exome sequencing (#23149) are accessible for all participants, along with extensive phenotypic assessment [105, 106]. Presented analyses were performed on various subsets of “white British” individuals (N = 409,703), defined as in white british ancestry subset==1 in ukb sqc v2.txt. UKBB has approval from the NorthWest Multicentre Research Ethics Committee as a Research Tissue Bank. All participants signed a broad informed consent form. Data were accessed under application 16389.

#### CNV association data

Focal CNV-traits pairs stem from previous work by our group [19]. Briefly, CNV genome-wide association studies (GWASs) were performed in the UKBB to explore the association between the copy-number of genome-wide CNV-proxy probes derived from microarray-based CNV calls and 57 quantitative traits according to three association models: a mirror model in which deletions and duplications have opposing phenotypic consequences, as well as deletion-only and duplication-only models assessing the isolated impact of the deletion or duplication, respectively. We identified 131 genome-wide significant signals characterized by a lead CNV-proxy probe [19]. These were filtered for autosomal CNV regions and phenotypes measured in both males and females, resulting in 119 associations involving 43 phenotypes and 27 CNV regions.

#### Phenotype transformation

Raw phenotypic measurements for the 43 analyzed quantitative traits (see CNV association data) were extracted and averaged over available instances. Grip strength was defined as the mean of right and left hand grip strength (#47 and #46). WHR was defined as the ratio between waist and hip circumference (#48 and #49); WHR adjusted for body mass index (WHRadjBMI), by regressing WHR on BMI (#21001). To develop PGS scoring files, phenotypes were rank-transformed (`rntransform` from the GenABEL v1.8.0 R package) within the training sample set (see Polygenic scores calculation). For association studies with CNVs, measurements were inverse normal transformed across the test set (see Polygenic scores calculation) before correcting traits for sex, age (#21003), age^2^, genotyping batch, and principal components (PCs) 1-40 [19]. The latter are referred to as adjusted phenotypes throughout the manuscript.

#### CNV carrier identification & CNV burden calculation

We called genotype microarray-based CNVs (GRCh37/hg19) using standard PennCNV v1.0.5 settings [107] for all UKBB participants [19]. Briefly, each CNV call was attributed a quality score ranging between -1 (likely deletion) and 1 (likely duplication) [108]. Only high-confidence CNVs with absolute value of quality score > 0.5 were retained. CNV carrier status was established for each UKBB participant across the 119 assessed CNV-trait pairs (see CNV association data): individuals were considered as deletion or duplication carriers when harboring a high-quality deletion (copy-number = 1) or duplication (copy-number = 3), respectively, overlapping the lead CNV-proxy probe for that signal. Copy-neutral individuals were defined as having no evidence of a CNV (even with low quality score) overlapping the lead CNV-proxy probe. For association studies, CNV carrier status was encoded according to the model yielding the most significant CNV-trait association [19], using the following deletion/copy-neutral/duplication encoding for the mirror (-1/0/1), deletion-only (1/0/NA), and duplication-only (NA/0/1) models. Individual-level CNV, deletion, and duplication burdens were calculated as previously described [19], reflecting the number of hg19 RefSeq genes disrupted by high-confidence autosomal CNVs, deletions, and duplications, respectively (≥ 1 bp overlap with exons, splice sites, non-coding RNAs, or UTRs).

#### Polygenic scores calculation

Due to the lack of standardized scoring files for all analyzed phenotypes in other cohorts, we divided the “white British” UKBB samples into a training set (n = 264,372; 54% females), used to develop PGS scoring files, and a CNV-carrier-enriched test set (n = 96,716; 54% females) (Supplementary Fig.1). Individuals carrying a high-confidence CNV (absolute value of quality score: > 0.5) overlapping the lead probe of one of the 119 analyzed CNV-trait pairs or an overall high CNV burden (≥ 10 genes) (see CNV carrier identification & CNV burden calculation) were pre-selected for the test set and pruned to 38,236 unrelated individuals (kinship ≤ 0.0884) by iteratively removing the sample with the highest number of relatives. These individuals and all their relatives (≤ 3^rd^ degree, regardless of CNV carrier status) were excluded from the 409,703 “white British” samples, along with individuals carrying a lower-confidence CNV call (absolute value of quality score: 0.2–0.5). Remaining individuals were randomly split into training (80%) and control (20%) sets with matching age and sex distributions. Individuals in the control set were pruned to unrelated individuals and merged with pre-selected CNV carriers to form the complete test set. Non-carriers in the test set related to samples in the training set were excluded from the test set. Finally, individuals whose reported sex did not match their genetically inferred sex (`Submitted.Gender` and `Inferred.Gender` in `ukb sqc v2.txt`) were excluded from both sets.

GWASs were conducted in the training set for the 43 rank-transformed phenotypes (see Phenotype selection). Briefly, PLINK v2.00a4.3 [109] was used to filter genotype microarray data, excluding variants with minor allele frequency (MAF) < 1% (`--maf 0.01`), missingness rate > 10% (`--geno 0.1`), or Hardy-Weinberg p < 1 × 10^-15^ (`--hwe 1e-15`). Samples with > 10% missing genotype calls (`--mind 0.1`) were excluded. For imputed variants, those with MAF < 0.5% (`--maf 0.005`) or low imputation quality (`--mach-r2-filter 0.3 2`) were removed. GWASs were carried out with Regenie v3.2.9 [110], including age, sex, and PC1-20 as covariates. Only quality-controlled geno-typed variants were used for model fitting (i.e., Regenie step 1). We then tested both genotyped and imputed variant dosages for association, using default parameters except for minimum minor allele count (`--minMAC 1`) (step 2). PGS weights were derived from the resulting GWAS summary statistics, filtered for common (MAF > 1%) high imputation quality (INFO > 0.8) variants present in the HapMap3+ extended set. LDpred2 (snp ldpred2 auto implemented in the bigsnpr v1.12.18 R package) was run using the pre-computed linkage disequilibrium matrix based on UKBB participants of European ancestry, a heritability estimate from LD score regression (using snp ldsc), and default parameters, except for `num_iter=500, shrink_corr=0.95, allow_jump_sign=FALSE, use_MLE=TRUE`, and `vec_p_init=seq_log(1e-4,0.2,length.out=10)` [111–113]. PGS_GW_, defined as the sum of effect allele dosages (from imputed genotypes) weighted by their corresponding effect sizes (derived from LDpred2), were computed for the test set using `pgsc_calc v1.3.2` (output: SUM) [114].

To assess PGS_GW_ performance, we fitted two linear regressions that explain the inverse-normal transformed phenotypic values as a function of covariates-only (i.e., sex, age, age^2^, genotyping batch, and 40 PCs) or covariates in combination with PGS_GW_. Confidence intervals (CI; 95%) were computed for the model’s *R*^2^ using the ci.R2() function from the MBESS v4.9.3 R package [115]. The difference in *R*^2^ between the models (Δ*R*^2^) – which represents the contribution of PGS_GW_ to phenotypic variability – was computed by subtracting the *R*^2^ of the covariate-only model from the *R*^2^ of the model including PGS_GW_. The 95%-CI for Δ*R*^2^ was computed with `ci.R2()`, using a single predictor (K = 1).

#### Rare pLoF burden calculation

A subset of 319,161 unrelated (> 3^rd^ degree) “white British” UKBB participants [25] with available exome sequencing data were used to compute the pLoF burden. pLoF variants were annotated using LOFTEE [116] and SnpEff (retrieved from `ukb23158_500k_OQFE.annotations.txt.gz`) [117]. Only those classified as high-confidence pLoF by LOFTEE and either annotated as pLoF or absent in SnpEff were retained. These were filtered for MAF ≤ 1% among non-related “white British” individuals. Rare pLoF burden was defined as the number of autosomal pLoF variants in an individual’s genome (`plink2 –score --no-mean-imputation`).

### CNV-PGS modeling

#### Significance threshold

Analyses modeling the interplay between CNV and PGS are repeated for the 119 selected CNV-trait pairs (see CNV association data). Significance was determined using Bonferroni correction (p ≤ 0.05/119 = 4.2 × 10^-4^), unless specified otherwise. We report nominally significant signals (p ≤ 0.05).

#### Additive contribution of PGS_GW_ to CNV effect

We modeled the joint contribution of CNV and PGS_GW_ to phenotypic variability by fitting a linear regression model explaining adjusted phenotypes as a function of CNV carrier status (encoded according to the most significant CNV-GWAS model [19]) and PGS_GW_. To specifically assess the contribution of PGS_GW_ among CNV carriers, a linear regression modeling raw phenotypes as a function of PGS_GW_, age, age^2^, sex, and PC1-10 was fitted among CNV, deletion, or duplication carriers, depending on whether the CNV-trait association had previously been found to be most significant under the mirror, deletion-only, or duplication-only model, respectively [19]. The standard error of the PGS_GW_ effect was calculated as the standard deviation of 5,000 bootstrap estimates of the coefficient and was plugged into the Wald test to obtain two-sided p-values. Comparison of effects across phenotypes was done based on standardized effect sizes along with their 95%-CIs. These were obtained by dividing the effect size and the CI endpoints by the standard deviation of the corresponding phenotype in the test set. We used logistic regression to jointly estimate the impact of the number of CNV carriers and the fraction of phenotypic variance explained by PGS_GW_ on the likelihood of a CNV-trait pair to be found Bonferroni significant in the above-described modeling strategy. For visualization, the distribution of the raw phenotypic values is shown for each PGS_GW_ quintile among CNV carriers and copy-neutral individuals. Differences in prevalence of abnormally elevated GGT values (> 50 U/L) among carriers of a 22q11.23 duplication versus copy-neutral individuals was evaluated by two-sided Fisher’s exact test.

#### CNV-PGS_GW_ interaction

For each CNV-trait pair, a linear regression model was fitted that explains the adjusted phenotype as a function of CNV carrier status (encoded according to the most significant CNV-GWAS model [19]), PGS_GW_, and an interaction term between the two. Similarly, we tested for an interaction between the deletion or duplication burden and PGS_GW_, defining significance based on Bonferroni correction (p ≤ 0.05/(43 × 2) = 5.8 × 10^-4^). As interactions can be scale-dependent, we recomputed linear regressions explaining raw phenotypic values as a function of CNV carrier status, PGS_GW_, an interaction term between the two, and covariates (age, age^2^, sex, genotype batch, and PCs 1-40). For significant interactions, the regression model was recomputed using a PGS*_trans_* that excludes all SNVs within ± 250 kb of the focal CNV region to ensure that the interaction was not driven by errors in genotyping caused by the presence of the overlapping CNV. Analogously, interactions between the deletion or duplication burden and PGS_GW_ on the raw phenotypic scale were assessed.

To assess our power to detect significant interactions at current sample size (N = 96,716; i.e., equivalent to our test set), we conducted simulations across a grid of CNV frequencies, q, and interaction effects sizes, *β*, that overlap empirically observed data, i.e., q and *β* varied between 1 × 10^-5^ and 1 × 10^-2^ and 0.001 and 1, respectively. For each combination of q and *β*, 1,000 simulations were performed. Specifically, CNV and PGS_GW_ genotypes were simulated for N individuals, following CNV ∼ Bernoulli(q) and PGS*_GW_* ∼ N (0, 1). These were used to model phenotypic values *Y*, as *Y* ∼ *β*(*CNV* ×*PGS_GW_*)+ɛ. We then fitted a linear regression *Y* ∼ *CNV* +*PGS* +(*CNV* × *PGS*) and deemed the interaction significant at p ≤ 0.05/119. Power was assessed as the fraction of the simulations for which the interaction term reached significance.

#### PGS_GW_ distribution among CNV carriers

For each CNV-trait pair, a linear regression model was fitted explaining PGS_GW_ as a function of CNV carrier status (encoded according to the most significant CNV-GWAS model [19]). Similarly, we fitted linear regressions explaining PGS_GW_ as a function of the CNV burden for the 21 phenotypes that were significantly (p ≤ 0.05/43 = 1.2 × 10^-3^) impacted by the latter. Significant effects were defined based on a Bonferroni correction (p ≤ 0.05/21 = 2.4 × 10^-3^). Hypothesizing that selective participation bias would induce a negative (spurious) correlation between CNV carrier status and PGS_GW_ (Fig. 1c), we assessed whether the fraction of CNV-trait pairs with a CNV effect on PGS_GW_ whose direction *aligned* with the CNV effect on the adjusted phenotype deviates from the expectation, accounting for the fact that some phenotypes are correlated. To do so, we simulated PGSs by drawing *N* samples from a multivariate normal distribution *PGS_GW_* ∼ 𝒩 (*µ*, **Σ_PGS_**) using rmvnorm from the mvtnorm v1.3.3 R package [118], with sample size *N*, mean vector *µ*, and covariance matrix **Σ_PGS_** derived from real data. CNV status were sampled as *CNV* ∼ Bernoulli(*q*) with probabilities *q* corresponding to the empirically observed CNV frequencies. Phenotypes were generated under a linear model *Y* ∼ *CNV* + *PGS* + ɛ, with errors ɛ coming from a multivariate normal distribution ɛ ∼ 𝒩 (**0**, **Σ_Y_**) with mean ^⃗^0 and empirical phenotypic covariance **Σ_Y_**. Using the simulated data, we fitted the same two models as for the real-data analyses: i) phenotype as a function of CNV status, and ii) PGS_GW_ as a function of CNV status. The proportion of the 119 CNV-trait pairs for which the CNV’s effect shows directional agreement in the two models was computed and the procedure was repeated 1,000 times. A p-value was obtained as the proportion of trials with a ratio greater than the empirical one.

#### Cistagging of CNVs by PGS

For each CNV-trait pair, using a similar approach as for PGS_GW_ (see Polygenic scores calculation), a local PGS (PGS*_cis_*) using only SNVs mapping to the CNV region ± 50 kb (PGS*_cis_*_-50kb_), ± 100 kb (PGS*_cis_*_-100kb_), ± 250 kb (PGS*_cis_*_-250kb_), or the entire chromosome (PGS*_cis_*_-chr_) were calculated. Similarly, a PGS*_trans_* was computed excluding all SNVs that map to the CNV region ± 50 kb (PGS*_trans_*_-50kb_), ± 100 kb (PGS*_trans_*_-100kb_), ± 250 kb (PGS*_trans_*_-250kb_), or the entire chromosome (PGS*_trans_*_-chr_). We next computed the Pearson correlation between the CNV carrier status (encoded according to the most significant CNV-GWAS model [19]) and various PGS*_cis_* and PGS*_trans_*. Standard errors for the Pearson correlations were computed through Fisher’s z-transformation and correlations were deemed Bonferroni-significant at p ≤ 0.05/119 = 4.2 × 10^-4^.

#### Conditioning of CNV-trait relations on PGS_GW_

For each CNV-trait pair, a linear regression model was fitted explaining the adjusted phenotype as a function of either CNV carrier status-only (model_M1_) or CNV carrier status and PGS_GW_ (model_M2_). If conditioning the CNV-trait relation on PGS_GW_ results in a significant change in the CNV-trait association, this would indicate that CNV and PGS_GW_ are not independent, supporting tagging of the CNV by PGS_GW_. Differences in effect size of the CNV carrier status were assessed using a t-statistic:

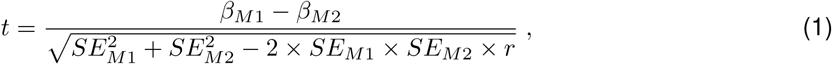

where *β* and SE are the effect size and standard error of the effect of the CNV on the adjusted phenotype estimated through either model_M1_ or model_M2_. We adjust our test statistic for the fact that *β_M_*_1_ and *β_M_*_2_ are estimated from the same data, by accounting for r, the correlation between 5,000 bootstrap estimates of *β_M_*_1_ and *β_M_*_2_. Two-sided p-values were computed in R `(2*pnorm(-abs(t), mean = 0, sd = 1))`.

### Assortative mating

#### Phenotypic similarity within couples

Couples were defined as “white British” individuals in households with two unrelated, opposite-sex in-dividuals who identified as “husband, partner or wife” (#6141) [76]. Of the 51,664 identified couples, 3,396 have both partners in the test sample set. Focusing on the latter subset, same-trait and cross-trait phenotypic similarity among couples was assessed through Pearson correlation across 43 adjusted phenotypic measurements (see Phenotype selection), with significance defined through Bonferroni correction (p ≤ 0.05/43^2^ = 2.7 × 10^-5^). To ensure that downsampling did not impact the accuracy of our estimates, we compared same-trait phenotypic correlations computed from the test set to those obtained from the 51,664 couples. Furthermore, we compared estimates of within-couple phenotypic correlation to those obtained using a stricter definition of couples [81, 119], requiring individuals to have matching home location East and North coordinates (#22702 and #22704), Townsend deprivation index (#22189), number of smokers in household (#1259), number in household (#709), length of time at the current address (#699), average total household income before tax (#738), and living with husband or wife or partner (#6141). Couples were further required to be of opposite sex, of “white British” ancestry, unrelated, and have partners appear in a single couple, resulting in 22,068 couples, among which 1,213 had both partners in our test set.

#### Assortative mating modeling

We explored assortative mating as a potential mechanism to explain positive CNV-PGS correlation by comparing the total CNV-PGS*_trans_*_-250kb_ correlation to an indirect path mediated by assortment, defined as:

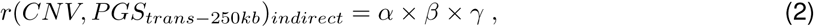

where *α* is the Pearson correlation between the adjusted phenotype and CNV carrier status (encoded by the most significant CNV-GWAS model [19]), *β* is the within-couple Pearson correlation of adjusted phenotypic values, and *γ* is the Pearson correlation between the adjusted phenotype and PGS*_trans_*_-250kb_ (Fig. 1c). By using PGS*_trans_*_-250kb_ instead of PGS_GW_, we can ensure that our framework does not model local linkage disequilibrium. CIs for the total CNV-PGS*_trans_*_-250kb_ correlation were computed through Fisher’s z-transformation. SE of α, *β*, and *γ* were estimated using Bonett’s approximation 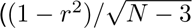, where *N* corresponds to the sample size). For the indirect path, the SE was derived from the variance of the product of independent variables as:

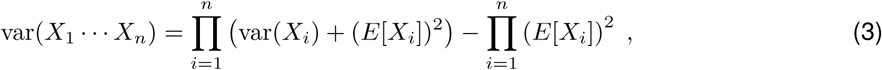

where 95%-CIs were then obtained from propagated errors.

#### Significance assessment

We first compared the calculated total-indirect CNV-PGS*_trans_*_-250kb_ with two competing hypotheses: a null scenario in which the true correlation is zero (cor.test() in R), and a scenario in which our data is better explained by selective participation. To test the latter, we computed the correlation between the total CNV-PGS*_trans_*_-250kb_ correlation and −*α* × *γ*, which is proportional to the bias induced by selective participation. In a second time, significance of the indirect path through assortative mating was assessed by comparing the total-indirect Pearson correlation to a set of three null distributions derived from 50,000 permutations, in which one of α, *β*, or *γ* was randomized. For *α* and γ, these were obtained by sampling without replacement, so that each phenotype was paired with a random CNV status or PGS*_trans_*_-250kb_, respectively, across the 119 CNV-trait pairs. For *β*, within-couple phenotypic correlation was recomputed after randomly repairing couples without replacement. The product of *α* × *β* × *γ* was recomputed, maintaining the empirical values for the two other coefficients. Finally, the correlation between the total and indirect CNV-PGS*_trans_*_-250kb_ correlations was computed and p-values were calculated as the proportion of permutations yielding as or more extreme correlations than the empirical one.

#### Sensitivity analyses for assortative mating modeling

We conducted a series of followup analyses to ensure that our results were robust to analytical variations. First, we repeated the analysis estimating *α* and *γ* excluding couples used to calculate *β*. Second, we repeated the analysis adjusting phenotypes for sex × covariates interactions. Third, to account for residual correlation between phenotypes and geographic location, we included participants’ home location East and North coordinates at 1 km resolution (#22702 and #22704) as additional covariates when correcting phenotypic values and repeated the analysis. Fourth, Winner’s curse could affect the CNV-trait pairs that were retained for this study, leading to an overestimation of the CNV-phenotype correlation, α. To ensure this was not the case, we repeated the analysis focusing on 98 CNV-trait pairs that in the original study would survive a stricter significance threshold of p ≤ 0.05/(11,804 × 3), which accounts for the three tested association models [19].

From a theoretical perspective, to assess whether the observed CNV-PGS correlation could arise solely from algebraic constraints, we first obtained bounds for *r*(*CNV*, *PGS_GW_*) based on the positive semidefiniteness of the full correlation matrix (comprising phenotype, *CNV*, and *PGS*_GW_). These limits define the range of allowed *r*(*CNV*, *PGS_GW_*) values under the assumed causal structure (Supplementary Note 1). We then derived an analytical formula and conducted simulations showing that under the assortative mating model assumptions, the CNV-PGS_GW_ correlation corresponds to the product of *α* × *β* × *γ* (Supplementary Notes 2 and 3).

Finally, we repeated the assortative mating framework (see Assortative mating modeling) at the phenotypic level for 21 traits that were significantly (p ≤ 0.05/43 = 1.2 × 10^-3^) impacted by the CNV burden (see CNV carrier identification & CNV burden calculation).

### CNV inheritance rate estimation

#### Sample selection

We used KING [120] to identify related UKBB participants (≤ 3^rd^ degree; kinship > 0.044). Among the 67,958 pairs of relatives (103,726 individuals) in which at least one individual carried a high-confidence CNV, 12,831 pairs (21,216 individuals) harbored a CNV that overlaps (≥ 1 bp) one of the 27 assessed CNV regions (see CNV carrier identification & CNV burden calculation).

#### Identity-by-descent estimation

For each CNV carrier with a close relative, we estimated the probability that the region harboring the CNV is identity-by-descent (IBD) with its relative, using THORIN [121, 122]. Namely, average IBD probability *IBD_avg_* within a 1 Mb window around the CNV was computed by summing per-SNV IBD probability on each haplotype. We classified the IBD status of the CNV carrier and its relative as either *IBD*0 (i.e., no shared haplotypes; *IBD_avg_* < 0.25), IBD1 (i.e., one shared haplotype; 0.75 < *IBD_avg_* < 1.25), *IBD*2 (i.e., two shared haplotypes; *IBD_avg_* > 1.75). IBD status was set to unknown for all other probabilities (]0.25, 0.75[∪]1.25, 1.75[).

#### CNV sharing among relatives

Accurate CNV breakpoint estimation is notoriously challenging from microarray data, so that even inherited CNVs could be detected with different breakpoints. Relatives were considered to carry the same CNV (*CNV* 2) if they harbor a CNV of the same type showing over 50% reciprocal overlap. Otherwise, the pair is considered to have one CNV carrier (CNV 1).

#### Inheritance rate estimation

We combined information on IBD and CNV sharing between two relatives to classify our samples:

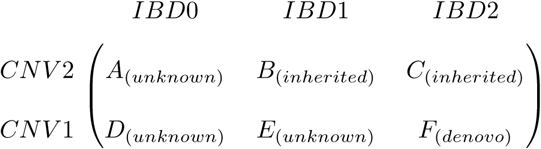

where *IBD*0, *IBD*1, or *IBD*2 indicate whether none, one, or two haplotypes are shared, respectively; *CNV* 1 or *CNV* 2 indicate whether one or both individuals in a relative pair carry the same CNV, respectively. Specifically, *B* and *C* represent cases where the CNV is likely to be inherited, *F* represents cases where the CNV is likely to be *de novo* (or a false positive), while *A*, *D*, and *E*, represent cases where we do not have sufficient data to infer the inheritance status. Based on this, we estimated the average CNV inheritance rate as:

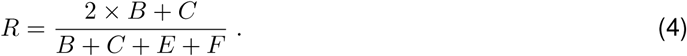

We estimated the standard error *SE*(*R*) of the inheritance rate using parametric simulations from binomial distributions. For each locus *i*, we used the counts of correctly assigned *IBD*1 and *IBD*2 segments (*B* and *C*, respectively) and the total number of individuals in IBD1 and IBD2 status (*n*_1_ = *B* + *E* and *n*_2_ = *C* + *F*, respectively). We estimated inheritance rates separately for each IBD status:

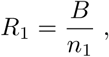

and

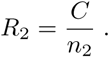

We then sampled 1,000 replicates of B and C from a binomial distribution:

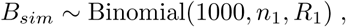

and

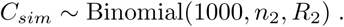

We computed the simulated inheritance rate as:

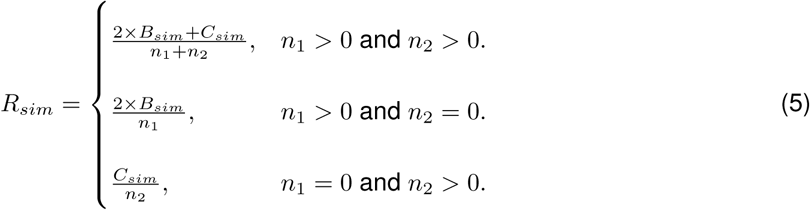

and estimated the standard error of the inheritance rate *SE*(*R*) as the standard deviation of *R_sim_*. The pooled inheritance rate estimate and its standard error for duplications and deletions was calculated using inverse-variance weighted metaanalysis for testable CNVs (i.e., CNVs with ≥ 20 carriers with a ≤ 3^rd^ degree relative who shares *IBD*1 or *IBD*2) with a non-null inheritance rate.

### pLoF burden analyses

Linear regression models explaining adjusted phenotypes as a function of pLoF burden identified 15 traits subject to modulation by the latter among unrelated “white British” samples with available exome sequencing data (p ≤ 0.05/43 = 1.2 × 10^-3^). For these traits, linear regression models explaining PGS_GW_ as a function of pLoF burden were fitted, with significance defined based on Bonferroni correction (p ≤ 0.05/15 = 3.3 × 10^-3^). Pearson correlation between the CNV and the pLoF burdens was assessed. To assess our power to detect a correlation between the CNV and pLoF burdens, we performed 1,000 simulation trials across a grid of correlations, *r*, centered on the empirically observed correlation (*r* = 0.005) and ranging from 0.0005 to 0.01 with a step size of 0.0005. In each trial, two samples of 319,161 individuals (i.e., the size of the unrelated “white British” UKBB participants with available exome sequencing data) were drawn from a bivariate normal distribution with means (0, 0), unit variance and a Pearson correlation *r*. After selecting the most appropriate distribution to represent each burden type, we rank-transformed the sample describing the CNV burden to follow a negative binomial distribution with mean *µ_CNV_* = 1.51 and size = 0.18. The second sample, representing the pLoF burden, was rank-transformed to match a normal distribution with mean *µ_pLoF_* = 13.71 and standard deviation *σ* = 3.78. Parameters of these distributions were estimated by fitting negative binomial (CNV burden) or normal (pLoF burden) models to our data (filtered for “white British” UKBB participants) `using fitdistr()` from the MASS v7.3.65 R package [123]. The Pearson correlations between these two samples were computed, and the corresponding power was estimated using `wp.correlation()` from the WebPower v0.9.4 R package [124] at a significance level of *α* = 0.05. For each value of *r*, we averaged correlation and power across 1,000 trials, and reported the average power at the empirically observed correlation (*r* = 0.005). We further explored how the discrepancy between true and measured correlation was affected by the CNV burden frequency by repeating the previous procedure with a true Pearson correlation fixed at *r* = 1 and varying CNV burden mean (*µ_CNV_*; ranging from 0.1 to 1 in 0.1 step increments and 1 to 10 in unit step increments).

## Data Availability

Data used in this study is publicly available in tabular format. The code to reproduce analyses is accessible on Github.

https://github.com/cauwerx/CNV_PGS_interaction

## Data and code Availability

Data used in this study are publicly available, as described in the Methods. Code to reproduce analyses is available at https://github.com/cauwerx/CNV_PGS_interaction.

## Acknowledgments

We thank UKBB participants for sharing their data. Computations were carried out on the Urblauna High-Performance Computing server from the University of Lausanne and the UK Biobank Research Analysis Platform. This work was supported by funding from the Department of Computational Biology (ZK) and the Center for Integrative Genomics (AR) from the University of Lausanne, as well as grants from the Swiss National Science Foundation (P500-3 235131 to C.A.; 31003A 182632 and IZSTZ0 216615 to AR) and Horizon2020 Twinning projects (ePerMed 692145, AR). The funders had no role in study design, data collection and analysis, decision to publish, or preparation of the manuscript.

## Author contributions

C.C., C.A., A.R., and Z.K. conceived and designed the study; C.C. and C.A. performed the main analyses; R.H. and T.C. derived the PGS weights; R.H. estimated the *de novo* rate of CNVs; T.S. provided guidance for assortative mating analyses; Z.K. supervised all statistical analyses; C.C. and C.A. drafted, and A.R. and Z.K. made critical revisions to the manuscript; All authors read, approved, and provided feedback on the final manuscript.

## Supplementary information

Supplementary Figures 1-11.

Supplementary Notes 1-3.

Supplementary Tables 1-16.

## Supplementary Figures

**Figure S1.**
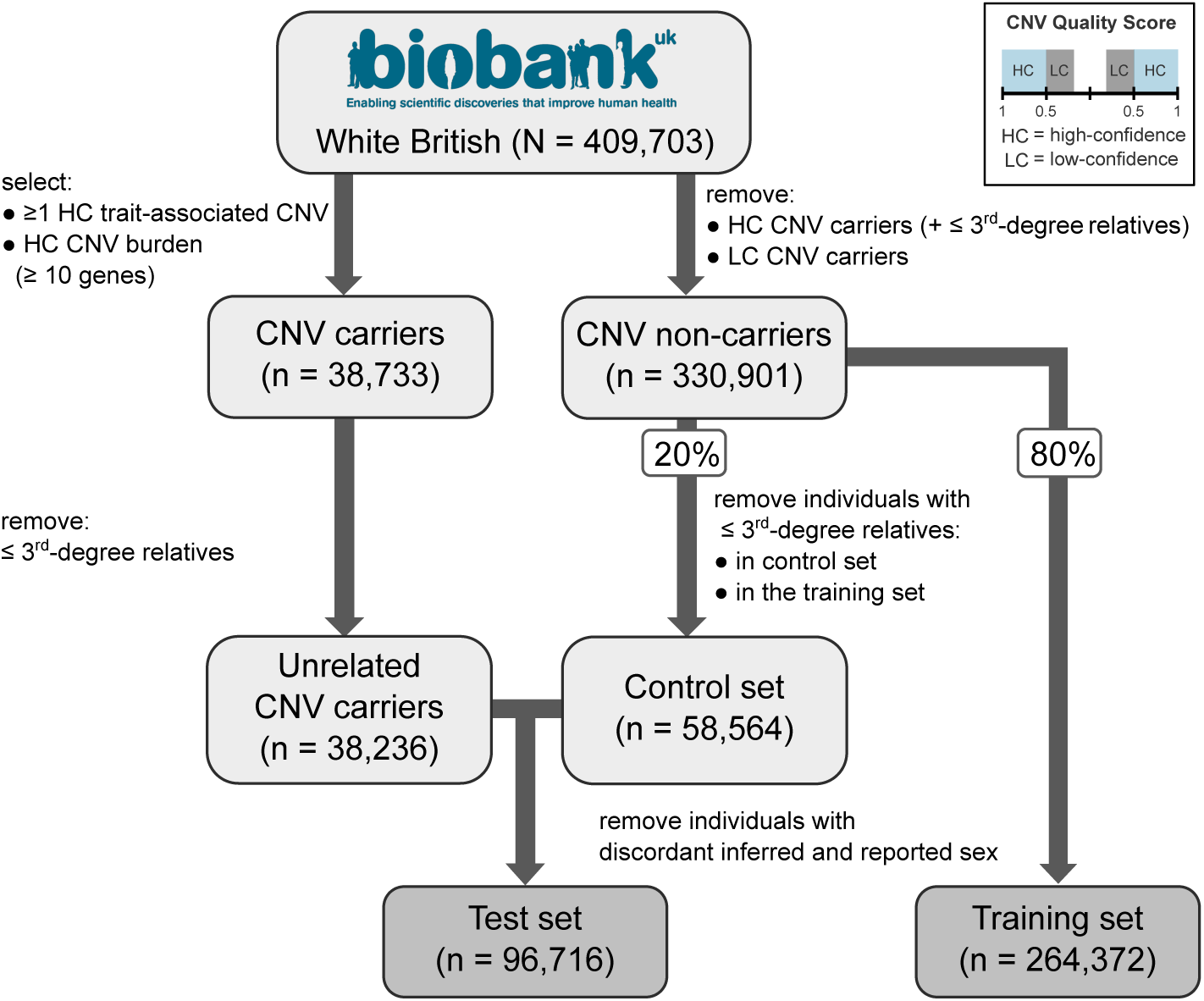
Study design. Schematic representation of the study design. We split 409,703 “white British” UK Biobank samples into a training set used to develop polygenic score (PGS) weights, and a CNV-carrier-enriched test set used to perform analyses. Individuals carrying a high-confidence (HC) CNV call (absolute value of quality score: *≥* 0.5) overlapping the lead probe of one of the assessed CNV-trait pairs or a high CNV burden (*≥* 10 genes) were pre-selected for the test set and related individuals were pruned. These individuals and all their relatives were excluded from the 409,703 “white British” samples, along with individuals carrying a lower-confidence (LC) CNV call (absolute value of quality score: 0.2-0.5). Remaining individuals were randomly split into training (80%) and control (20%) sets with matching age and sex distributions. The control set was pruned to unrelated individuals, samples related to samples in the training set were excluded, and remaining individuals were merged with pre-selected CNV carriers to form the test set. Finally, individuals with mismatched reported versus genetically inferred sex were excluded.

**Figure S2.**
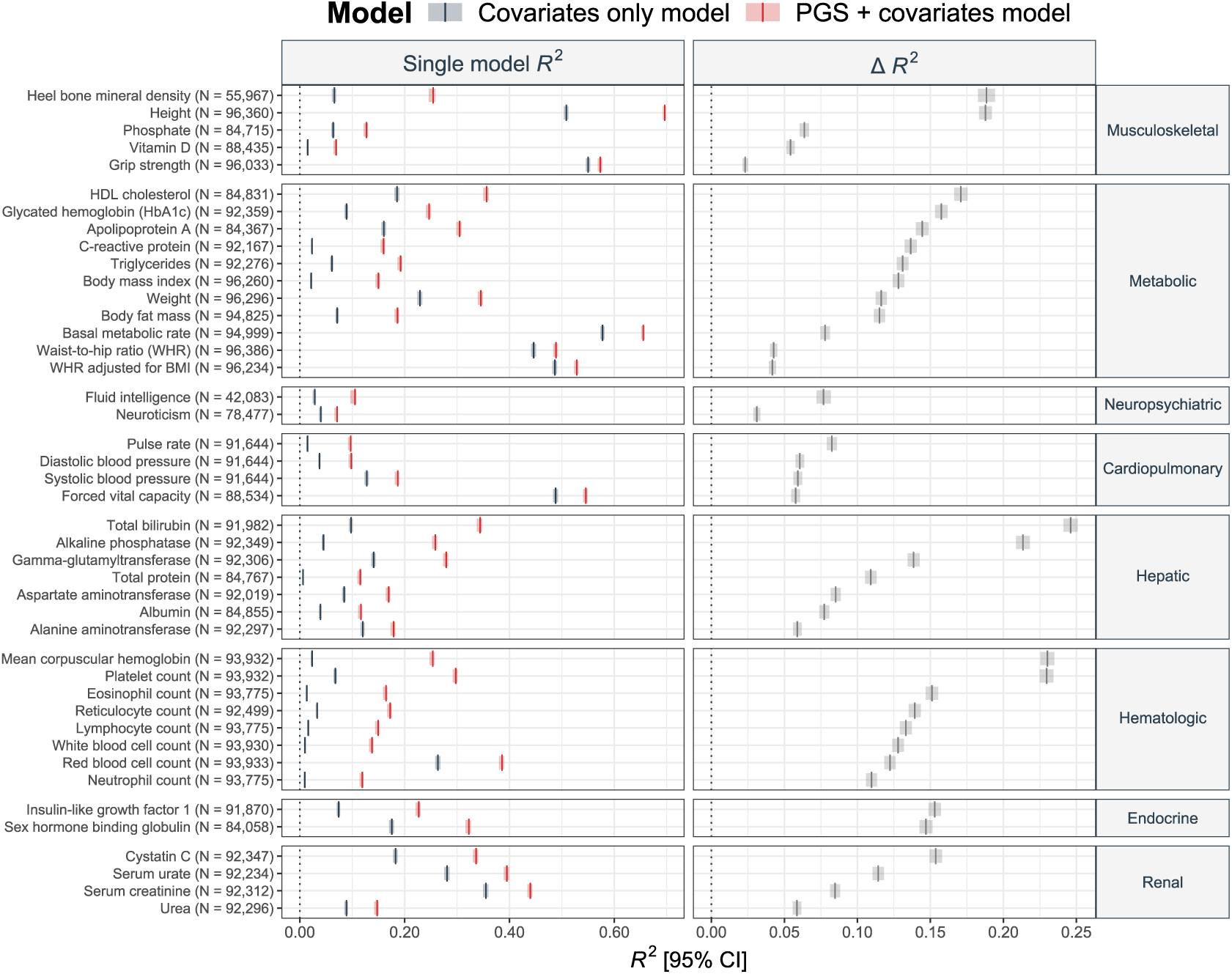
Phenotypic variance explained by PGSGW. The left panel shows the *R*^2^ (x-axis) with 95% confidence interval (CI) of a linear regression explaining adjusted phenotypic values for the 43 traits assessed in this study (y-axis, left) as a function of covariates only (i.e., sex, age, age2, genotyping batch, and 40 principal components; in grey) or covariates and genome-wide polygenic score (PGSGW; in red). The right panel depicts the difference in *R*^2^ between these two models (Δ*R*^2^), with 95%-CI. Phenotypes are ordered by decreasing Δ*R*^2^ within each phenotype category (y-axis, right) and the number of samples in the test set used to fit the model is indicated as N (y-axis, left).

**Figure S3.**
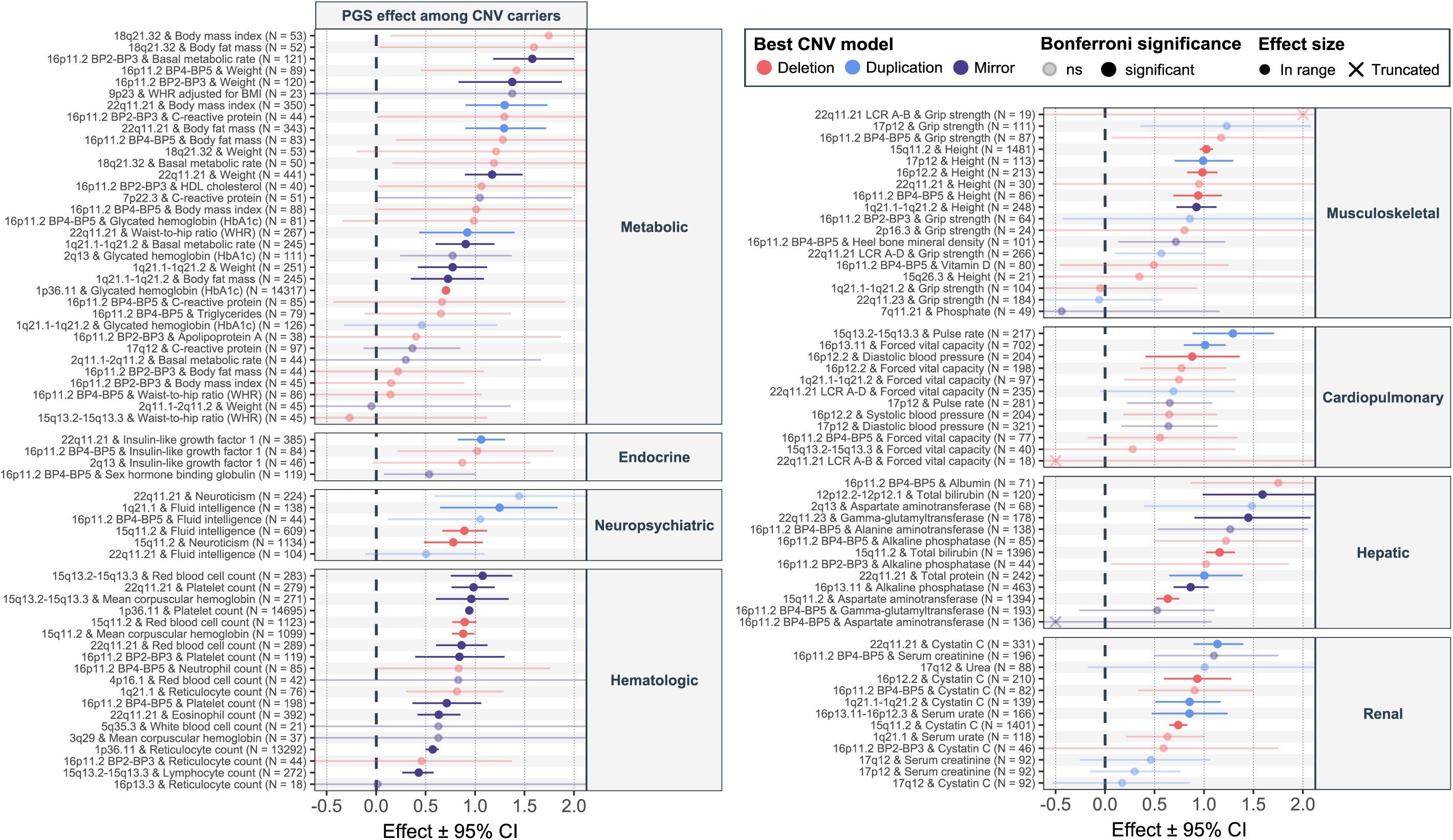
PGSGW effect among CNV carriers. Standardized effect sizes with 95% confidence intervals (CI) of the PGSGW effect on the phenotype (x-axis) among CNV carriers for the 119 unique CNV-trait pairs assessed in this study (y-axis, left ordered by decreasing effect size within each trait categories (y-axis, right). PGSGW effects were assessed among deletion (red), duplication (blue), or CNV (purple) carriers depending on whether the CNV-trait association was previously found to be most significant according to a deletion-only, duplication-only, or mirror model, respectively in Auwerx *et al.*, 2022. The number of considered carriers is indicated as N (y-axis, left). Transparency indicates whether the effect is significant under Bonferroni correction (p *≤* 0.05/119 = 4.2 *×* 10^-4^_)_ or not. Some non-significant effects were truncated to improve readability and are indicated with a cross instead of a circle.

**Figure S4.**
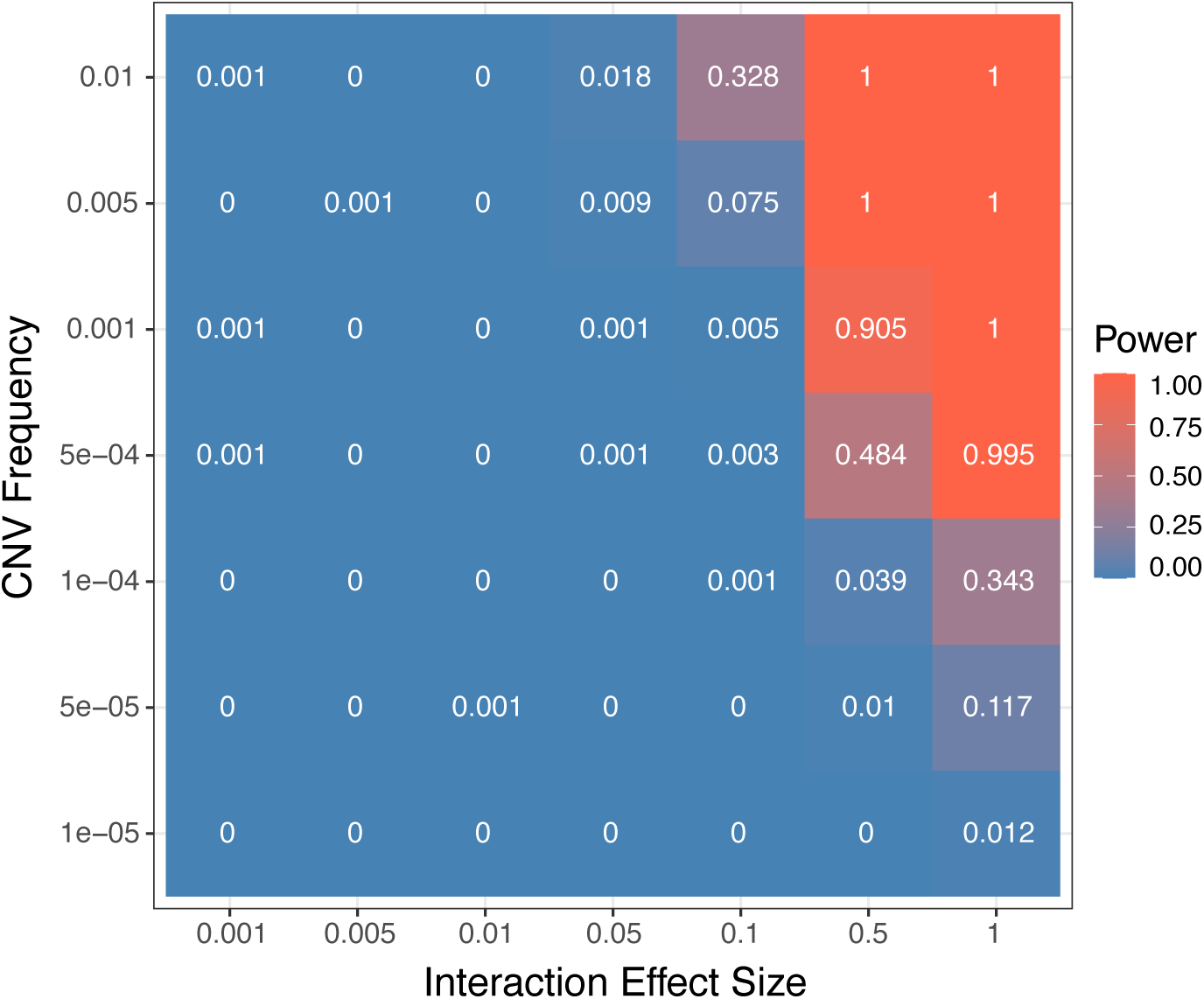
Power to detect CNV *×* PGS interactions. Simulations were conducted across a grid of CNV frequencies (y-axis) and interaction effect sizes (x-axis; absolute values) using a sample size equivalent to our test set (N = 96,716). Color represents the power to detect an interaction of a given effect size at a specific CNV frequency based on 1,000 simulations.

**Figure S5.**
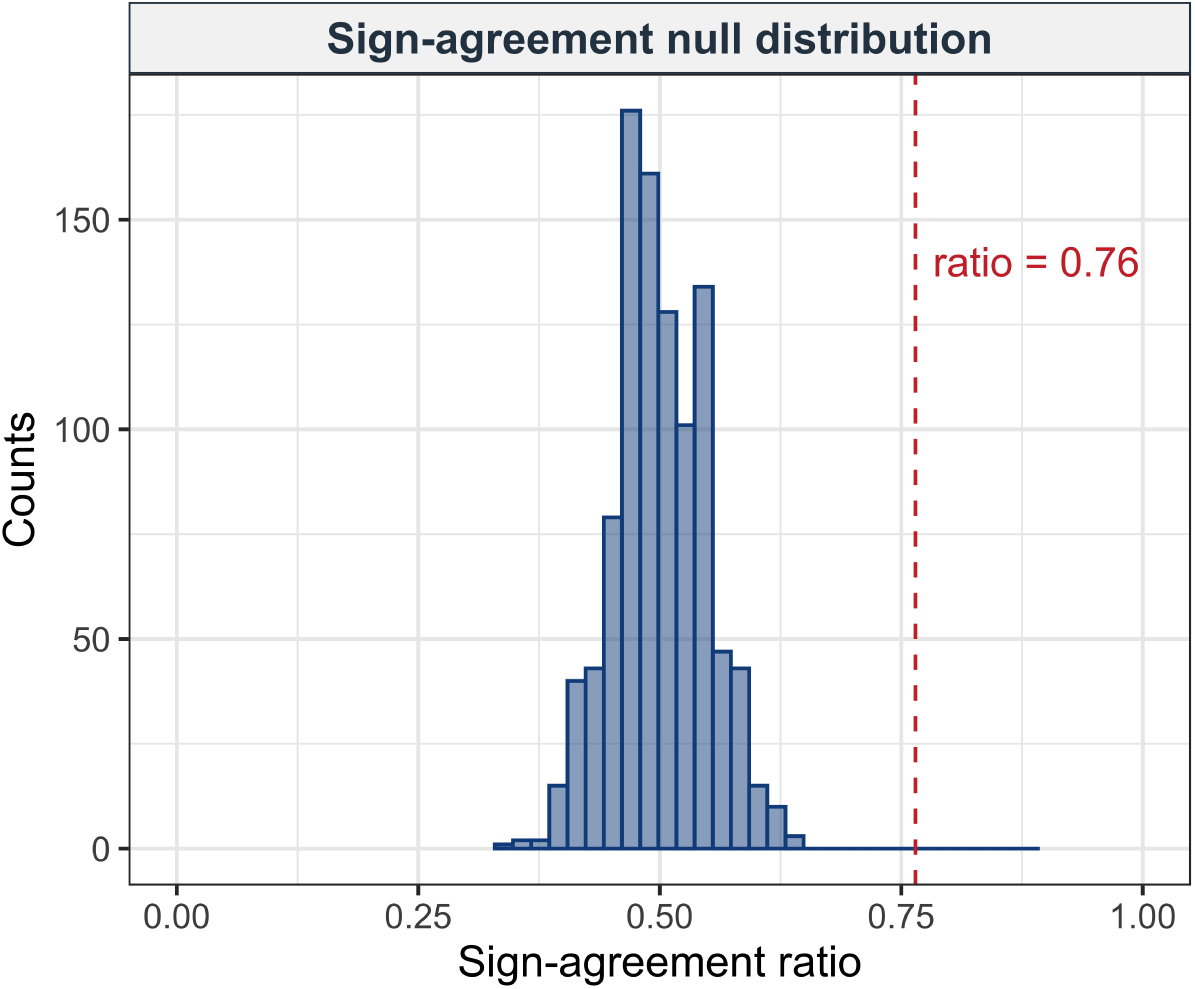
Directional concordance of CNV effects on phenotypes and PGSGW. Histogram showing the null distribution of CNV effects on phenotype and PGSGW sign agreement, accounting for the PGSGW and phenotype covariance structure across all 43 traits. The empirical sign-agreement ratio is indicated by the dashed red vertical dashed line.

**Figure S6.**
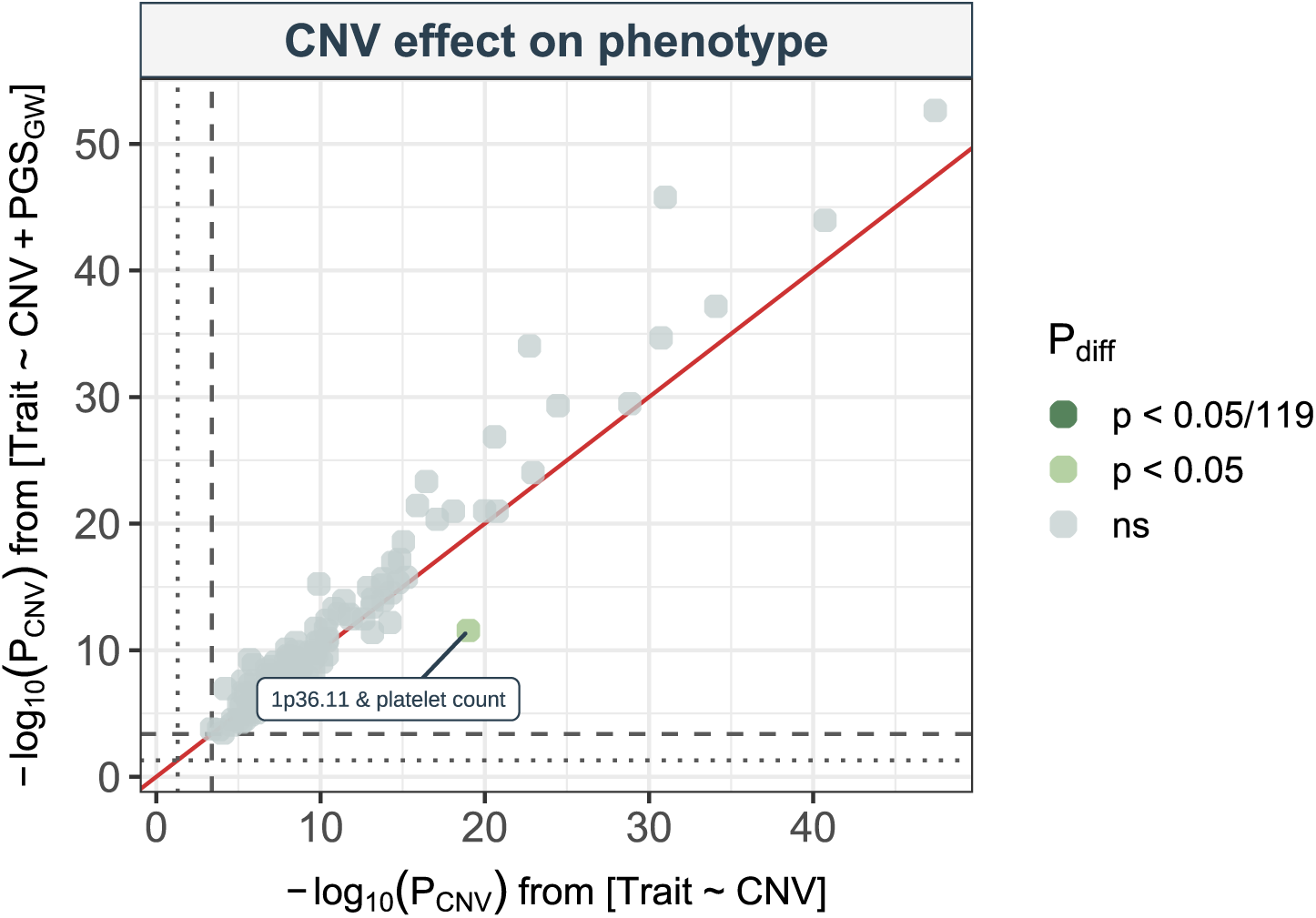
Conditioning of the CNV-phenotype relation on PGSGW. **c**, Negative logarithm of the p-value of the effect of the CNV on adjusted phenotypes, when conditioning on PGSGW (y-axis) or not (x-axis). Data points are colored according to the significance degree of a t-test assessing whether the CNV effect on the adjusted phenotype is different in the two compared models, with “ns” indicating p *>* 0.05. The single nominally significant effect is labeled. The red diagonal represents the identity, while the grey dashed and dotted lines mark the threshold for Bonferroni (-log10(0.05/119) = 3.38) and nominal (-log10(0.05) = 1.30) significance, respectively.

**Figure S7.**
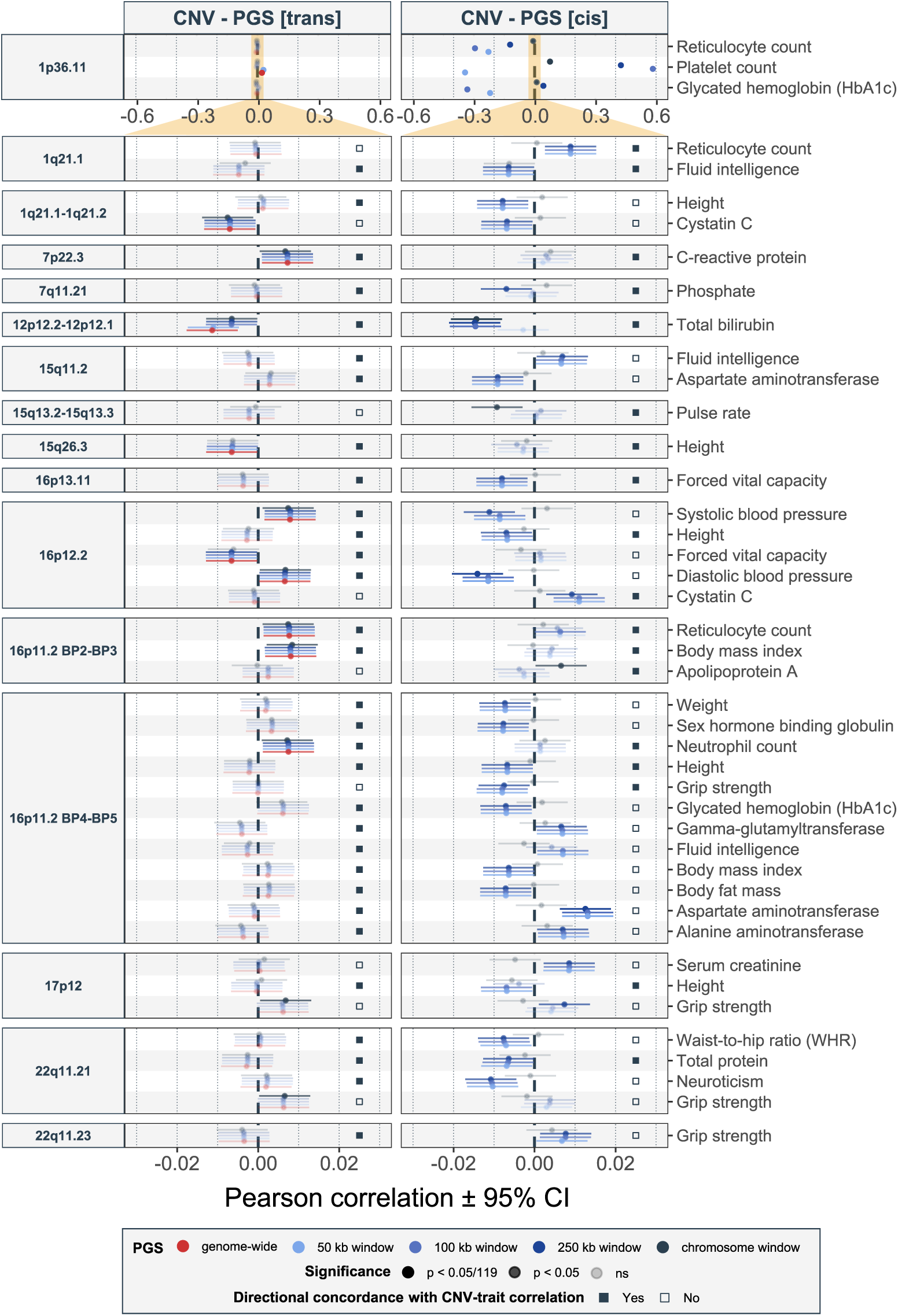
Partial PGS-CNV correlations. Pearson correlation with 95% confidence interval (CI; x-axis) between polygenic scores (PGSs; y-axis, right) and CNV carrier status (y-axis, left) for all CNV-trait pairs with at least one nominally significant CNV-PGSGW, CNV-PGS*trans*, or CNV-PGS*cis* correlation. The left panel shows correlation with the genome-wide PGSGW (red) and various PGS*trans* (blue shades), while the right panel shows correlations with various PGS*cis* (blue shades). Transparency reflects the significance level, with “ns” indicating p *>* 0.05. The small grey square indicates whether the correlation between the CNV carrier status and PGS*trans*-250kb (left panel) or PGS*cis*-250kb (right panel) are directionally concordant with the CNV’s effect on the trait. We use a different x-axis scale for the 1p36.11 CNV region for better visualization (orange area).

**Figure S8.**
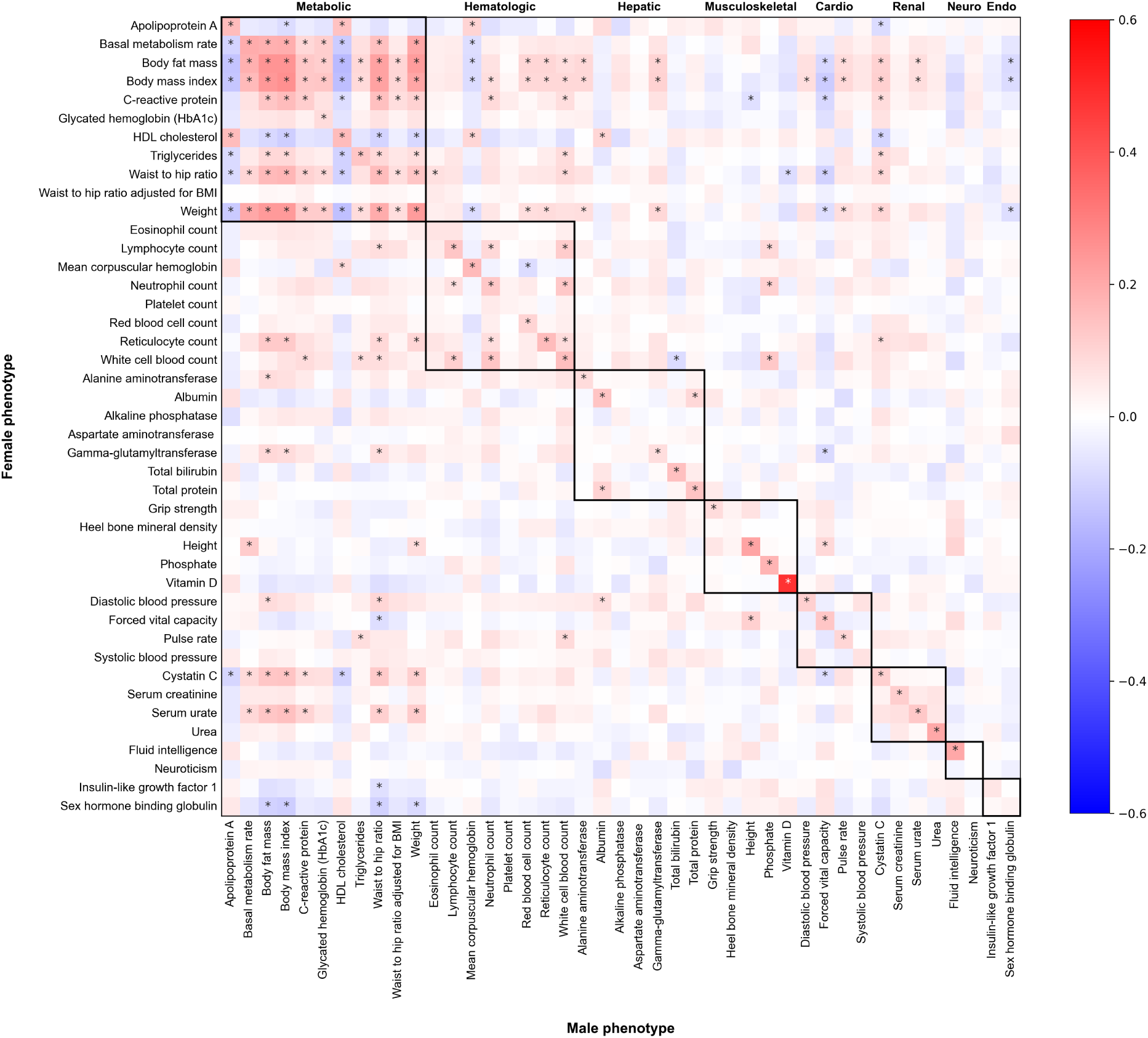
Pairwise partner phenotypic correlation. Heatmap of Pearson correlation coefficients between 43 adjusted phenotypes in males (x-axis) versus the corresponding phenotype in their female partner (y-axis). Stars indicate Bonferroni-significant correlations (p *≤* 0.05/43^2^ = 2.7 *×* 10^-5^). Phenotypes are clustered by trait categories, as depicted by bolded squares and top labels. Cardio = Cardiometabolic; Neuro = Neuropsychiatric; Endo = Endocrine.

**Figure S9.**
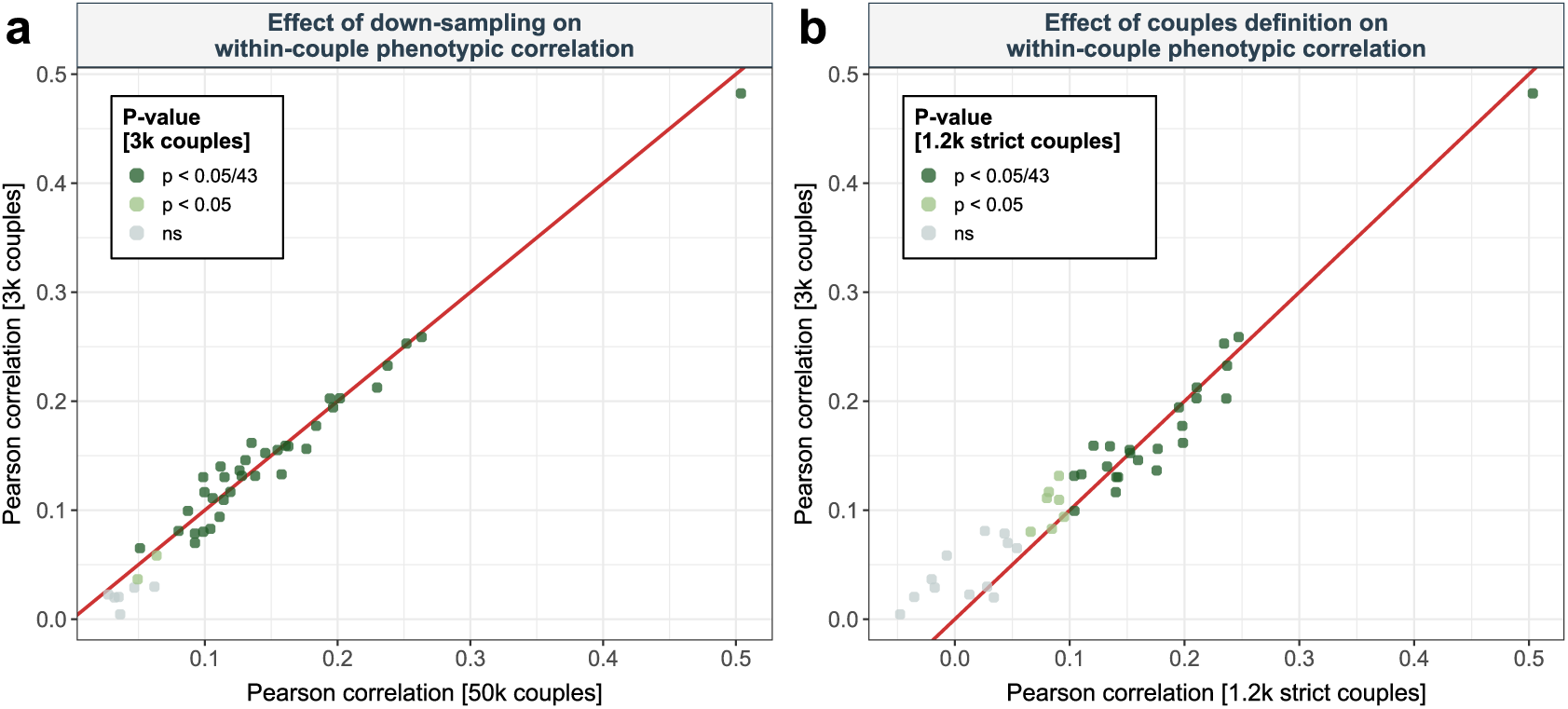
Impact of couples selection on same-trait within-couple phenotypic correlation. **a**, Impact of down-sampling: Same-trait, within-couple Pearson phenotypic correlation among the 3,396 couples present in our testing subset (y-axis, [3k]) against same-trait, within-couple Pearson phenotypic correlation among the 51,664 couples present in the entire “white British” subset of the UKBB (x-axis, [50k]). All phenotypes show Bonferroni-significant (p *≤* 0.05/43 = 1.2 *×* 10^-3^) within-couple correlation among the 50k couple subset and data points are colored according to the significance level of the within-couple phenotypic correlation among the 3k subset, with “ns” indicating p *>* 0.05. The red diagonal represents the identity line. **b**, Impact of stricter couples definition: Same-trait, within-couple Pearson phenotypic correlation among the 3,396 couples defined based on our original couples definition (y-axis, [3k]) against same-trait, within-couple Pearson phenotypic correlation among the 1,213 couples defined according to a strict couples definition present in our test subset (x-axis, [1.2k strict]). Data points are colored according to the significance level of the within-couple phenotypic correlation based on the strict couples definition, with “ns” indicating p *>* 0.05. The red diagonal represents the identity line.

**Figure S10.**
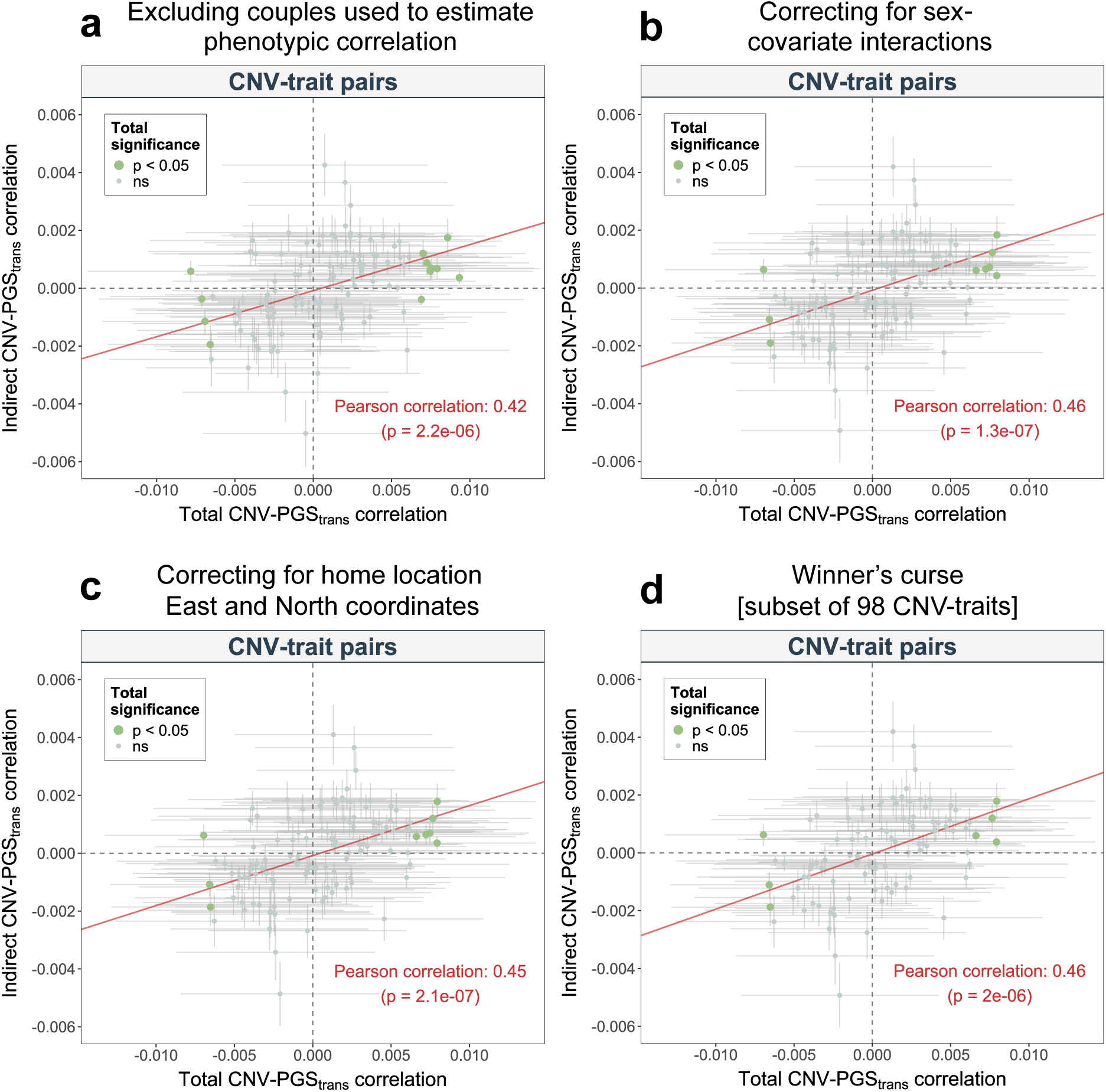
Assortative mating sensitivity analyses. Sensitivity analyses probing the robustness of the correlation between total (x-axis) and indirect (y-axis; through assortative mating) CNV-PGS*trans*-250kb correlation. **a**, Excluding the couples used for *β* estimation (i.e., within-couple phenotype correlation) from the estimation of *α* and *γ*. **b**, Using phenotypic values corrected for sex *×* covariate interactions. **c**, Using phenotypic values corrected for home location East and North coordinates (1 km resolution). **d**, Restricting the analysis to 98 CNV-trait pairs with p *≤* 0.05/(11,804 *×* 3) in the original CNV-GWAS study (Auwerx *et al.*, 2022) to assess the impact of Winner’s curse. Error bars represent 95% confidence intervals. Size and color reflect the significance level of the total CNV-PGS*trans*-250kb correlation, with “ns” indicating p *>* 0.05. The red line represents the fitted linear regression of the indirect CNV-PGS*trans*-250kb correlation against the total CNV-PGS*trans*-250kb correlation.

**Figure S11.**
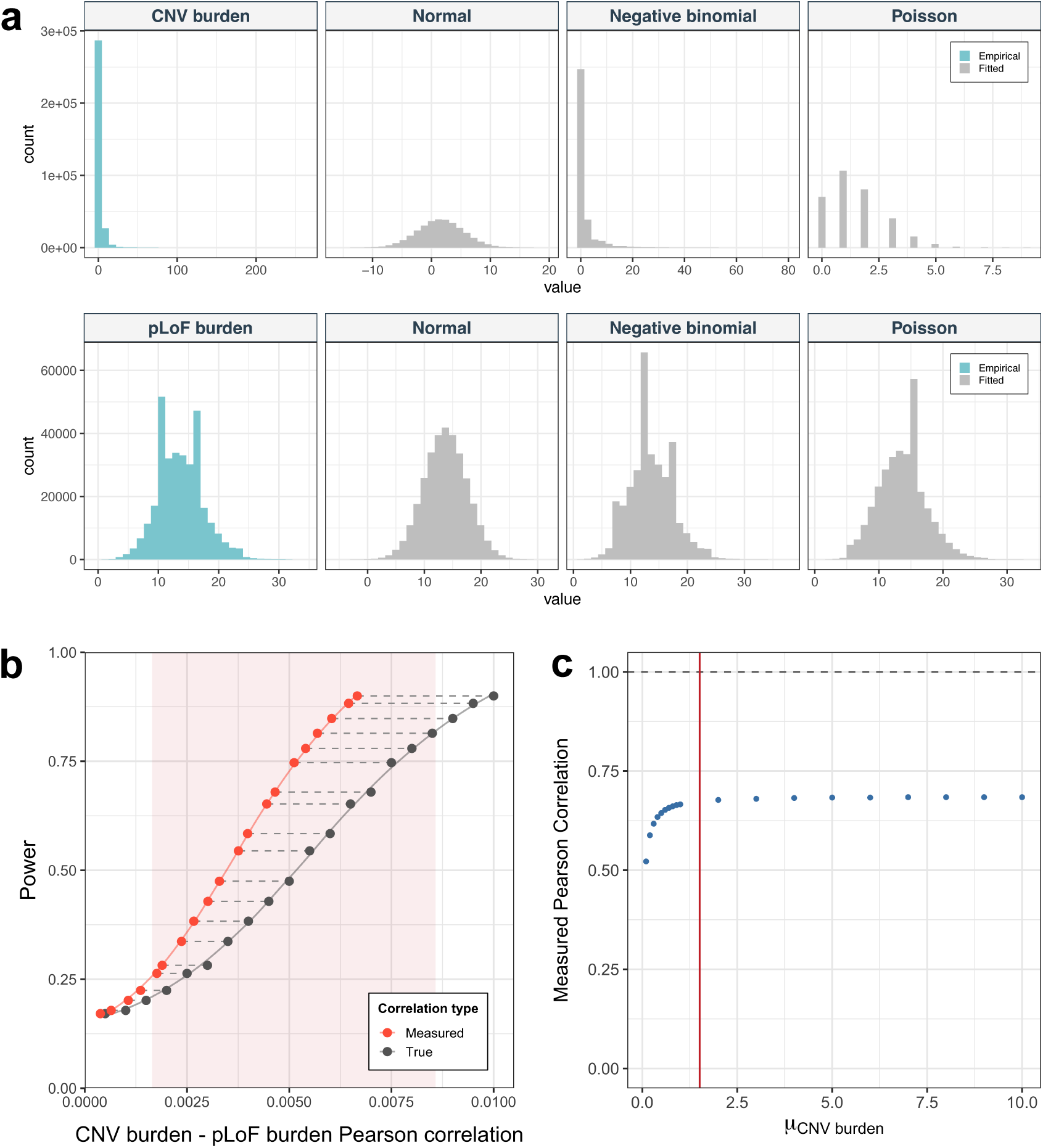
Power to detect correlation between CNV burden and pLoF burden. **a**, Normal, negative binomial, and Poisson distributions (grey) were fitted to the observed (blue) CNV burden (top row) and pLoF burden (bottom row) by maximum likelihood parameter estimation. **b**, Power (y-axis) as a function of the Pearson correlation (x-axis) between simulated CNV burden and pLoF burden. True correlations are shown in grey and measured correlations in red, with dashed lines connecting corresponding values. CNV burden and pLoF burden were generated from a bivariate distribution with a given correlation *r* (i.e., “true correlation”) and rank-transformed to match empirical negative binomial and normal distributions, respectively. The shaded region corresponds to the 95% confidence interval centered on the empirically observed correlation in our data (*r* = 0.005). **c**, CNV burden and pLoF burden were generated from a bivariate normal distribution with fixed correlation *r* = 1 and rank-transformed to approximate negative binomial and normal distributions with means *µCNV* and *µpLoF*, respectively. Correlation strength was measured across a grid of *µCNV* values, with remaining parameters *sizeCNV* and *σpLoF* held constant at their empirical values. The red vertical line depicts the empirical *µCNV* = 1.51.

## Supplementary Notes

### Supplementary Note 1

#### Theoretical limits of the CNV-PGS_GW_ correlation

Given the following causal structure between CNV, PGS_GW_, and phenotype (Fig. N1.1):

**Figure N1.1.**
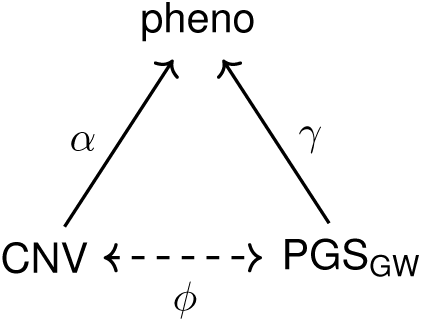
Assumed CNV-PGSGW relationship via their influence on the phenotype. Directed acyclic graph (DAG) in which the solid arrows depict the causal effects of a copy-number variant (CNV) and the genome-wide polygenic score (PGSGW) on phenotype (pheno) for a single individual, represented by correlations *α* = *r*(*CNV, pheno*) and *γ* = *r*(*PGSGW, pheno*), respectively. The bidirectional dashed arrow denotes the observed correlation *ϕ* = *r*(*CNV, PGSGW*) between CNV and PGSGW.

Since correlation matrices are positive semi-definite (PSD, i.e., they are symmetric and all principal minors are non-negative), we can obtain an expression for the boundaries of the correlation coefficients. Consider the associated correlation matrix:

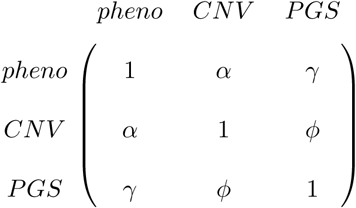

where *α*, *γ*, and *ϕ* are bounded between [-1,1]. Since all eigenvalues must be ≥ 0, the determinant is also ≥ 0:

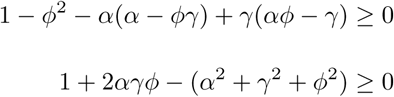

Solving the inequality for *ϕ* gives:

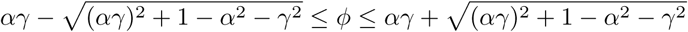

This relationship, as established by [125], imposes a lower and upper bound for ϕ, when both PGS_GW_ and CNV impact the trait of a single individual.

We next extend this logic to an assortative mating scenario, where CNV affects the phenotype of one partner and PGS_GW_ the phenotype of the other, as depicted in Fig. N1.2:

**Figure N1.2.**
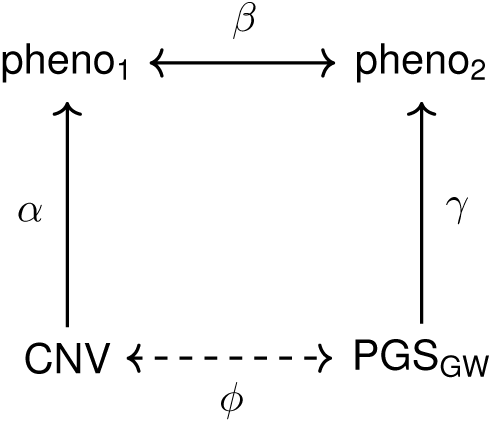
Assumed CNV-PGSGW relationship via assortative mating. DAG in which the solid arrows depict the causal effects of copy-number variants (CNV) and the genome-wide polygenic score (PGSGW) on the phenotypes of the two partners (pheno1 and pheno2), represented by correlations *α* = *r*(*CNV, pheno*1) and *γ* = *r*(*PGSGW, pheno*2), respectively. The solid bidirectional arrow between the phenotypes indicates the within-couple, same-trait, phenotypic correlation, *β*. The dashed bidirectional arrow denotes the observed correlation *ϕ* = *r*(*CNV, PGSGW*).

Assuming that CNV-pheno_2_ and PGS_GW_-pheno_1_ have no direct relationship, only through pheno_1_ and pheno_2_, their correlation can be approximated using path-tracing rules, leading to the correlation matrix A:

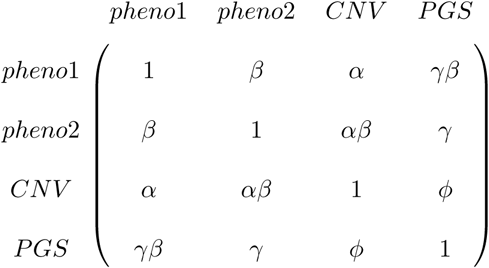

Computing all 15 principal minors M = det(A*_I_*), where A*_I_* is generated from A by selecting the rows and columns in each subset I ⊆ {1, 2, 3, 4} (Table N1), we can once again obtain boundaries for ϕ from the restriction that *M*_10_, *M*_13_, *M*_14_ and *M*_15_ should all be ≥ 0.

We thus obtain the following bounds for the total CNV-PGS_GW_ correlation (ϕ):

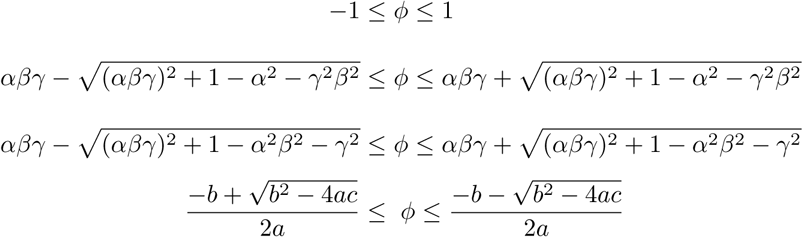

Where *a* = *β*^2^ − 1, *b* = 2*αβγ*(1 − *β*^2^), and *c* = 1 − (*α*^2^ + *β*^2^ + *γ*^2^) + (*α*^2^*β*^2^ + *α*^2^*γ*^2^ + *β*^2^*γ*^2^) − 2*α*^2^*β*^2^*γ*^2^ + *α*^2^*β*^4^*γ*^2^.

The most strict bounds are given by: *ϕ ∈* [*max(L*_10_*, L*_13_*, L*_14_*, L*_15_*), min(U*_10_*, U*_13_*, U*_14_*, U*_15_)] where L*_i_* and U*_i_* correspond to the the lower and upper bounds obtained from principal minor M*_i_*. We can then assess whether the correlation between the indirect and total CNV-PGS_GW_ correlation is expected within this mathematical constraint.

**Table N1.**
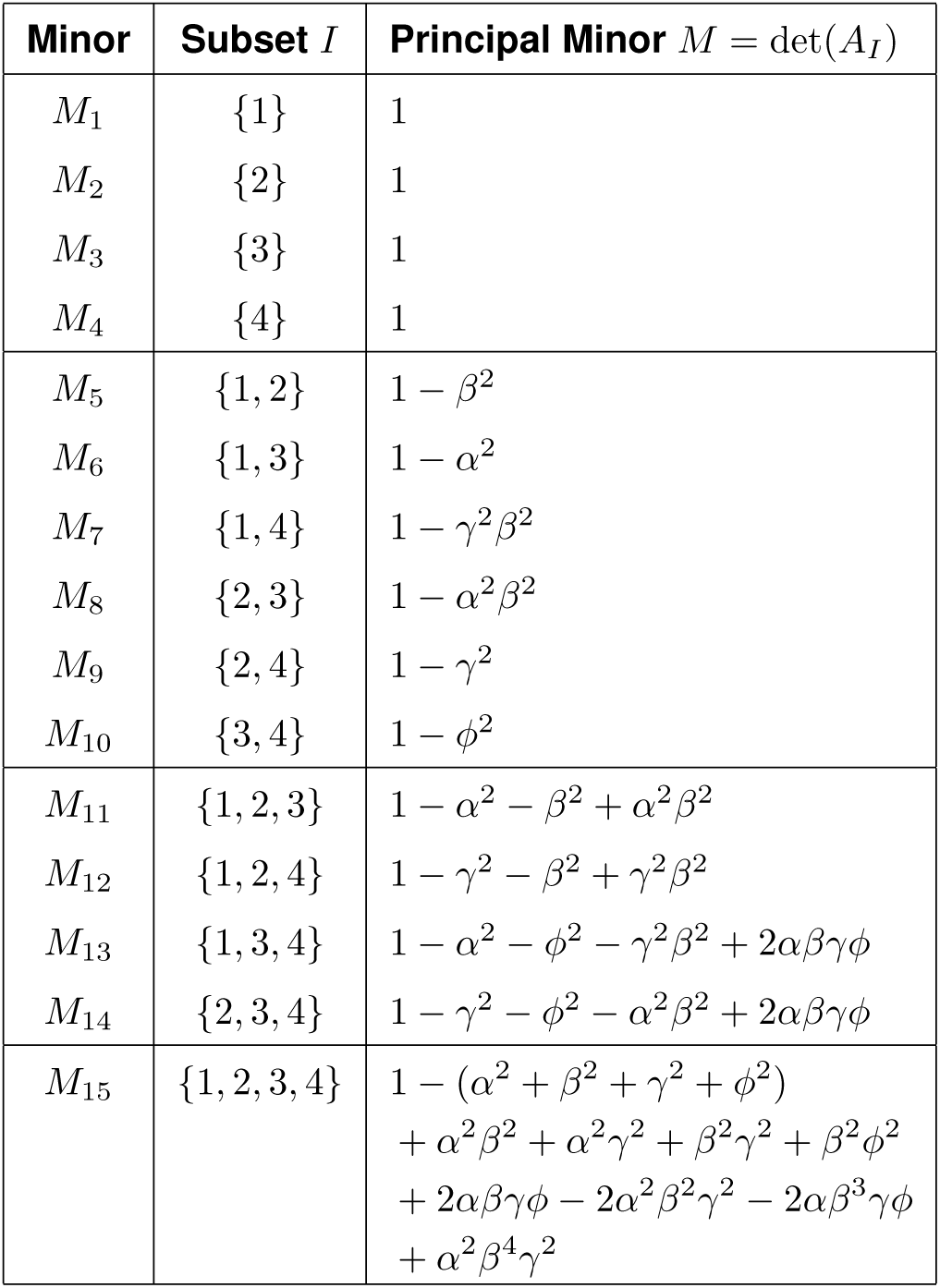
Principal minors of the 4 *×* 4 correlation matrix A.

As shown in Fig. N1.3a, the predicted range for *r*(*CNV*, *PGS_GW_*) for each trait is broad, with the narrowest interval across the 119 assessed CNV-trait pairs spanning -0.79 to 0.78. Similar minimum ranges are observed for the 21 phenotypes affected by the CNV burden (Fig. N1.3b). This supports that the observed correlation between indirect and total paths is not trivially dictated by PSD bounds alone.

**Figure N1.3.**
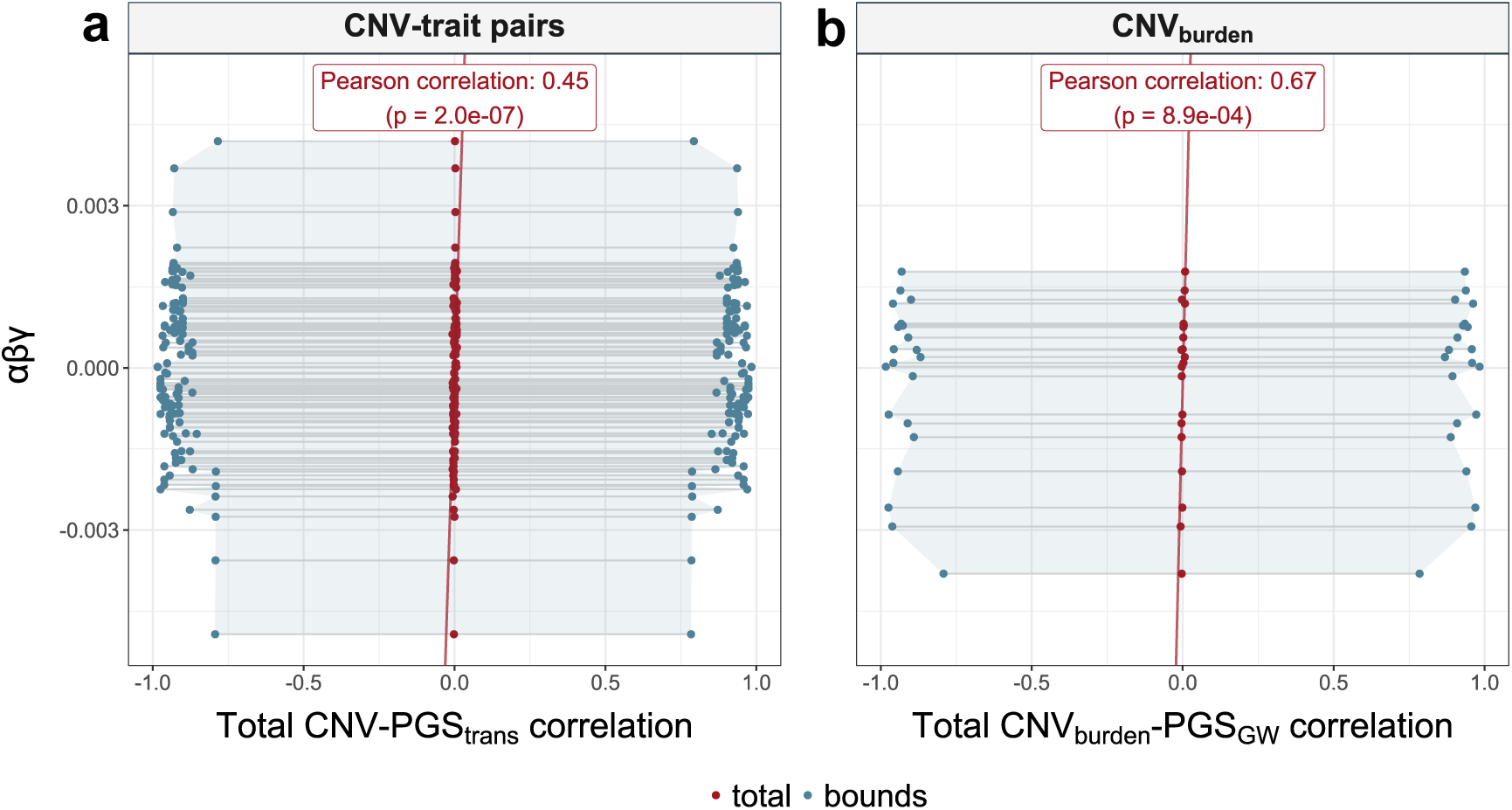
Theoretical bounds for the CNV-PGSGW correlation. **a,** *α × β × γ* (y-axis) against the total observed CNV-PGStrans correlation (x-axis) are shown as red dots. Lower (*max*(*L*10*, L*13*, L*14*, L*15)) and upper (*min*(*U*10*, U*13*, U*14*, U*15)) bounds for total CNV-PGSGW correlation are shown in blue. Line segments connect values corresponding to each of the 119 CNV-trait pairs. The Pearson correlation coefficient between *α×β ×γ* and the total CNV-PGStrans correlation is displayed. **b,** same analysis as in **a,** using the CNV burden instead of individual CNV-trait pairs, and PGSGW instead of PGStrans. Each connected segment corresponds to one of the 21 phenotypes significantly affected by CNV burden.

### Supplementary Note 2

#### Analytical approximation of the CNV-PGS correlation

We define the random variables X and Y as the phenotypes of each partner with C representing the CNV carrier status of the first partner, and G the PGS_GW_ of the second partner (Fig. N2).

**Figure N2.**
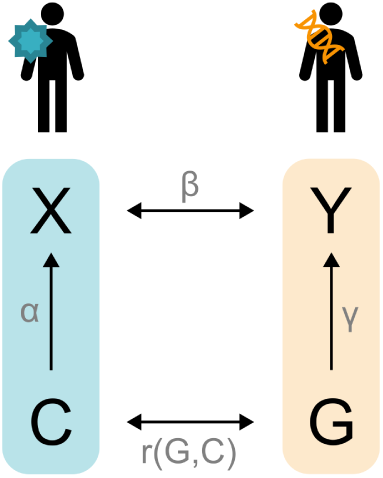
Schematic representation of assortative mating modeling. *X* and *Y* represent the phenotypic values for the same trait among two partners, with *X* being influenced by the first individual’s CNV (C; correlation between the variables is denoted *α*) and *Y* being influenced by the second individual’s PGS (*G*; correlation between the variables is denoted *γ*). The phenotypic correlation strength between *X* and *Y* is denoted *β*.

Each variable is approximated by a standard normal distribution:

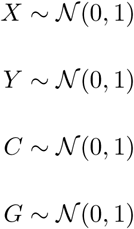

We then assume a causal model in which the CNV directly influences the phenotype of one partner (*C* → *X*) and the PGS_GW_ directly influences the phenotype of the other partner (*G* → *Y*) so that:

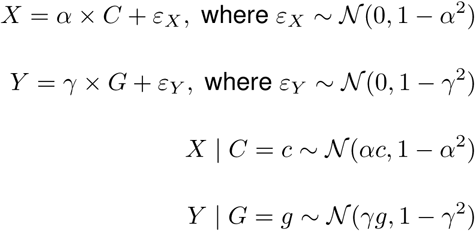

Partners’ phenotypes *X* and *Y* are correlated due to assortative mating, such that *r*(*X*, *Y*) = *β*. To compute the joint probability density function (PDF) of *C* and *G*, we apply the law of total probability by conditioning on the couple’s phenotypes and integrating over all possible values of *X* and *Y*. This yields:

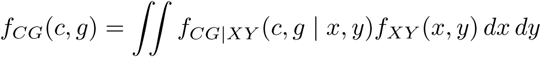

Since *C* and *G* are conditionally independent, the joint conditional probability can be factorized as:

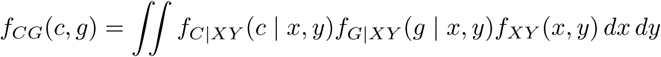

Given that *C* is independent of *Y*, and *G* is independent of *X*, we can further simplify the expression:

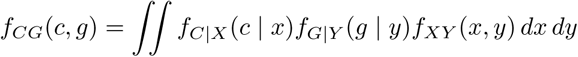

Using Bayes’ theorem to express the PDFs *f_C|X_* (*c | x*) and *f_G|Y_* (*g | y*) in terms of their corresponding inverse forms, we obtain:

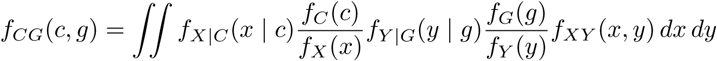

Finally, under the assumption that *f_XY_* is bivariate normal:

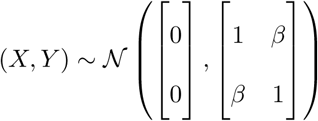

The integral becomes:

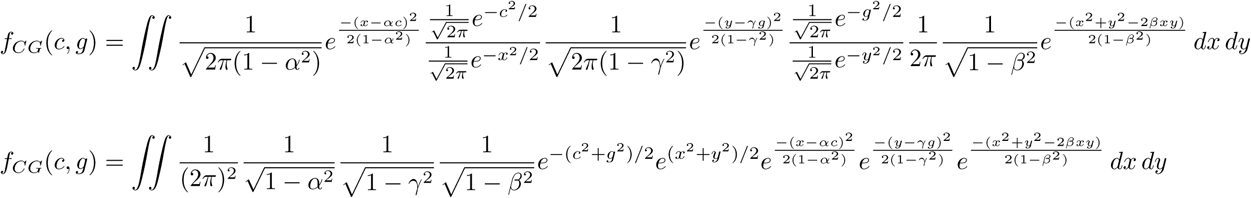

Symbolic integration with Maple yields the following closed-form solution for the PDF of G and C:

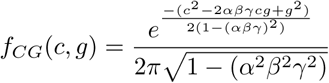

This expression corresponds to the joint PDF of two random variables that follow a bivariate normal distribution. The general form of such distribution is given by:

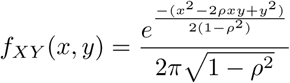

Where *ρ* denotes the correlation between *X* and *Y*. Comparing the two expressions, it follows that under the specified assumptions:

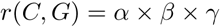

### Supplementary Note 3

#### Total vs indirect CNV–PGS_GW_ correlations in mating simulations

To assess whether *α* × *β* × *γ* reliably predicts the CNV-PGS_GW_ correlation under assortative mating, we performed several forward simulations. In each of the 100 runs, 10,000 individuals with CNV-determined phenotype selected a partner from a pool of 500 candidates, each with a PGS_GW_-determined phenotype. The matching criterion was to pair individuals whose phenotypic correlation was the closest to a fixed value *β*, which defined the strength of assortative mating. We ran four replicates for *β* = 0 (no phenotypic correlation), *β* = 0.25 (moderate correlation), *β* = 0.5 (strong correlation), and *β* = 1 (perfect correlation). We computed both the predicted, indirect (*α×β ×γ*) and empirical, total (*r*(*CNV, PGS_GW_*)) correlations for four different values of *β*, across a grid of *α* and *γ* values. For each coordinate, the total correlation was averaged across simulation replicates. Finally, we fit the simulation results to non-linear models (specifically, rotated hyperbolic paraboloids) using the nlsLM function from the minpack.lm R package, except for *β* = 0 results, which were fitted with a linear model, using the lm function. Figure N3 shows the predictions of these models for a larger grid of *α* and *γ* values. Simulations showed that *α* × *β* × *γ* predicts well the CNV-PGS_GW_ correlation for each value of *β*. Total and indirect correlations exhibit the largest positive correlation when both *α* and *γ* are simultaneously 1 or -1. Conversely, they are maximally anticorrelated when they present opposite signs (total = 1, indirect = -1, or vice versa). We can also observe that, in the absence of assortment, we expect no CNV-PGS_GW_ correlation, as correctly predicted by the indirect path.

**Figure N3.**
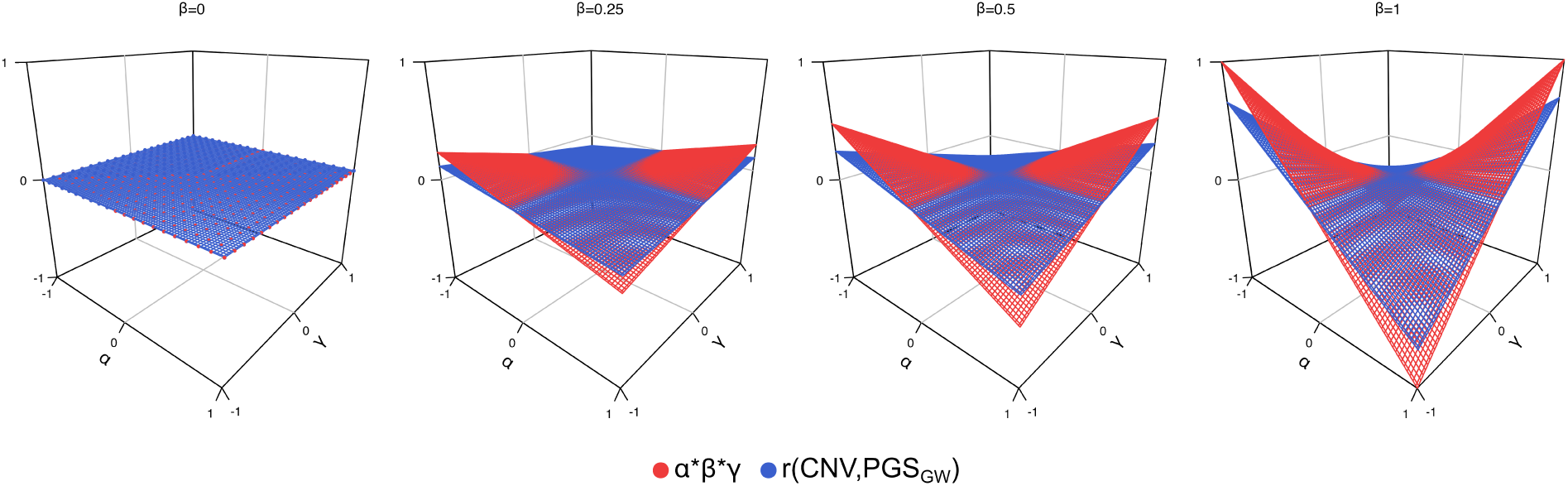
Simulated total versus indirect CNV-PGSGW under various degrees of assortative mating. Mean of the empirical, total CNV–PGSGW correlations (red) and corresponding *α × β × γ* predicted indirect correlation (blue) (z-axis) across four levels of assortative mating strength (*β* = 0, 0.25, 0.5, and 1) plotted over a range of *α* (x-axis) and *γ* (y-axis) values.

