## Extended Data 1 for "Compounding of rare pathogenic copy-number variants and polygenic background is consistent with assortative mating"

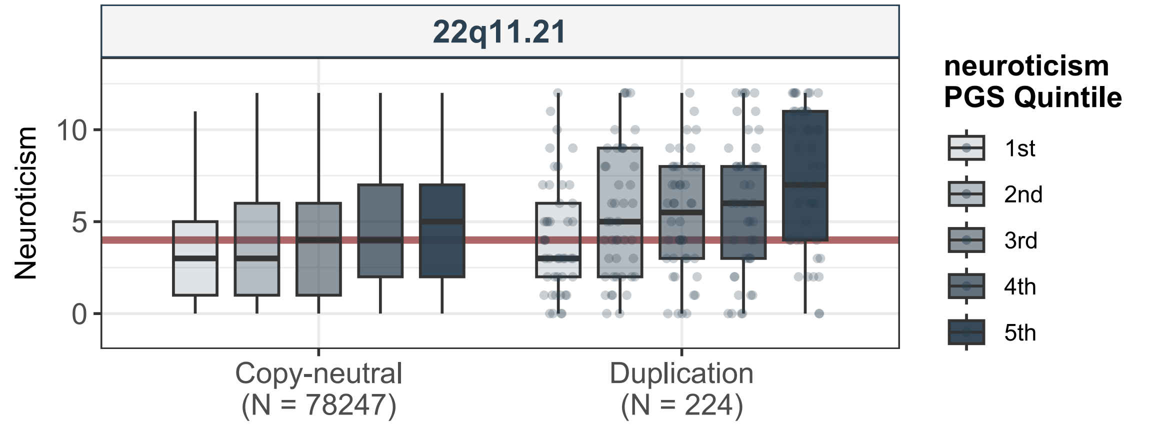
22q11.21
Neuroticism
10
5
0
Copy-neutral
(N = 78247)
Duplication
(N = 224)
neuroticism
PGS Quintile
1st
2nd
3rd
4th
5th

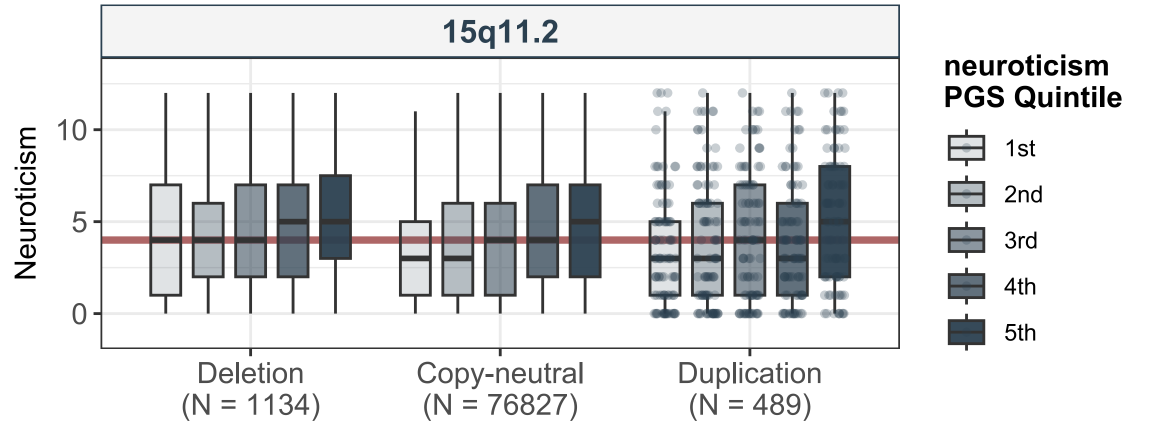
15q11.2
Neuroticism
10
5
0
Deletion
(N = 1134)
Copy-neutral
(N = 76827)
Duplication
(N = 489)
neuroticism
PGS Quintile
1st
2nd
3rd
4th
5th

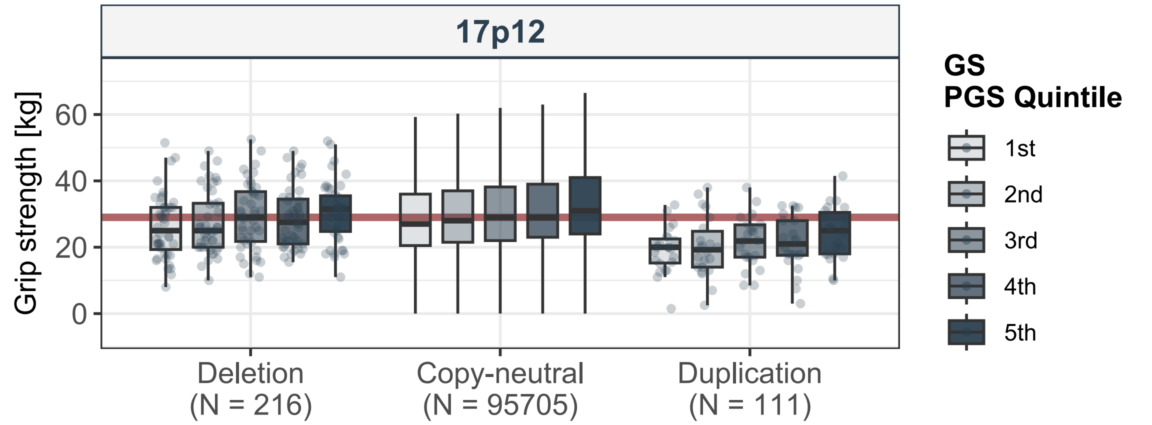

17p12
Grip strength [kg]
60
40
20
0
Deletion
(N = 216)
Copy-neutral
(N = 95705)
Duplication
(N = 111)
GS
PGS Quintile
1st
2nd
3rd
4th
5th

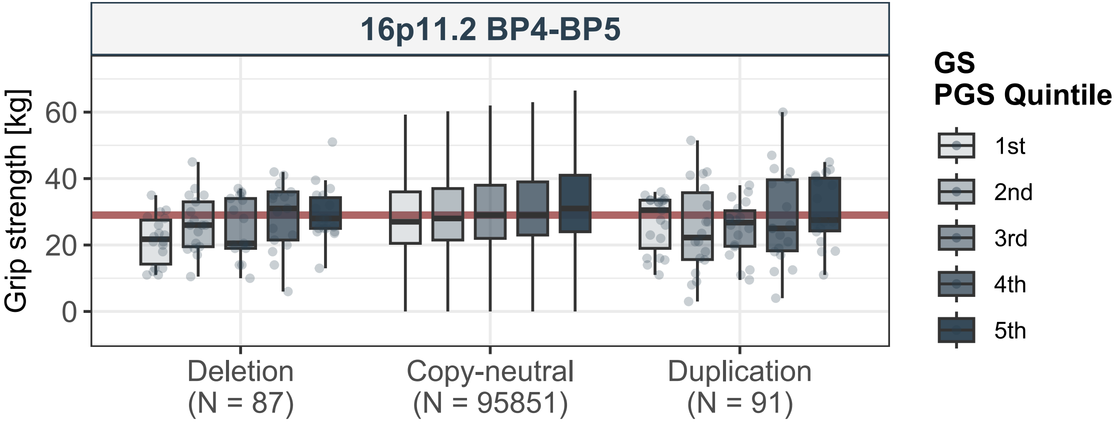
16p11.2 BP4-BP5
Grip strength [kg]
60
40
20
0
Deletion
(N = 87)
Copy-neutral
(N = 95851)
Duplication
(N = 91)
GS
PGS Quintile
1st
2nd
3rd
4th
5th

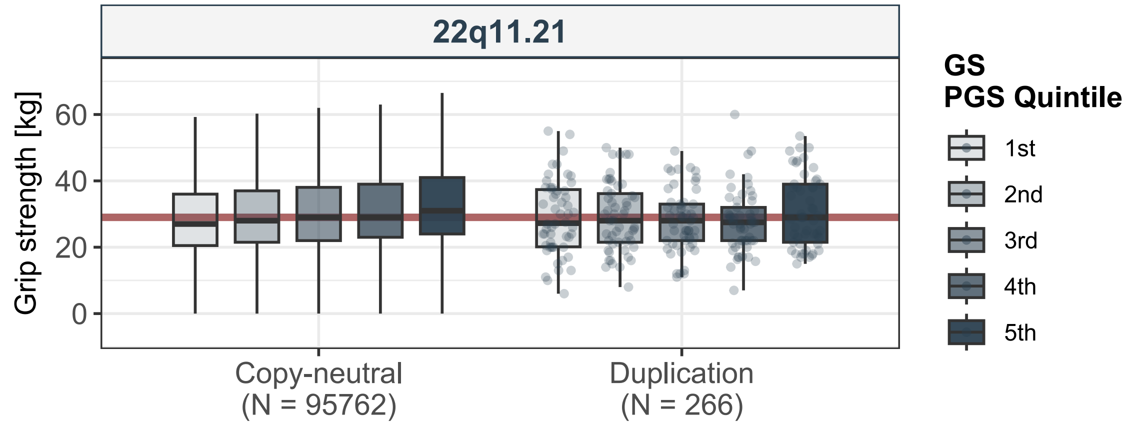
22q11.21
Grip strength [kg]
60
40
20
0
Copy-neutral
(N = 95762)
Duplication
(N = 266)
GS
PGS Quintile
1st
2nd
3rd
4th
5th

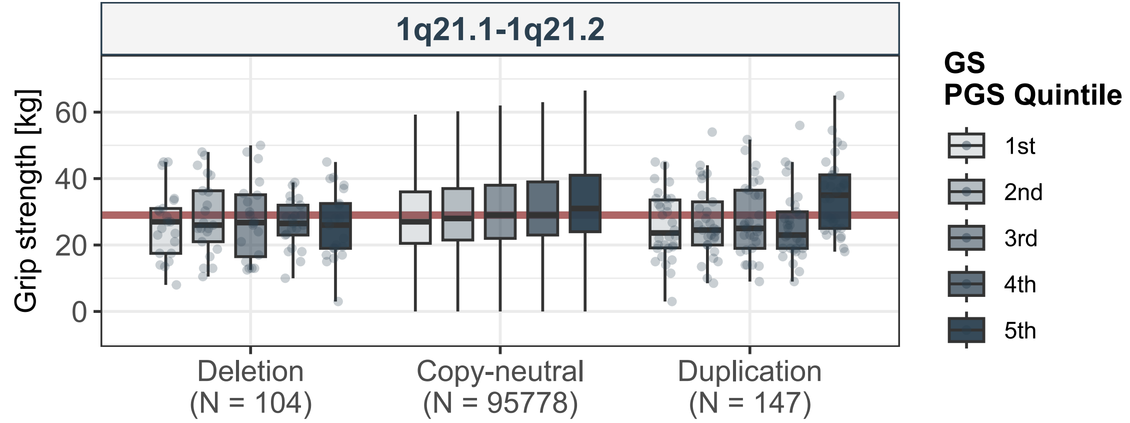
1q21.1-1q21.2
Grip strength [kg]
60
40
20
0
Deletion
(N = 104)
Copy-neutral
(N = 95778)
Duplication
(N = 147)
GS
PGS Quintile
1st
2nd
3rd
4th
5th

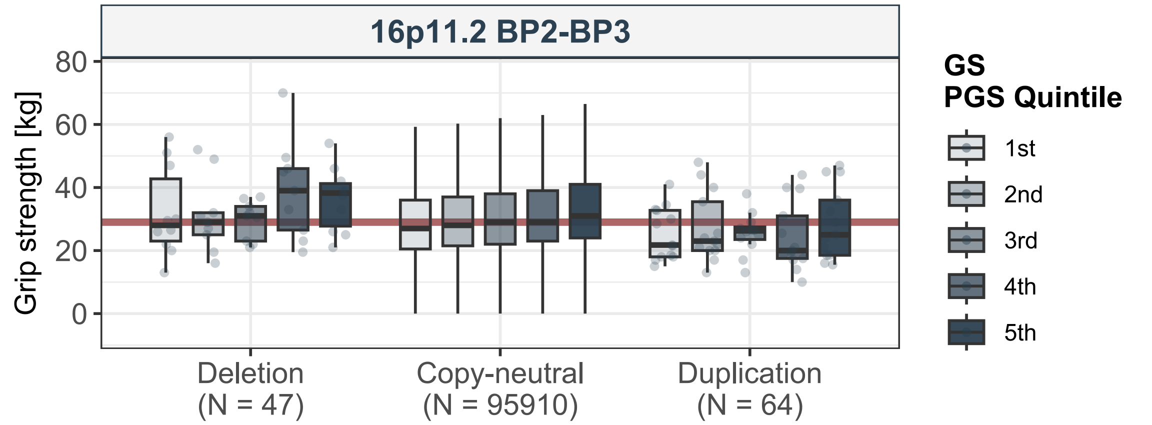
16p11.2 BP2-BP3
Grip strength [kg]
80
60
40
20
0
Deletion
(N = 47)
Copy-neutral
(N = 95910)
Duplication
(N = 64)
GS
PGS Quintile
1st
2nd
3rd
4th
5th

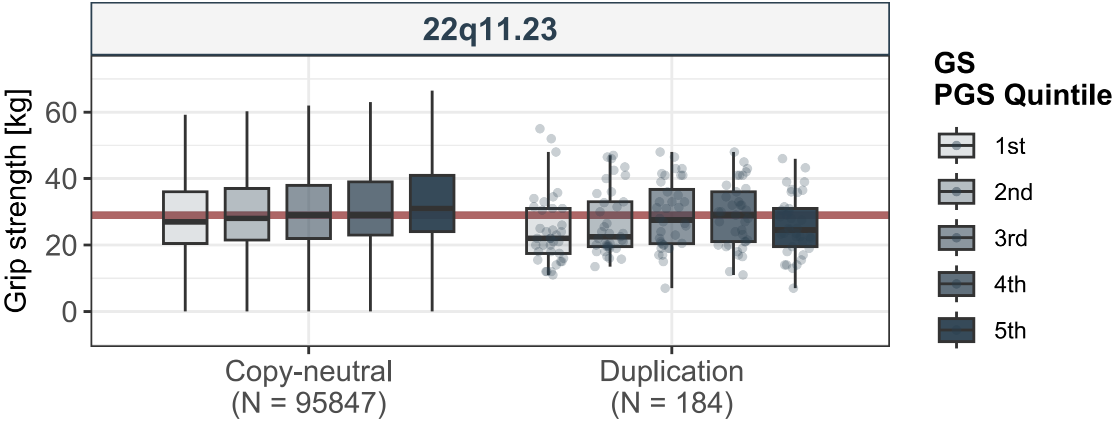
22q11.23
Grip strength [kg]
0
20
40
60
Copy-neutral
(N = 95847)
Duplication
(N = 184)
GS
PGS Quintile
1st
2nd
3rd
4th
5th

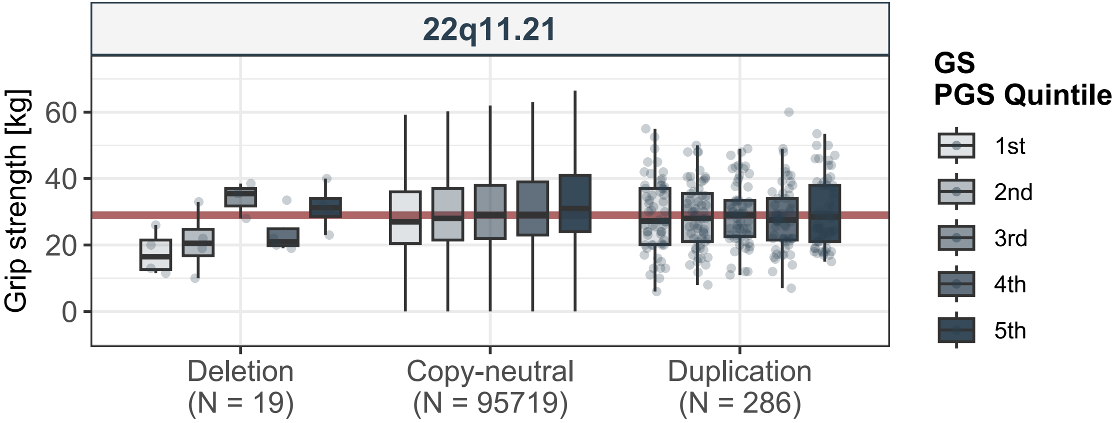
22q11.21
Grip strength [kg]
60
40
20
0
Deletion
(N = 19)
Copy-neutral
(N = 95719)
Duplication
(N = 286)
GS
PGS Quintile
1st
2nd
3rd
4th
5th

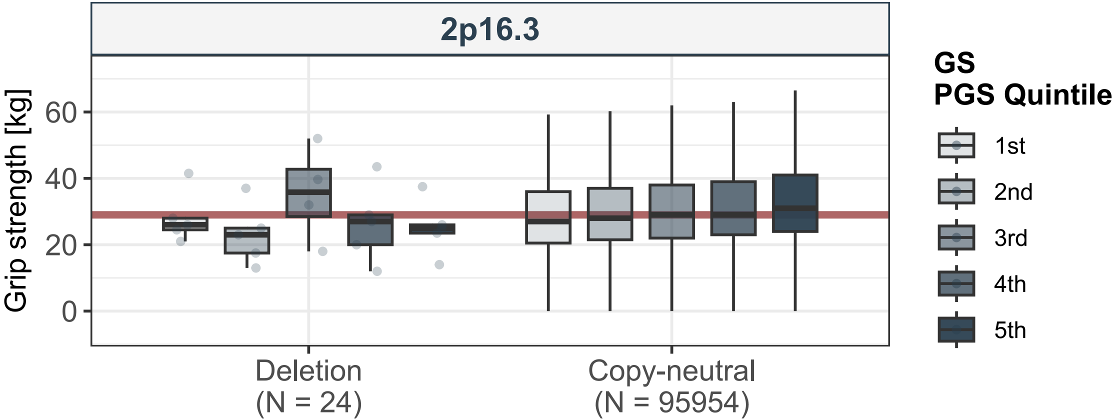
2p16.3
Grip strength [kg]
60
40
20
0
Deletion
(N = 24)
Copy-neutral
(N = 95954)
GS
PGS Quintile
1st
2nd
3rd
4th
5th

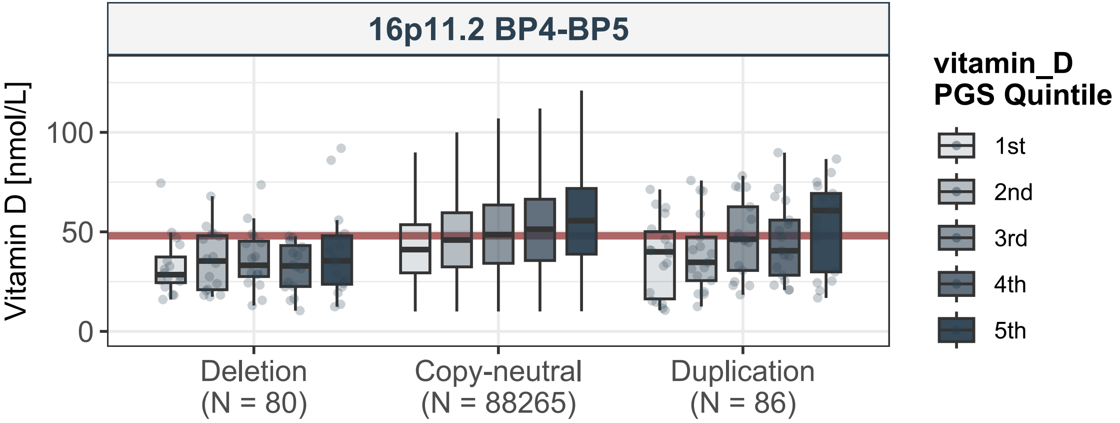

16p11.2 BP4-BP5
Vitamin D [nmol/L]
100
50
0
Deletion
(N = 80)
Copy-neutral
(N = 88265)
Duplication
(N = 86)
vitamin_D
PGS Quintile
1st
2nd
3rd
4th
5th

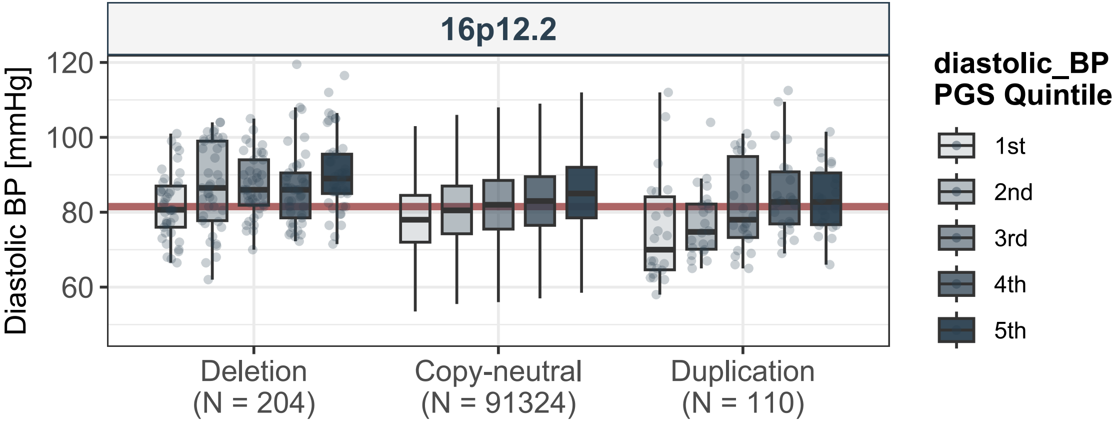

16p12.2
Diastolic BP [mmHg]
120
100
80
60
Deletion
(N = 204)
Copy-neutral
(N = 91324)
Duplication
(N = 110)
diastolic_BP
PGS Quintile
1st
2nd
3rd
4th
5th

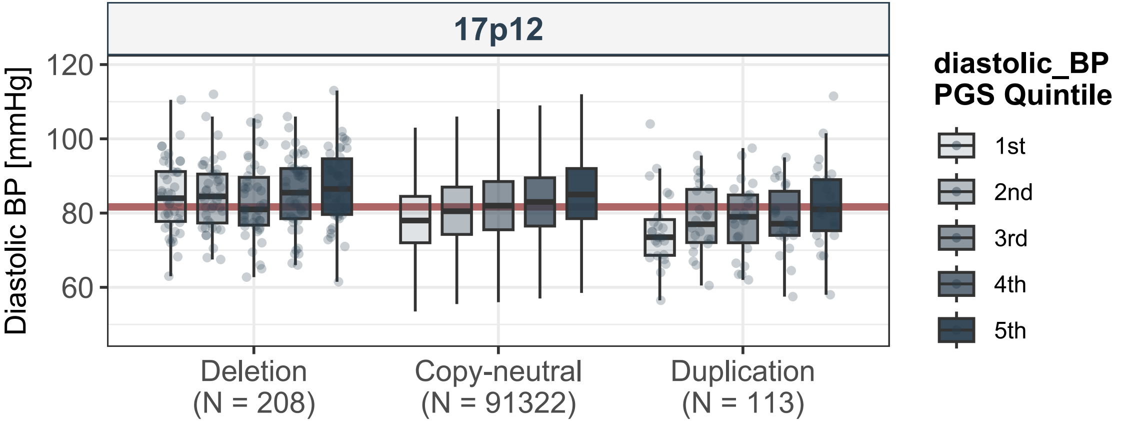
17p12
Diastolic BP [mmHg]
120
100
80
60
Deletion
(N = 208)
Copy-neutral
(N = 91322)
Duplication
(N = 113)
diastolic_BP
PGS Quintile
1st
2nd
3rd
4th
5th

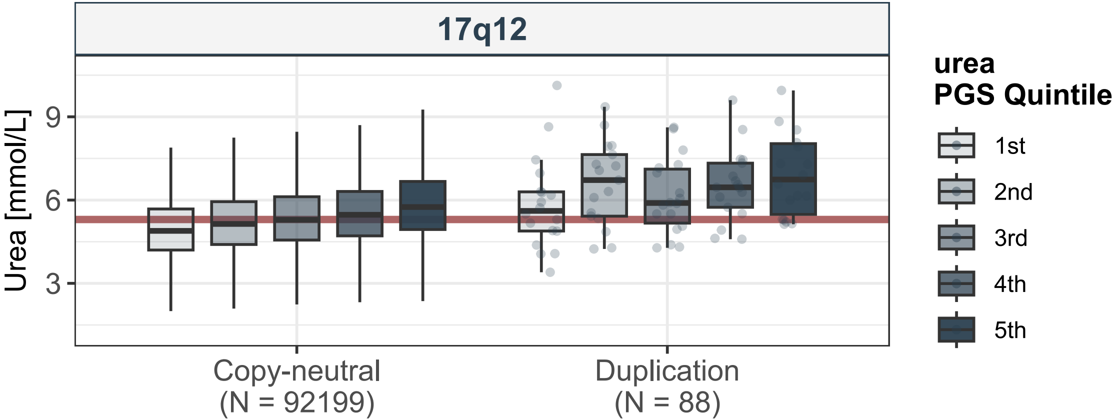
17q12
Urea [mmol/L]
9
6
3
Copy-neutral
(N = 92199)
Duplication
(N = 88)
urea
PGS Quintile
1st
2nd
3rd
4th
5th

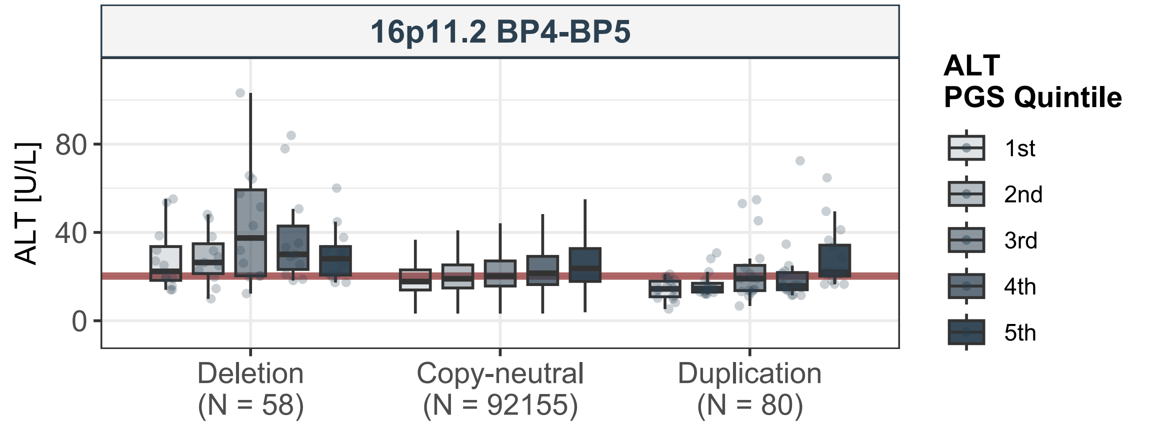
16p11.2 BP4-BP5
ALT [U/L]
80
40
0
Deletion
(N = 58)
Copy-neutral
(N = 92155)
Duplication
(N = 80)
ALT
PGS Quintile
1st
2nd
3rd
4th
5th

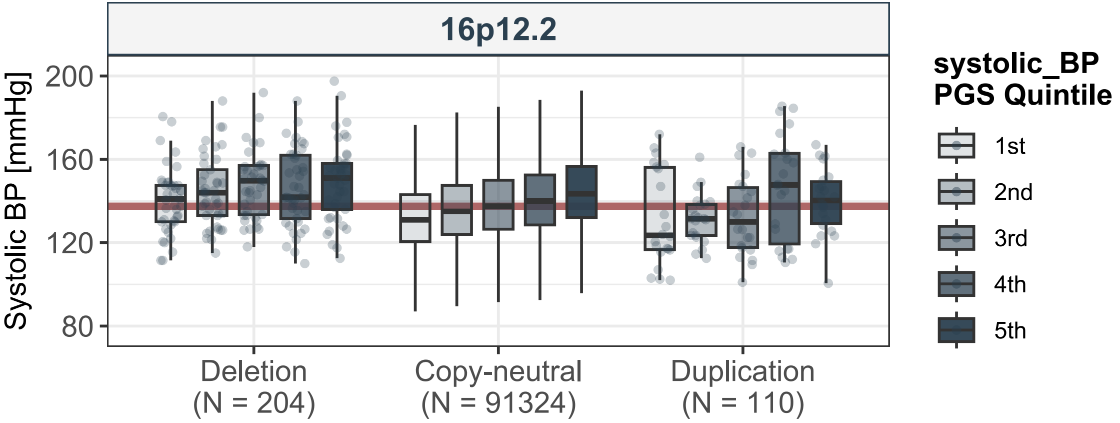

16p12.2
Systolic BP [mmHg]
200
160
120
80
Deletion
(N = 204)
Copy-neutral
(N = 91324)
Duplication
(N = 110)
systolic_BP
PGS Quintile
1st
2nd
3rd
4th
5th

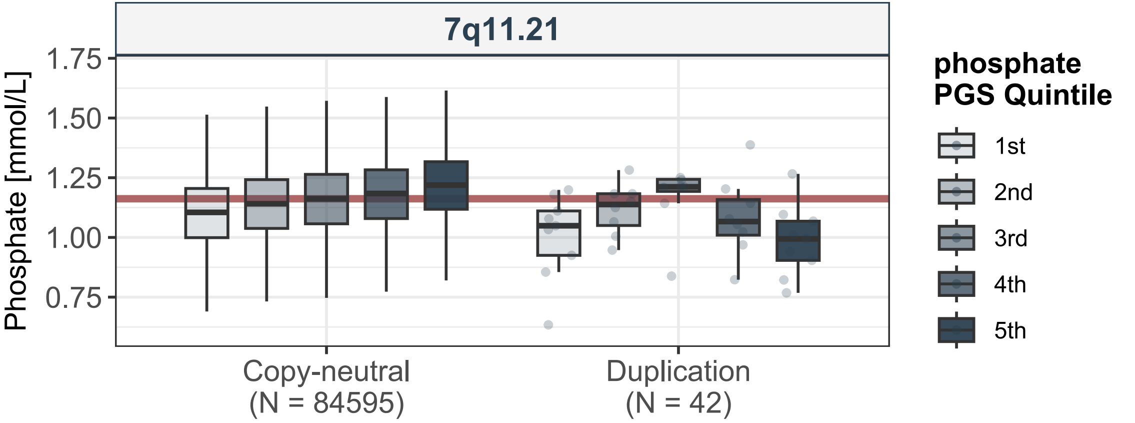
7q11.21
Phosphate [mmol/L]
1.75
1.50
1.25
1.00
0.75
Copy-neutral
(N = 84595)
Duplication
(N = 42)
phosphate
PGS Quintile
1st
2nd
3rd
4th
5th

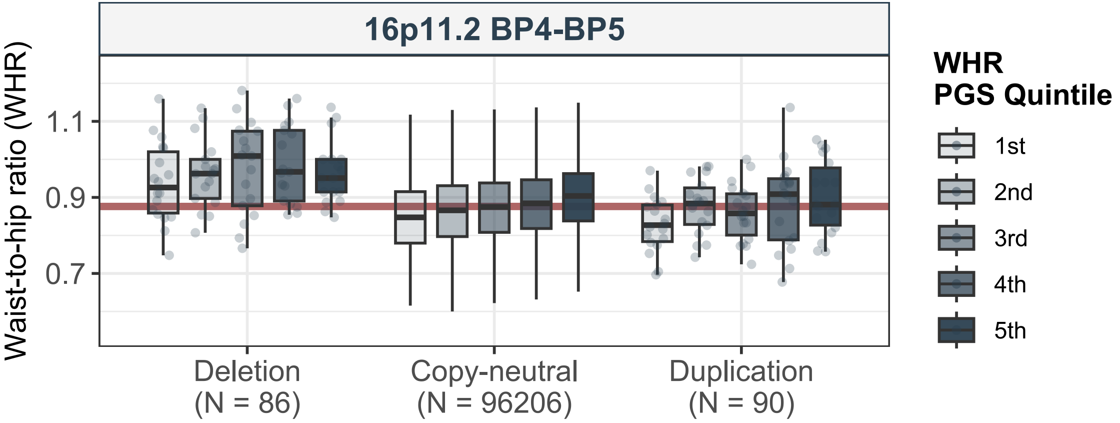
16p11.2 BP4-BP5
Waist-to-hip ratio (WHR)
1.1
0.9
0.7
Deletion
(N = 86)
Copy-neutral
(N = 96206)
Duplication
(N = 90)
WHR
PGS Quintile
1st
2nd
3rd
4th
5th

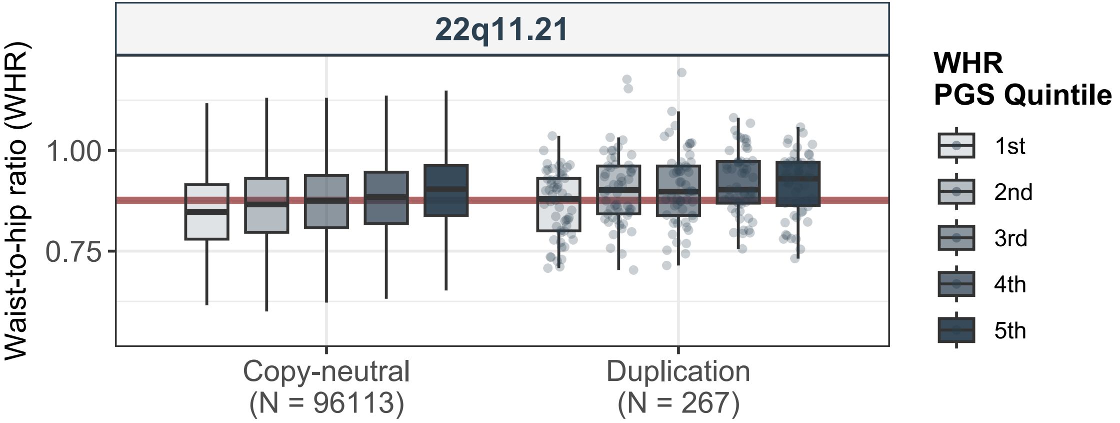
22q11.21
Waist-to-hip ratio (WHR)
1.00
0.75
Copy-neutral
(N = 96113)
Duplication
(N = 267)
WHR
PGS Quintile
1st
2nd
3rd
4th
5th

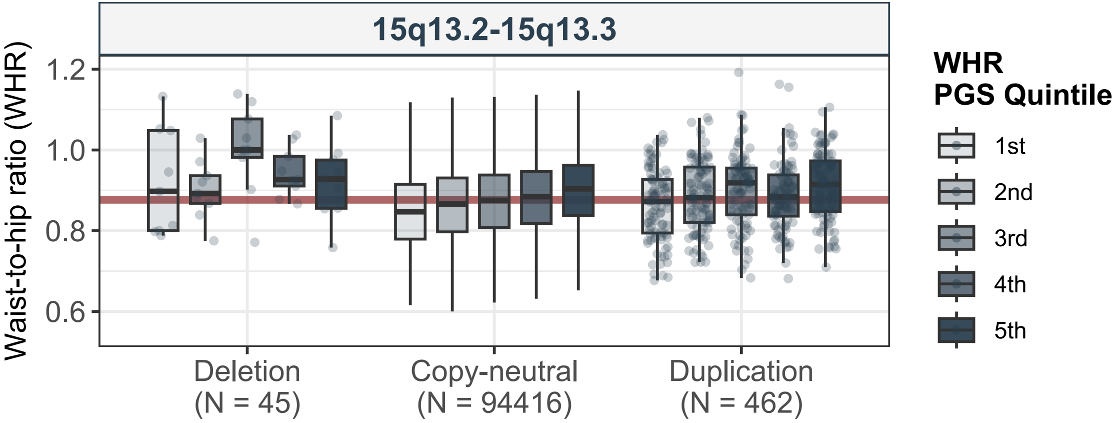
15q13.2-15q13.3
Waist-to-hip ratio (WHR)
1.2
1.0
0.8
0.6
Deletion
(N = 45)
Copy-neutral
(N = 94416)
Duplication
(N = 462)
WHR
PGS Quintile
1st
2nd
3rd
4th
5th

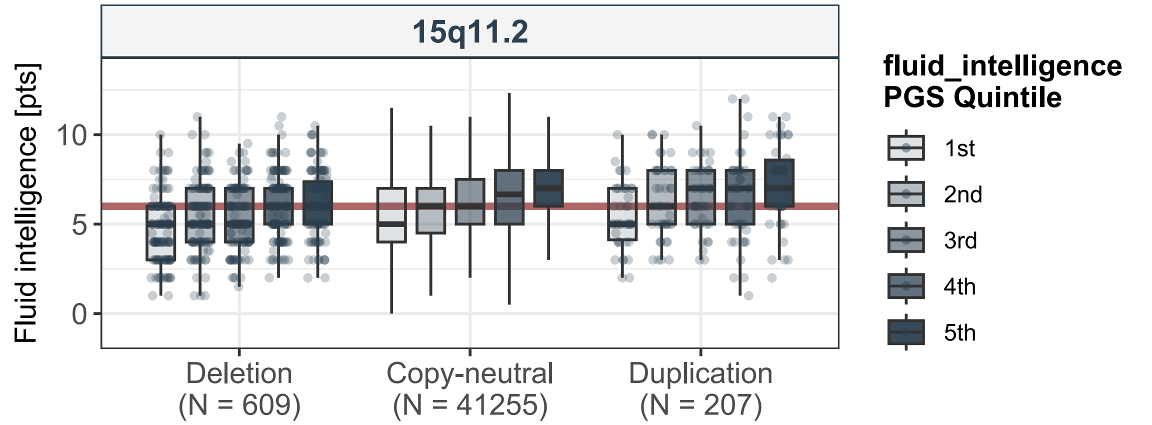
15q11.2
Fluid intelligence [pts]
0
5
10
Deletion
(N = 609)
Copy-neutral
(N = 41255)
Duplication
(N = 207)
fluid_intelligence
PGS Quintile
1st
2nd
3rd
4th
5th

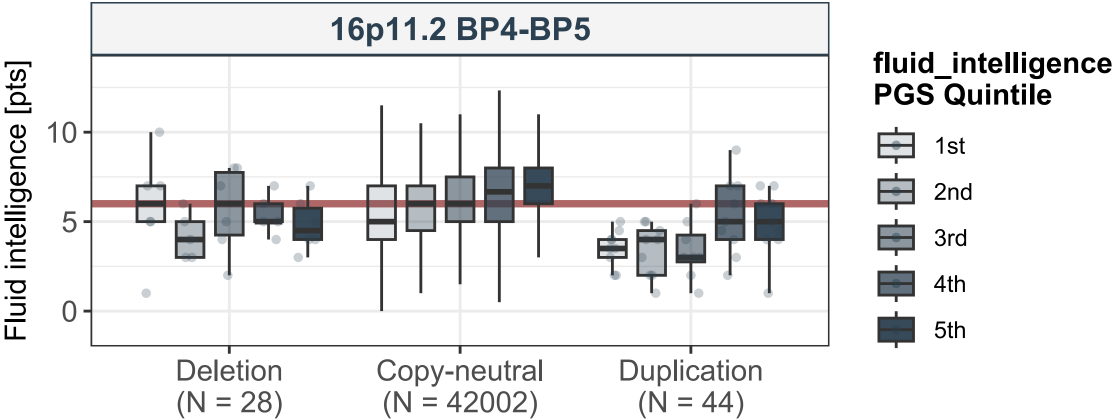

16p11.2 BP4-BP5
Fluid intelligence [pts]
10
5
0
Deletion
(N = 28)
Copy-neutral
(N = 42002)
Duplication
(N = 44)
fluid_intelligence
PGS Quintile
1st
2nd
3rd
4th
5th

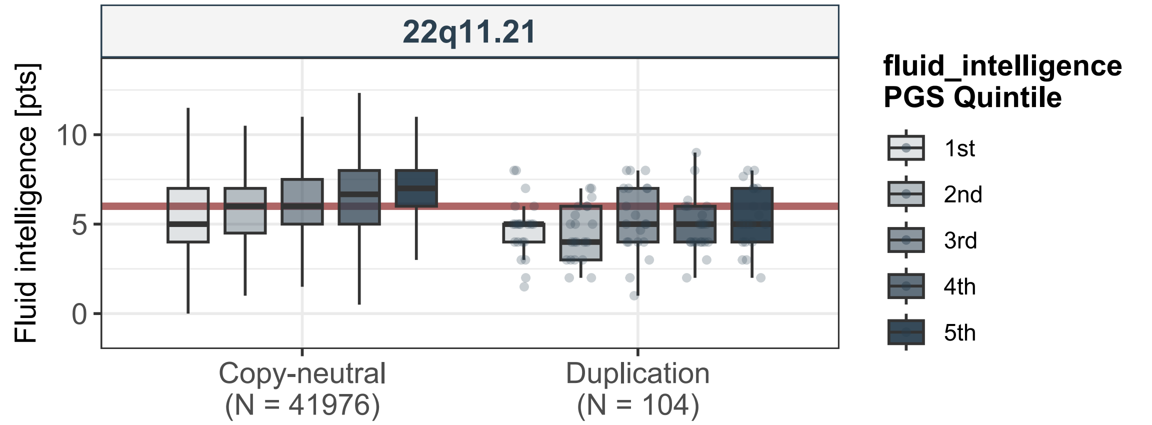
22q11.21
Fluid intelligence [pts]
0
5
10
Copy-neutral
(N = 41976)
Duplication
(N = 104)
fluid_intelligence
PGS Quintile
1st
2nd
3rd
4th
5th

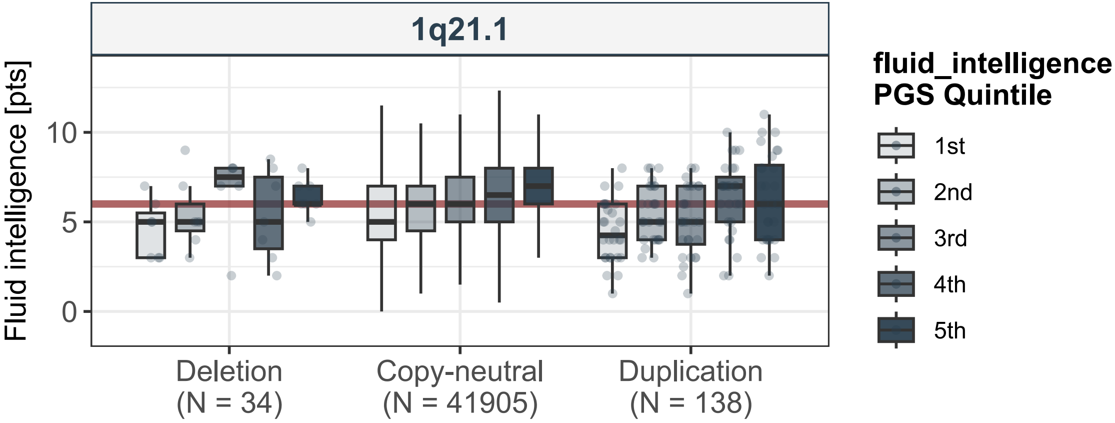
1q21.1
Fluid intelligence [pts]
10
5
0
Deletion
(N = 34)
Copy-neutral
(N = 41905)
Duplication
(N = 138)
fluid_intelligence
PGS Quintile
1st
2nd
3rd
4th
5th

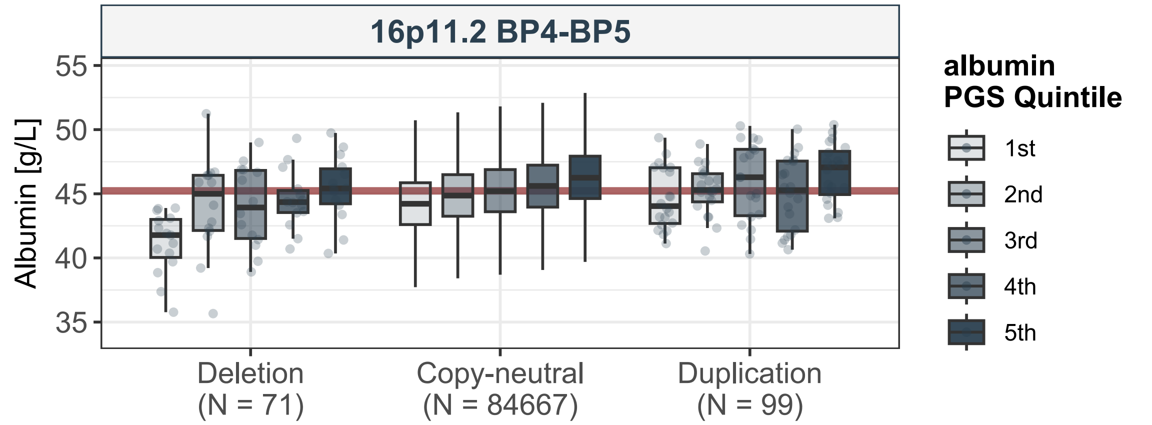
16p11.2 BP4-BP5
Albumin [g/L]
55
50
45
40
35
Deletion
(N = 71)
Copy-neutral
(N = 84667)
Duplication
(N = 99)
albumin
PGS Quintile
1st
2nd
3rd
4th
5th

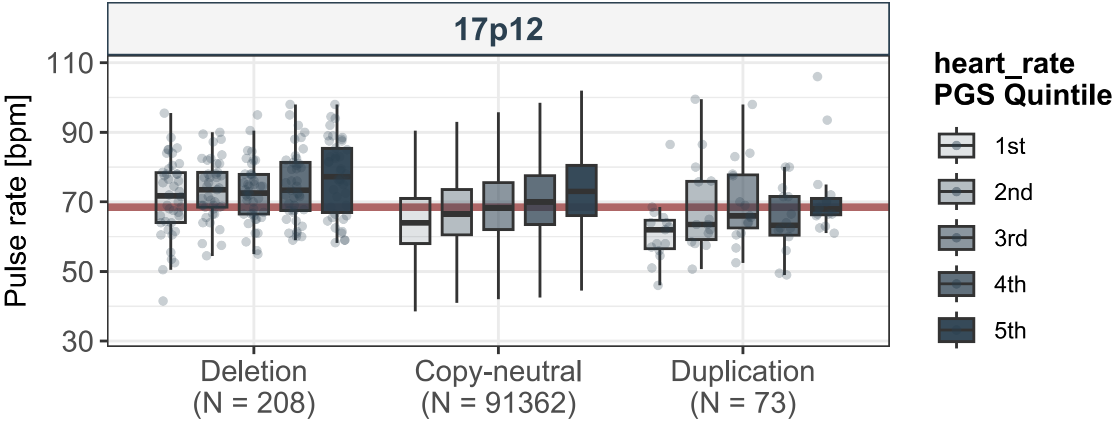
17p12
Pulse rate [bpm]
110
90
70
50
30
Deletion
(N = 208)
Copy-neutral
(N = 91362)
Duplication
(N = 73)
heart_rate
PGS Quintile
1st
2nd
3rd
4th
5th

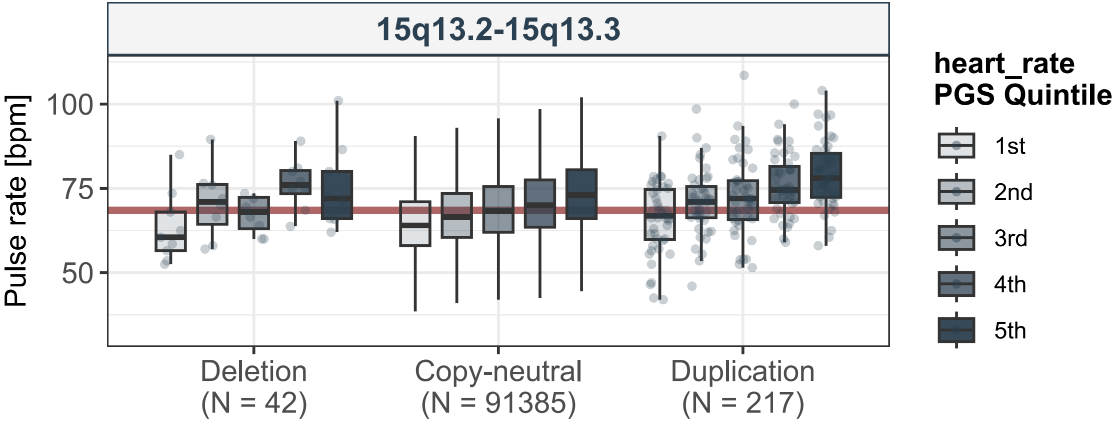
15q13.2-15q13.3
Pulse rate [bpm]
100
75
50
Deletion
(N = 42)
Copy-neutral
(N = 91385)
Duplication
(N = 217)
heart_rate
PGS Quintile
1st
2nd
3rd
4th
5th

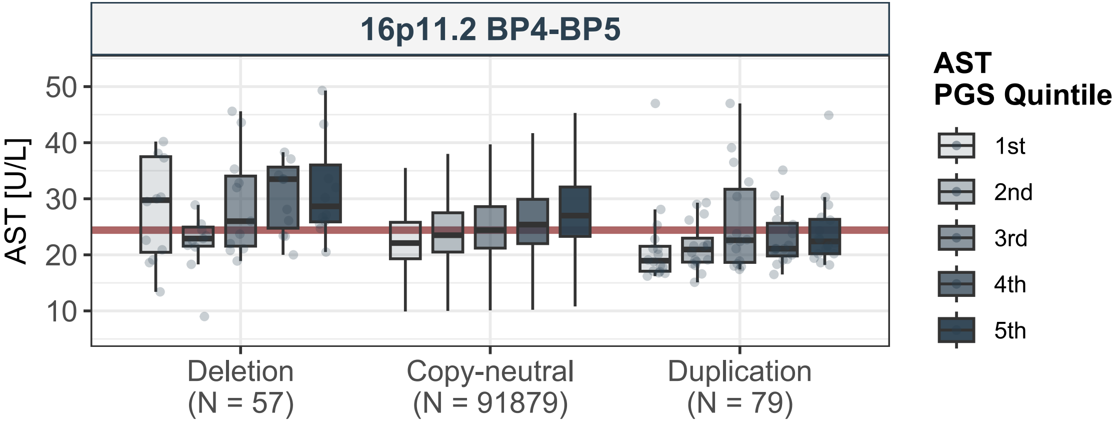
16p11.2 BP4-BP5
AST [U/L]
50
40
30
20
10
Deletion
(N = 57)
Copy-neutral
(N = 91879)
Duplication
(N = 79)
AST
PGS Quintile
1st
2nd
3rd
4th
5th

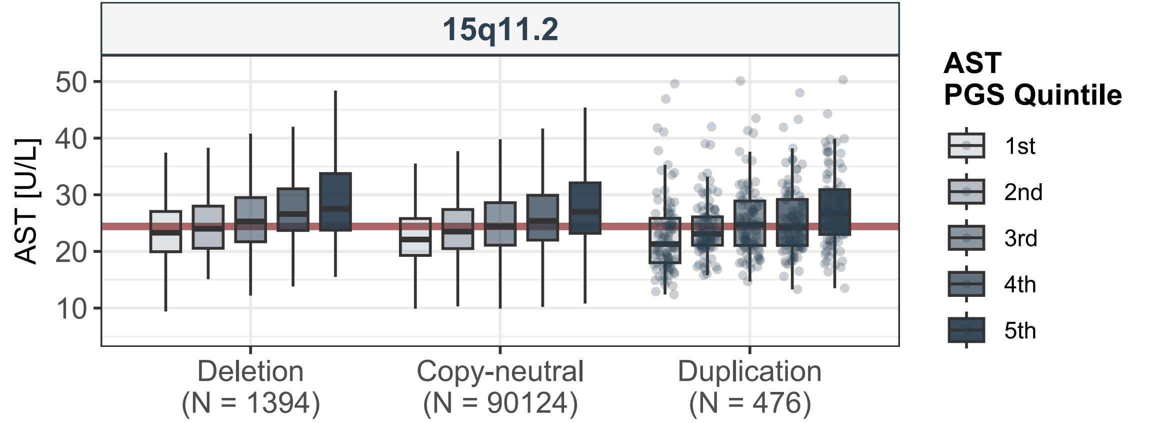
15q11.2
AST [U/L]
50
40
30
20
10
Deletion
(N = 1394)
Copy-neutral
(N = 90124)
Duplication
(N = 476)
AST
PGS Quintile
1st
2nd
3rd
4th
5th

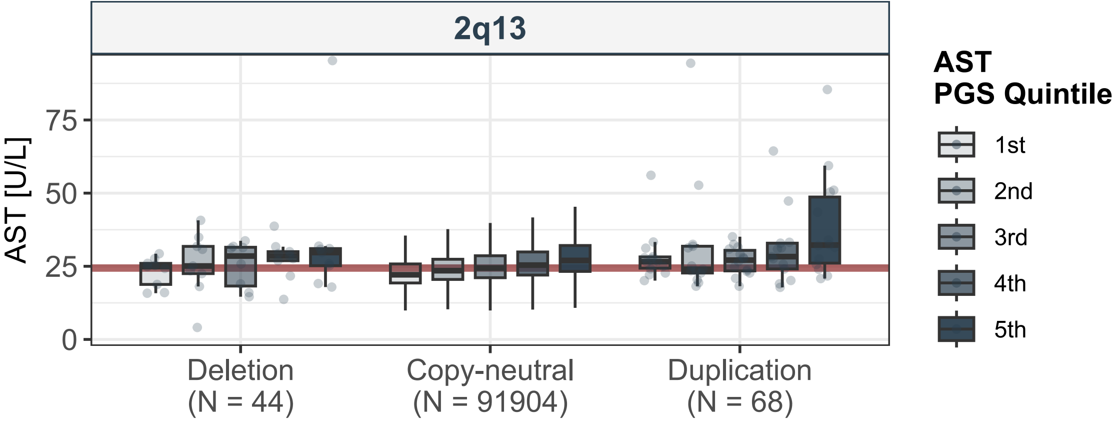
2q13
AST [U/L]
0
25
50
75
Deletion
(N = 44)
Copy-neutral
(N = 91904)
Duplication
(N = 68)
AST
PGS Quintile
1st
2nd
3rd
4th
5th

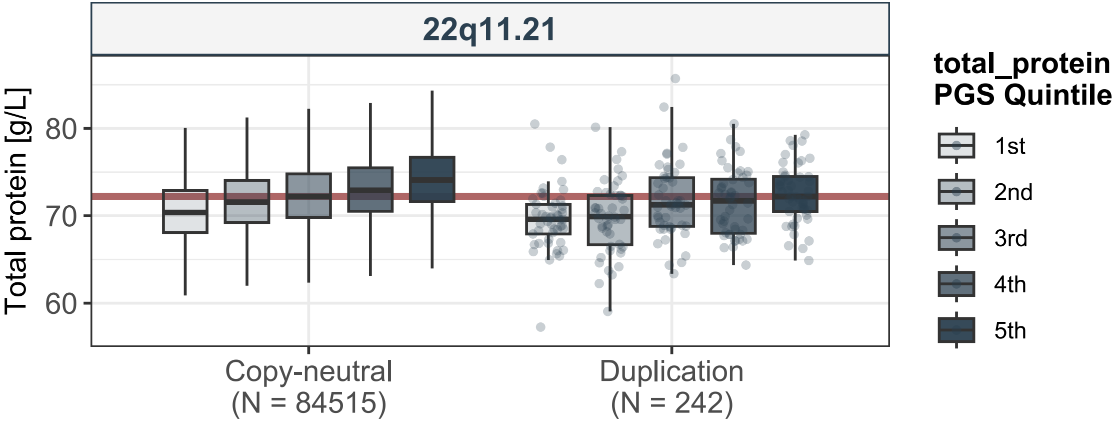
22q11.21
Total protein [g/L]
80
70
60
Copy-neutral
(N = 84515)
Duplication
(N = 242)
total_protein
PGS Quintile
1st
2nd
3rd
4th
5th

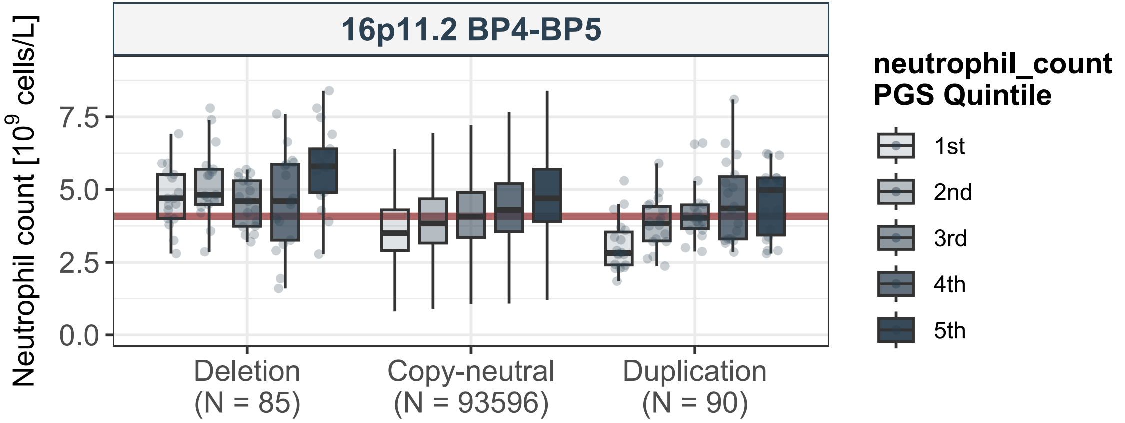
16p11.2 BP4-BP5
Neutrophil count [$10^9$ cells/L]
7.5
5.0
2.5
0.0
Deletion
(N = 85)
Copy-neutral
(N = 93596)
Duplication
(N = 90)
neutrophil_count
PGS Quintile
1st
2nd
3rd
4th
5th

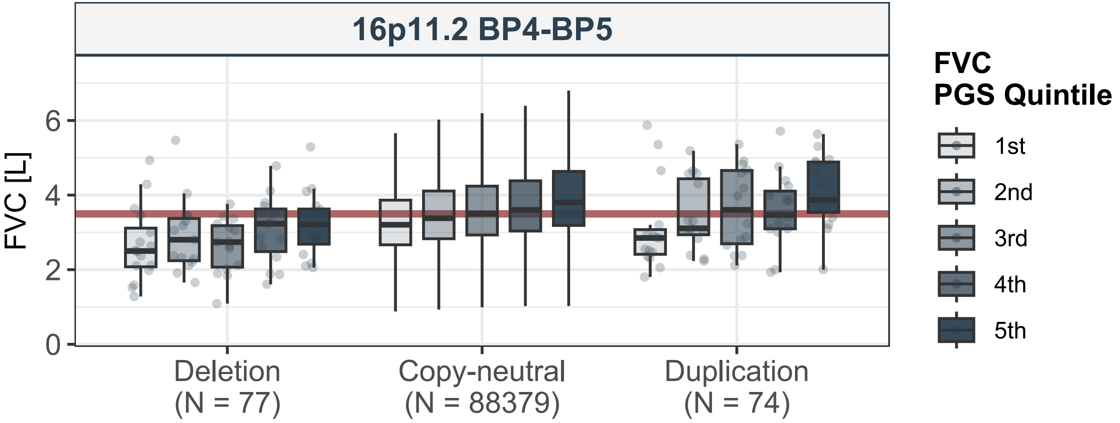
16p11.2 BP4-BP5
FVC [L]
0
2
4
6
Deletion
(N = 77)
Copy-neutral
(N = 88379)
Duplication
(N = 74)
FVC
PGS Quintile
1st
2nd
3rd
4th
5th

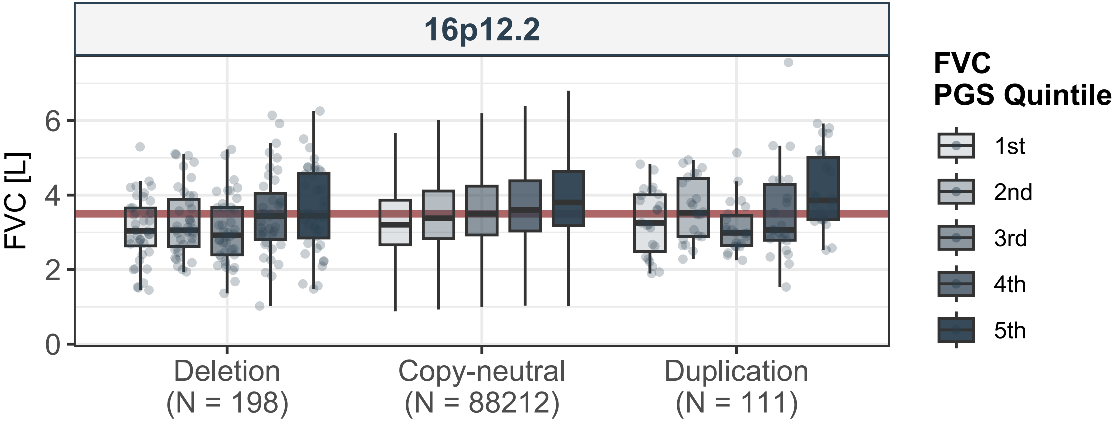

16p12.2
FVC [L]
0
2
4
6
Deletion
(N = 198)
Copy-neutral
(N = 88212)
Duplication
(N = 111)
FVC
PGS Quintile
1st
2nd
3rd
4th
5th

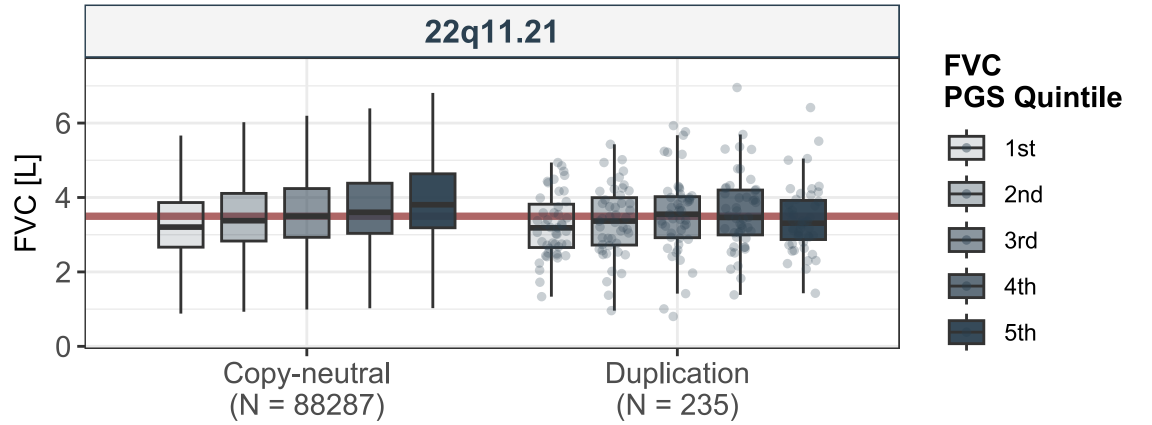
22q11.21
FVC [L]
0
2
4
6
Copy-neutral
(N = 88287)
Duplication
(N = 235)
FVC
PGS Quintile
1st
2nd
3rd
4th
5th

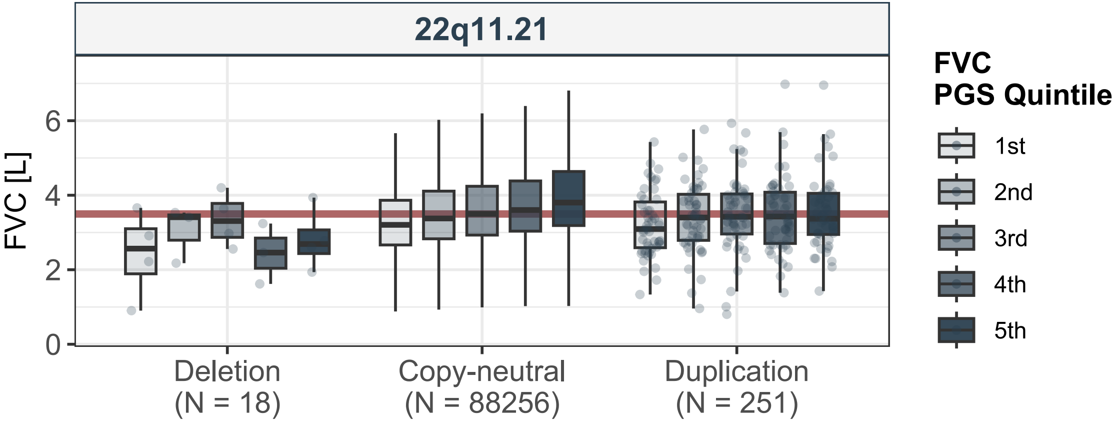
22q11.21
FVC [L]
0
2
4
6
Deletion
(N = 18)
Copy-neutral
(N = 88256)
Duplication
(N = 251)
FVC
PGS Quintile
1st
2nd
3rd
4th
5th

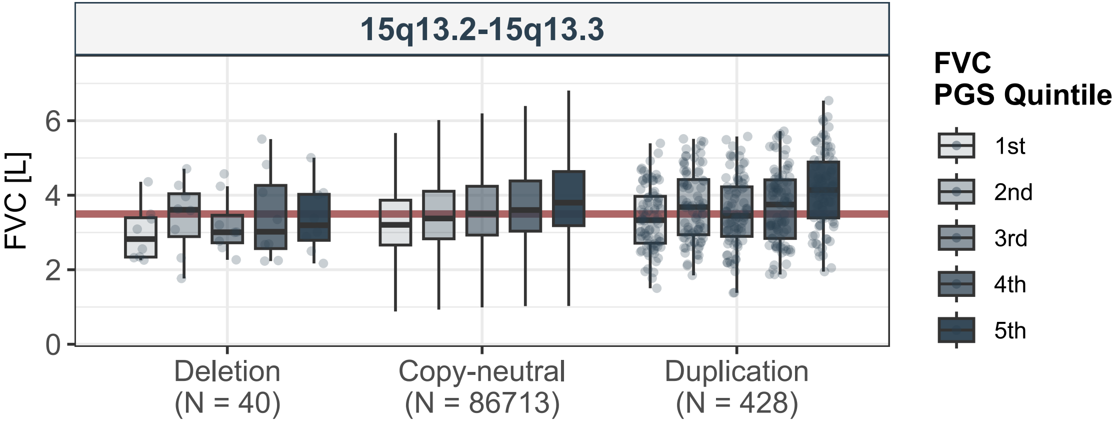

15q13.2-15q13.3
FVC [L]
0
2
4
6
Deletion
(N = 40)
Copy-neutral
(N = 86713)
Duplication
(N = 428)
FVC
PGS Quintile
1st
2nd
3rd
4th
5th

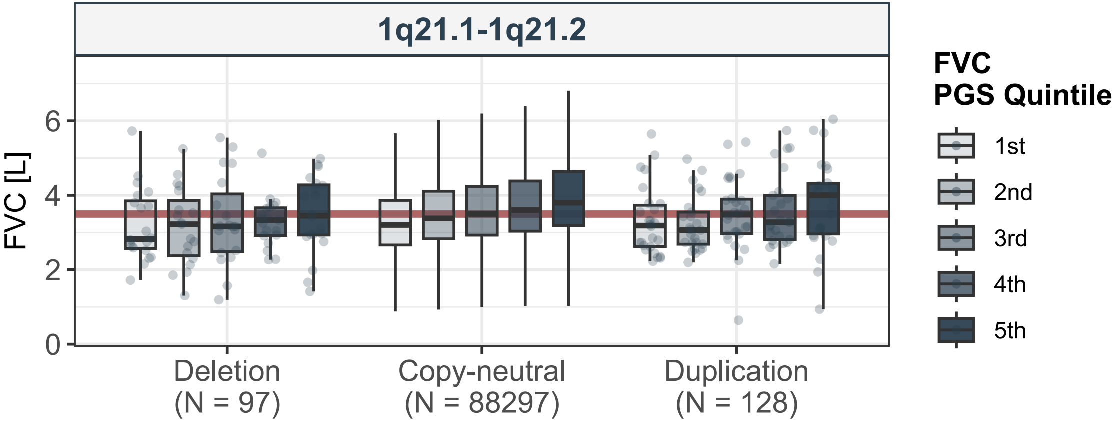
1q21.1-1q21.2
FVC [L]
0
2
4
6
Deletion
(N = 97)
Copy-neutral
(N = 88297)
Duplication
(N = 128)
FVC
PGS Quintile
1st
2nd
3rd
4th
5th

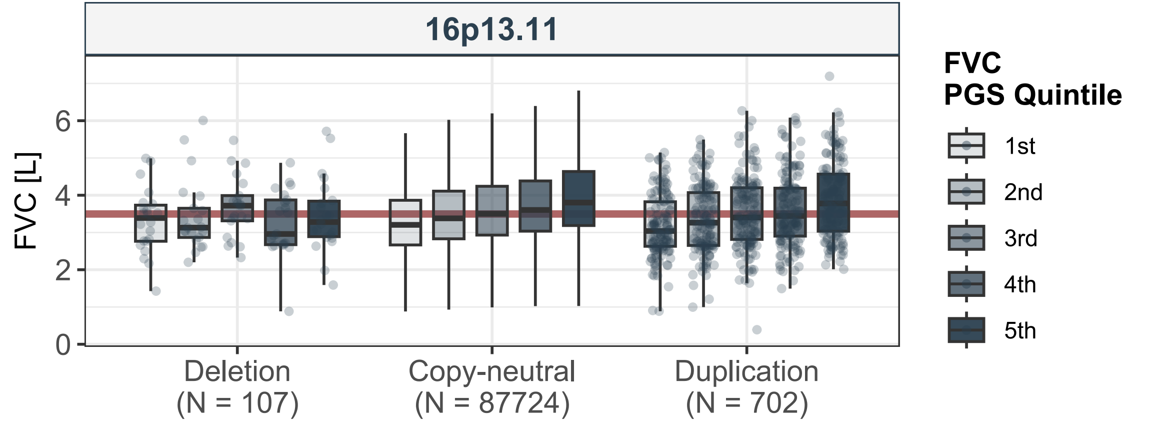
16p13.11
FVC [L]
0
2
4
6
Deletion
(N = 107)
Copy-neutral
(N = 87724)
Duplication
(N = 702)
FVC
PGS Quintile
1st
2nd
3rd
4th
5th

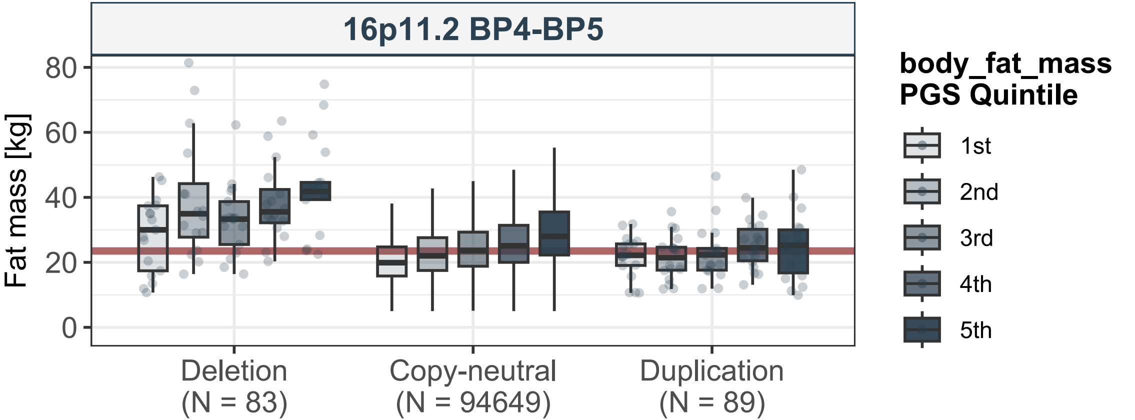

16p11.2 BP4-BP5
Fat mass [kg]
80
60
40
20
0
Deletion
(N = 83)
Copy-neutral
(N = 94649)
Duplication
(N = 89)
body_fat_mass
PGS Quintile
1st
2nd
3rd
4th
5th

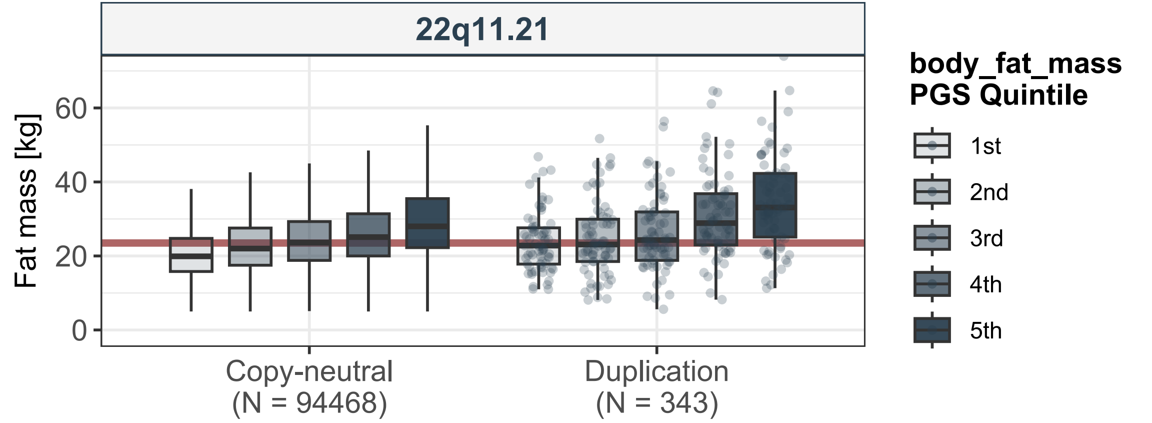
22q11.21
Fat mass [kg]
0
20
40
60
Copy-neutral
(N = 94468)
Duplication
(N = 343)
body_fat_mass
PGS Quintile
1st
2nd
3rd
4th
5th

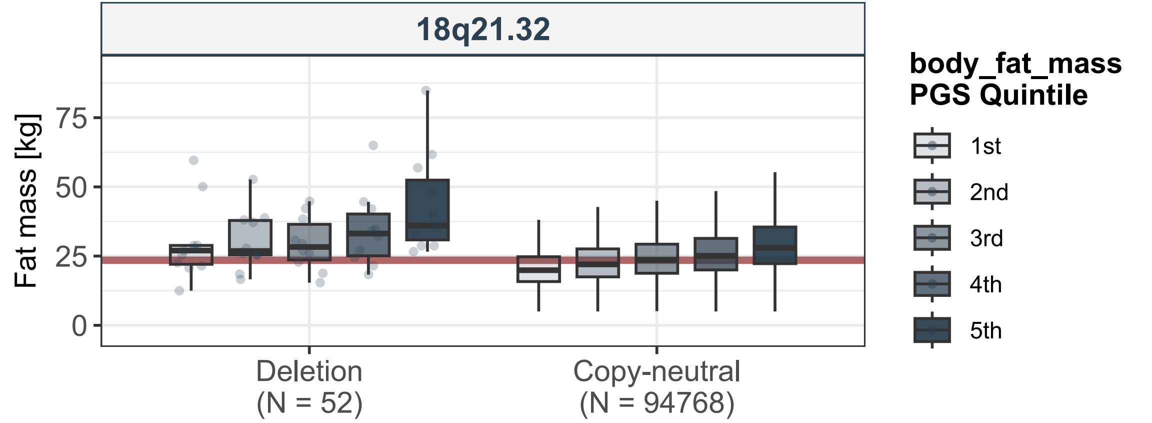
18q21.32
Fat mass [kg]
75
50
25
0
Deletion
(N = 52)
Copy-neutral
(N = 94768)
body_fat_mass
PGS Quintile
1st
2nd
3rd
4th
5th

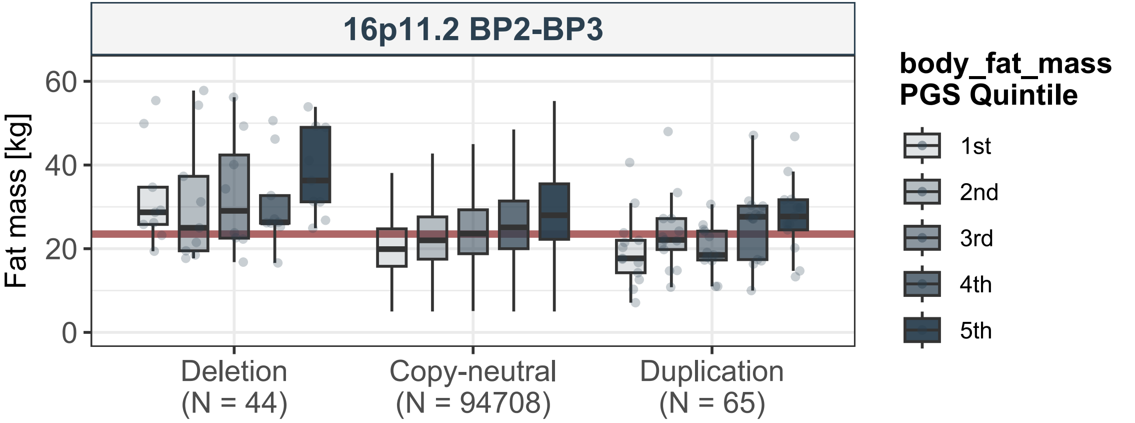
16p11.2 BP2-BP3
Fat mass [kg]
60
40
20
0
Deletion
(N = 44)
Copy-neutral
(N = 94708)
Duplication
(N = 65)
body_fat_mass
PGS Quintile
1st
2nd
3rd
4th
5th

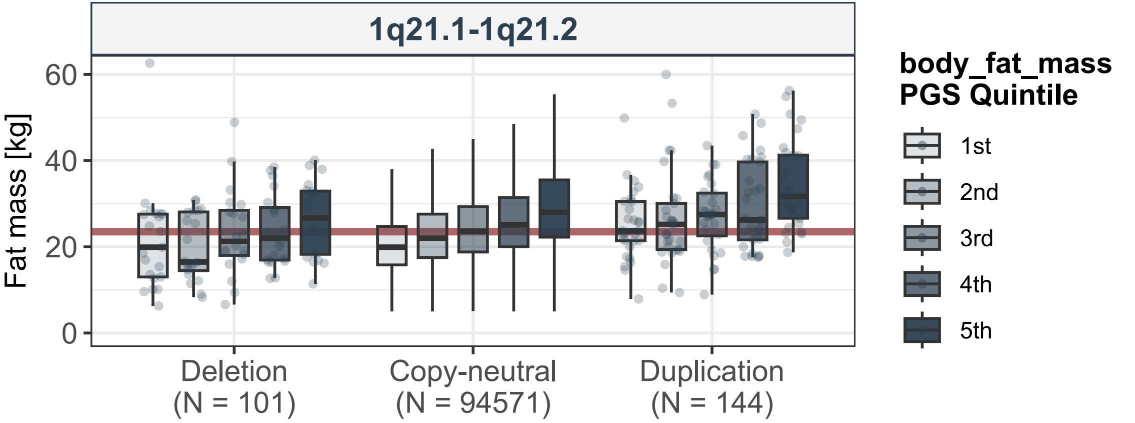

1q21.1-1q21.2
Fat mass [kg]
0
20
40
60
Deletion
(N = 101)
Copy-neutral
(N = 94571)
Duplication
(N = 144)
body_fat_mass
PGS Quintile
1st
2nd
3rd
4th
5th

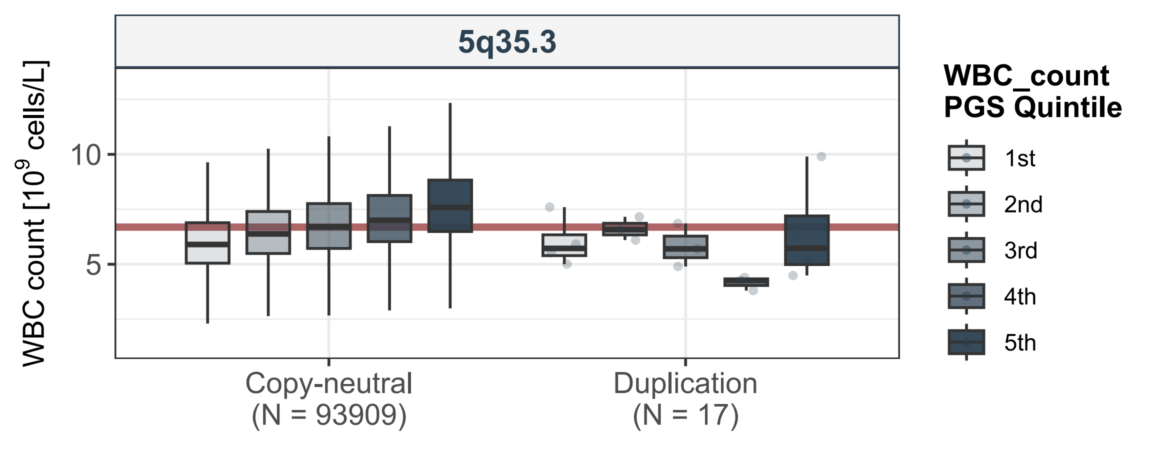
5q35.3
WBC count [$10^9$ cells/L]
10
5
Copy-neutral
(N = 93909)
Duplication
(N = 17)
WBC_count
PGS Quintile
1st
2nd
3rd
4th
5th

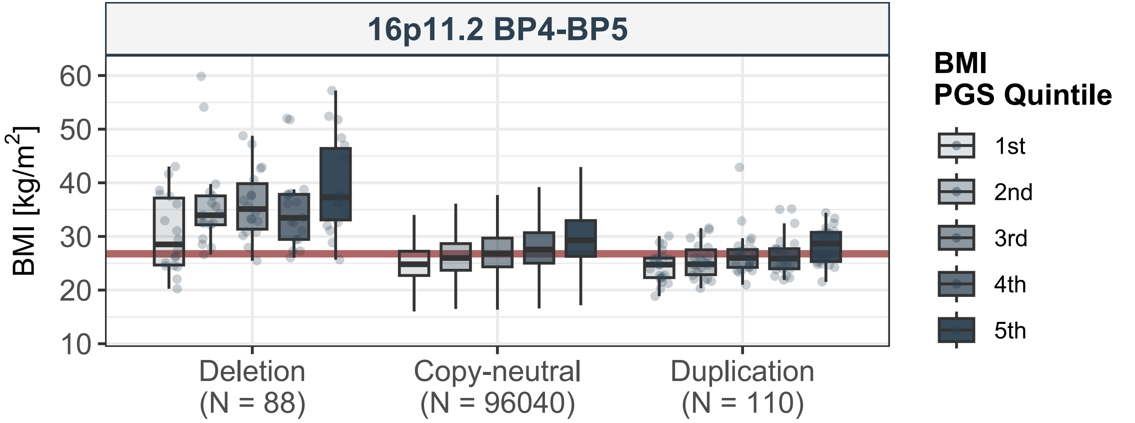
16p11.2 BP4-BP5
BMI [kg/m2]
60
50
40
30
20
10
Deletion
(N = 88)
Copy-neutral
(N = 96040)
Duplication
(N = 110)
BMI
PGS Quintile
1st
2nd
3rd
4th
5th

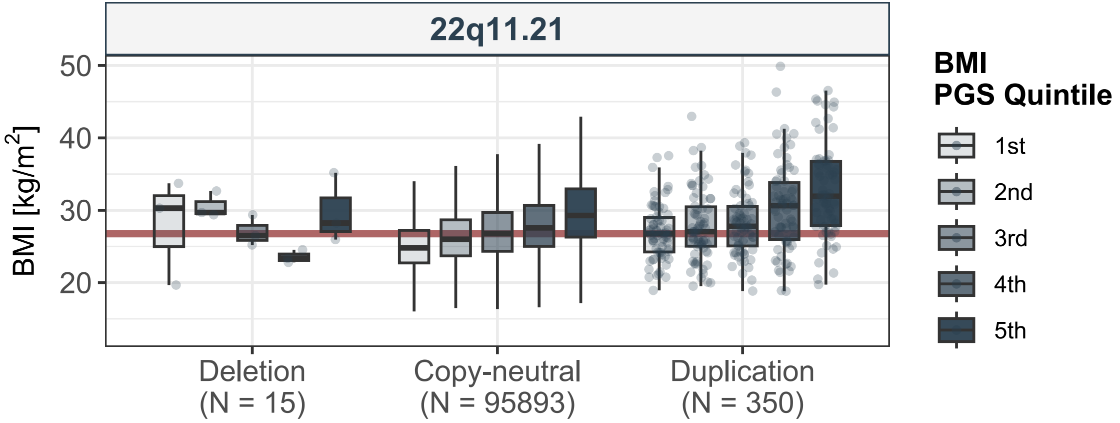

22q11.21
BMI [kg/m2]
50
40
30
20
Deletion
(N = 15)
Copy-neutral
(N = 95893)
Duplication
(N = 350)
BMI
PGS Quintile
1st
2nd
3rd
4th
5th

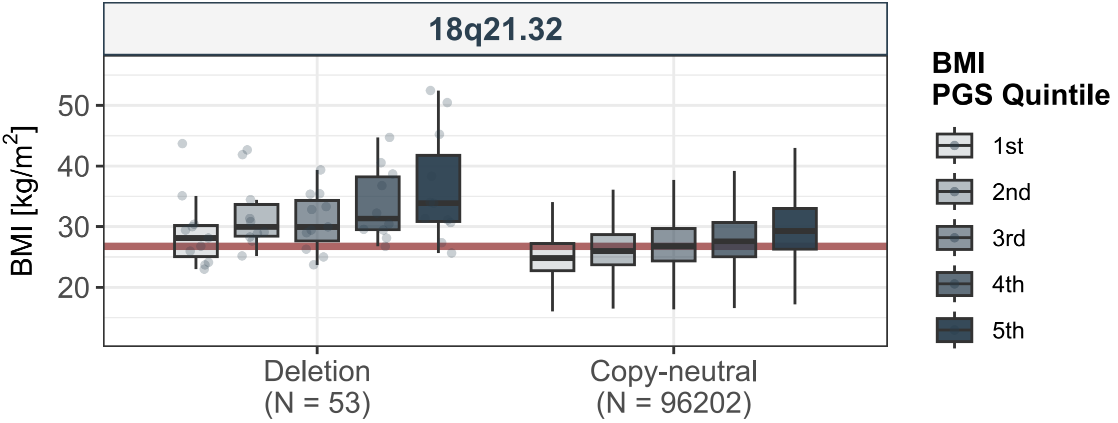
18q21.32
BMI [kg/m2]
50
40
30
20
Deletion
(N = 53)
Copy-neutral
(N = 96202)
BMI
PGS Quintile
1st
2nd
3rd
4th
5th

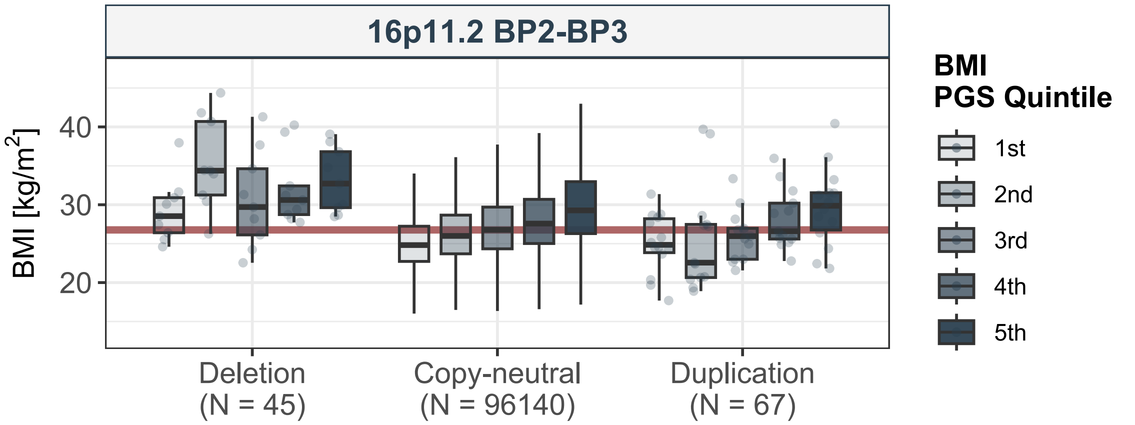
16p11.2 BP2-BP3
BMI [kg/m2]
40
30
20
Deletion
(N = 45)
Copy-neutral
(N = 96140)
Duplication
(N = 67)
BMI
PGS Quintile
1st
2nd
3rd
4th
5th

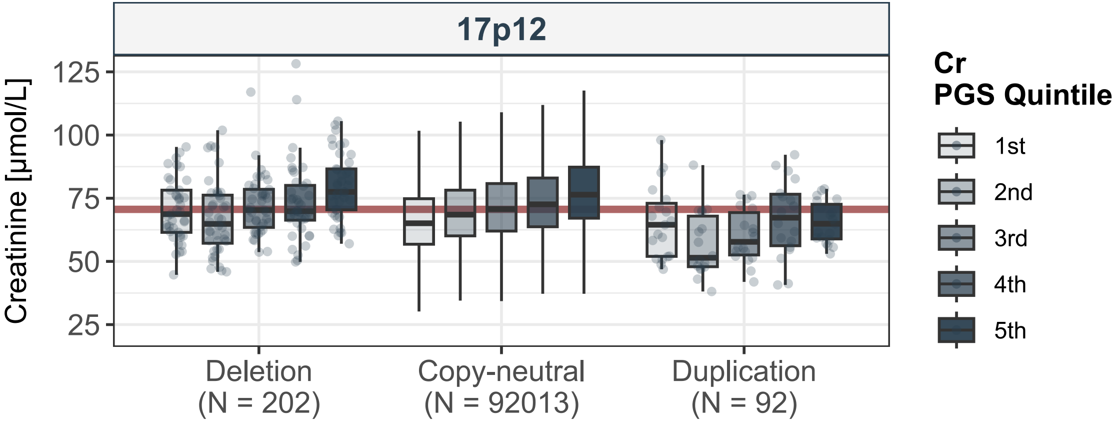
17p12
Creatinine [µmol/L]
125
100
75
50
25
Deletion
(N = 202)
Copy-neutral
(N = 92013)
Duplication
(N = 92)
Cr
PGS Quintile
1st
2nd
3rd
4th
5th

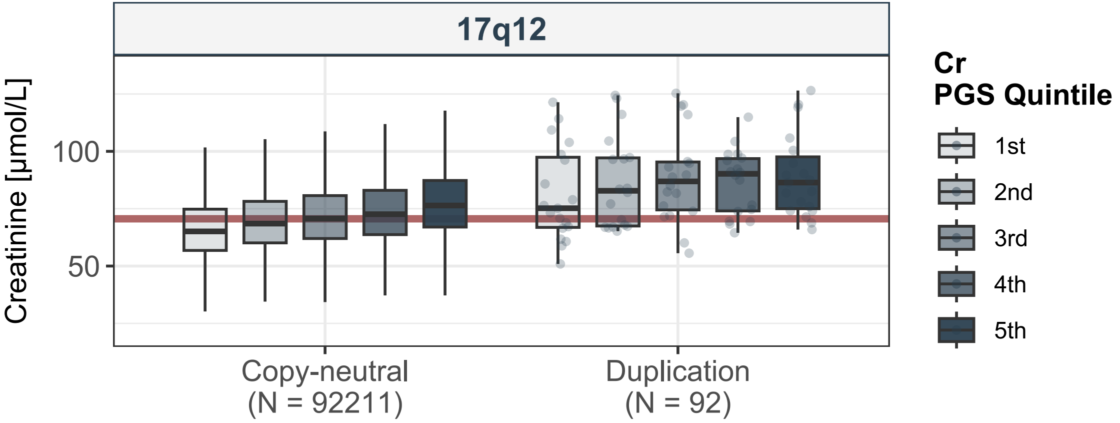
17q12
Creatinine [µmol/L]
100
50
Copy-neutral
(N = 92211)
Duplication
(N = 92)
Cr
PGS Quintile
1st
2nd
3rd
4th
5th

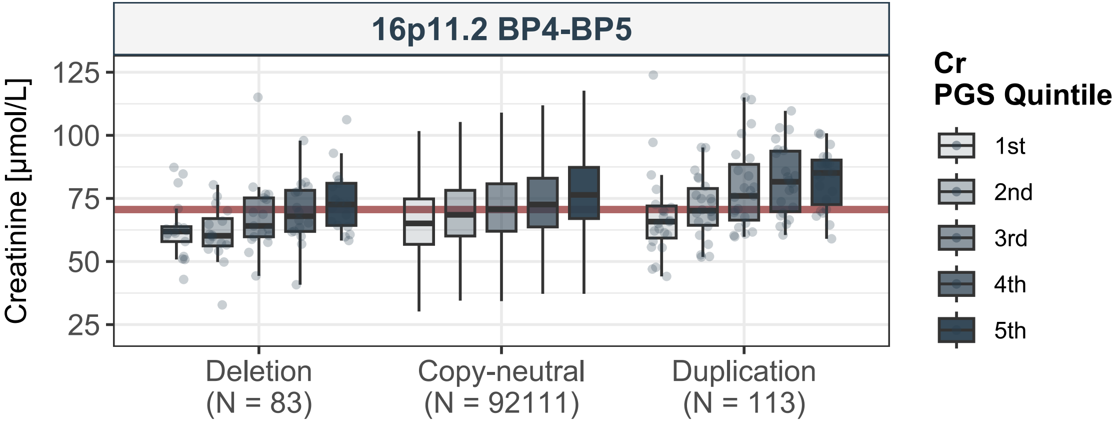

16p11.2 BP4-BP5
Creatinine [µmol/L]
125
100
75
50
25
Deletion
(N = 83)
Copy-neutral
(N = 92111)
Duplication
(N = 113)
Cr
PGS Quintile
1st
2nd
3rd
4th
5th

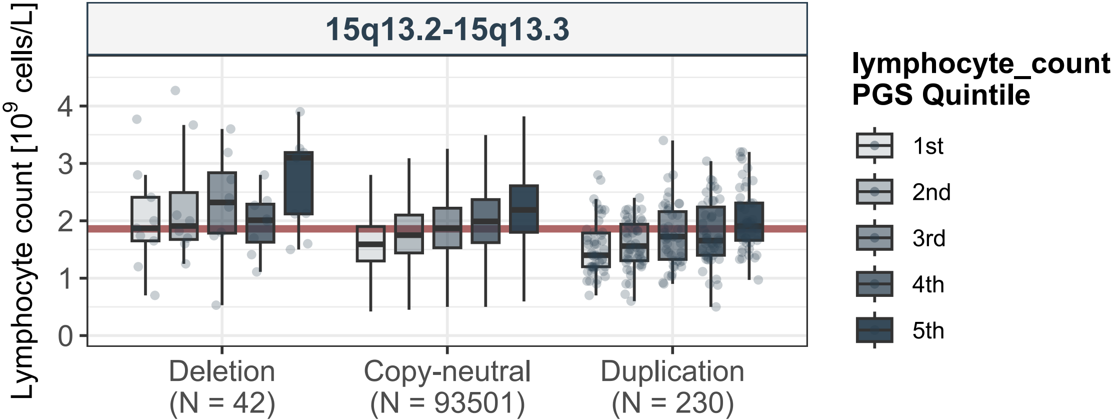

15q13.2-15q13.3
Lymphocyte count [$10^9$ cells/L]
0
1
2
3
4
Deletion
(N = 42)
Copy-neutral
(N = 93501)
Duplication
(N = 230)
lymphocyte_count
PGS Quintile
1st
2nd
3rd
4th
5th

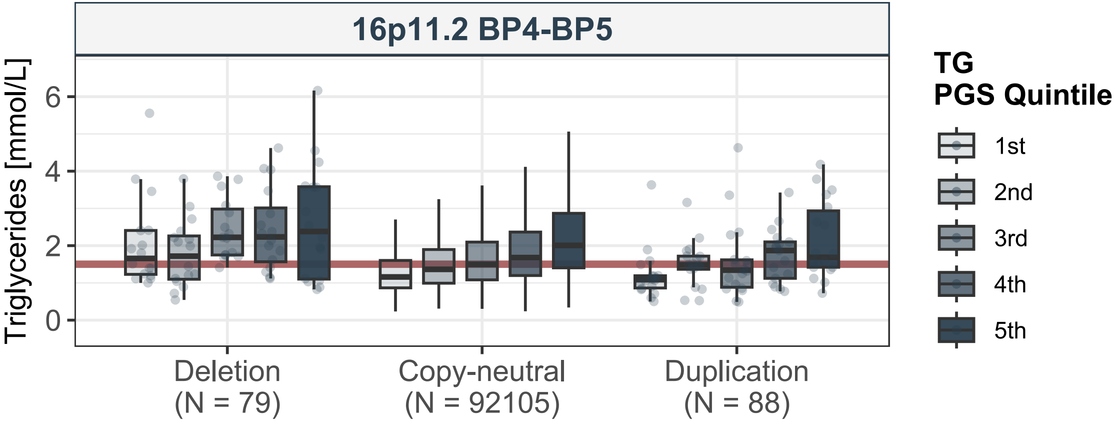
16p11.2 BP4-BP5
Triglycerides [mmol/L]
0
2
4
6
Deletion
(N = 79)
Copy-neutral
(N = 92105)
Duplication
(N = 88)
TG
PGS Quintile
1st
2nd
3rd
4th
5th

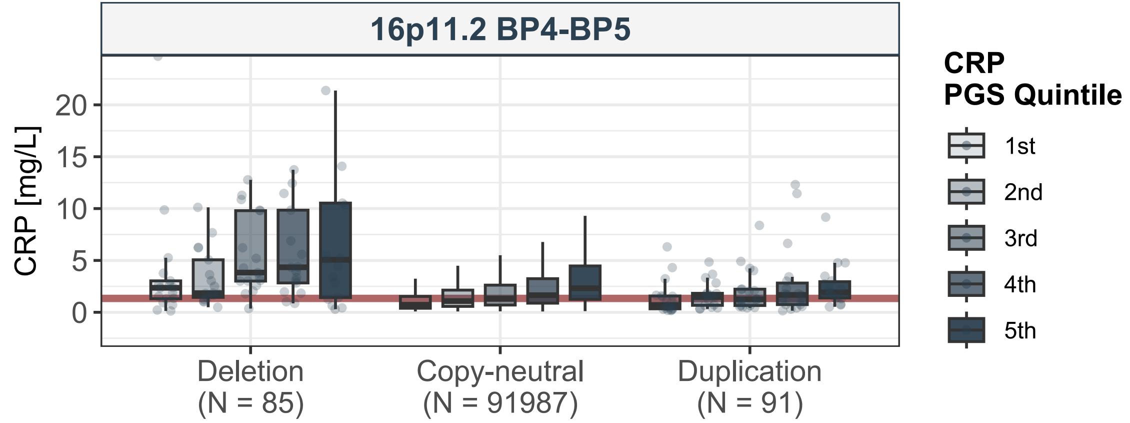

16p11.2 BP4-BP5
CRP [mg/L]
20
15
10
5
0
Deletion
(N = 85)
Copy-neutral
(N = 91987)
Duplication
(N = 91)
CRP
PGS Quintile
1st
2nd
3rd
4th
5th

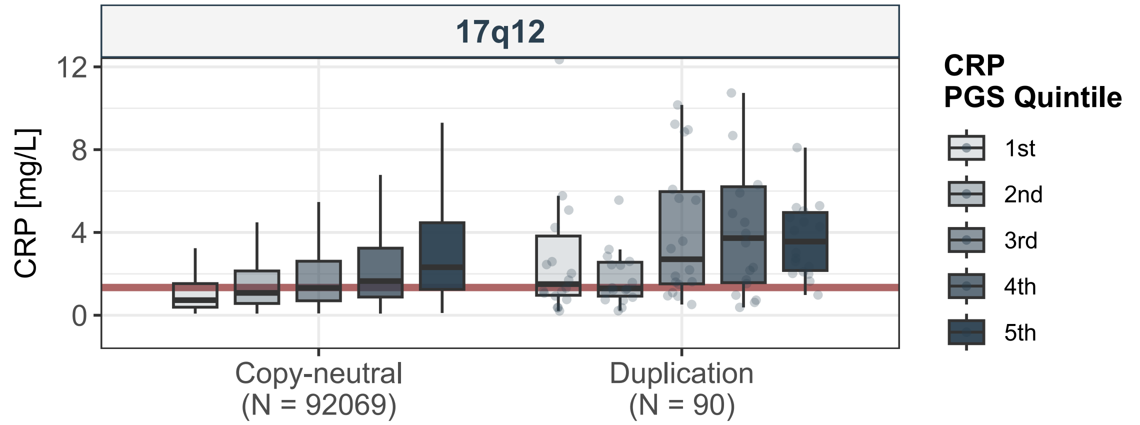

17q12
CRP [mg/L]
12
8
4
0
Copy-neutral
(N = 92069)
Duplication
(N = 90)
CRP
PGS Quintile
1st
2nd
3rd
4th
5th

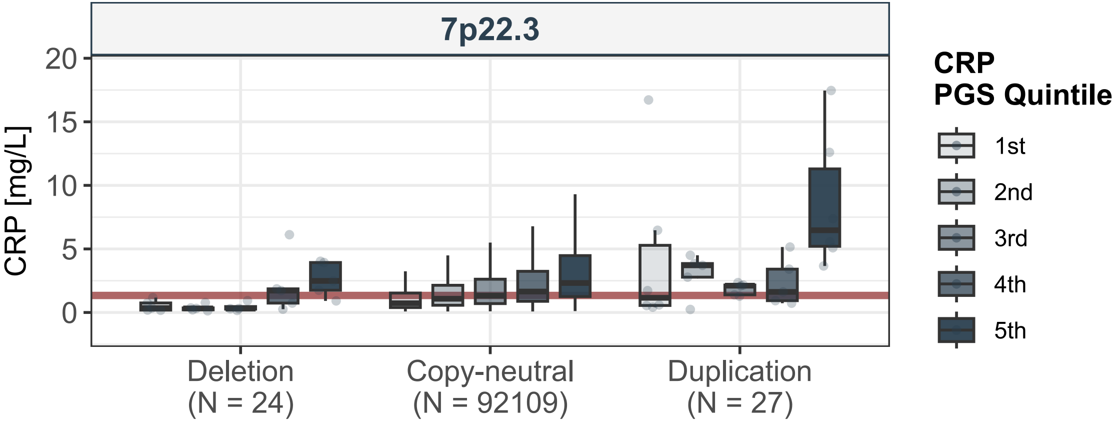
7p22.3
CRP [mg/L]
20
15
10
5
0
Deletion
(N = 24)
Copy-neutral
(N = 92109)
Duplication
(N = 27)
CRP
PGS Quintile
1st
2nd
3rd
4th
5th

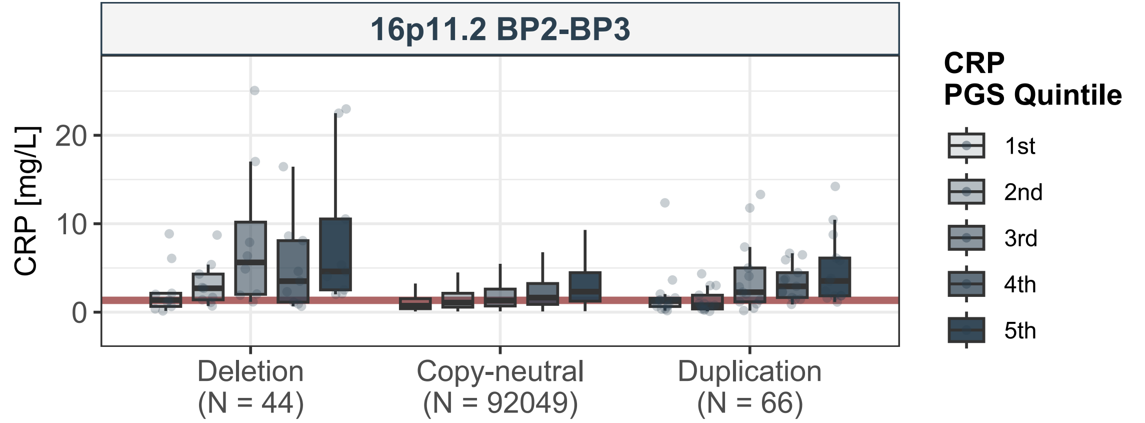

16p11.2 BP2-BP3
CRP [mg/L]
20
10
0
Deletion
(N = 44)
Copy-neutral
(N = 92049)
Duplication
(N = 66)
CRP
PGS Quintile
1st
2nd
3rd
4th
5th

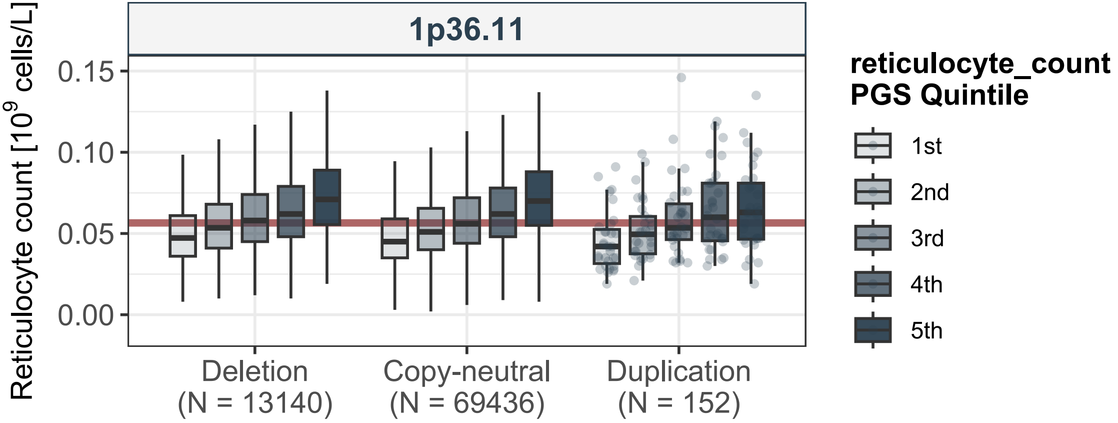

1p36.11
Reticulocyte count [10^9 cells/L]
0.15
0.10
0.05
0.00
Deletion
(N = 13140)
Copy-neutral
(N = 69436)
Duplication
(N = 152)
reticulocyte_count
PGS Quintile
1st
2nd
3rd
4th
5th

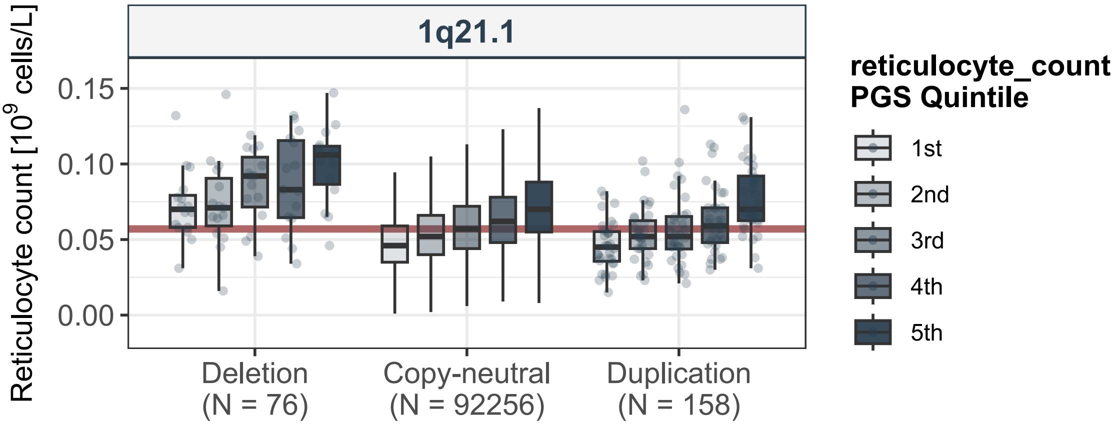

1q21.1
Reticulocyte count [10^9 cells/L]
0.15
0.10
0.05
0.00
Deletion
(N = 76)
Copy-neutral
(N = 92256)
Duplication
(N = 158)
reticulocyte_count
PGS Quintile
1st
2nd
3rd
4th
5th

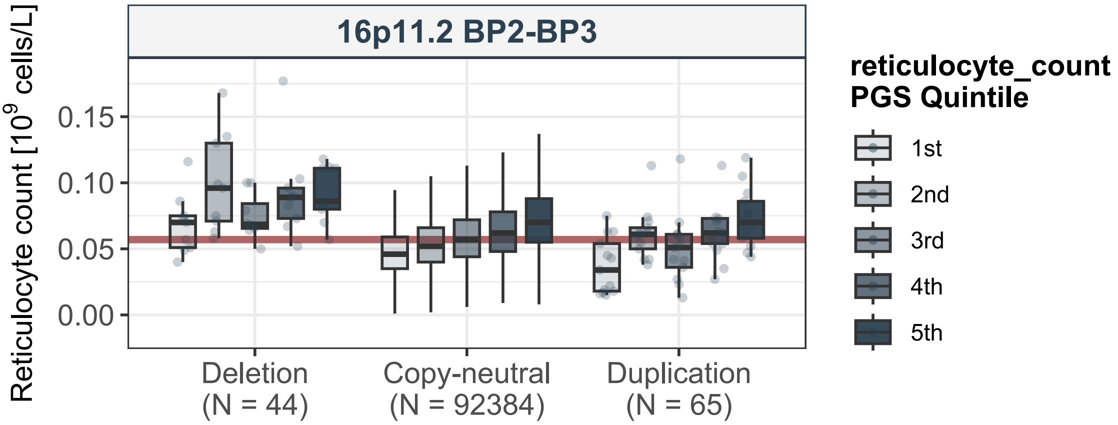
16p11.2 BP2-BP3
Reticulocyte count [10^9 cells/L]
0.15
0.10
0.05
0.00
Deletion
(N = 44)
Copy-neutral
(N = 92384)
Duplication
(N = 65)
reticulocyte_count
PGS Quintile
1st
2nd
3rd
4th
5th

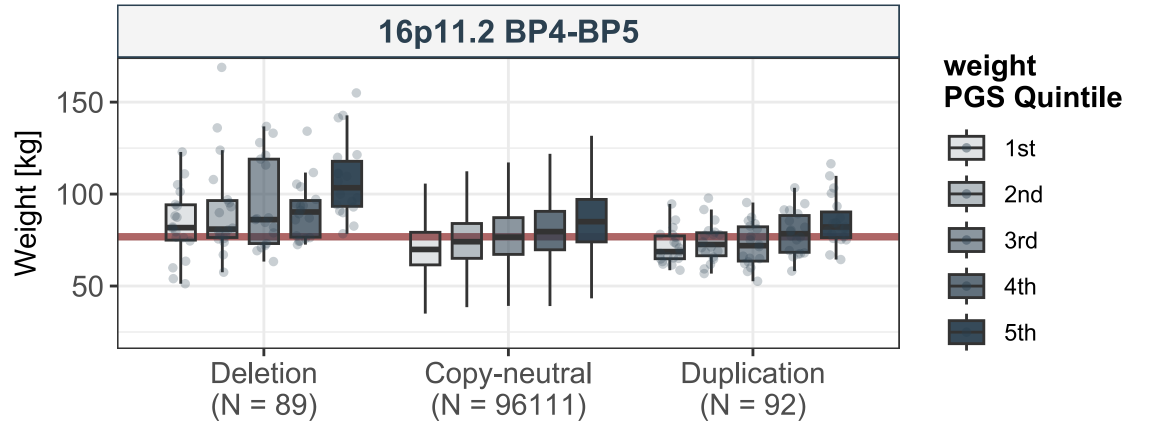

16p11.2 BP4-BP5
Weight [kg]
150
100
50
Deletion
(N = 89)
Copy-neutral
(N = 96111)
Duplication
(N = 92)
weight
PGS Quintile
1st
2nd
3rd
4th
5th

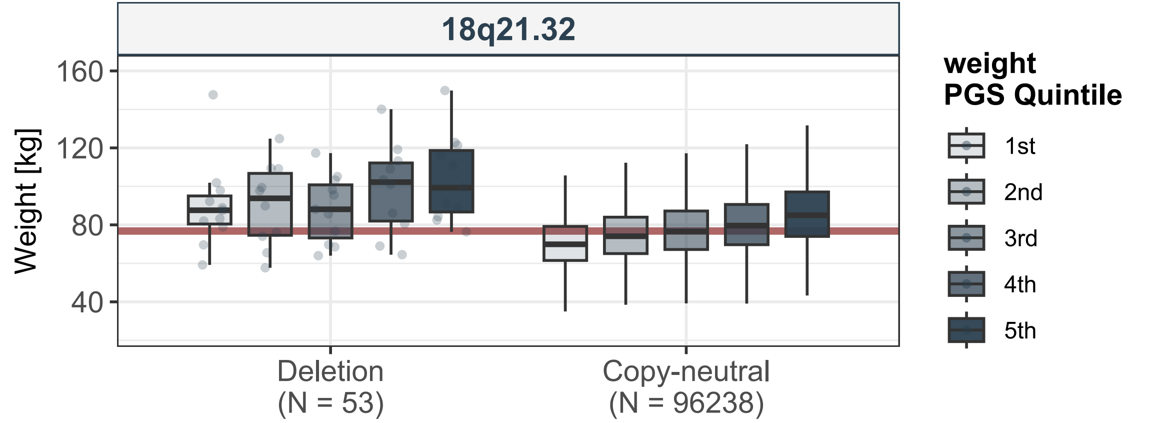

18q21.32
Weight [kg]
160
120
80
40
Deletion
(N = 53)
Copy-neutral
(N = 96238)
weight
PGS Quintile
1st
2nd
3rd
4th
5th

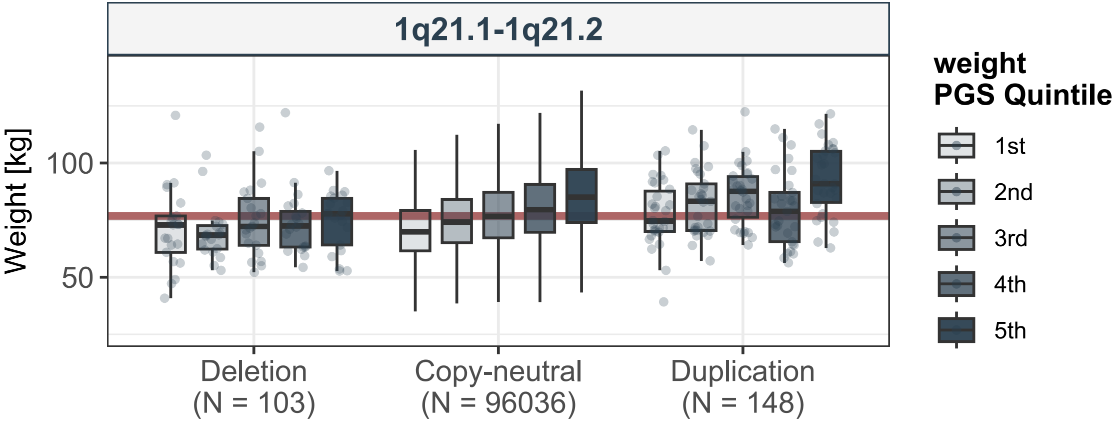

1q21.1-1q21.2
Weight [kg]
100
50
Deletion
(N = 103)
Copy-neutral
(N = 96036)
Duplication
(N = 148)
weight
PGS Quintile
1st
2nd
3rd
4th
5th

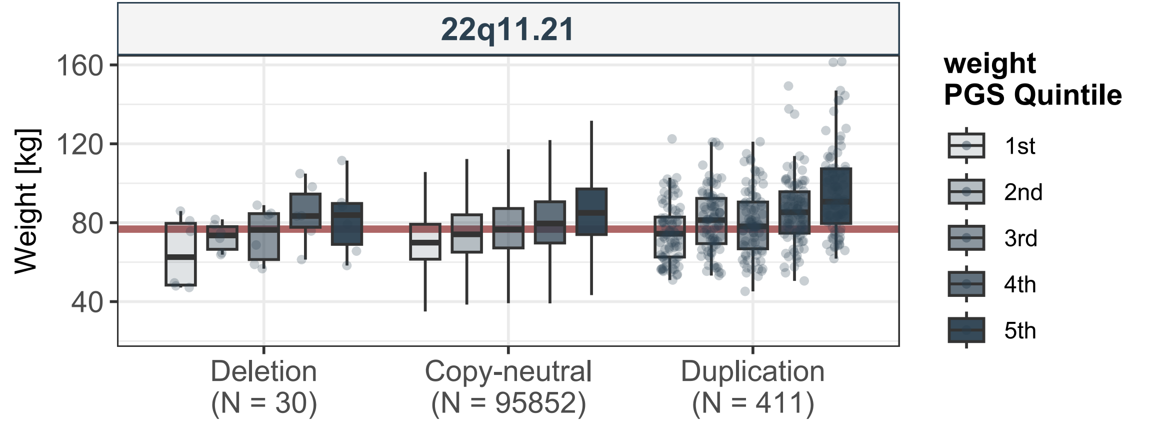
22q11.21
Weight [kg]
160
120
80
40
Deletion
(N = 30)
Copy-neutral
(N = 95852)
Duplication
(N = 411)
weight
PGS Quintile
1st
2nd
3rd
4th
5th

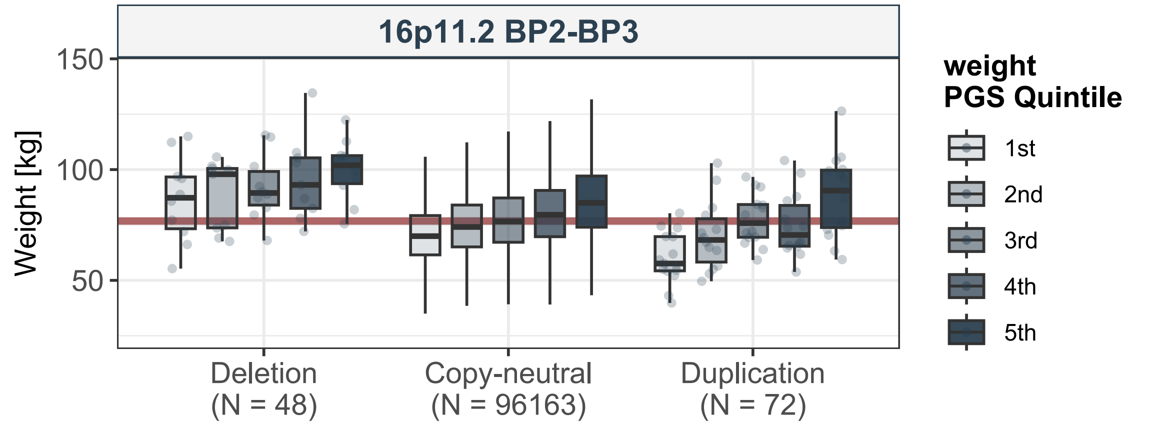

16p11.2 BP2-BP3
Weight [kg]
150
100
50
Deletion
(N = 48)
Copy-neutral
(N = 96163)
Duplication
(N = 72)
weight
PGS Quintile
1st
2nd
3rd
4th
5th

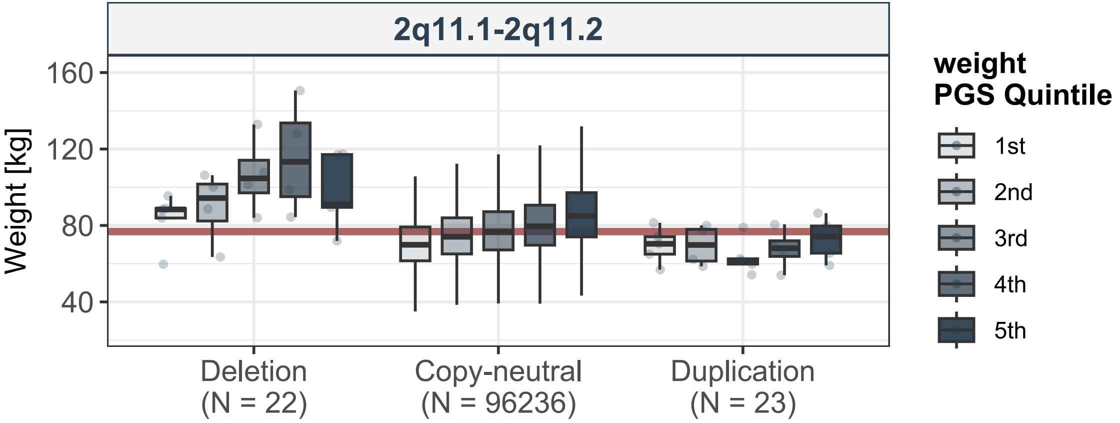

2q11.1-2q11.2
Weight [kg]
160
120
80
40
Deletion
(N = 22)
Copy-neutral
(N = 96236)
Duplication
(N = 23)
weight
PGS Quintile
1st
2nd
3rd
4th
5th

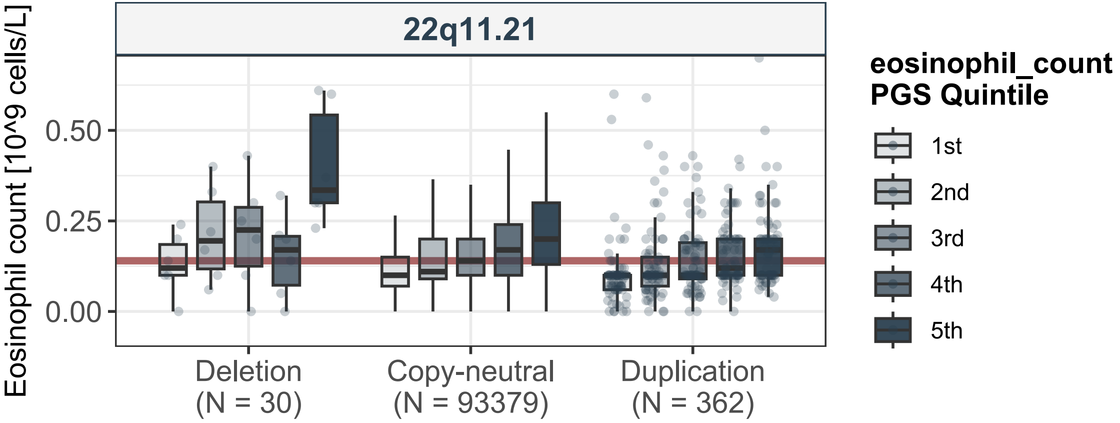
22q11.21
Eosinophil count [10^9 cells/L]
0.50
0.25
0.00
Deletion
(N = 30)
Copy-neutral
(N = 93379)
Duplication
(N = 362)
eosinophil_count
PGS Quintile
1st
2nd
3rd
4th
5th

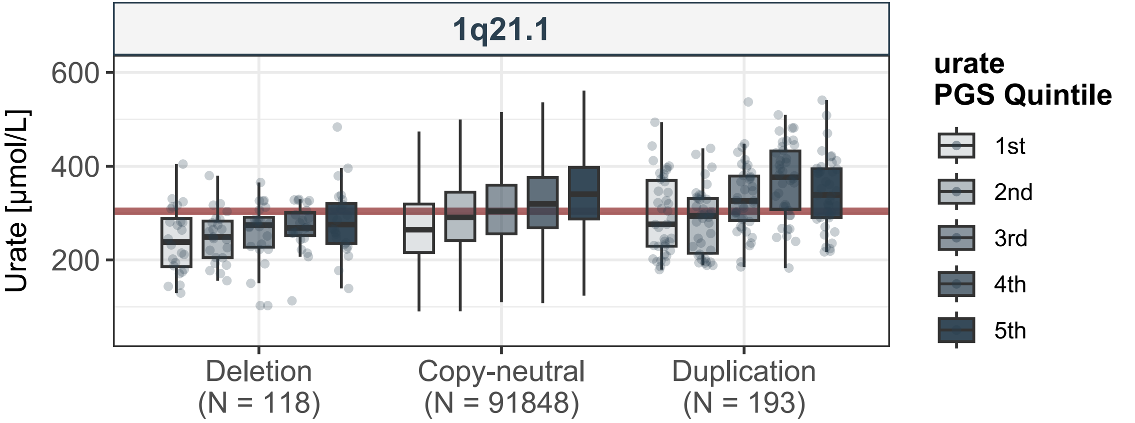
1q21.1
Urate [µmol/L]
600
400
200
Deletion
(N = 118)
Copy-neutral
(N = 91848)
Duplication
(N = 193)
urate
PGS Quintile
1st
2nd
3rd
4th
5th

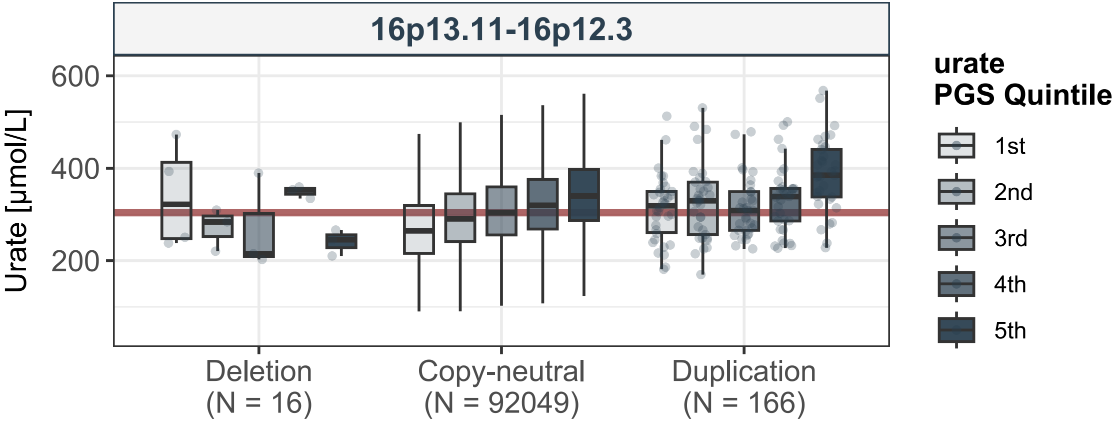
16p13.11-16p12.3
Urate [µmol/L]
600
400
200
Deletion
(N = 16)
Copy-neutral
(N = 92049)
Duplication
(N = 166)
urate
PGS Quintile
1st
2nd
3rd
4th
5th

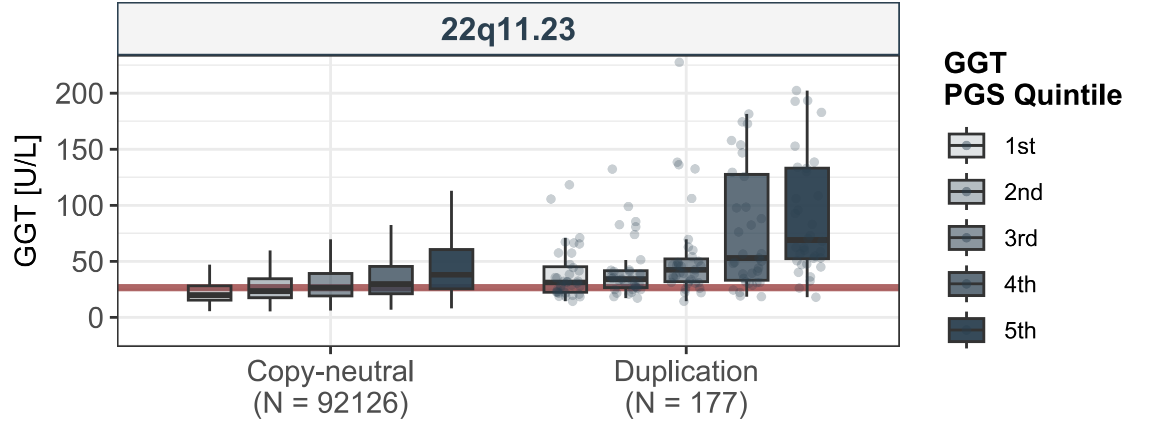

22q11.23
GGT [U/L]
200
150
100
50
0
Copy-neutral
(N = 92126)
Duplication
(N = 177)
GGT
PGS Quintile
1st
2nd
3rd
4th
5th

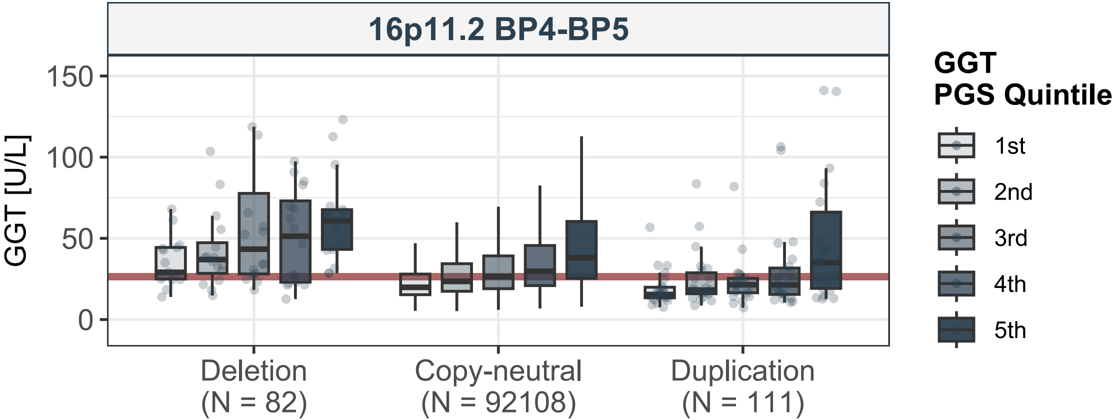
16p11.2 BP4-BP5
GGT [U/L]
150
100
50
0
Deletion
(N = 82)
Copy-neutral
(N = 92108)
Duplication
(N = 111)
GGT
PGS Quintile
1st
2nd
3rd
4th
5th

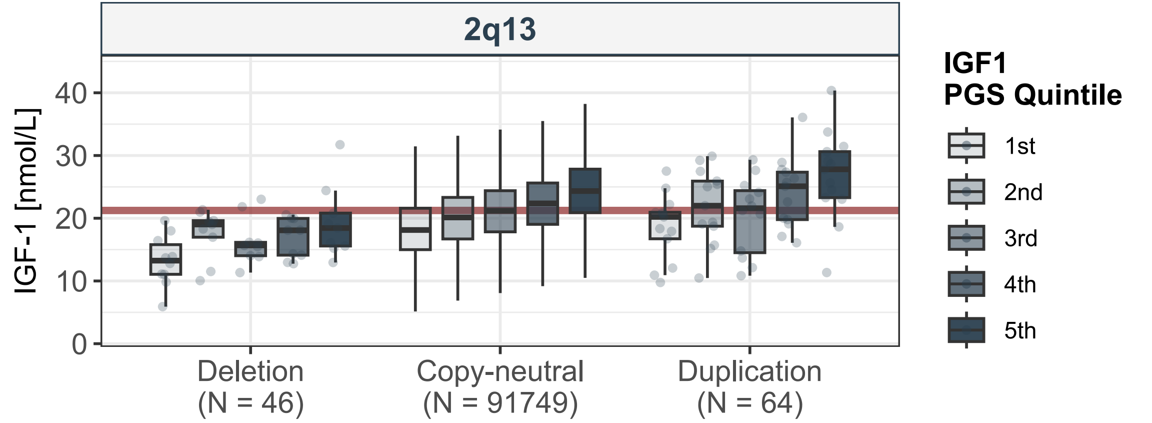
2q13
IGF-1 [nmol/L]
0
10
20
30
40
Deletion
(N = 46)
Copy-neutral
(N = 91749)
Duplication
(N = 64)
IGF1
PGS Quintile
1st
2nd
3rd
4th
5th

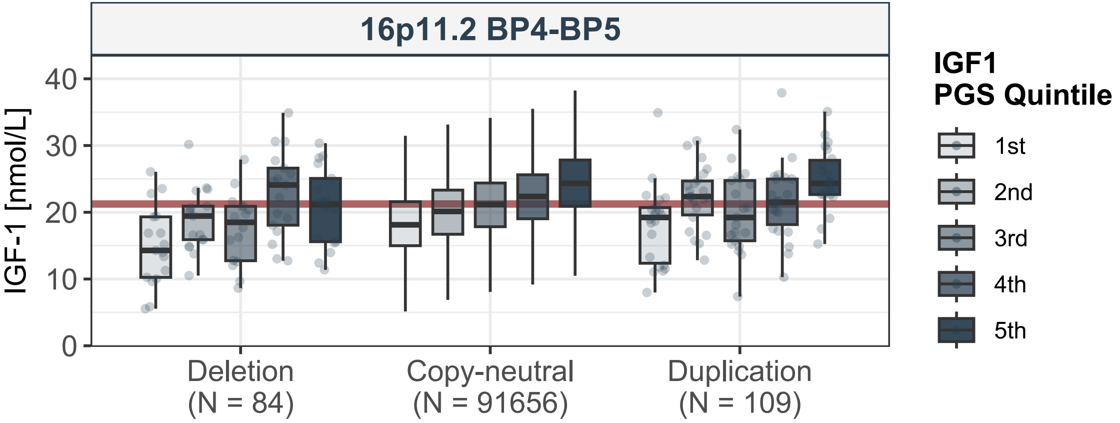

16p11.2 BP4-BP5
IGF-1 [nmol/L]
0
10
20
30
40
Deletion
(N = 84)
Copy-neutral
(N = 91656)
Duplication
(N = 109)
IGF1
PGS Quintile
1st
2nd
3rd
4th
5th

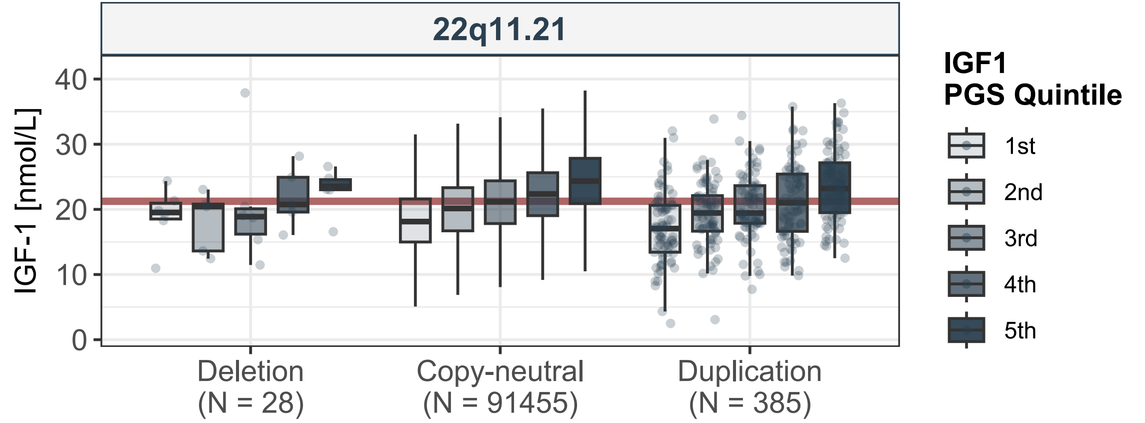
22q11.21
IGF-1 [nmol/L]
0
10
20
30
40
Deletion
(N = 28)
Copy-neutral
(N = 91455)
Duplication
(N = 385)
IGF1
PGS Quintile
1st
2nd
3rd
4th
5th

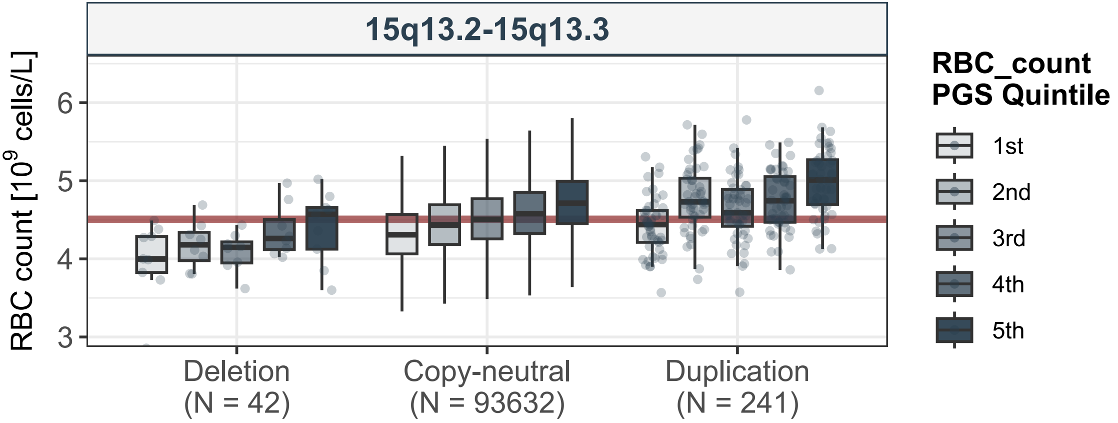

15q13.2-15q13.3
RBC count [10^9 cells/L]
6
5
4
3
Deletion
(N = 42)
Copy-neutral
(N = 93632)
Duplication
(N = 241)
RBC_count
PGS Quintile
1st
2nd
3rd
4th
5th

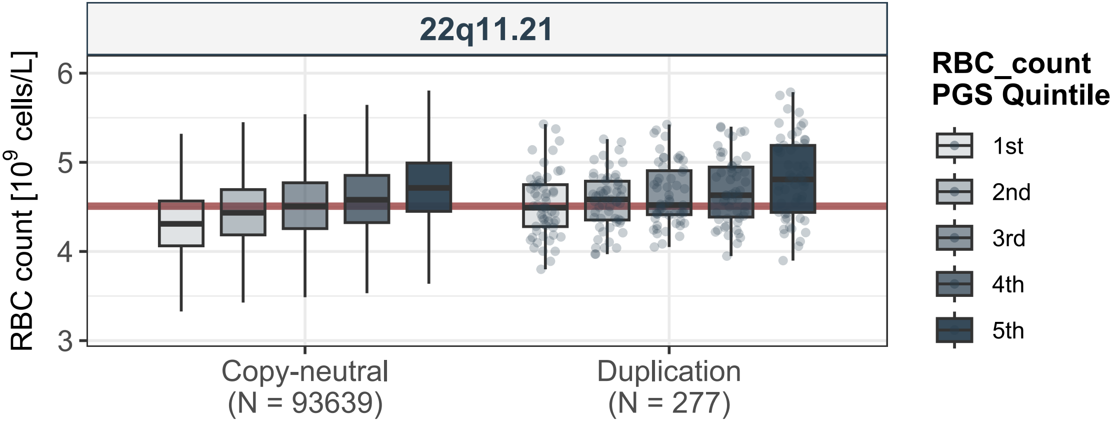
22q11.21
RBC count [10^9 cells/L]
6
5
4
3
Copy-neutral
(N = 93639)
Duplication
(N = 277)
RBC_count
PGS Quintile
1st
2nd
3rd
4th
5th

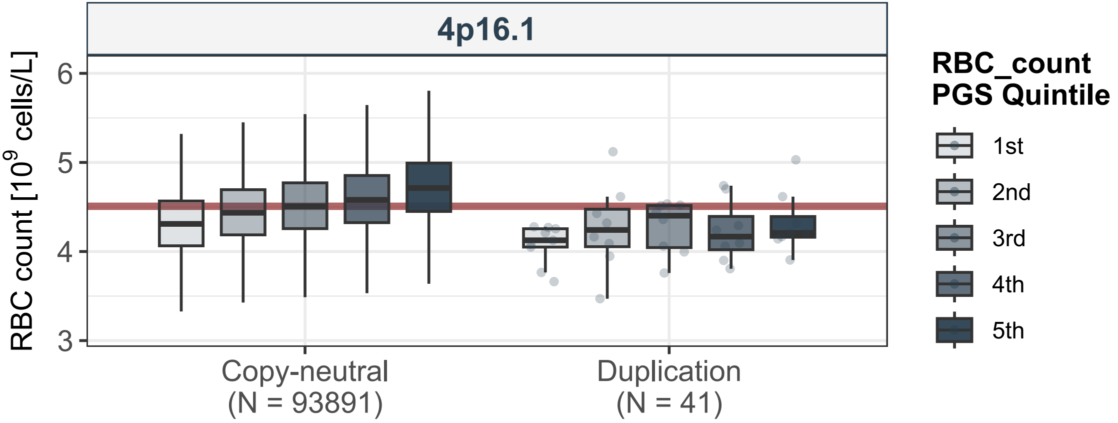
4p16.1
RBC count [10^9 cells/L]
6
5
4
3
Copy-neutral
(N = 93891)
Duplication
(N = 41)
RBC_count
PGS Quintile
1st
2nd
3rd
4th
5th

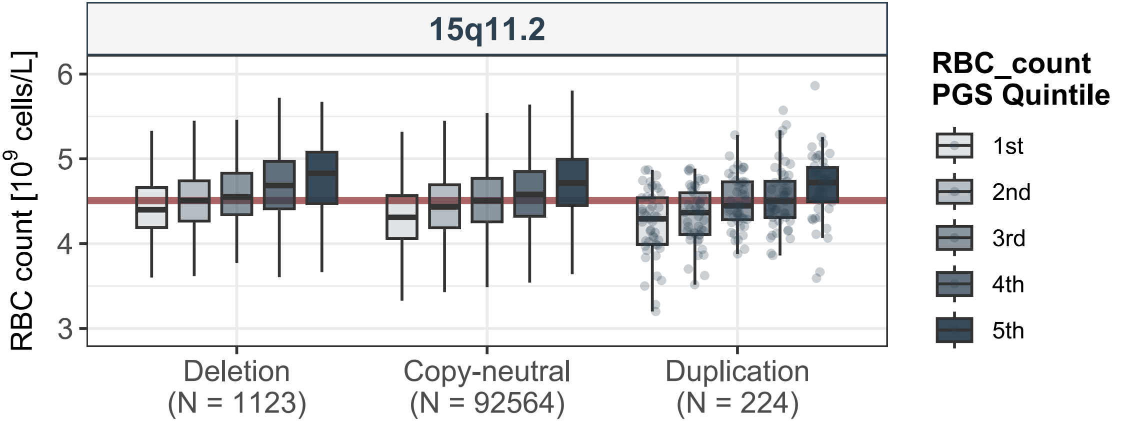
15q11.2
RBC count [10^9 cells/L]
6
5
4
3
Deletion
(N = 1123)
Copy-neutral
(N = 92564)
Duplication
(N = 224)
RBC_count
PGS Quintile
1st
2nd
3rd
4th
5th

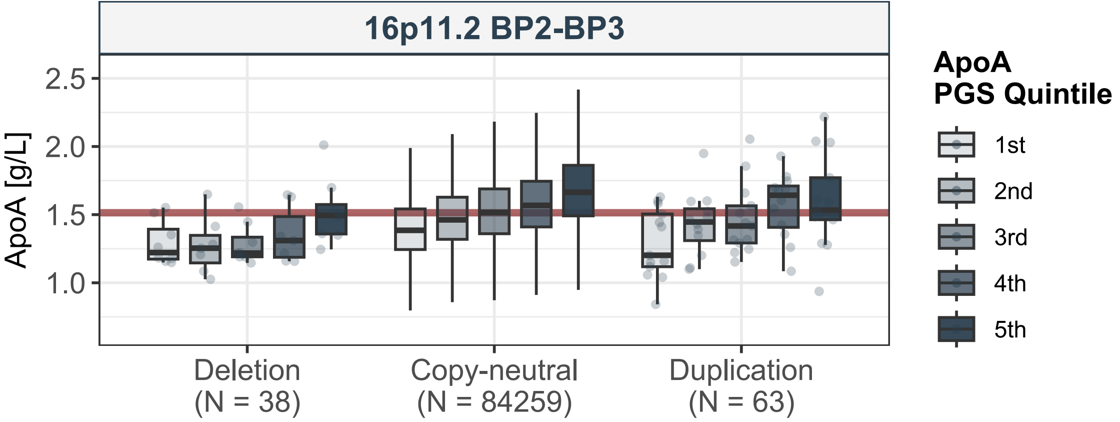
16p11.2 BP2-BP3
ApoA [g/L]
2.5
2.0
1.5
1.0
Deletion
(N = 38)
Copy-neutral
(N = 84259)
Duplication
(N = 63)
ApoA
PGS Quintile
1st
2nd
3rd
4th
5th

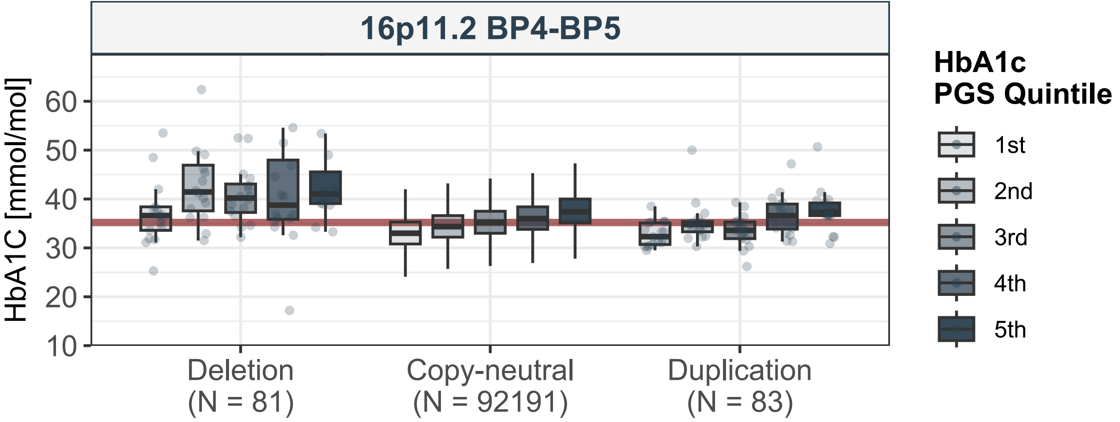
16p11.2 BP4-BP5
HbA1C [mmol/mol]
60
50
40
30
20
10
Deletion
(N = 81)
Copy-neutral
(N = 92191)
Duplication
(N = 83)
HbA1c
PGS Quintile
1st
2nd
3rd
4th
5th

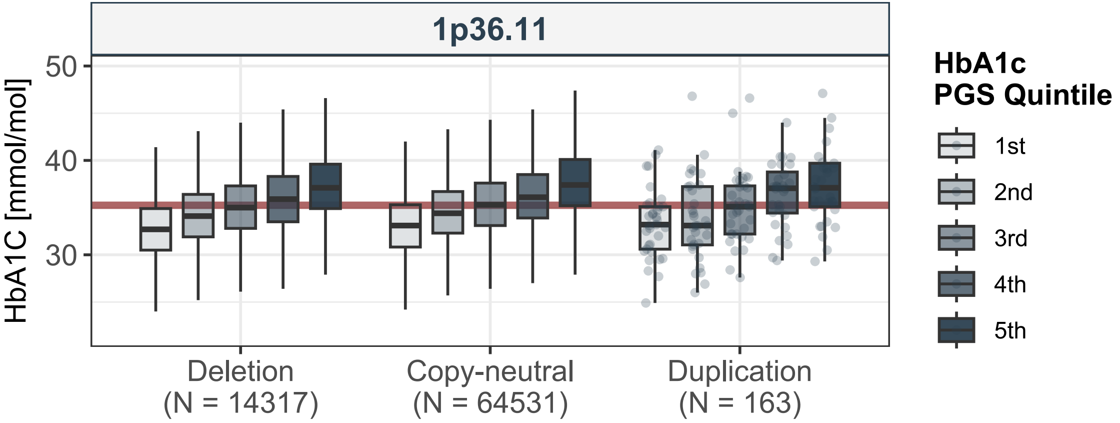

1p36.11
HbA1C [mmol/mol]
50
40
30
Deletion
(N = 14317)
Copy-neutral
(N = 64531)
Duplication
(N = 163)
HbA1c
PGS Quintile
1st
2nd
3rd
4th
5th

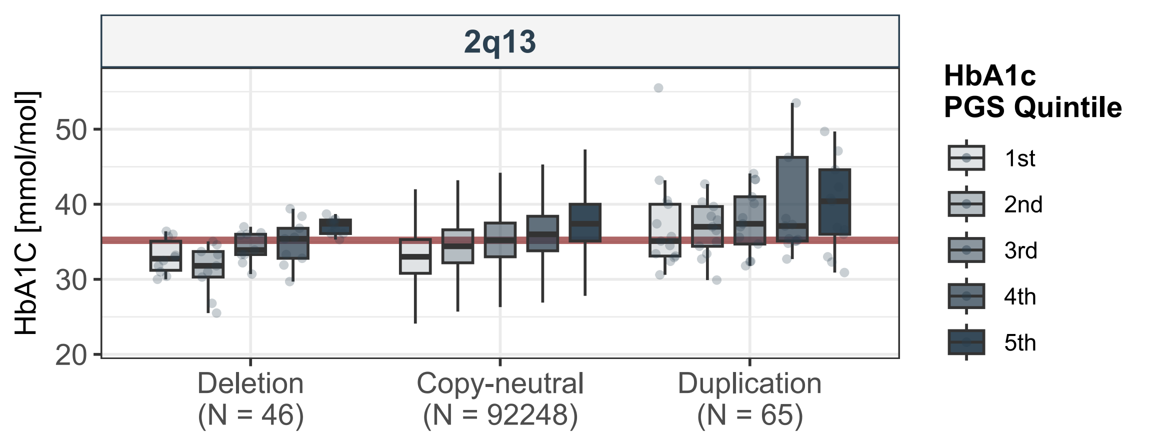
2q13
HbA1C [mmol/mol]
50
40
30
20
Deletion
(N = 46)
Copy-neutral
(N = 92248)
Duplication
(N = 65)
HbA1c
PGS Quintile
1st
2nd
3rd
4th
5th

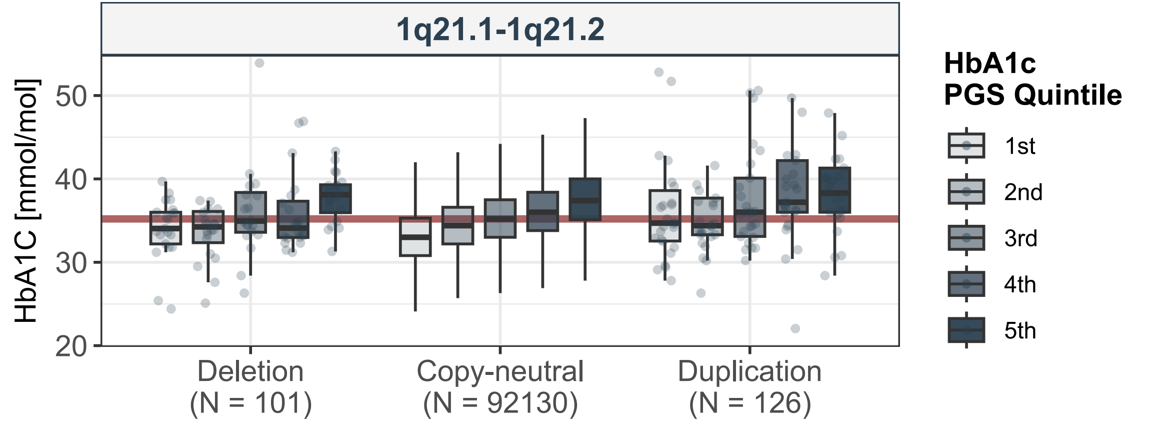

1q21.1-1q21.2
HbA1C [mmol/mol]
50
40
30
20
Deletion
(N = 101)
Copy-neutral
(N = 92130)
Duplication
(N = 126)
HbA1c
PGS Quintile
1st
2nd
3rd
4th
5th

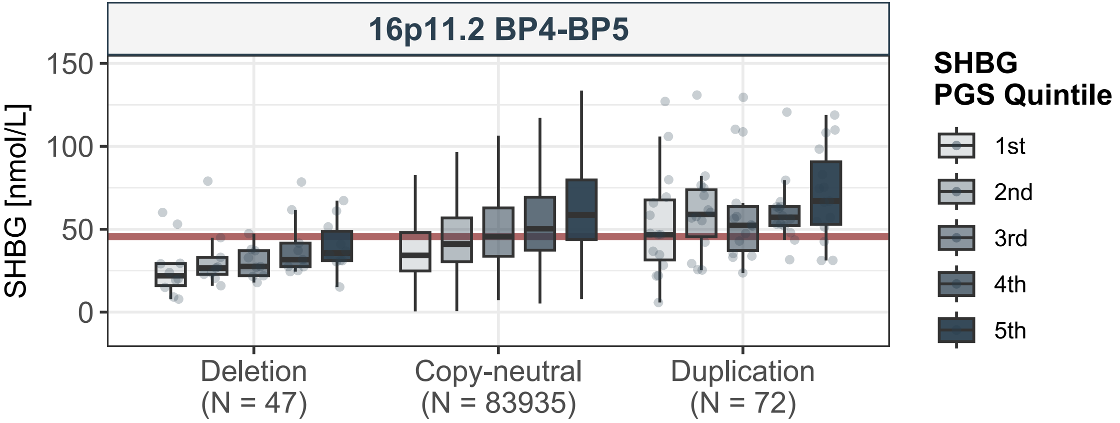

16p11.2 BP4-BP5
SHBG [nmol/L]
150
100
50
0
Deletion
(N = 47)
Copy-neutral
(N = 83935)
Duplication
(N = 72)
SHBG
PGS Quintile
1st
2nd
3rd
4th
5th

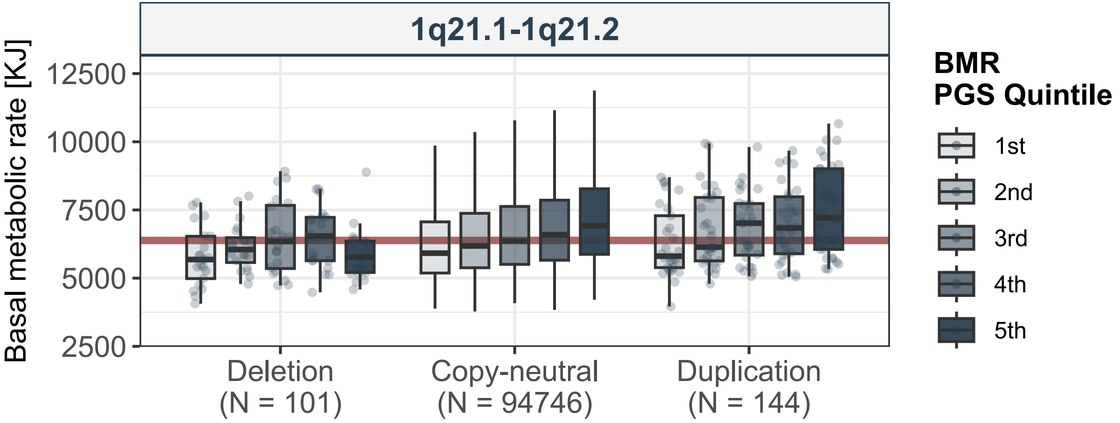
1q21.1-1q21.2
Basal metabolic rate [KJ]
12500
10000
7500
5000
2500
Deletion
(N = 101)
Copy-neutral
(N = 94746)
Duplication
(N = 144)
BMR
PGS Quintile
1st
2nd
3rd
4th
5th

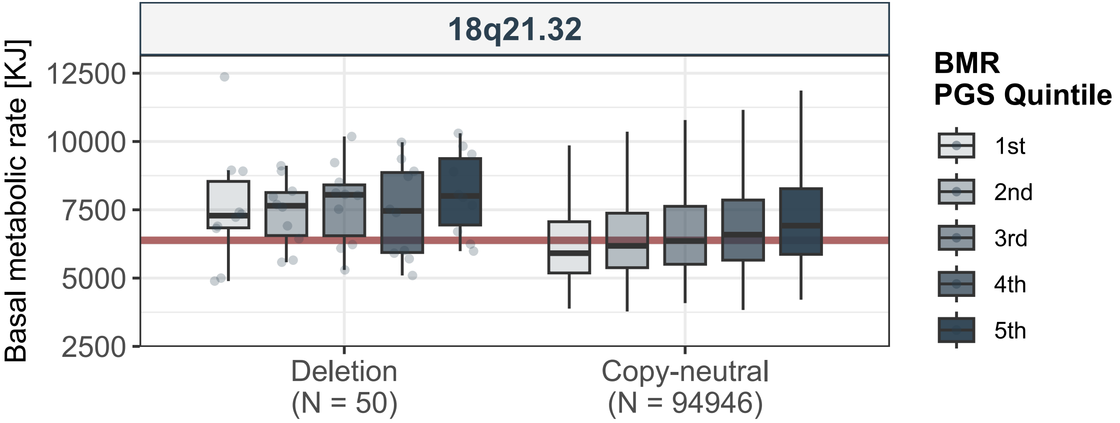
18q21.32
Basal metabolic rate [KJ]
12500
10000
7500
5000
2500
Deletion
(N = 50)
Copy-neutral
(N = 94946)
BMR
PGS Quintile
1st
2nd
3rd
4th
5th

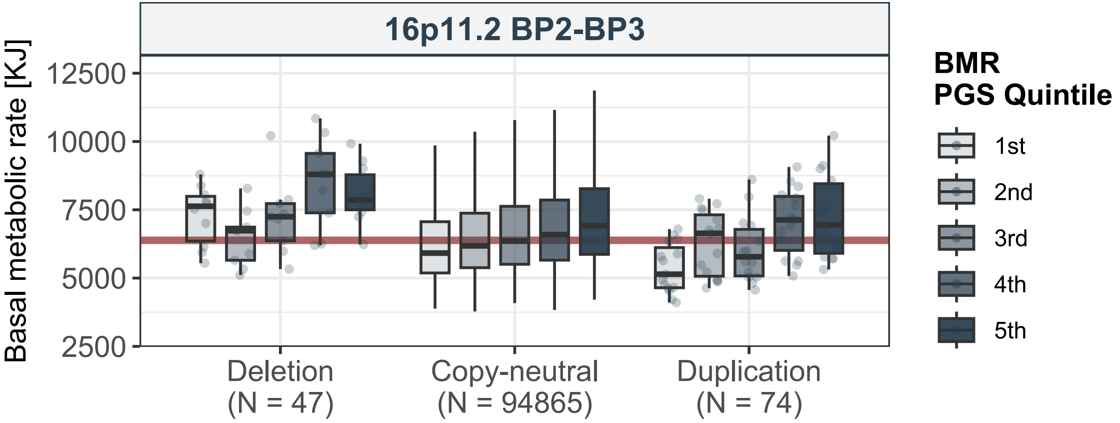
16p11.2 BP2-BP3
Basal metabolic rate [KJ]
12500
10000
7500
5000
2500
Deletion
(N = 47)
Copy-neutral
(N = 94865)
Duplication
(N = 74)
BMR
PGS Quintile
1st
2nd
3rd
4th
5th

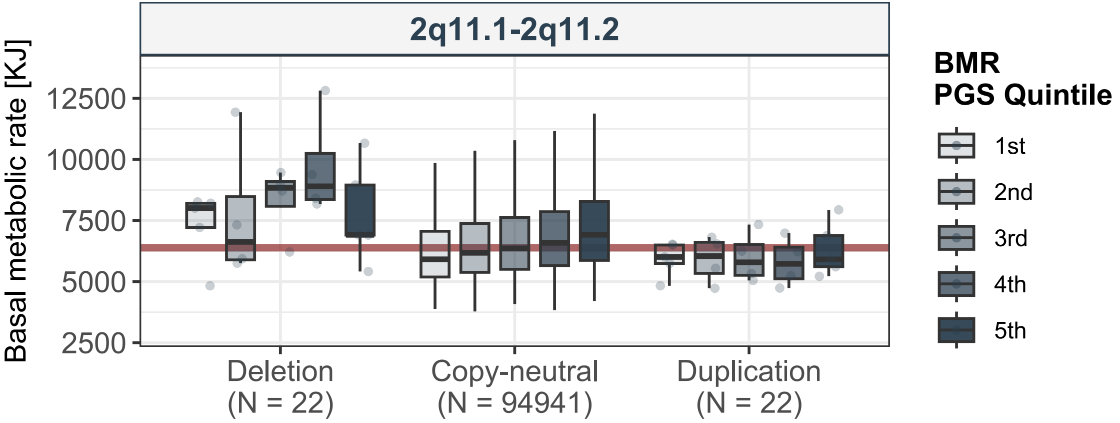
2q11.1-2q11.2
Basal metabolic rate [KJ]
12500
10000
7500
5000
2500
Deletion
(N = 22)
Copy-neutral
(N = 94941)
Duplication
(N = 22)
BMR
PGS Quintile
1st
2nd
3rd
4th
5th

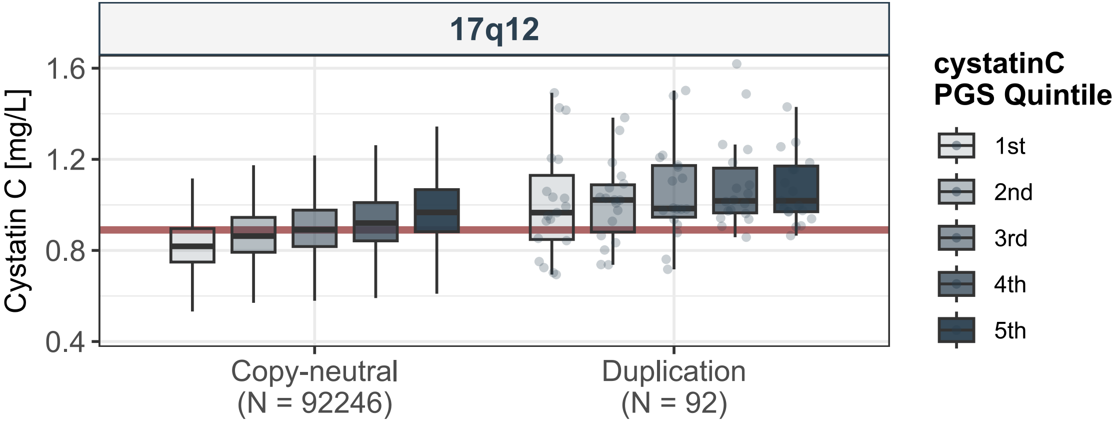

17q12
Cystatin C [mg/L]
1.6
1.2
0.8
0.4
Copy-neutral
(N = 92246)
Duplication
(N = 92)
cystatinC
PGS Quintile
1st
2nd
3rd
4th
5th

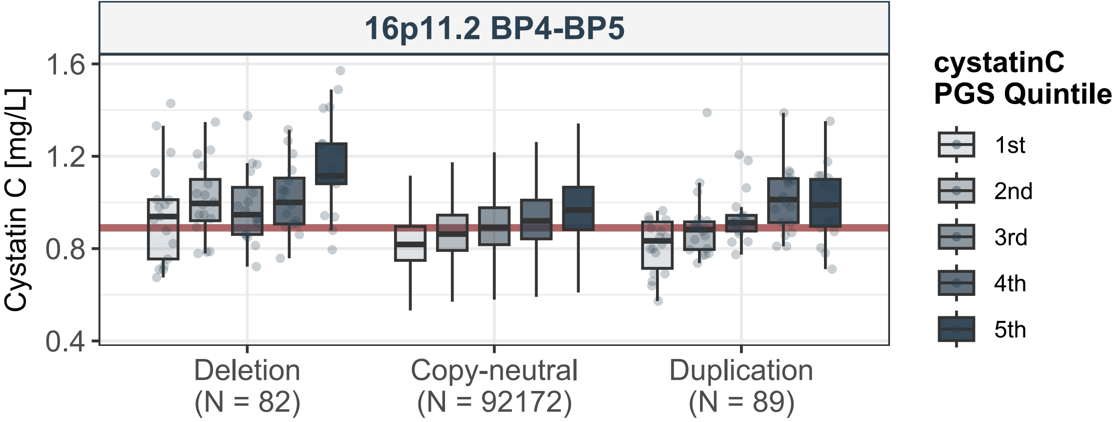
16p11.2 BP4-BP5
Cystatin C [mg/L]
1.6
1.2
0.8
0.4
Deletion
(N = 82)
Copy-neutral
(N = 92172)
Duplication
(N = 89)
cystatinC
PGS Quintile
1st
2nd
3rd
4th
5th

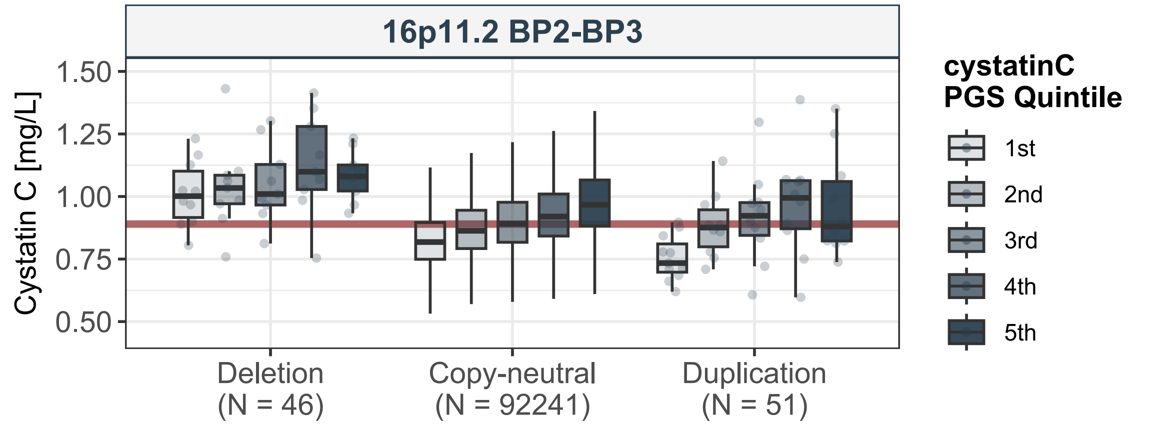
16p11.2 BP2-BP3
Cystatin C [mg/L]
1.50
1.25
1.00
0.75
0.50
Deletion
(N = 46)
Copy-neutral
(N = 92241)
Duplication
(N = 51)
cystatinC
PGS Quintile
1st
2nd
3rd
4th
5th

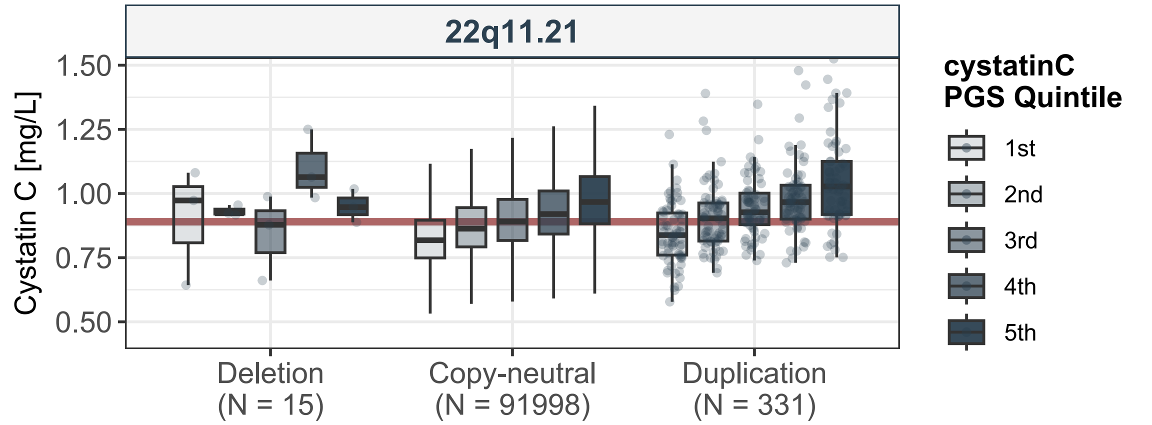
22q11.21
Cystatin C [mg/L]
1.50
1.25
1.00
0.75
0.50
Deletion
(N = 15)
Copy-neutral
(N = 91998)
Duplication
(N = 331)
cystatinC
PGS Quintile
1st
2nd
3rd
4th
5th

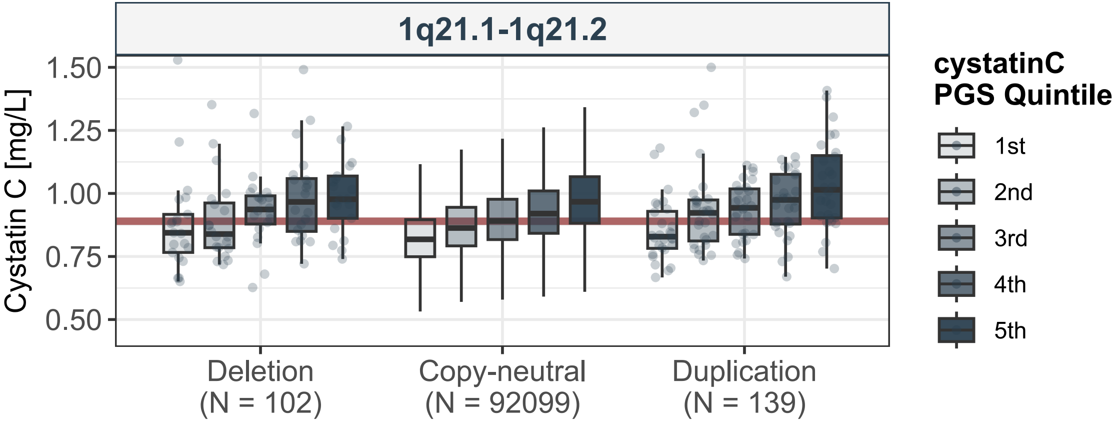
1q21.1-1q21.2
Cystatin C [mg/L]
1.50
1.25
1.00
0.75
0.50
Deletion
(N = 102)
Copy-neutral
(N = 92099)
Duplication
(N = 139)
cystatinC
PGS Quintile
1st
2nd
3rd
4th
5th

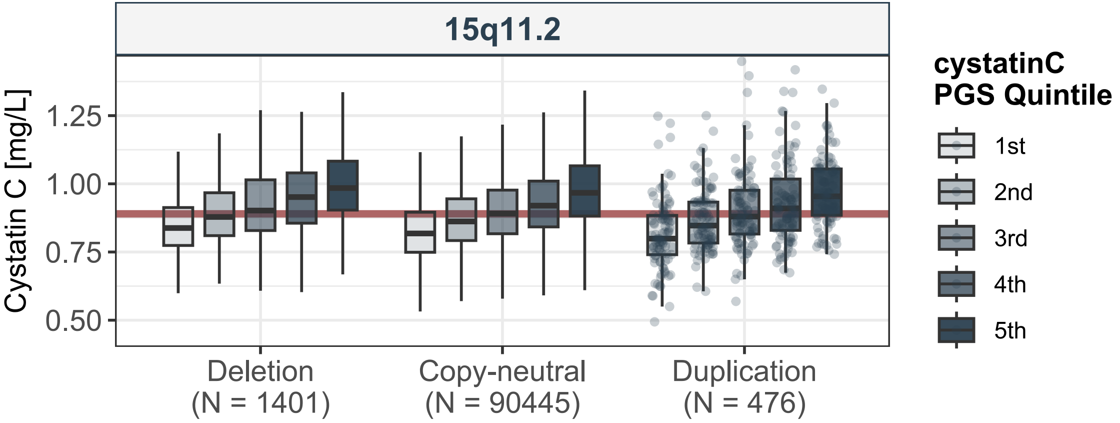

15q11.2
Cystatin C [mg/L]
1.25
1.00
0.75
0.50
Deletion
(N = 1401)
Copy-neutral
(N = 90445)
Duplication
(N = 476)
cystatinC
PGS Quintile
1st
2nd
3rd
4th
5th

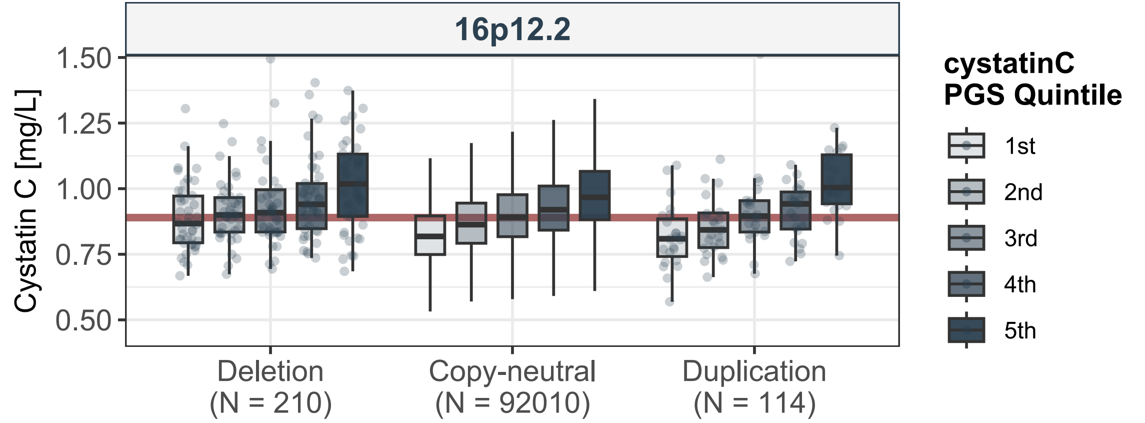

16p12.2
Cystatin C [mg/L]
1.50
1.25
1.00
0.75
0.50
Deletion
(N = 210)
Copy-neutral
(N = 92010)
Duplication
(N = 114)
cystatinC
PGS Quintile
1st
2nd
3rd
4th
5th

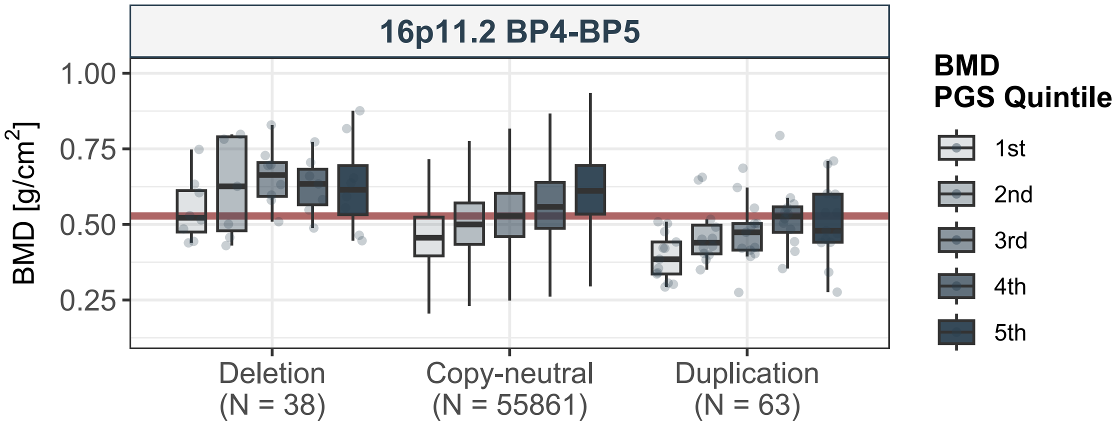
16p11.2 BP4-BP5
BMD [g/cm2]
1.00
0.75
0.50
0.25
Deletion
(N = 38)
Copy-neutral
(N = 55861)
Duplication
(N = 63)
BMD
PGS Quintile
1st
2nd
3rd
4th
5th

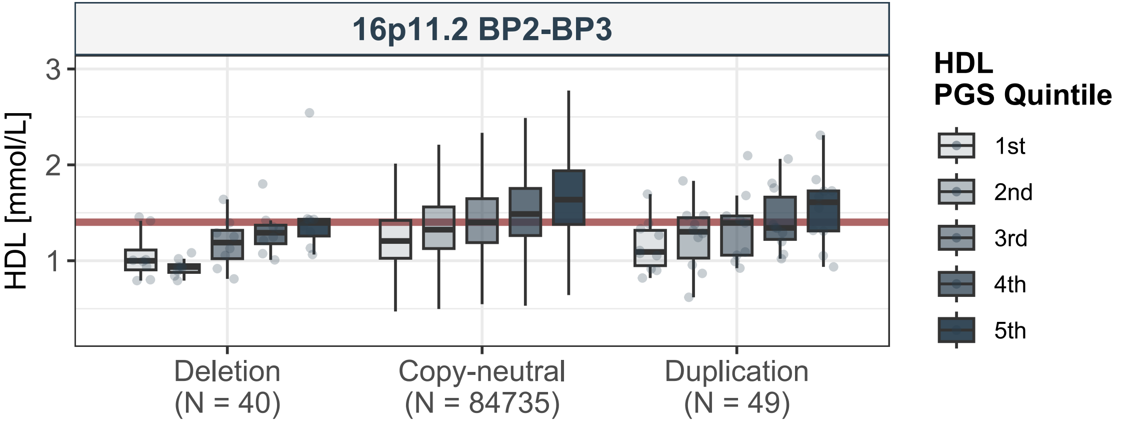
16p11.2 BP2-BP3
HDL [mmol/L]
3
2
1
Deletion
(N = 40)
Copy-neutral
(N = 84735)
Duplication
(N = 49)
HDL
PGS Quintile
1st
2nd
3rd
4th
5th

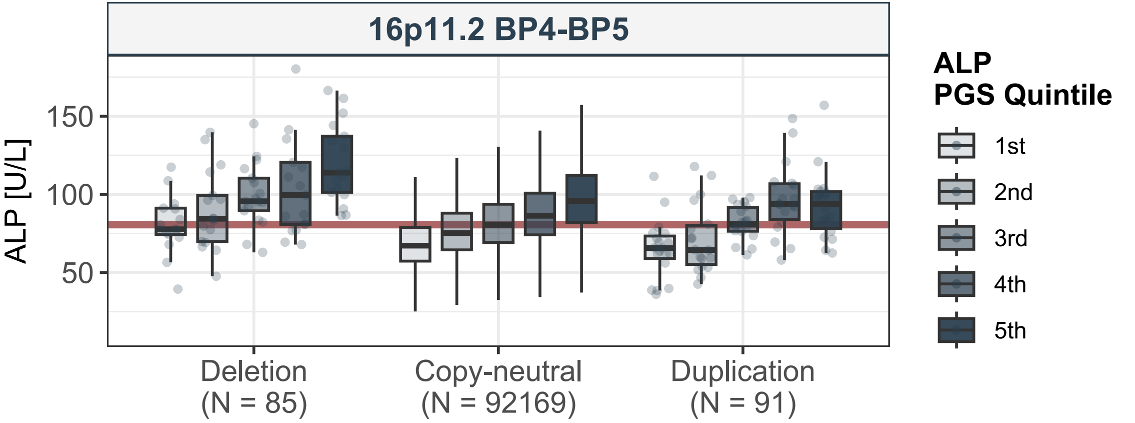
16p11.2 BP4-BP5
ALP [U/L]
150
100
50
Deletion
(N = 85)
Copy-neutral
(N = 92169)
Duplication
(N = 91)
ALP
PGS Quintile
1st
2nd
3rd
4th
5th

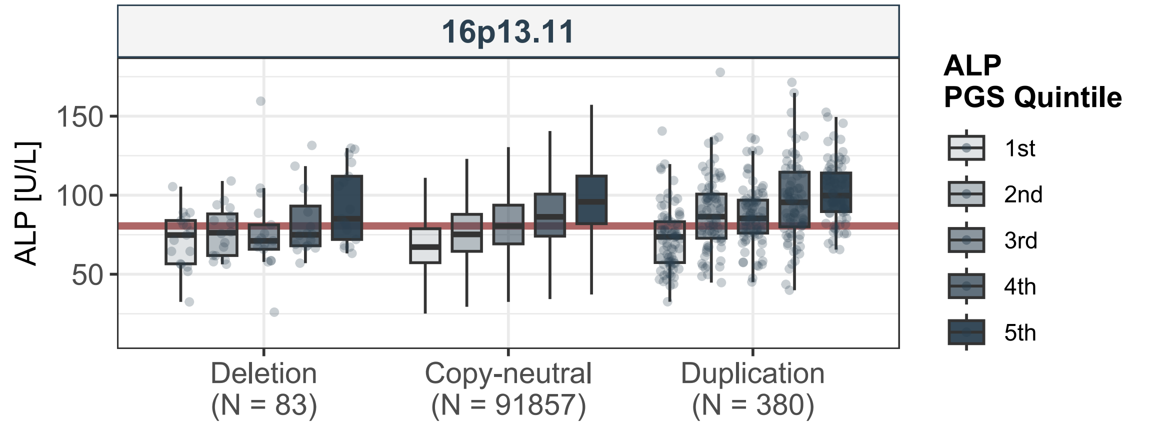

16p13.11
ALP [U/L]
150
100
50
Deletion
(N = 83)
Copy-neutral
(N = 91857)
Duplication
(N = 380)
ALP
PGS Quintile
1st
2nd
3rd
4th
5th

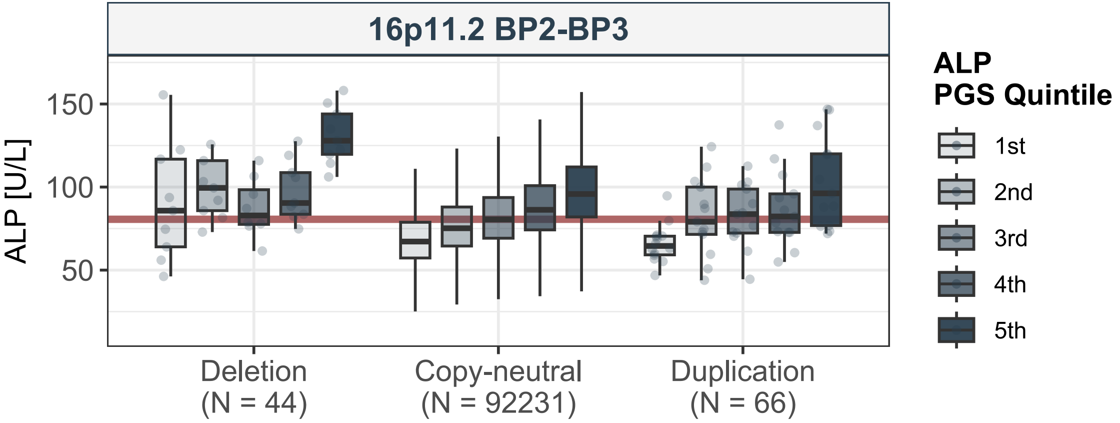

16p11.2 BP2-BP3
ALP [U/L]
150
100
50
Deletion
(N = 44)
Copy-neutral
(N = 92231)
Duplication
(N = 66)
ALP
PGS Quintile
1st
2nd
3rd
4th
5th

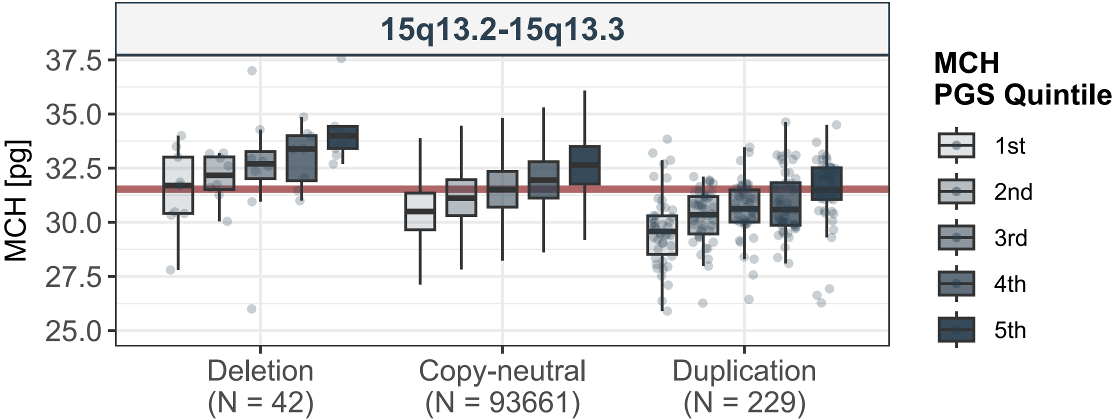
15q13.2-15q13.3
MCH [pg]
37.5
35.0
32.5
30.0
27.5
25.0
Deletion
(N = 42)
Copy-neutral
(N = 93661)
Duplication
(N = 229)
MCH
PGS Quintile
1st
2nd
3rd
4th
5th

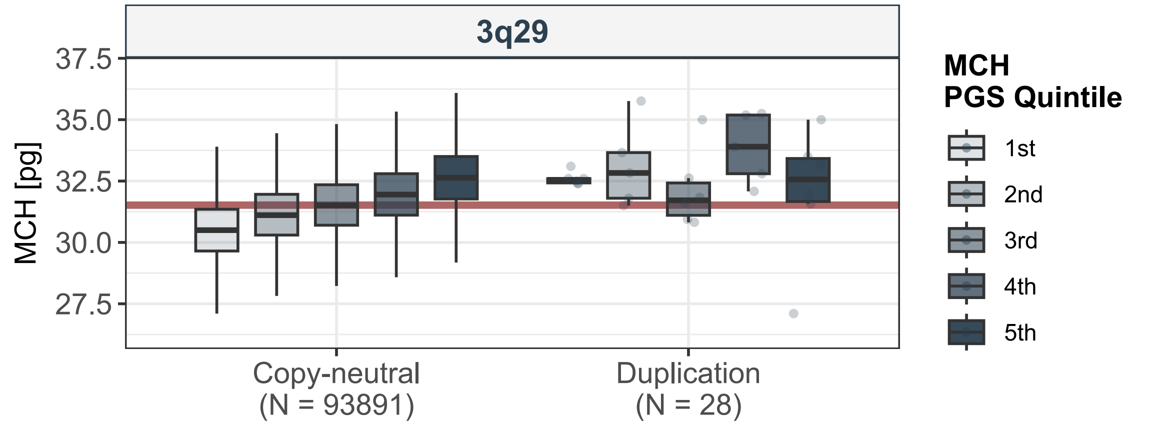
3q29
MCH [pg]
37.5
35.0
32.5
30.0
27.5
Copy-neutral
(N = 93891)
Duplication
(N = 28)
MCH
PGS Quintile
1st
2nd
3rd
4th
5th

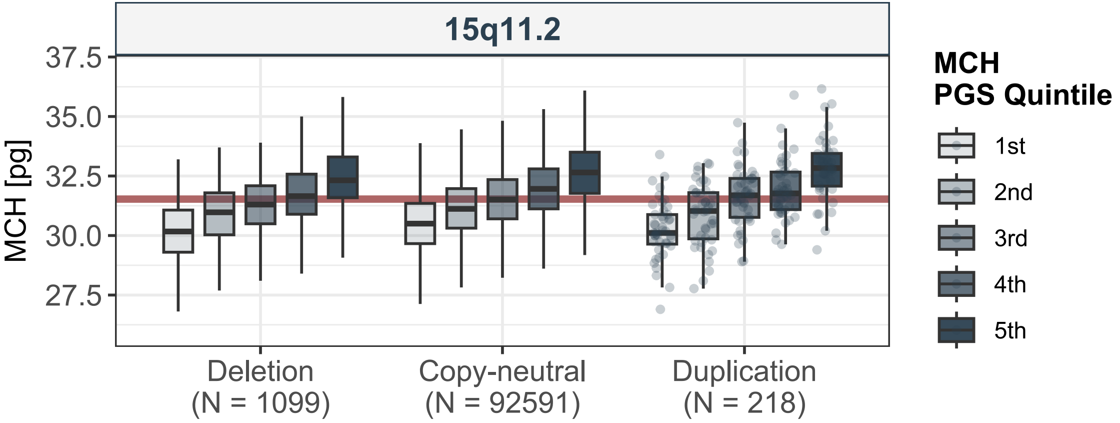
15q11.2
MCH [pg]
37.5
35.0
32.5
30.0
27.5
Deletion
(N = 1099)
Copy-neutral
(N = 92591)
Duplication
(N = 218)
MCH
PGS Quintile
1st
2nd
3rd
4th
5th

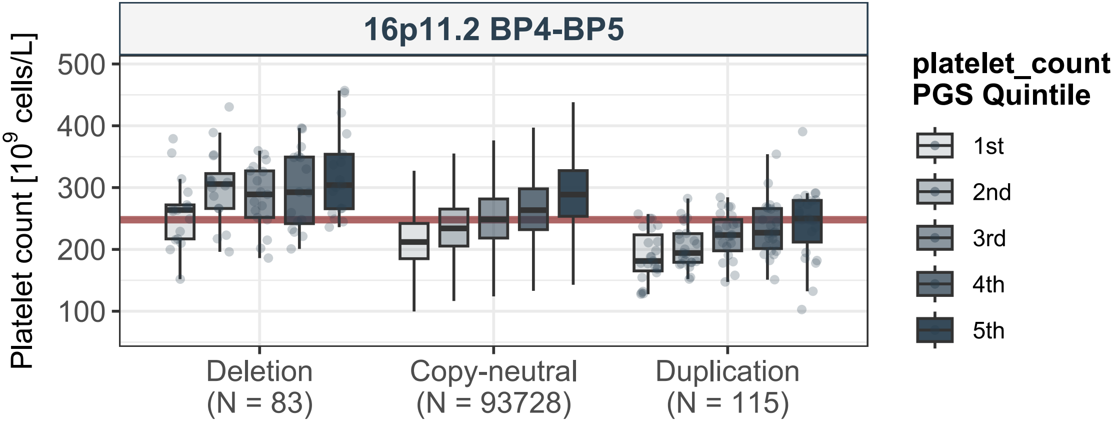
16p11.2 BP4-BP5
Platelet count [$10^9$ cells/L]
500
400
300
200
100
Deletion
(N = 83)
Copy-neutral
(N = 93728)
Duplication
(N = 115)
platelet_count
PGS Quintile
1st
2nd
3rd
4th
5th

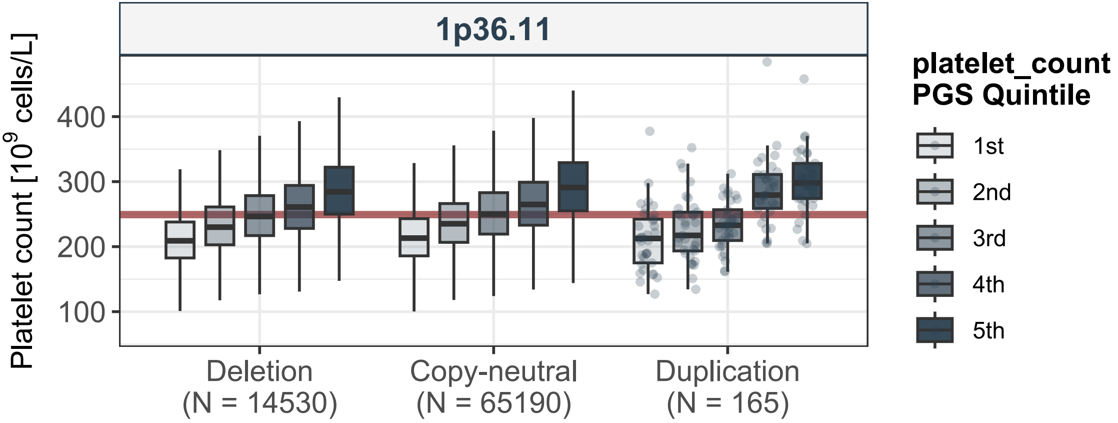

1p36.11
Platelet count [10^9 cells/L]
400
300
200
100
Deletion
(N = 14530)
Copy-neutral
(N = 65190)
Duplication
(N = 165)
platelet_count
PGS Quintile
1st
2nd
3rd
4th
5th

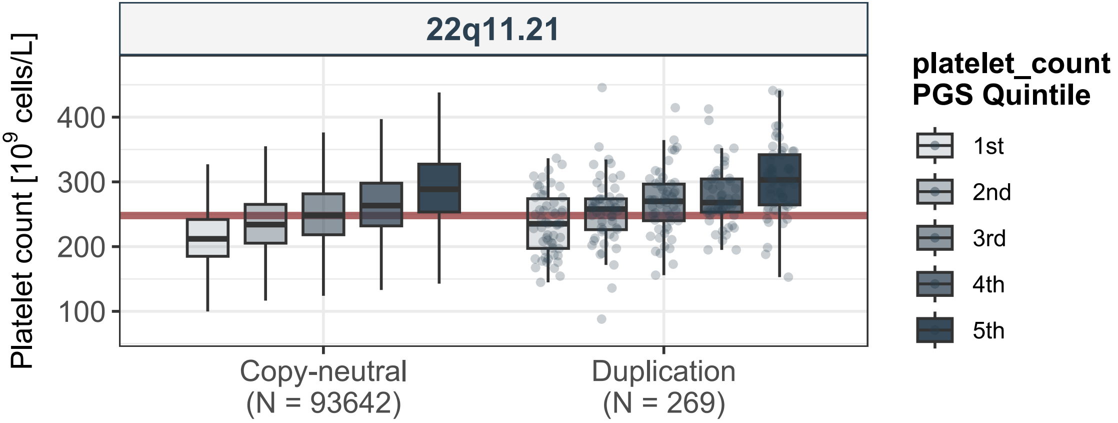

22q11.21
Platelet count [10^9 cells/L]
400
300
200
100
Copy-neutral
(N = 93642)
Duplication
(N = 269)
platelet_count
PGS Quintile
1st
2nd
3rd
4th
5th

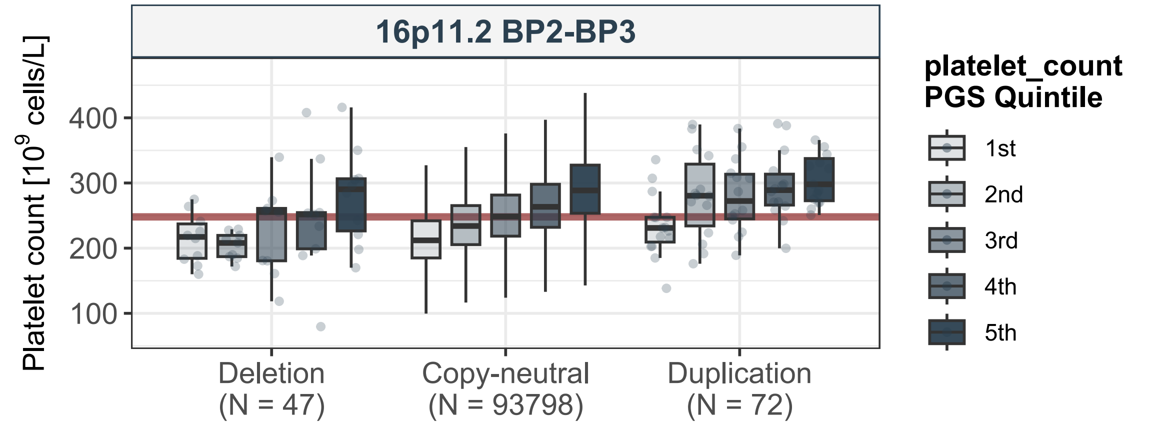

16p11.2 BP2-BP3
Platelet count [$10^9$ cells/L]
400
300
200
100
Deletion
(N = 47)
Copy-neutral
(N = 93798)
Duplication
(N = 72)
platelet_count
PGS Quintile
1st
2nd
3rd
4th
5th

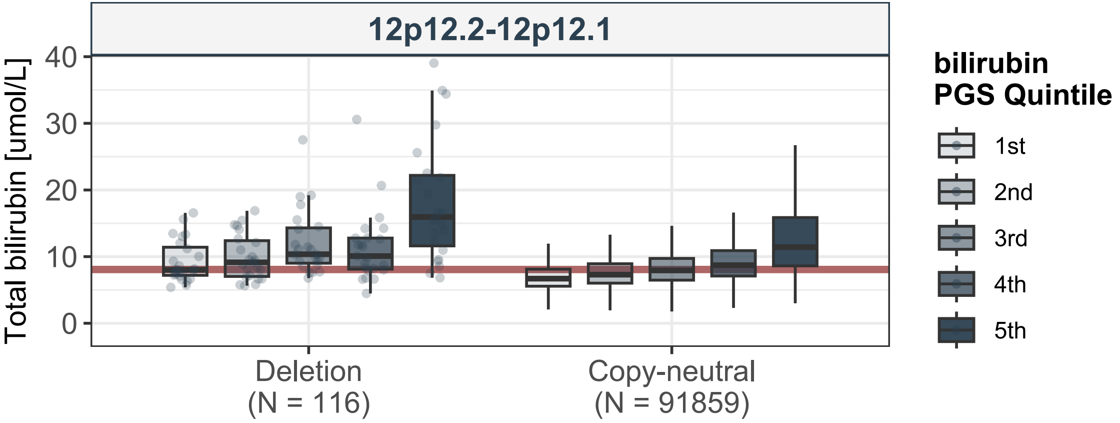

12p12.2-12p12.1
Total bilirubin [umol/L]
0
10
20
30
40
Deletion
(N = 116)
Copy-neutral
(N = 91859)
bilirubin
PGS Quintile
1st
2nd
3rd
4th
5th

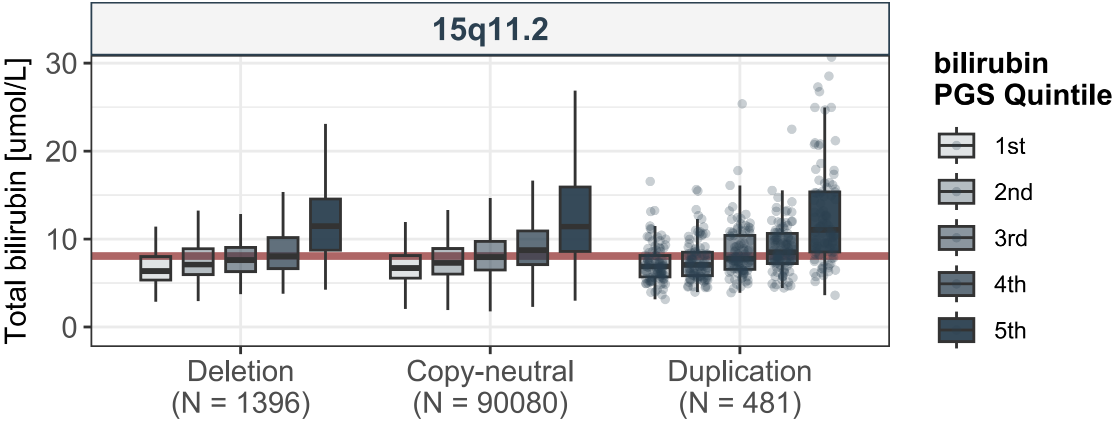
15q11.2
Total bilirubin [umol/L]
0
10
20
30
Deletion
(N = 1396)
Copy-neutral
(N = 90080)
Duplication
(N = 481)
bilirubin
PGS Quintile
1st
2nd
3rd
4th
5th

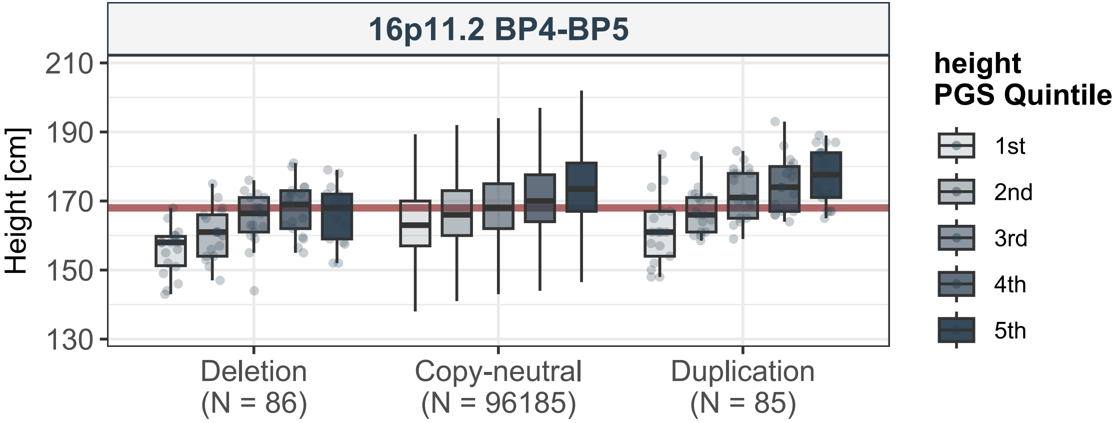

16p11.2 BP4-BP5
Height [cm]
210
190
170
150
130
Deletion
(N = 86)
Copy-neutral
(N = 96185)
Duplication
(N = 85)
height
PGS Quintile
1st
2nd
3rd
4th
5th

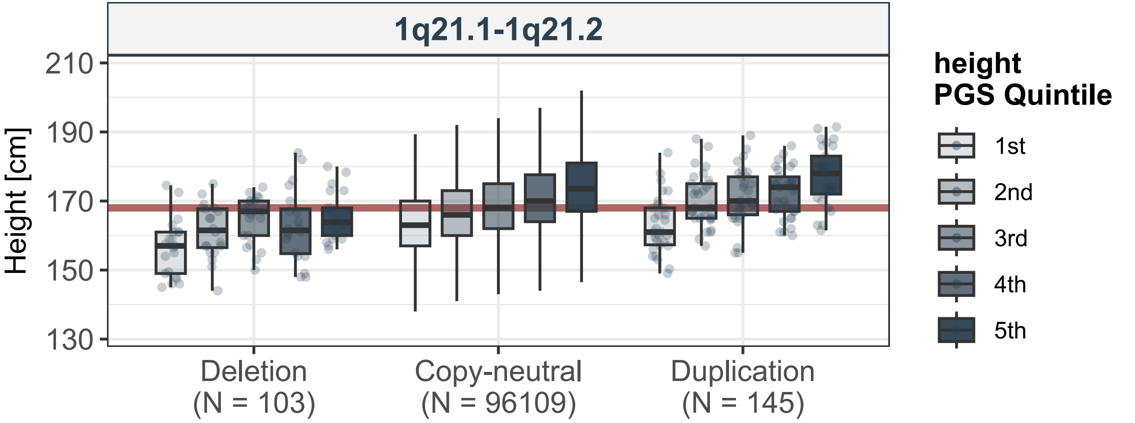

1q21.1-1q21.2
Height [cm]
210
190
170
150
130
Deletion
(N = 103)
Copy-neutral
(N = 96109)
Duplication
(N = 145)
height
PGS Quintile
1st
2nd
3rd
4th
5th

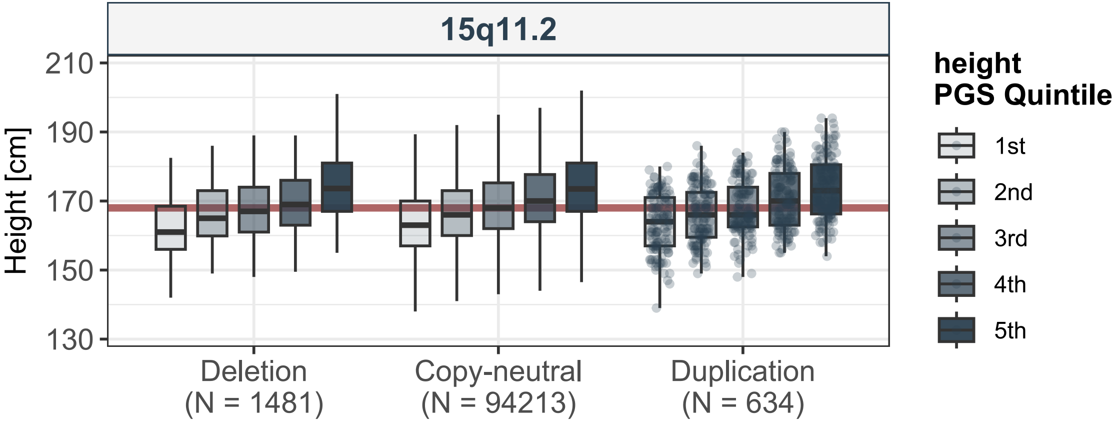
15q11.2
Height [cm]
210
190
170
150
130
Deletion
(N = 1481)
Copy-neutral
(N = 94213)
Duplication
(N = 634)
height
PGS Quintile
1st
2nd
3rd
4th
5th

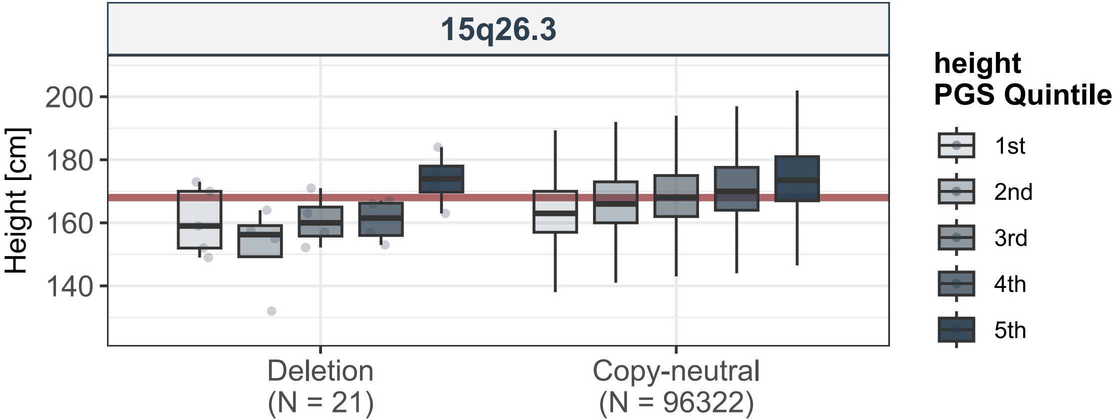
15q26.3
Height [cm]
200
180
160
140
Deletion
(N = 21)
Copy-neutral
(N = 96322)
height
PGS Quintile
1st
2nd
3rd
4th
5th

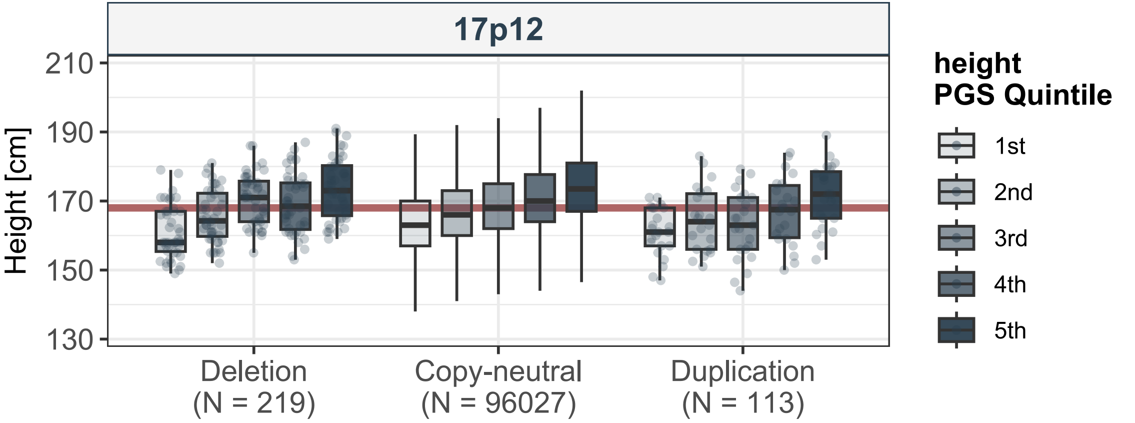
17p12
Height [cm]
210
190
170
150
130
Deletion
(N = 219)
Copy-neutral
(N = 96027)
Duplication
(N = 113)
height
PGS Quintile
1st
2nd
3rd
4th
5th

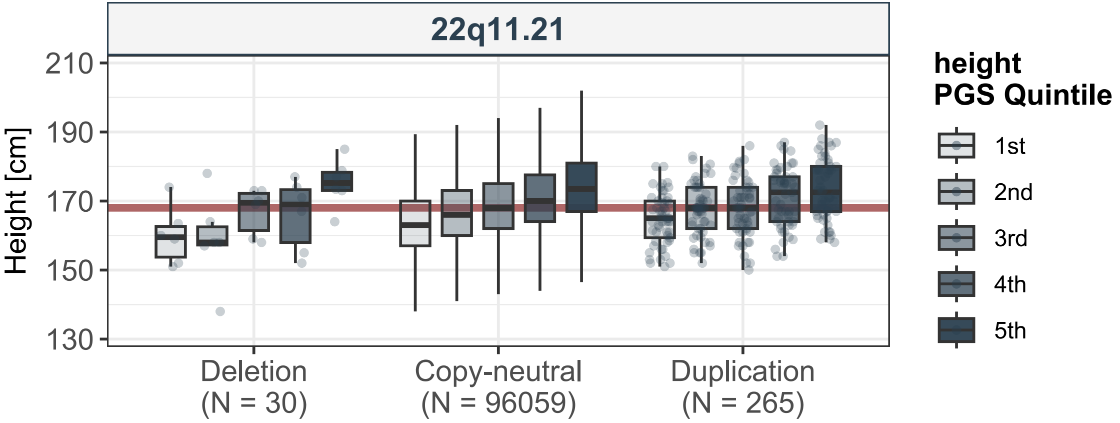
22q11.21
Height [cm]
210
190
170
150
130
Deletion
(N = 30)
Copy-neutral
(N = 96059)
Duplication
(N = 265)
height
PGS Quintile
1st
2nd
3rd
4th
5th

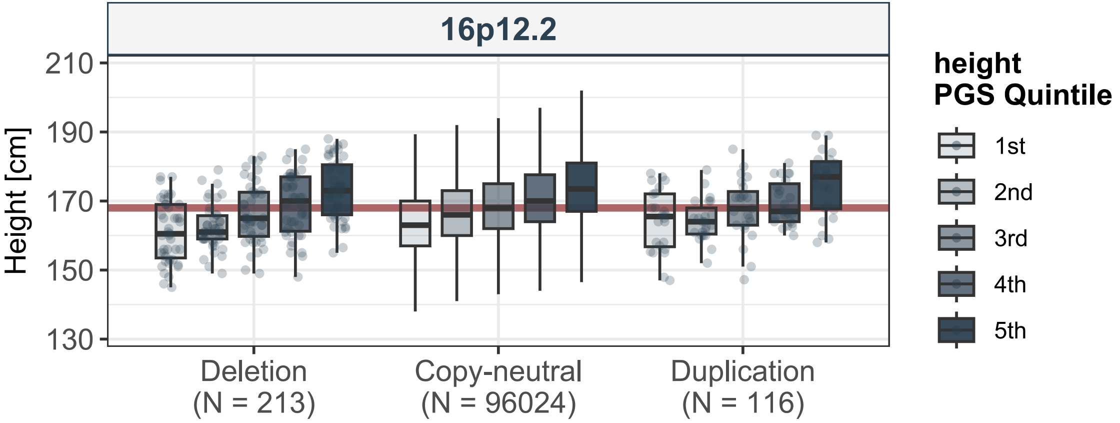
16p12.2
Height [cm]
210
190
170
150
130
Deletion
(N = 213)
Copy-neutral
(N = 96024)
Duplication
(N = 116)
height
PGS Quintile
1st
2nd
3rd
4th
5th
