## Extended Data 2 for "Compounding of rare pathogenic copy-number variants and polygenic background is consistent with assortative mating"

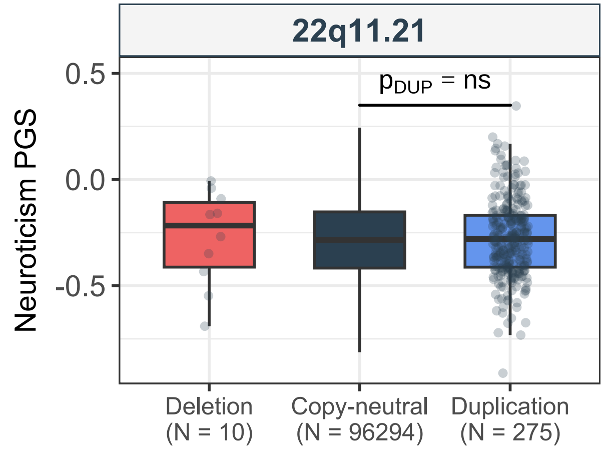
22q11.21
$p_{DUP}$ = ns
Neuroticism PGS
0.5
0.0
-0.5
Deletion
(N = 10)
Copy-neutral
(N = 96294)
Duplication
(N = 275)

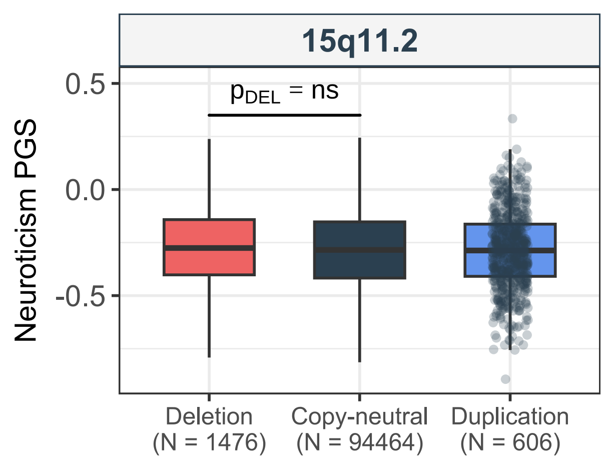
15q11.2
pDEL = ns
Neuroticism PGS
0.5
0.0
-0.5
Deletion
(N = 1476)
Copy-neutral
(N = 94464)
Duplication
(N = 606)

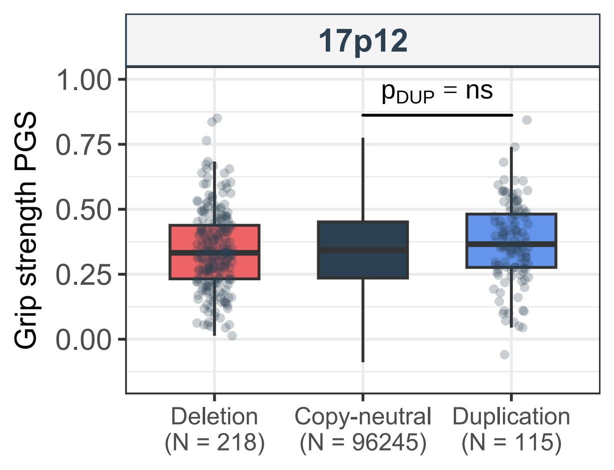
17p12
$p_{DUP}$ = ns
Grip strength PGS
1.00
0.75
0.50
0.25
0.00
Deletion
(N = 218)
Copy-neutral
(N = 96245)
Duplication
(N = 115)

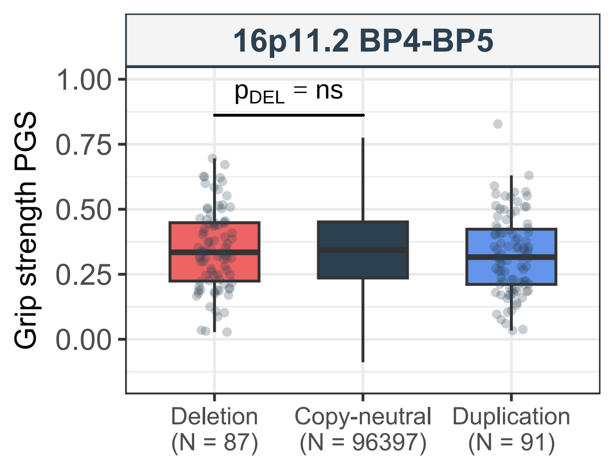
16p11.2 BP4-BP5
$p_{DEL}$ = ns
Grip strength PGS
1.00
0.75
0.50
0.25
0.00
Deletion
(N = 87)
Copy-neutral
(N = 96397)
Duplication
(N = 91)

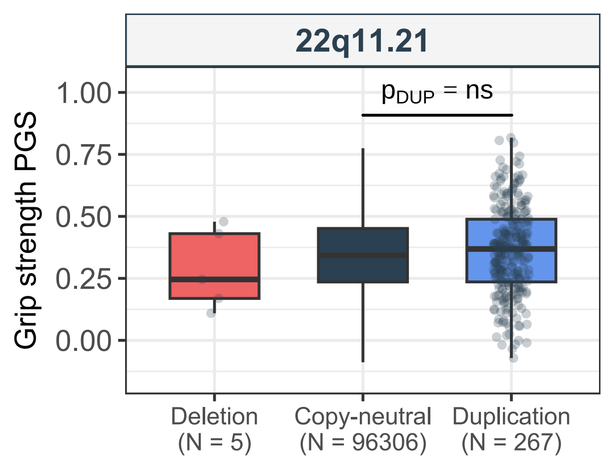

22q11.21
$p_{DUP}$ = ns
Grip strength PGS
1.00
0.75
0.50
0.25
0.00
Deletion (N = 5)
Copy-neutral (N = 96306)
Duplication (N = 267)

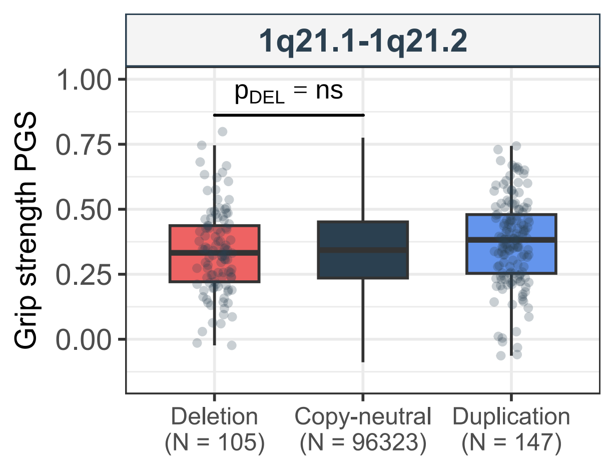
1q21.1-1q21.2
$p_{DEL}$ = ns
Grip strength PGS
1.00
0.75
0.50
0.25
0.00
Deletion
(N = 105)
Copy-neutral
(N = 96323)
Duplication
(N = 147)

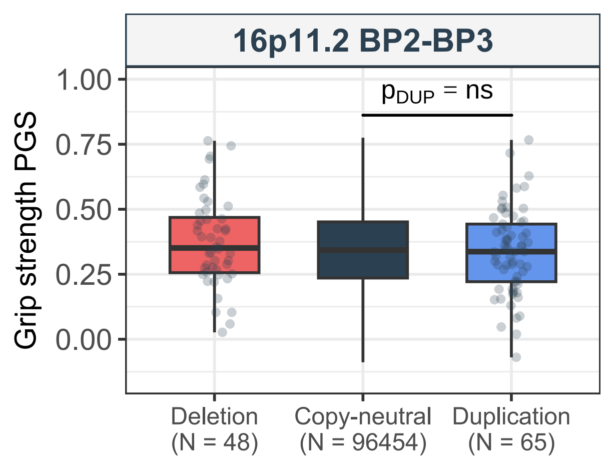
16p11.2 BP2-BP3
Grip strength PGS
1.00
0.75
0.50
0.25
0.00
$p_{DUP}$ = ns
Deletion (N = 48)
Copy-neutral (N = 96454)
Duplication (N = 65)

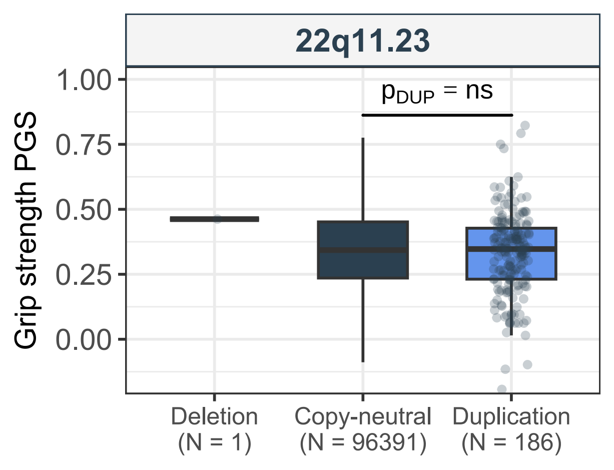

22q11.23
Grip strength PGS
1.00
0.75
0.50
0.25
0.00
$p_{DUP}$ = ns
Deletion
(N = 1)
Copy-neutral
(N = 96391)
Duplication
(N = 186)

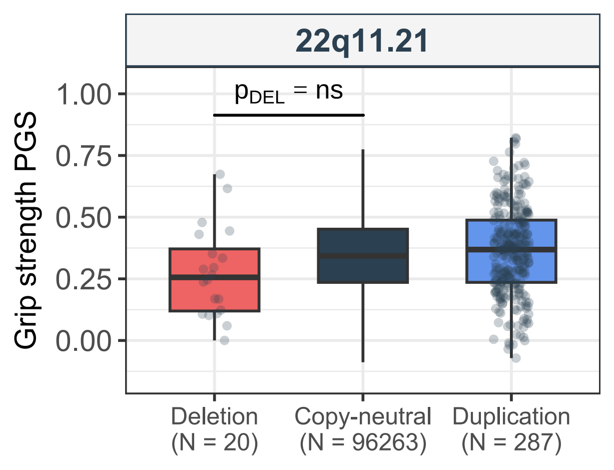

22q11.21
$p_{DEL}$ = ns
Grip strength PGS
1.00
0.75
0.50
0.25
0.00
Deletion
(N = 20)
Copy-neutral
(N = 96263)
Duplication
(N = 287)

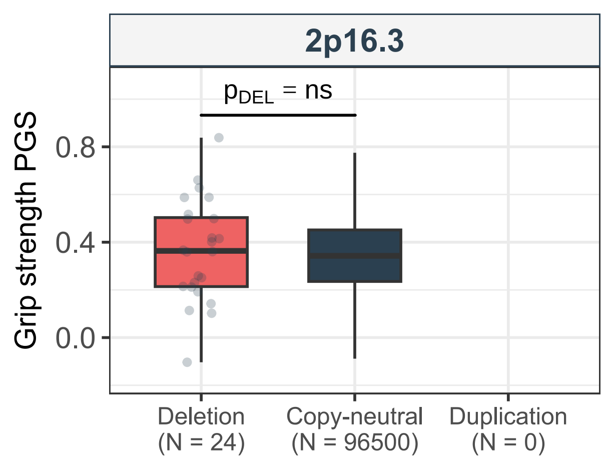
2p16.3
Grip strength PGS
p_DEL = ns
0.8
0.4
0.0
Deletion
(N = 24)
Copy-neutral
(N = 96500)
Duplication
(N = 0)

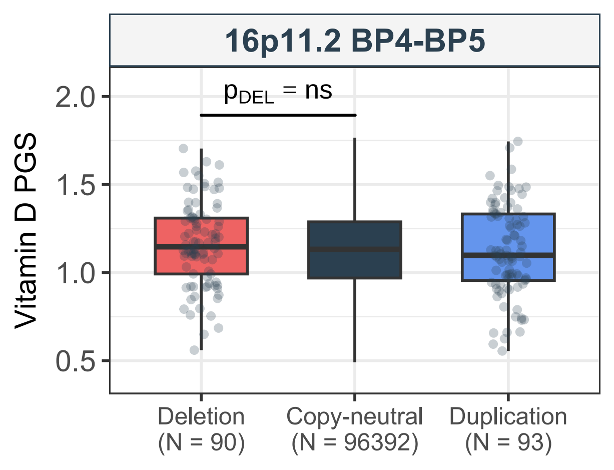

16p11.2 BP4-BP5
$p_{DEL}$ = ns
Vitamin D PGS
2.0
1.5
1.0
0.5
Deletion
(N = 90)
Copy-neutral
(N = 96392)
Duplication
(N = 93)

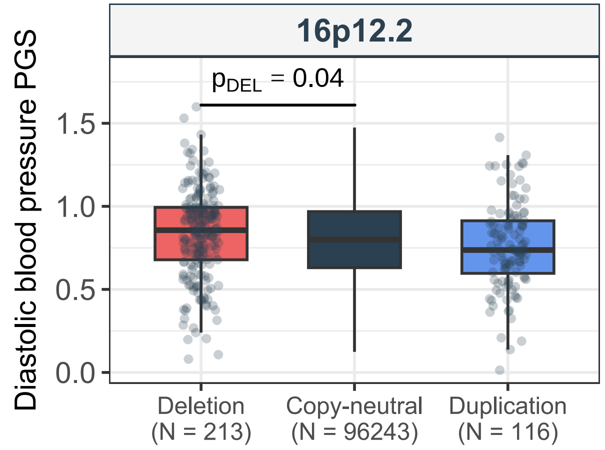

16p12.2
$p_{DEL}$ = 0.04
Diastolic blood pressure PGS
1.5
1.0
0.5
0.0
Deletion
(N = 213)
Copy-neutral
(N = 96243)
Duplication
(N = 116)

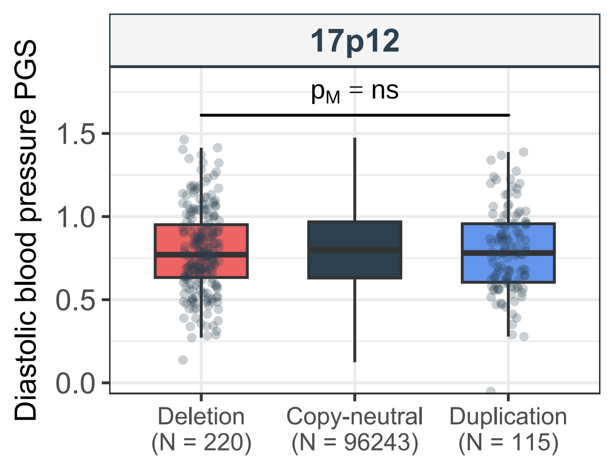
17p12
$p_M$ = ns
Diastolic blood pressure PGS
1.5
1.0
0.5
0.0
Deletion
(N = 220)
Copy-neutral
(N = 96243)
Duplication
(N = 115)

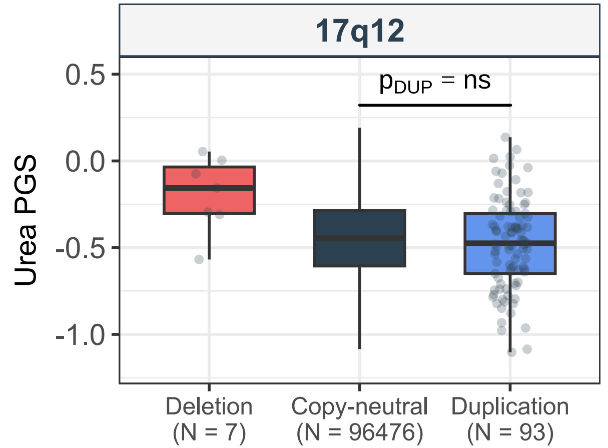
17q12
$p_{DUP}$ = ns
0.5
0.0
-0.5
-1.0
Urea PGS
Deletion (N = 7)
Copy-neutral (N = 96476)
Duplication (N = 93)

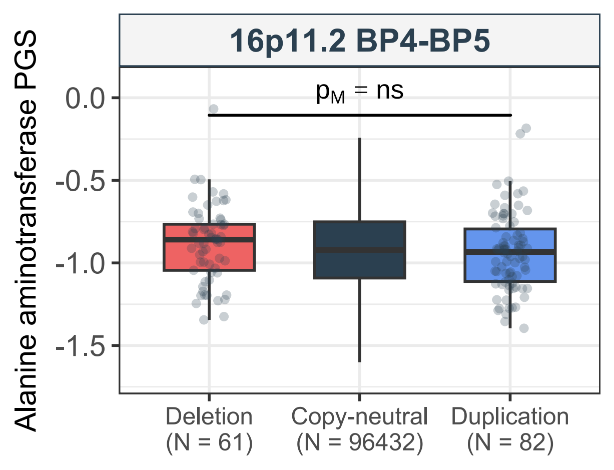
16p11.2 BP4-BP5
Alanine aminotransferase PGS
$p_M$ = ns
0.0
-0.5
-1.0
-1.5
Deletion
(N = 61)
Copy-neutral
(N = 96432)
Duplication
(N = 82)

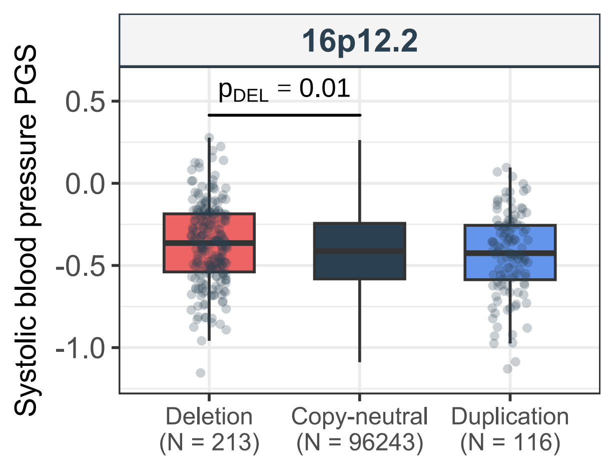

16p12.2
$p_{DEL}$ = 0.01
Systolic blood pressure PGS
0.5
0.0
-0.5
-1.0
Deletion
(N = 213)
Copy-neutral
(N = 96243)
Duplication
(N = 116)

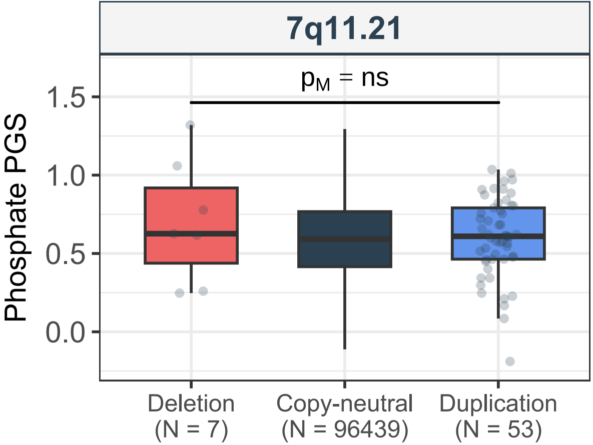
7q11.21
$p_M$ = ns
Phosphate PGS
1.5
1.0
0.5
0.0
Deletion
(N = 7)
Copy-neutral
(N = 96439)
Duplication
(N = 53)

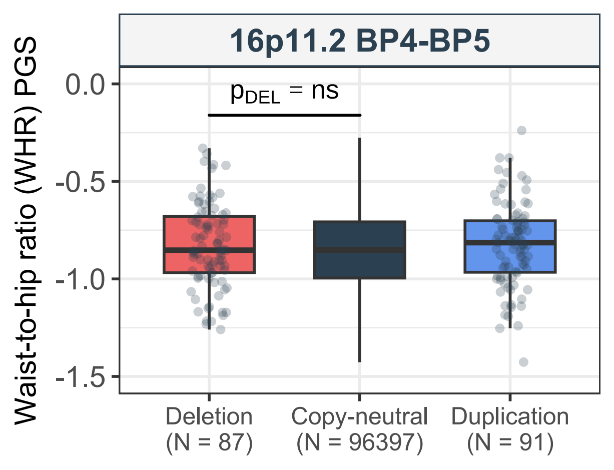
16p11.2 BP4-BP5
Waist-to-hip ratio (WHR) PGS
$p_{DEL}$ = ns
0.0
-0.5
-1.0
-1.5
Deletion
(N = 87)
Copy-neutral
(N = 96397)
Duplication
(N = 91)

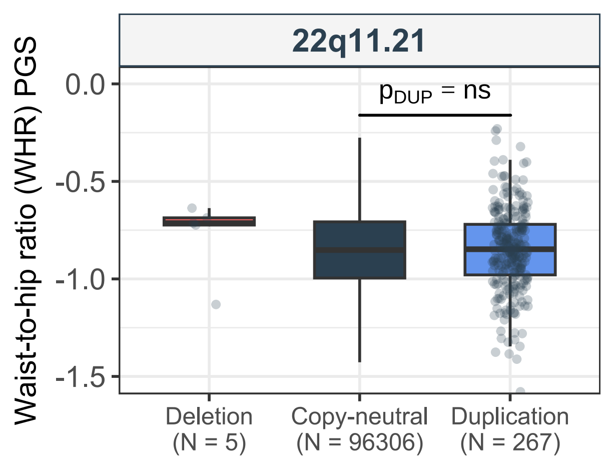
22q11.21
Waist-to-hip ratio (WHR) PGS
0.0
-0.5
-1.0
-1.5
$p_{DUP}$ = ns
Deletion
(N = 5)
Copy-neutral
(N = 96306)
Duplication
(N = 267)

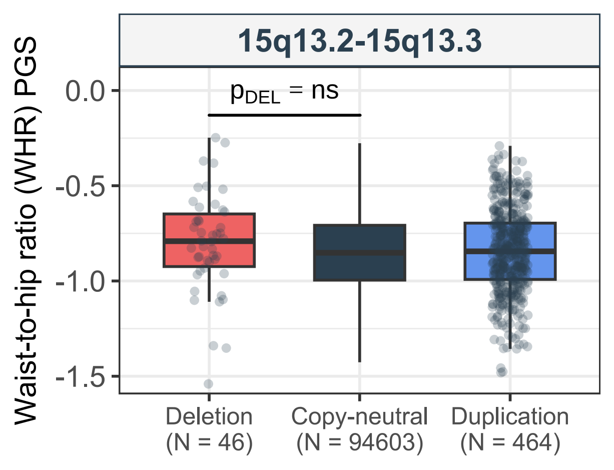
15q13.2-15q13.3
Waist-to-hip ratio (WHR) PGS
0.0
-0.5
-1.0
-1.5
p_DEL = ns
Deletion
(N = 46)
Copy-neutral
(N = 94603)
Duplication
(N = 464)

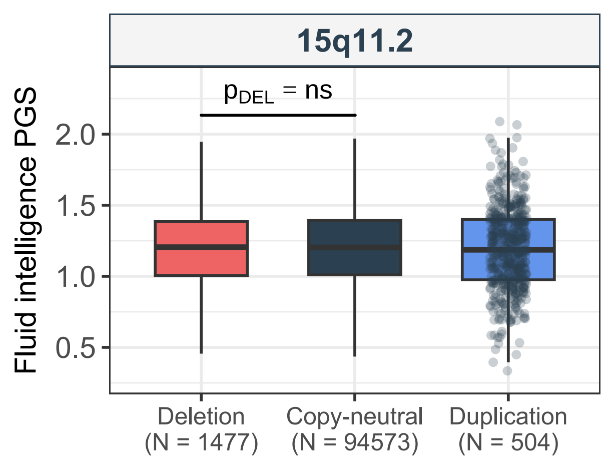

15q11.2
$p_{DEL}$ = ns
Fluid intelligence PGS
2.0
1.5
1.0
0.5
Deletion (N = 1477)
Copy-neutral (N = 94573)
Duplication (N = 504)

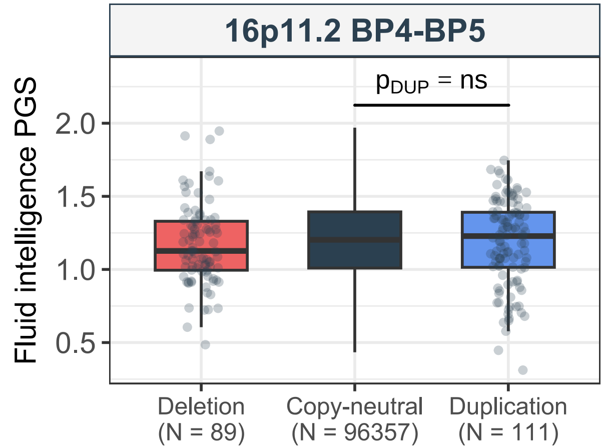
16p11.2 BP4-BP5
$p_{DUP}$ = ns
Fluid intelligence PGS
2.0
1.5
1.0
0.5
Deletion
(N = 89)
Copy-neutral
(N = 96357)
Duplication
(N = 111)

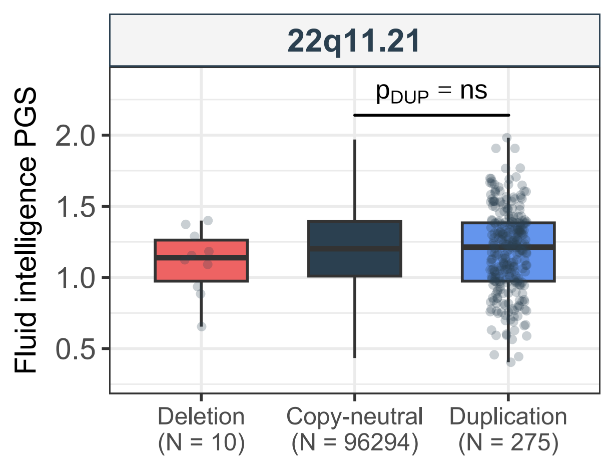
22q11.21
$p_{DUP}$ = ns
Fluid intelligence PGS
2.0
1.5
1.0
0.5
Deletion
(N = 10)
Copy-neutral
(N = 96294)
Duplication
(N = 275)

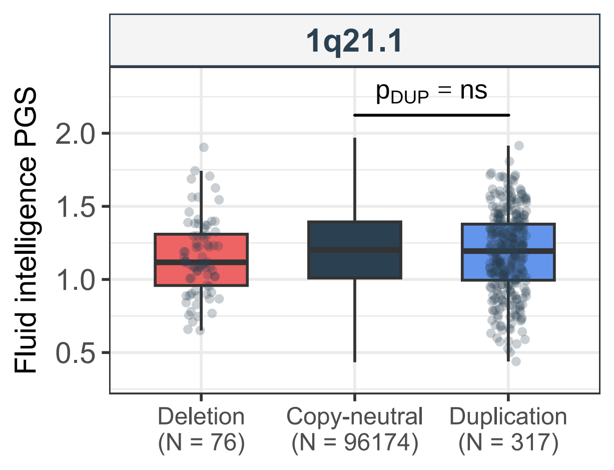
1q21.1
$p_{DUP}$ = ns
Fluid intelligence PGS
2.0
1.5
1.0
0.5
Deletion
(N = 76)
Copy-neutral
(N = 96174)
Duplication
(N = 317)

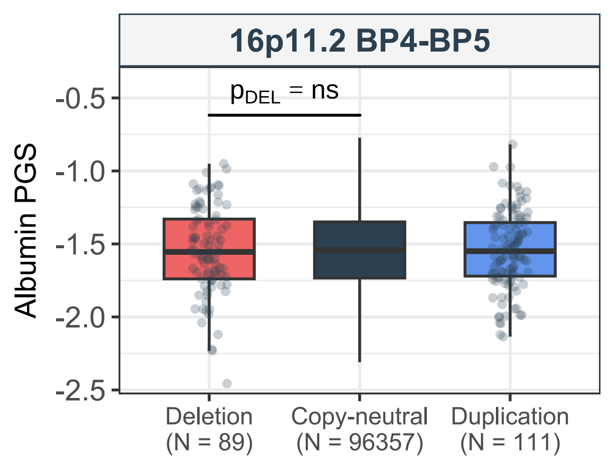

16p11.2 BP4-BP5
p_DEL = ns
Albumin PGS
-0.5
-1.0
-1.5
-2.0
-2.5
Deletion
(N = 89)
Copy-neutral
(N = 96357)
Duplication
(N = 111)

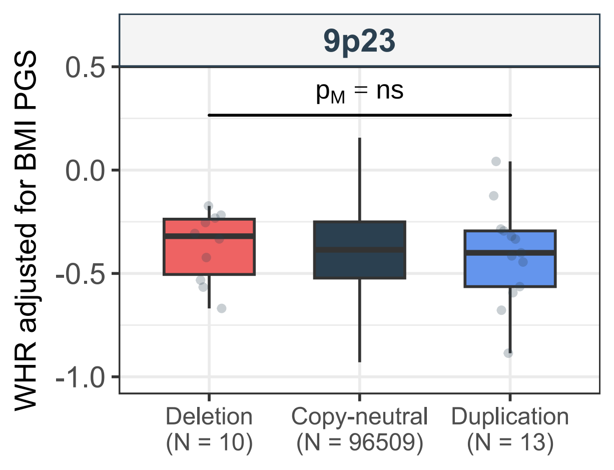

9p23
$p_M$ = ns
WHR adjusted for BMI PGS
0.5
0.0
-0.5
-1.0
Deletion
(N = 10)
Copy-neutral
(N = 96509)
Duplication
(N = 13)

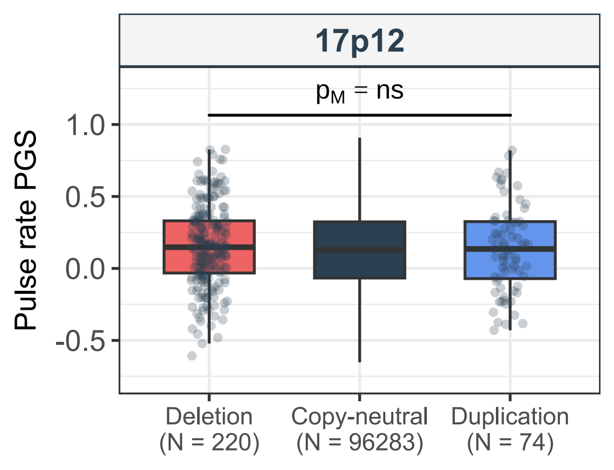
17p12
$p_M$ = ns
1.0
0.5
0.0
-0.5
Pulse rate PGS
Deletion
(N = 220)
Copy-neutral
(N = 96283)
Duplication
(N = 74)

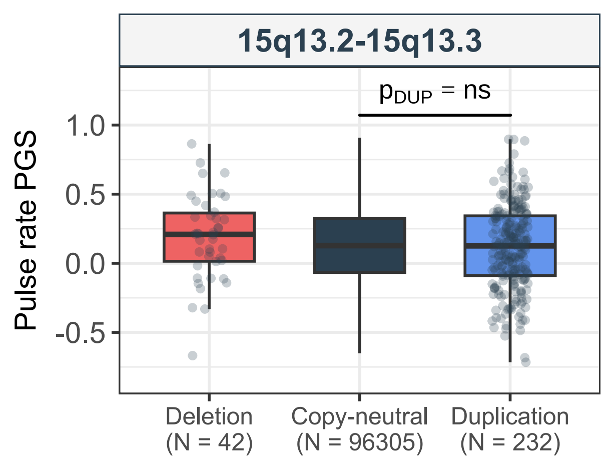

15q13.2-15q13.3
$p_{DUP}$ = ns
Pulse rate PGS
1.0
0.5
0.0
-0.5
Deletion (N = 42)
Copy-neutral (N = 96305)
Duplication (N = 232)

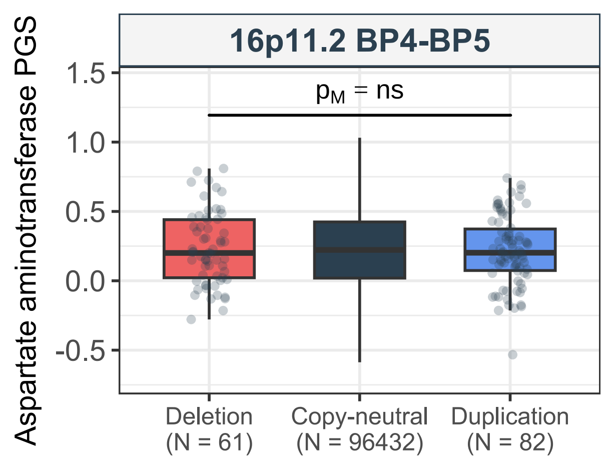

16p11.2 BP4-BP5
Aspartate aminotransferase PGS
$p_M$ = ns
1.5
1.0
0.5
0.0
-0.5
Deletion
(N = 61)
Copy-neutral
(N = 96432)
Duplication
(N = 82)

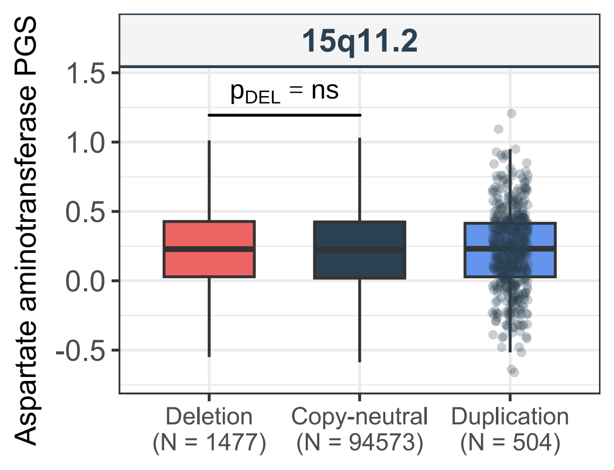

15q11.2
Aspartate aminotransferase PGS
p_DEL = ns
1.5
1.0
0.5
0.0
-0.5
Deletion
(N = 1477)
Copy-neutral
(N = 94573)
Duplication
(N = 504)

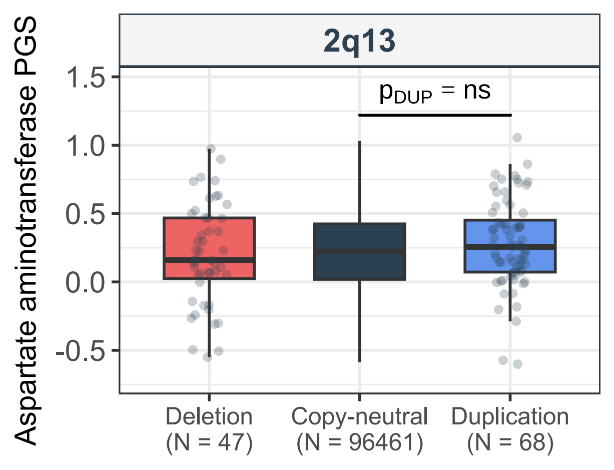
2q13
Aspartate aminotransferase PGS
$p_{DUP}$ = ns
1.5
1.0
0.5
0.0
-0.5
Deletion
(N = 47)
Copy-neutral
(N = 96461)
Duplication
(N = 68)

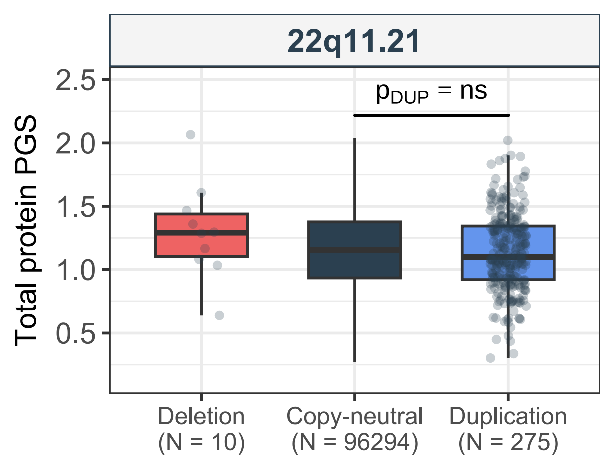

22q11.21
$p_{DUP}$ = ns
Total protein PGS
2.5
2.0
1.5
1.0
0.5
Deletion
(N = 10)
Copy-neutral
(N = 96294)
Duplication
(N = 275)

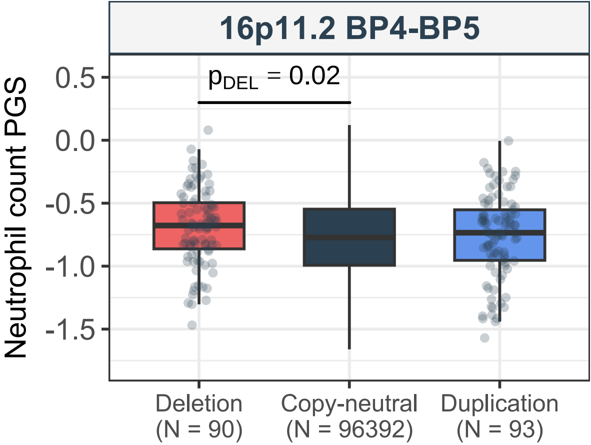
16p11.2 BP4-BP5
$p_{DEL}$ = 0.02
Neutrophil count PGS
0.5
0.0
-0.5
-1.0
-1.5
Deletion
(N = 90)
Copy-neutral
(N = 96392)
Duplication
(N = 93)

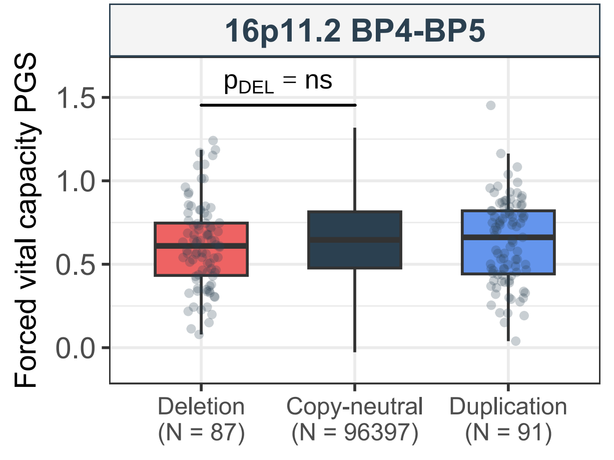

16p11.2 BP4-BP5
$p_{DEL}$ = ns
Forced vital capacity PGS
1.5
1.0
0.5
0.0
Deletion
(N = 87)
Copy-neutral
(N = 96397)
Duplication
(N = 91)

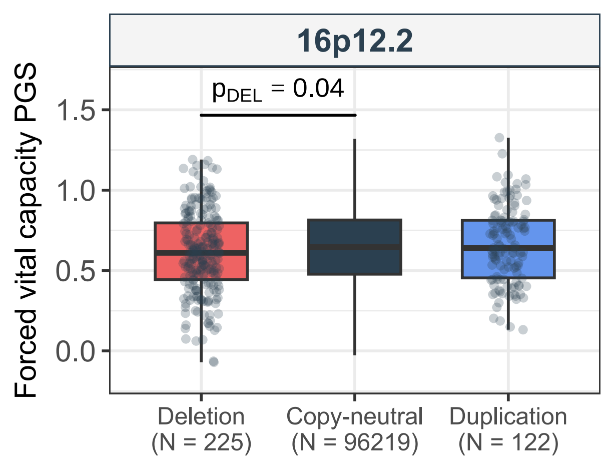
16p12.2
$p_{DEL}$ = 0.04
Forced vital capacity PGS
1.5
1.0
0.5
0.0
Deletion
(N = 225)
Copy-neutral
(N = 96219)
Duplication
(N = 122)

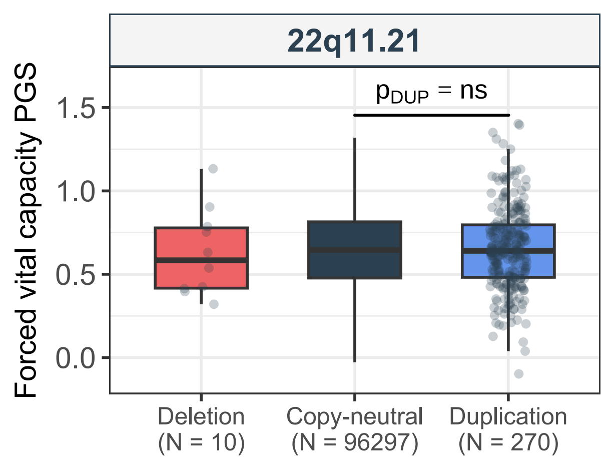

22q11.21
p_DUP = ns
Forced vital capacity PGS
1.5
1.0
0.5
0.0
Deletion
(N = 10)
Copy-neutral
(N = 96297)
Duplication
(N = 270)

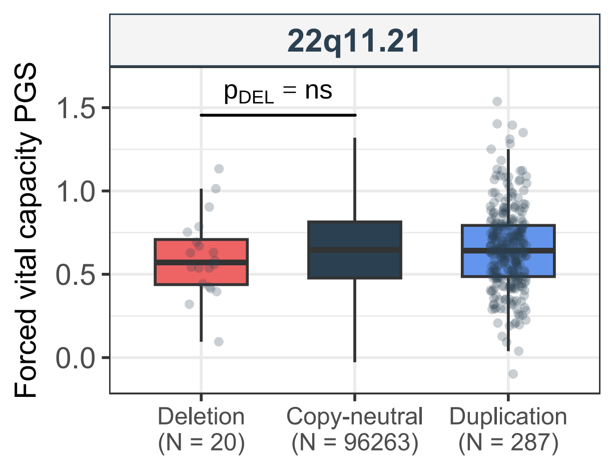
22q11.21
$p_{DEL}$ = ns
Forced vital capacity PGS
1.5
1.0
0.5
0.0
Deletion
(N = 20)
Copy-neutral
(N = 96263)
Duplication
(N = 287)

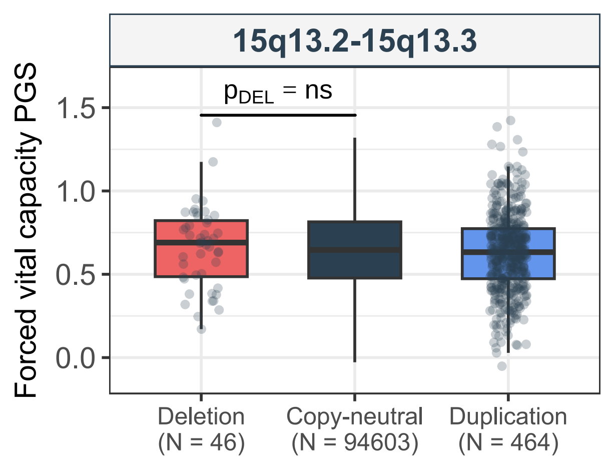

15q13.2-15q13.3
$p_{DEL}$ = ns
Forced vital capacity PGS
1.5
1.0
0.5
0.0
Deletion
(N = 46)
Copy-neutral
(N = 94603)
Duplication
(N = 464)

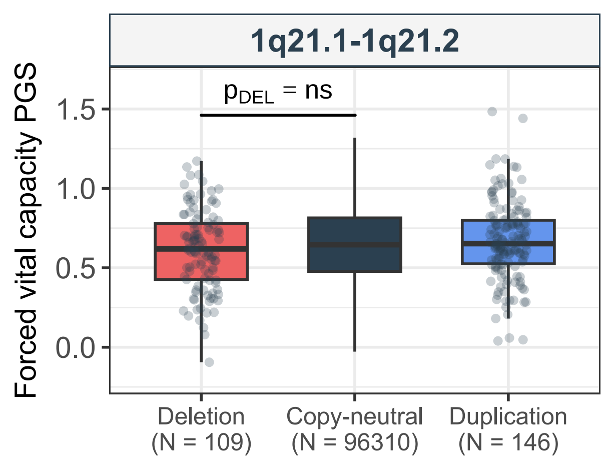
1q21.1-1q21.2
$p_{DEL}$ = ns
Forced vital capacity PGS
1.5
1.0
0.5
0.0
Deletion
(N = 109)
Copy-neutral
(N = 96310)
Duplication
(N = 146)

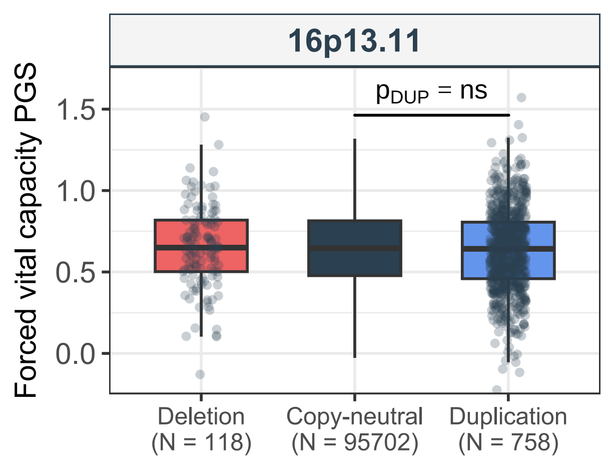
16p13.11
Forced vital capacity PGS
$p_{DUP}$ = ns
1.5
1.0
0.5
0.0
Deletion
(N = 118)
Copy-neutral
(N = 95702)
Duplication
(N = 758)

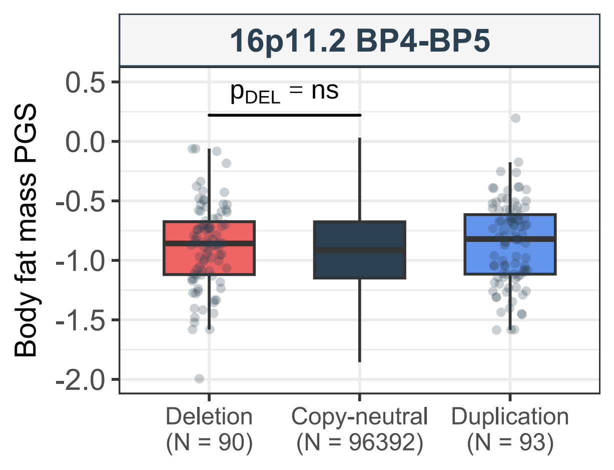

16p11.2 BP4-BP5
$p_{DEL}$ = ns
Body fat mass PGS
0.5
0.0
-0.5
-1.0
-1.5
-2.0
Deletion (N = 90)
Copy-neutral (N = 96392)
Duplication (N = 93)

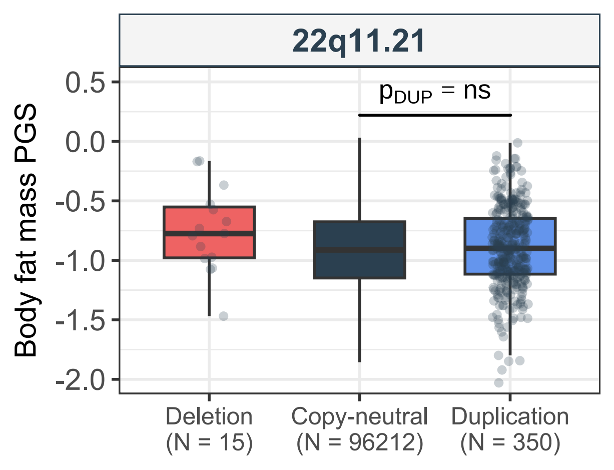
22q11.21
$p_{DUP}$ = ns
Body fat mass PGS
0.5
0.0
-0.5
-1.0
-1.5
-2.0
Deletion
(N = 15)
Copy-neutral
(N = 96212)
Duplication
(N = 350)

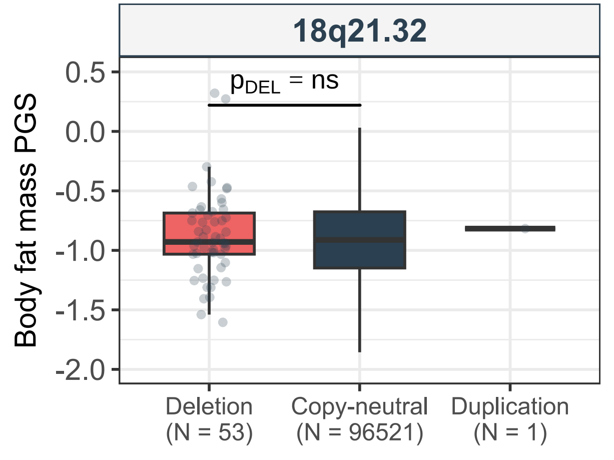

18q21.32
p_DEL = ns
Body fat mass PGS
0.5
0.0
-0.5
-1.0
-1.5
-2.0
Deletion (N = 53)
Copy-neutral (N = 96521)
Duplication (N = 1)

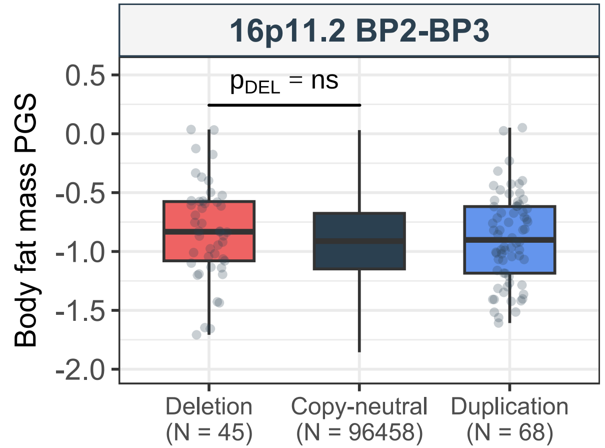
16p11.2 BP2-BP3
$p_{DEL}$ = ns
Body fat mass PGS
0.5
0.0
-0.5
-1.0
-1.5
-2.0
Deletion (N = 45)
Copy-neutral (N = 96458)
Duplication (N = 68)

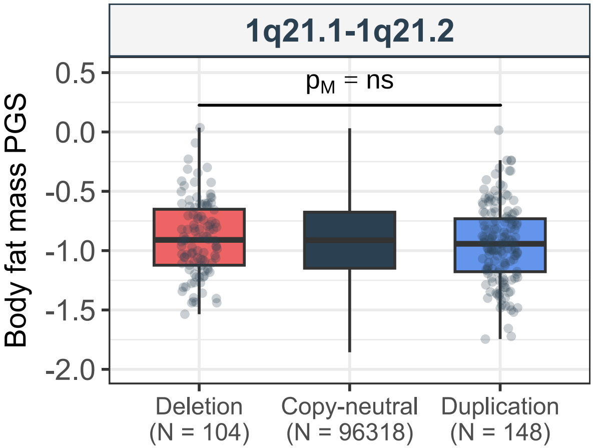

1q21.1-1q21.2
$p_M$ = ns
Body fat mass PGS
0.5
0.0
-0.5
-1.0
-1.5
-2.0
Deletion
(N = 104)
Copy-neutral
(N = 96318)
Duplication
(N = 148)

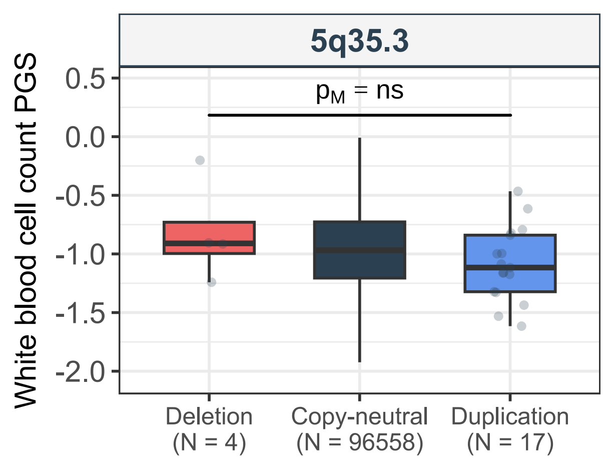

5q35.3
$p_M$ = ns
White blood cell count PGS
0.5
0.0
-0.5
-1.0
-1.5
-2.0
Deletion
(N = 4)
Copy-neutral
(N = 96558)
Duplication
(N = 17)

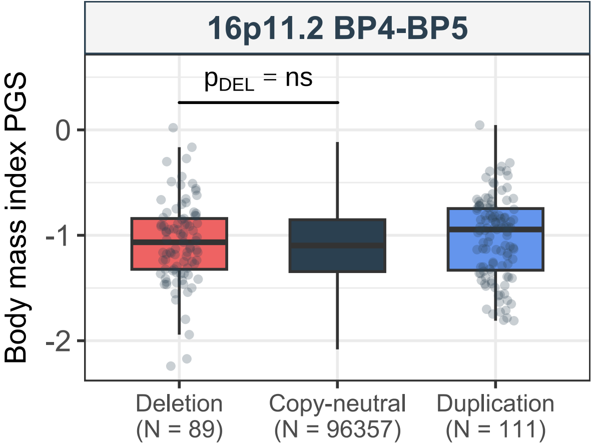

16p11.2 BP4-BP5
$p_{DEL}$ = ns
Body mass index PGS
0
-1
-2
Deletion
(N = 89)
Copy-neutral
(N = 96357)
Duplication
(N = 111)

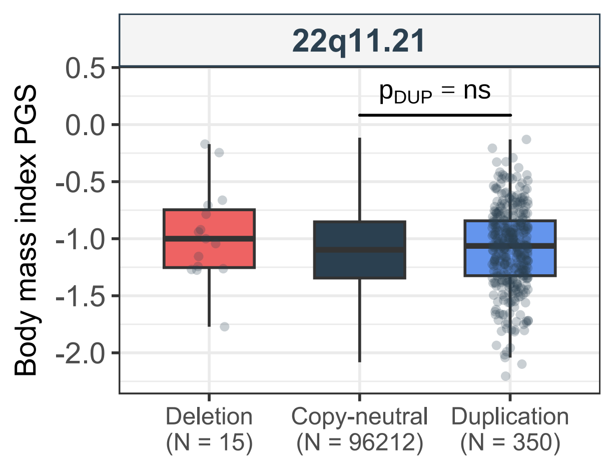

22q11.21
$p_{DUP}$ = ns
Body mass index PGS
0.5
0.0
-0.5
-1.0
-1.5
-2.0
Deletion (N = 15)
Copy-neutral (N = 96212)
Duplication (N = 350)

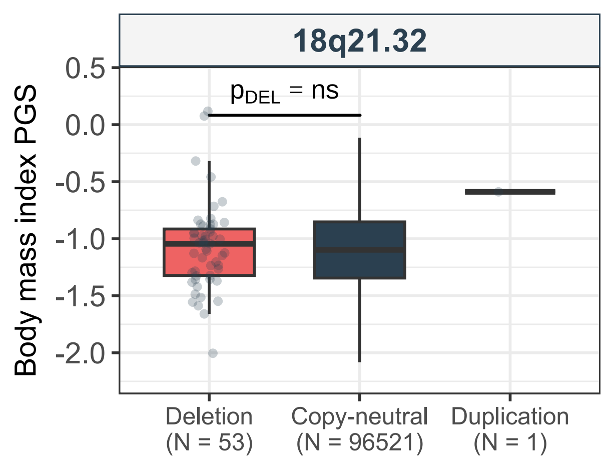
18q21.32
Body mass index PGS
0.5
0.0
-0.5
-1.0
-1.5
-2.0
p_DEL = ns
Deletion
(N = 53)
Copy-neutral
(N = 96521)
Duplication
(N = 1)

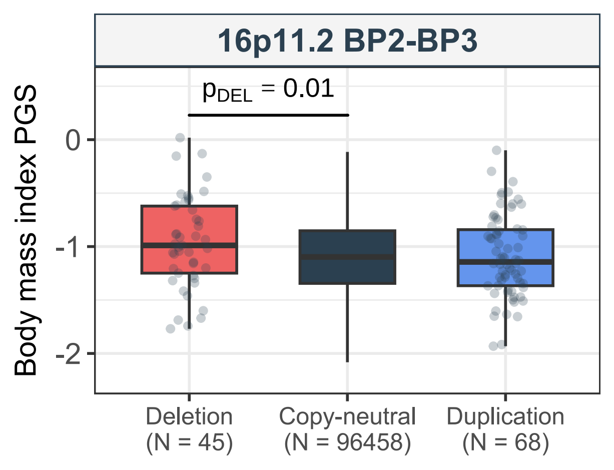

16p11.2 BP2-BP3
$p_{DEL}$ = 0.01
Body mass index PGS
0
-1
-2
Deletion
(N = 45)
Copy-neutral
(N = 96458)
Duplication
(N = 68)

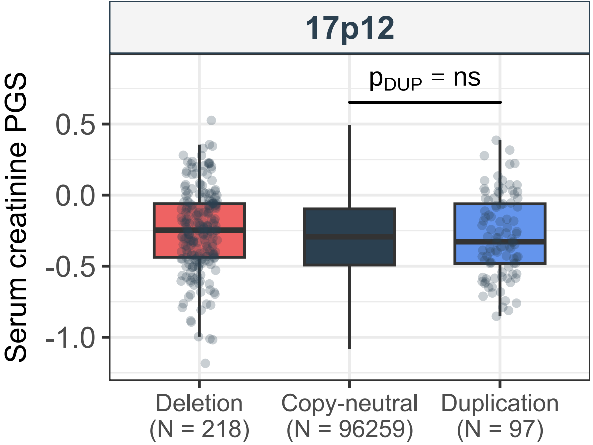
17p12
$p_{DUP}$ = ns
Serum creatinine PGS
0.5
0.0
-0.5
-1.0
Deletion
(N = 218)
Copy-neutral
(N = 96259)
Duplication
(N = 97)

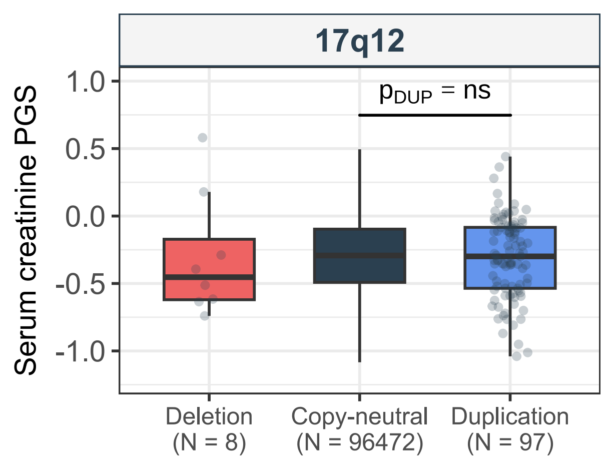

17q12
Serum creatinine PGS
$p_{DUP}$ = ns
1.0
0.5
0.0
-0.5
-1.0
Deletion
(N = 8)
Copy-neutral
(N = 96472)
Duplication
(N = 97)

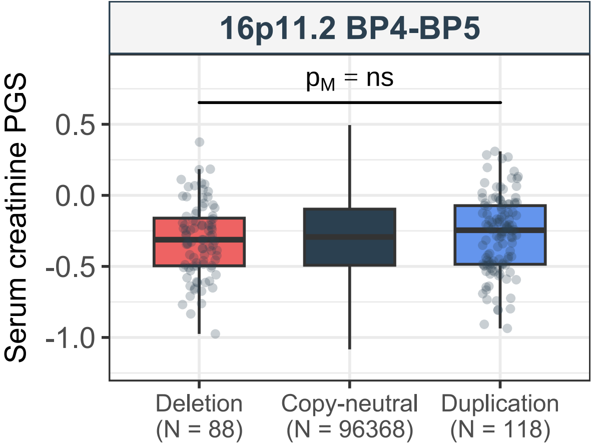
16p11.2 BP4-BP5
p_M = ns
Serum creatinine PGS
0.5
0.0
-0.5
-1.0
Deletion
(N = 88)
Copy-neutral
(N = 96368)
Duplication
(N = 118)

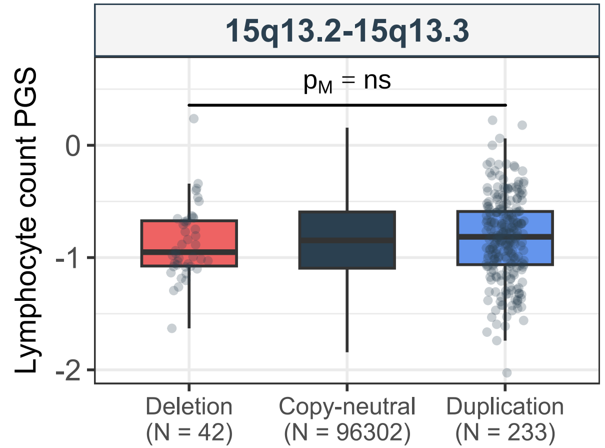
15q13.2-15q13.3
$p_M$ = ns
Lymphocyte count PGS
0
-1
-2
Deletion
(N = 42)
Copy-neutral
(N = 96302)
Duplication
(N = 233)

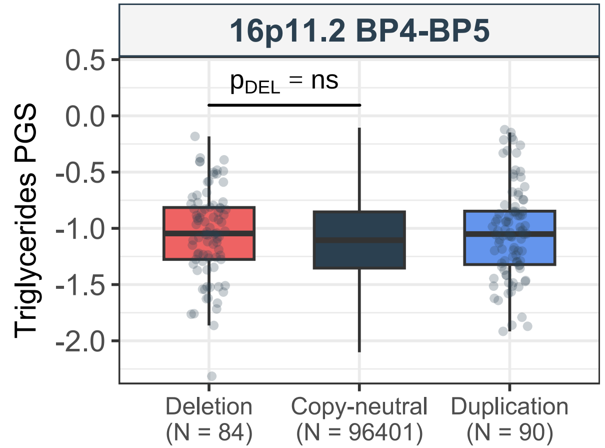
16p11.2 BP4-BP5
$p_{DEL}$ = ns
Triglycerides PGS
0.5
0.0
-0.5
-1.0
-1.5
-2.0
Deletion
(N = 84)
Copy-neutral
(N = 96401)
Duplication
(N = 90)

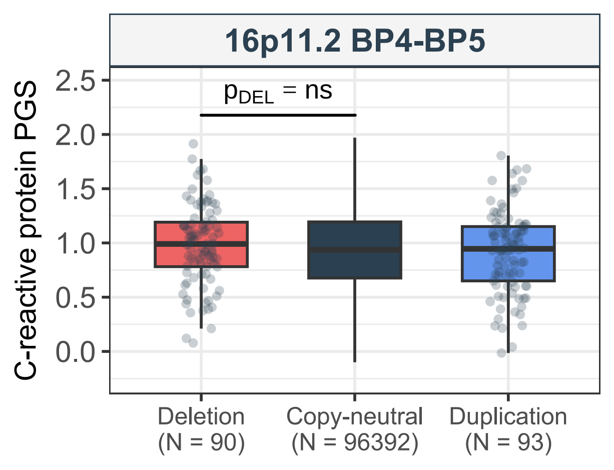
16p11.2 BP4-BP5
C-reactive protein PGS
p_DEL = ns
2.5
2.0
1.5
1.0
0.5
0.0
Deletion
(N = 90)
Copy-neutral
(N = 96392)
Duplication
(N = 93)

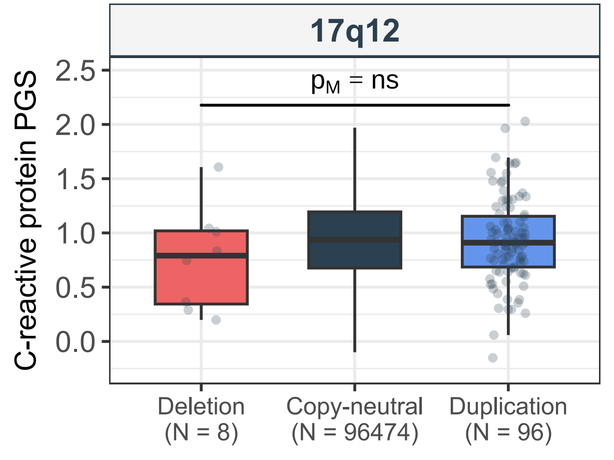

17q12
$p_M$ = ns
C-reactive protein PGS
2.5
2.0
1.5
1.0
0.5
0.0
Deletion
(N = 8)
Copy-neutral
(N = 96474)
Duplication
(N = 96)

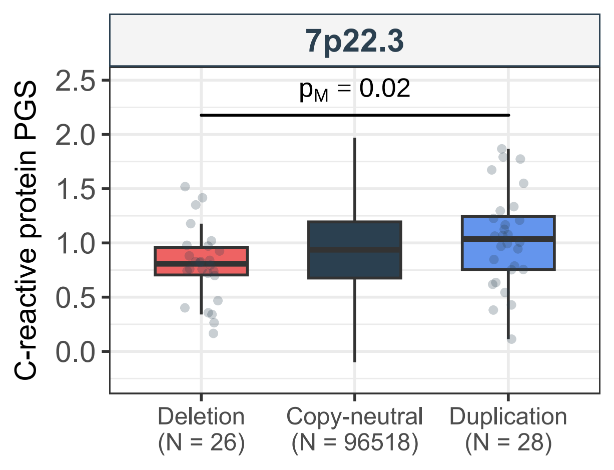

7p22.3
$p_M$ = 0.02
C-reactive protein PGS
2.5
2.0
1.5
1.0
0.5
0.0
Deletion
(N = 26)
Copy-neutral
(N = 96518)
Duplication
(N = 28)

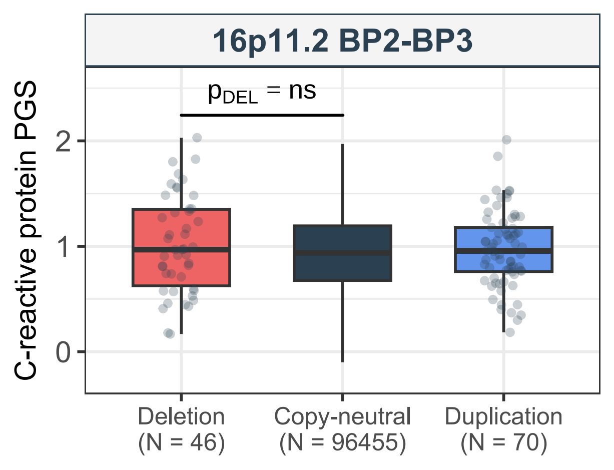
16p11.2 BP2-BP3
$p_{DEL}$ = ns
C-reactive protein PGS
2
1
0
Deletion
(N = 46)
Copy-neutral
(N = 96455)
Duplication
(N = 70)

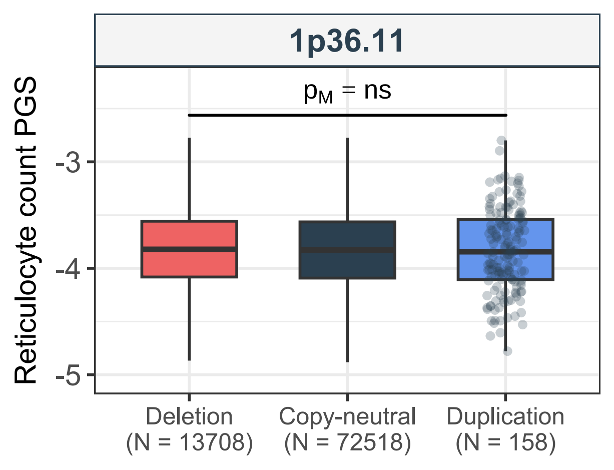
1p36.11
$p_M$ = ns
Reticulocyte count PGS
-3
-4
-5
Deletion
(N = 13708)
Copy-neutral
(N = 72518)
Duplication
(N = 158)

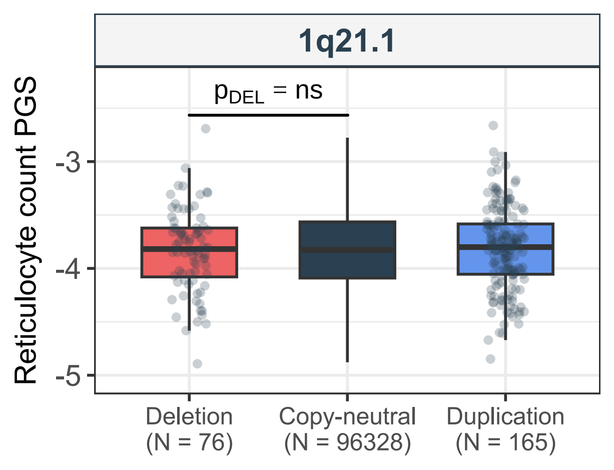
1q21.1
$p_{DEL}$ = ns
Reticulocyte count PGS
-3
-4
-5
Deletion
(N = 76)
Copy-neutral
(N = 96328)
Duplication
(N = 165)

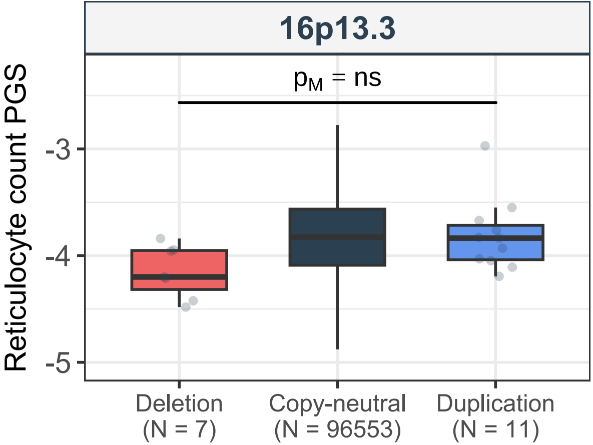
16p13.3
$p_M$ = ns
Reticulocyte count PGS
-3
-4
-5
Deletion
(N = 7)
Copy-neutral
(N = 96553)
Duplication
(N = 11)

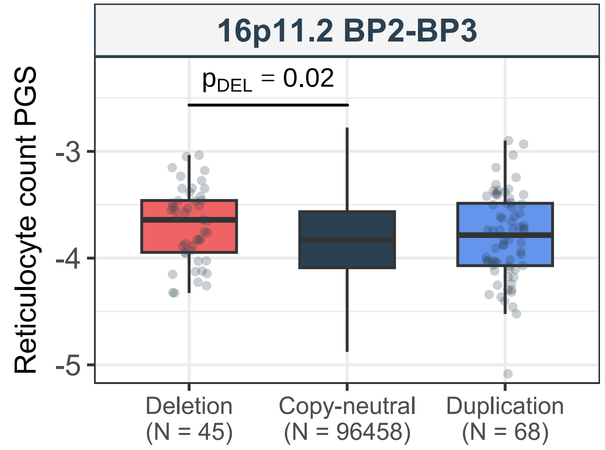

16p11.2 BP2-BP3
$p_{DEL}$ = 0.02
Reticulocyte count PGS
-3
-4
-5
Deletion
(N = 45)
Copy-neutral
(N = 96458)
Duplication
(N = 68)

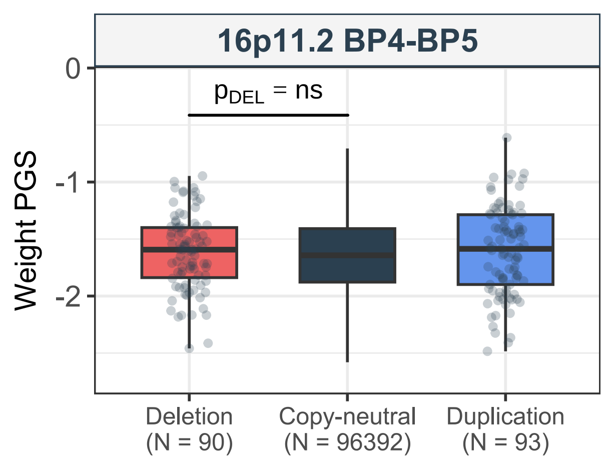
16p11.2 BP4-BP5
$p_{DEL}$ = ns
Weight PGS
0
-1
-2
Deletion
(N = 90)
Copy-neutral
(N = 96392)
Duplication
(N = 93)

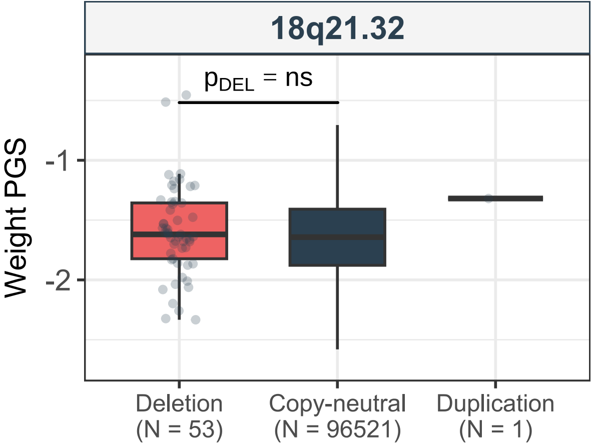
18q21.32
$p_{DEL}$ = ns
Weight PGS
-1
-2
Deletion
(N = 53)
Copy-neutral
(N = 96521)
Duplication
(N = 1)

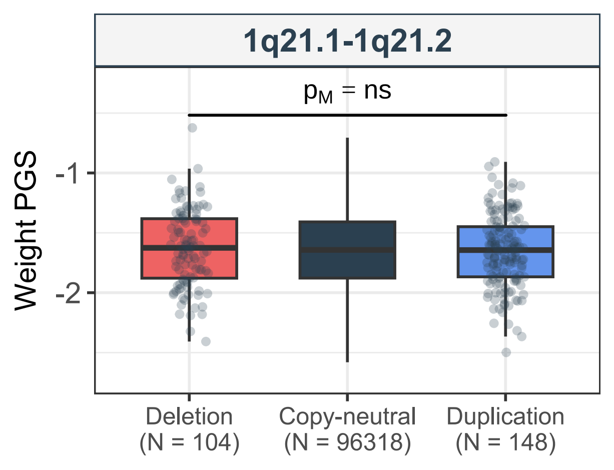

1q21.1-1q21.2
$p_M$ = ns
Weight PGS
-1
-2
Deletion (N = 104)
Copy-neutral (N = 96318)
Duplication (N = 148)

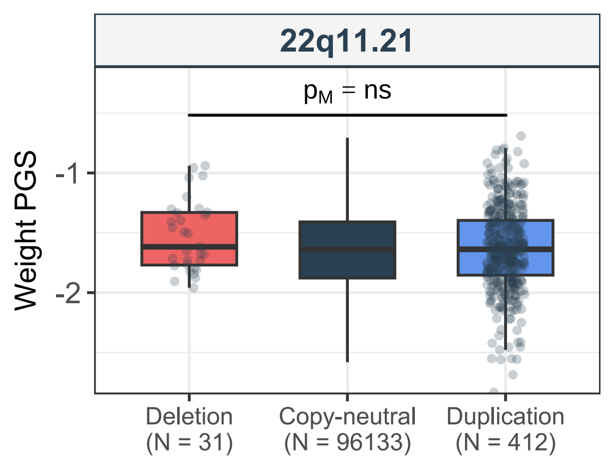
22q11.21
p_M = ns
Weight PGS
-1
-2
Deletion
(N = 31)
Copy-neutral
(N = 96133)
Duplication
(N = 412)

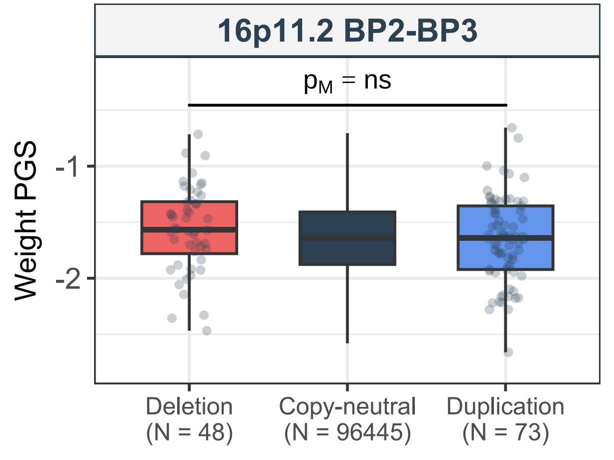

16p11.2 BP2-BP3
$p_M$ = ns
Weight PGS
-1
-2
Deletion
(N = 48)
Copy-neutral
(N = 96445)
Duplication
(N = 73)

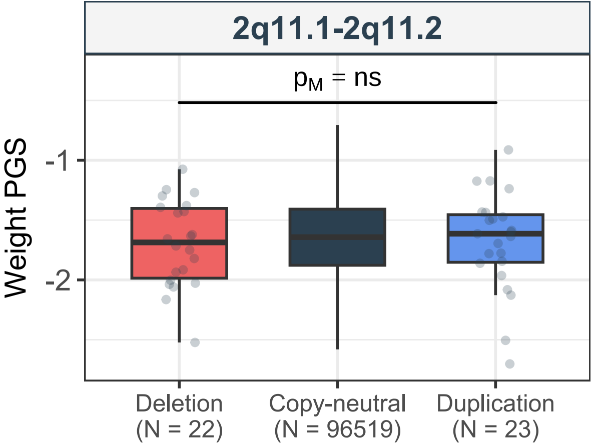
2q11.1-2q11.2
$p_M$ = ns
Weight PGS
-1
-2
Deletion
(N = 22)
Copy-neutral
(N = 96519)
Duplication
(N = 23)

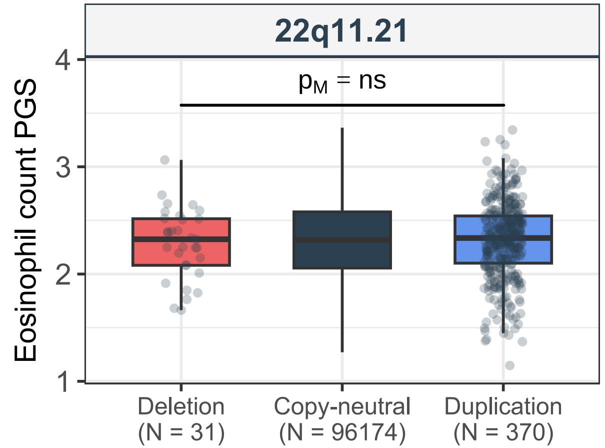

22q11.21
$p_M$ = ns
Eosinophil count PGS
4
3
2
1
Deletion
(N = 31)
Copy-neutral
(N = 96174)
Duplication
(N = 370)

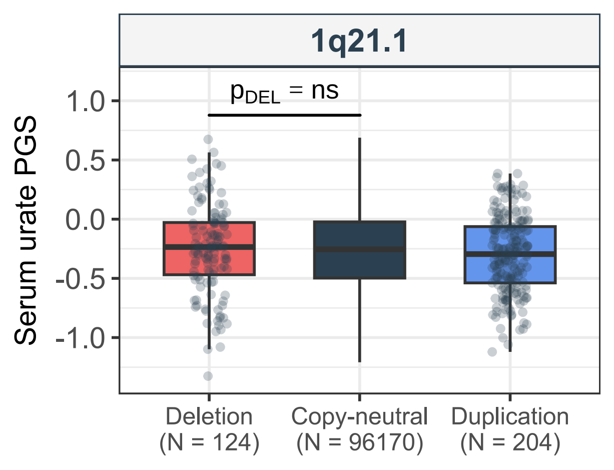
1q21.1
$p_{DEL}$ = ns
Serum urate PGS
1.0
0.5
0.0
-0.5
-1.0
Deletion
(N = 124)
Copy-neutral
(N = 96170)
Duplication
(N = 204)

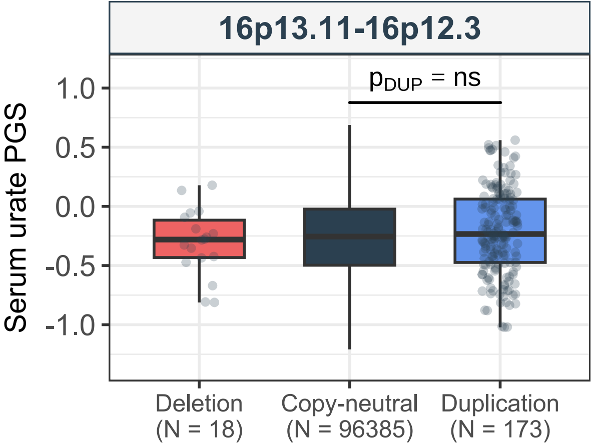

16p13.11-16p12.3
$p_{DUP}$ = ns
Serum urate PGS
1.0
0.5
0.0
-0.5
-1.0
Deletion
(N = 18)
Copy-neutral
(N = 96385)
Duplication
(N = 173)

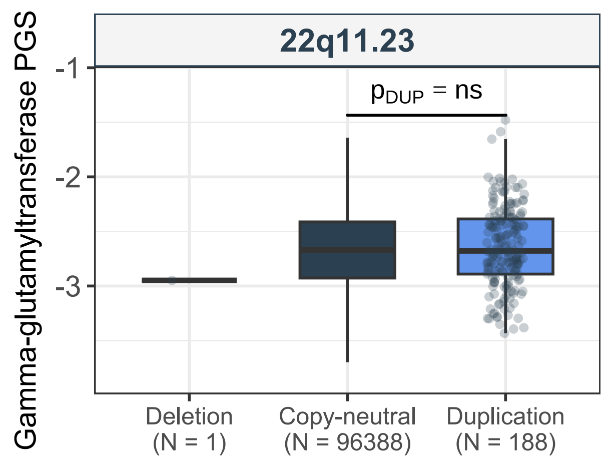
22q11.23
Gamma-glutamyltransferase PGS
$p_{DUP}$ = ns
-1
-2
-3
Deletion
(N = 1)
Copy-neutral
(N = 96388)
Duplication
(N = 188)

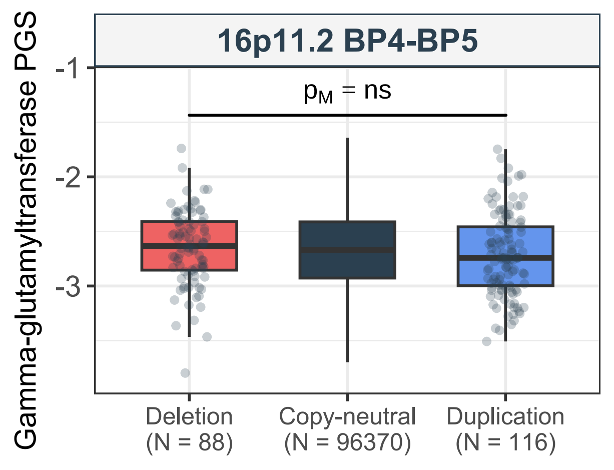
16p11.2 BP4-BP5
Gamma-glutamyltransferase PGS
$p_M$ = ns
-1
-2
-3
Deletion
(N = 88)
Copy-neutral
(N = 96370)
Duplication
(N = 116)

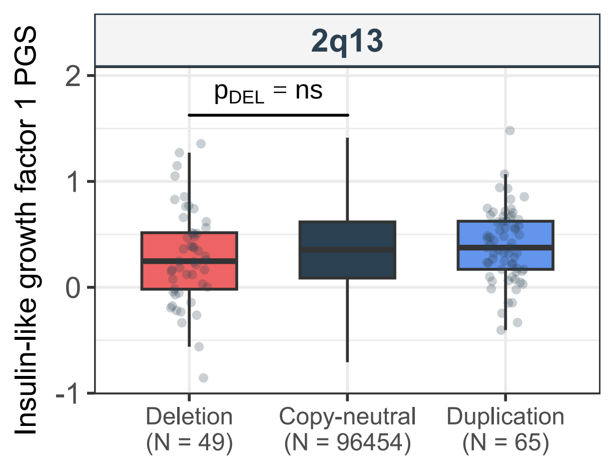

2q13
$p_{DEL}$ = ns
Insulin-like growth factor 1 PGS
2
1
0
-1
Deletion
(N = 49)
Copy-neutral
(N = 96454)
Duplication
(N = 65)

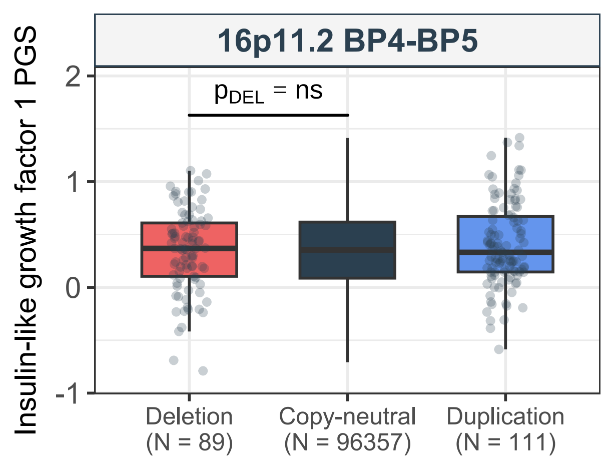
16p11.2 BP4-BP5
Insulin-like growth factor 1 PGS
$p_{DEL}$ = ns
2
1
0
-1
Deletion
(N = 89)
Copy-neutral
(N = 96357)
Duplication
(N = 111)

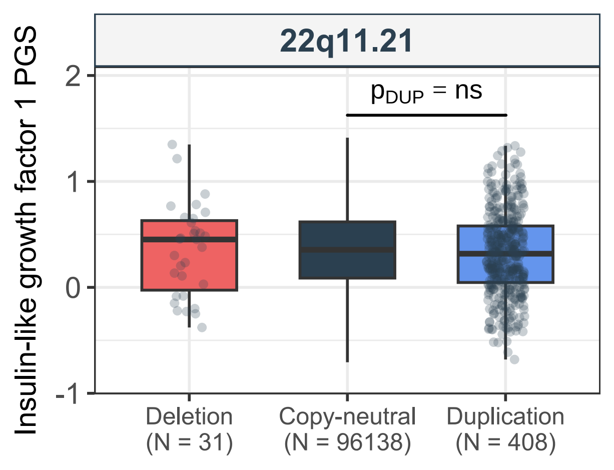
22q11.21
Insulin-like growth factor 1 PGS
$p_{DUP}$ = ns
2
1
0
-1
Deletion (N = 31)
Copy-neutral (N = 96138)
Duplication (N = 408)

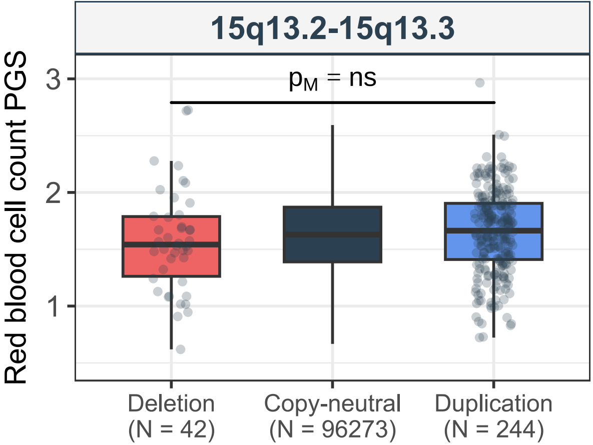

15q13.2-15q13.3
Red blood cell count PGS
$p_M$ = ns
3
2
1
Deletion (N = 42)
Copy-neutral (N = 96273)
Duplication (N = 244)

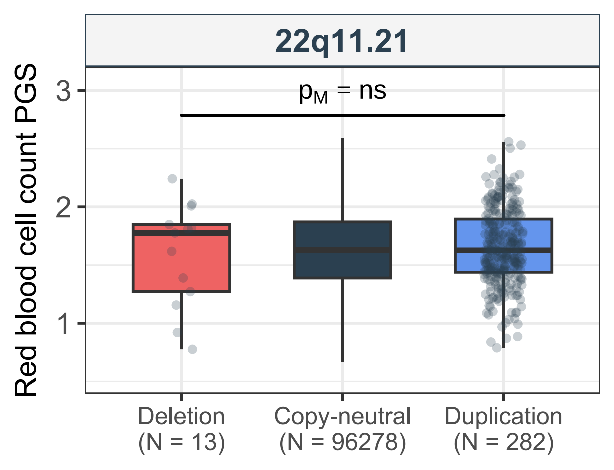

22q11.21
$p_M$ = ns
Red blood cell count PGS
3
2
1
Deletion
(N = 13)
Copy-neutral
(N = 96278)
Duplication
(N = 282)

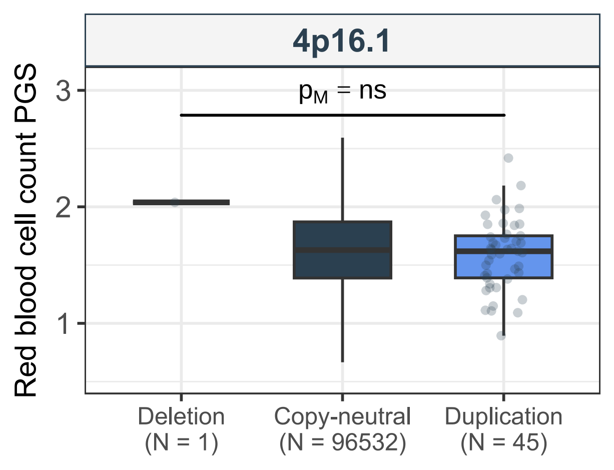
4p16.1
$p_M$ = ns
Red blood cell count PGS
3
2
1
Deletion
(N = 1)
Copy-neutral
(N = 96532)
Duplication
(N = 45)

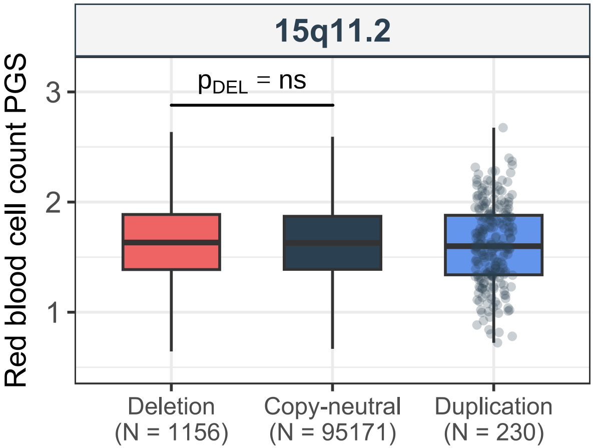
15q11.2
$p_{DEL}$ = ns
Red blood cell count PGS
3
2
1
Deletion
(N = 1156)
Copy-neutral
(N = 95171)
Duplication
(N = 230)

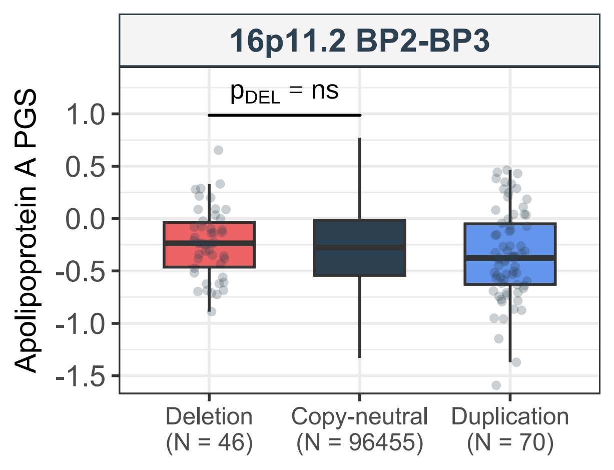
16p11.2 BP2-BP3
p_DEL = ns
Apolipoprotein A PGS
1.0
0.5
0.0
-0.5
-1.0
-1.5
Deletion
(N = 46)
Copy-neutral
(N = 96455)
Duplication
(N = 70)

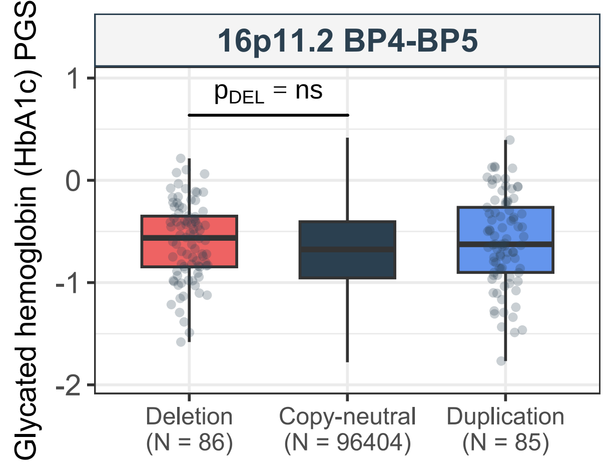
16p11.2 BP4-BP5
$p_{DEL}$ = ns
Glycated hemoglobin (HbA1c) PGS
1
0
-1
-2
Deletion
(N = 86)
Copy-neutral
(N = 96404)
Duplication
(N = 85)

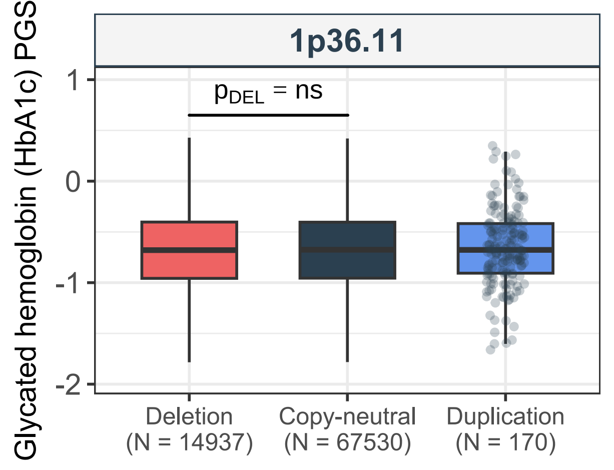
1p36.11
Glycated hemoglobin (HbA1c) PGS
$p_{DEL}$ = ns
1
0
-1
-2
Deletion
(N = 14937)
Copy-neutral
(N = 67530)
Duplication
(N = 170)

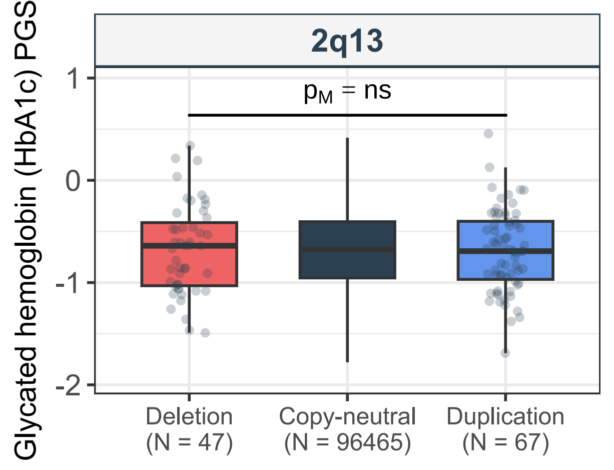
2q13
$p_M$ = ns
Glycated hemoglobin (HbA1c) PGS
1
0
-1
-2
Deletion
(N = 47)
Copy-neutral
(N = 96465)
Duplication
(N = 67)

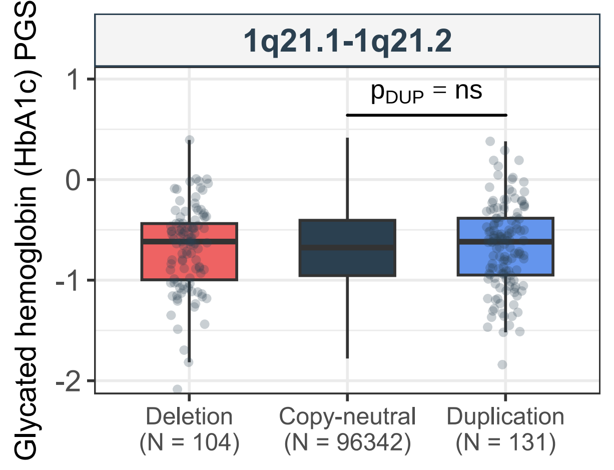

1q21.1-1q21.2
Glycated hemoglobin (HbA1c) PGS
$p_{DUP}$ = ns
1
0
-1
-2
Deletion
(N = 104)
Copy-neutral
(N = 96342)
Duplication
(N = 131)

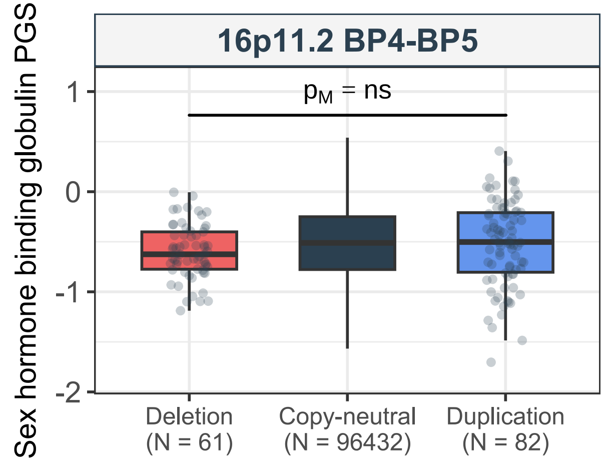

16p11.2 BP4-BP5
$p_M$ = ns
Sex hormone binding globulin PGS
1
0
-1
-2
Deletion
(N = 61)
Copy-neutral
(N = 96432)
Duplication
(N = 82)

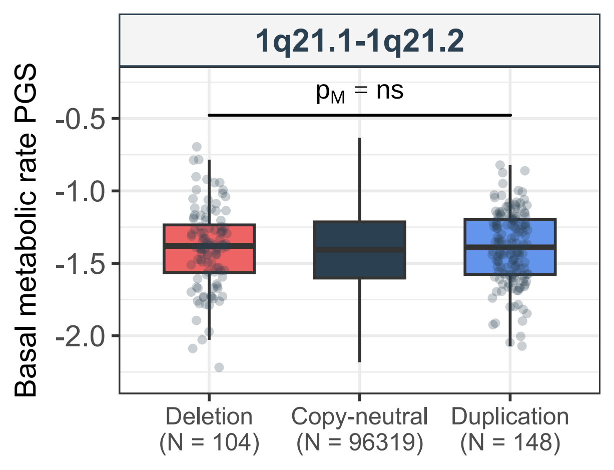

1q21.1-1q21.2
$p_M$ = ns
Basal metabolic rate PGS
-0.5
-1.0
-1.5
-2.0
Deletion
(N = 104)
Copy-neutral
(N = 96319)
Duplication
(N = 148)

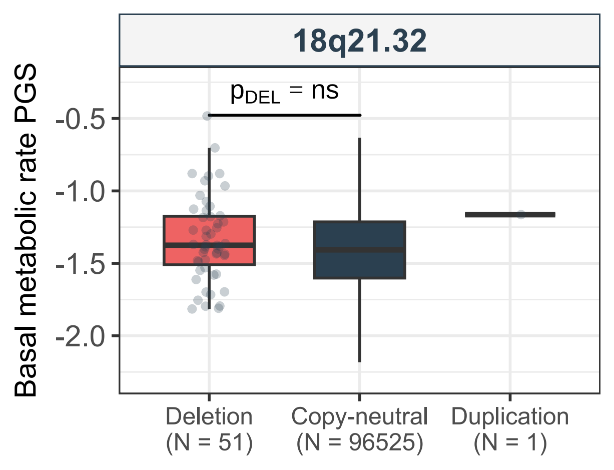
18q21.32
$p_{DEL}$ = ns
Basal metabolic rate PGS
-0.5
-1.0
-1.5
-2.0
Deletion
(N = 51)
Copy-neutral
(N = 96525)
Duplication
(N = 1)

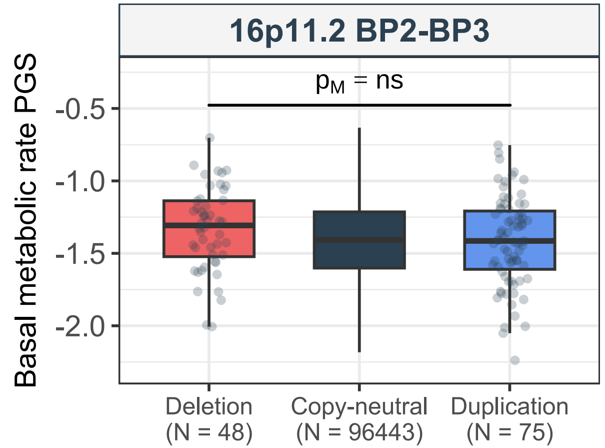

16p11.2 BP2-BP3
$p_M$ = ns
Basal metabolic rate PGS
-0.5
-1.0
-1.5
-2.0
Deletion
(N = 48)
Copy-neutral
(N = 96443)
Duplication
(N = 75)

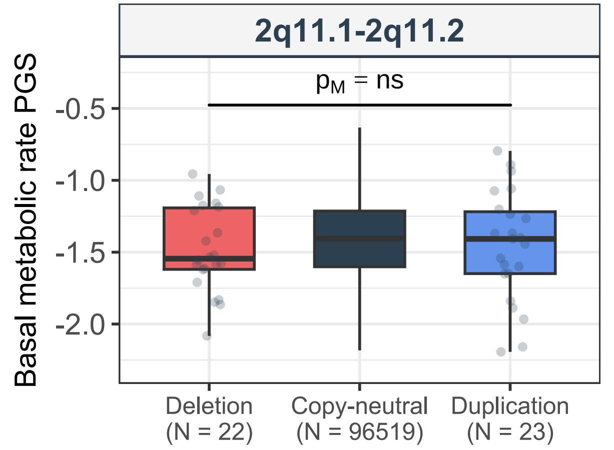
2q11.1-2q11.2
$p_M$ = ns
Basal metabolic rate PGS
-0.5
-1.0
-1.5
-2.0
Deletion
(N = 22)
Copy-neutral
(N = 96519)
Duplication
(N = 23)

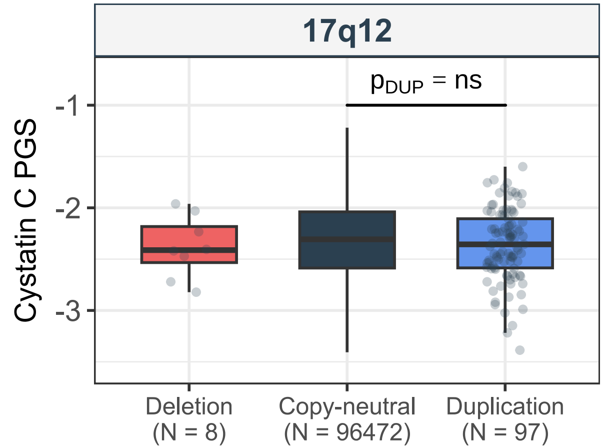

17q12
$p_{DUP}$ = ns
Cystatin C PGS
-1
-2
-3
Deletion
(N = 8)
Copy-neutral
(N = 96472)
Duplication
(N = 97)

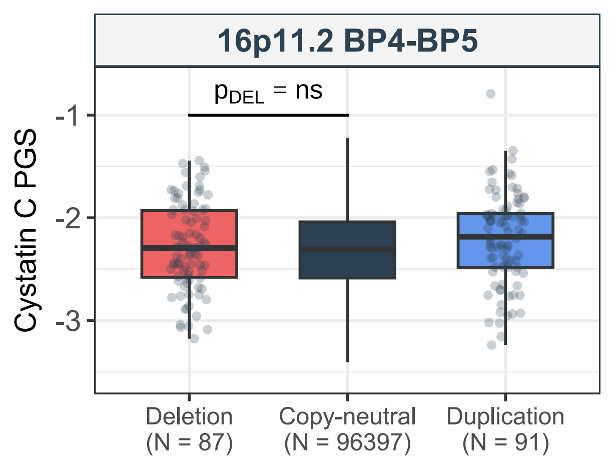
16p11.2 BP4-BP5
$p_{DEL}$ = ns
Cystatin C PGS
-1
-2
-3
Deletion
(N = 87)
Copy-neutral
(N = 96397)
Duplication
(N = 91)

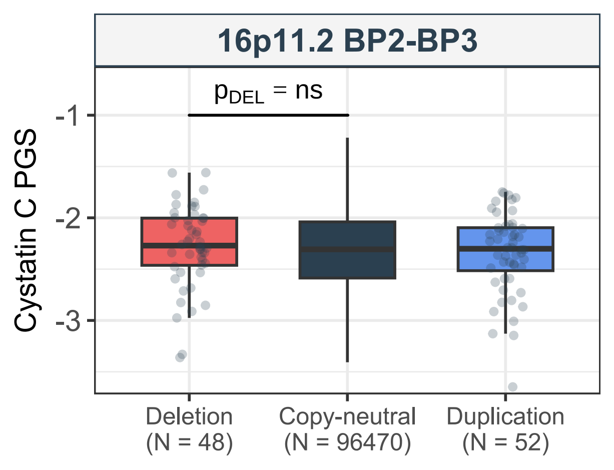
16p11.2 BP2-BP3
$p_{DEL}$ = ns
Cystatin C PGS
-1
-2
-3
Deletion
(N = 48)
Copy-neutral
(N = 96470)
Duplication
(N = 52)

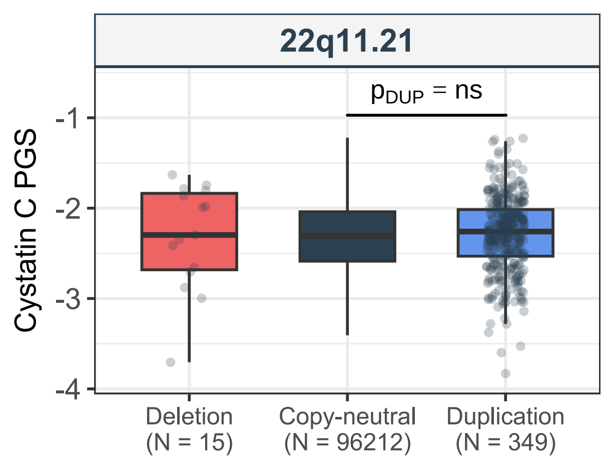
22q11.21
$p_{DUP}$ = ns
Cystatin C PGS
-1
-2
-3
-4
Deletion
(N = 15)
Copy-neutral
(N = 96212)
Duplication
(N = 349)

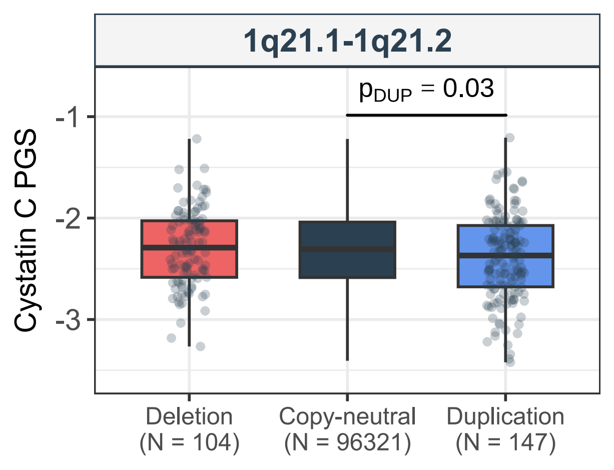
1q21.1-1q21.2
p_DUP = 0.03
Cystatin C PGS
-1
-2
-3
Deletion
(N = 104)
Copy-neutral
(N = 96321)
Duplication
(N = 147)

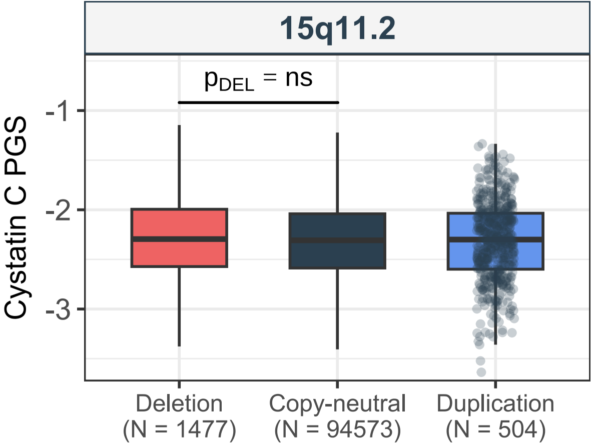

15q11.2
$p_{DEL}$ = ns
Cystatin C PGS
-1
-2
-3
Deletion
(N = 1477)
Copy-neutral
(N = 94573)
Duplication
(N = 504)

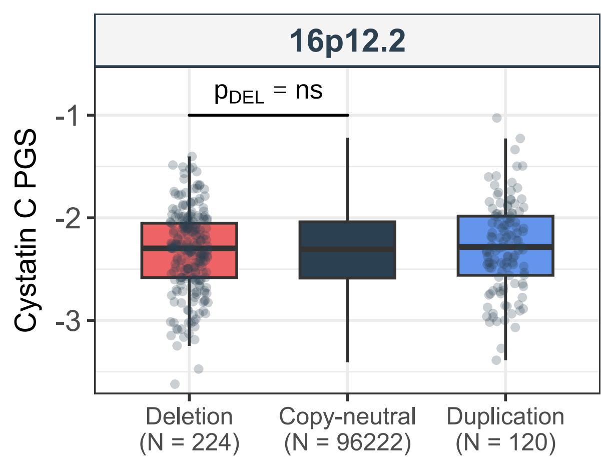
16p12.2
$p_{DEL}$ = ns
Cystatin C PGS
-1
-2
-3
Deletion
(N = 224)
Copy-neutral
(N = 96222)
Duplication
(N = 120)

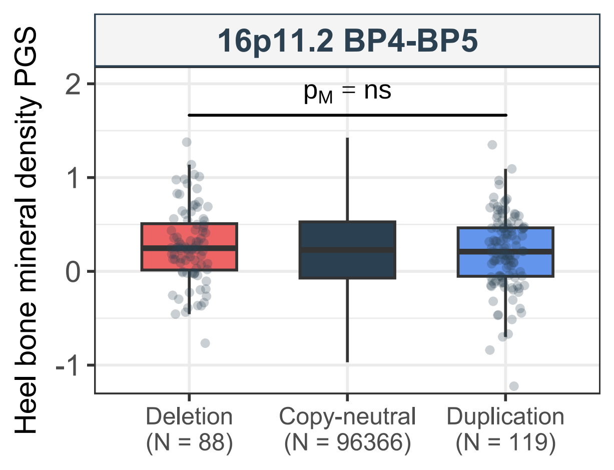

16p11.2 BP4-BP5
$p_M$ = ns
Heel bone mineral density PGS
2
1
0
-1
Deletion
(N = 88)
Copy-neutral
(N = 96366)
Duplication
(N = 119)

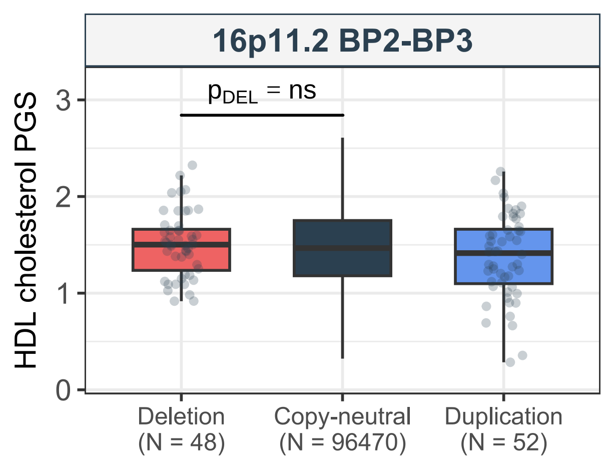
16p11.2 BP2-BP3
$p_{DEL}$ = ns
HDL cholesterol PGS
0
1
2
3
Deletion
(N = 48)
Copy-neutral
(N = 96470)
Duplication
(N = 52)

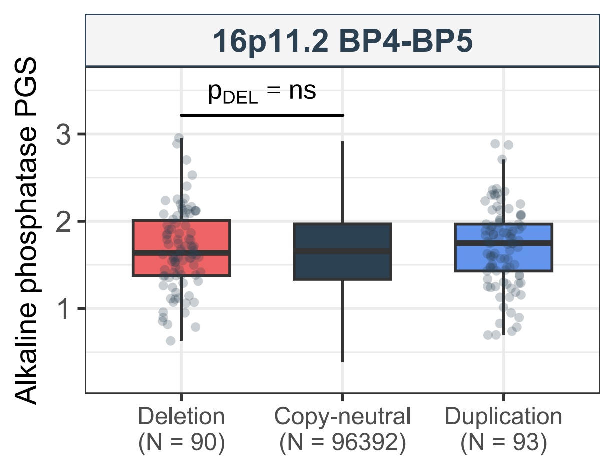

16p11.2 BP4-BP5
$p_{DEL}$ = ns
Alkaline phosphatase PGS
3
2
1
Deletion
(N = 90)
Copy-neutral
(N = 96392)
Duplication
(N = 93)

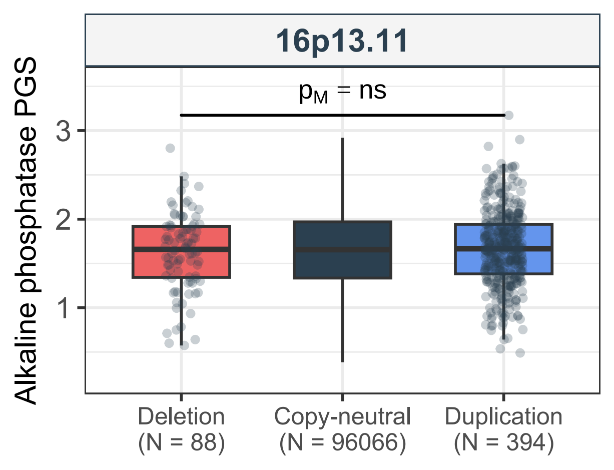
16p13.11
$p_M$ = ns
Alkaline phosphatase PGS
3
2
1
Deletion
(N = 88)
Copy-neutral
(N = 96066)
Duplication
(N = 394)

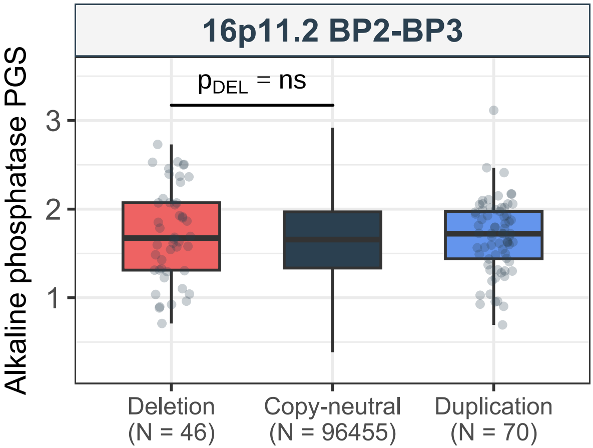
16p11.2 BP2-BP3
$p_{DEL}$ = ns
Alkaline phosphatase PGS
3
2
1
Deletion
(N = 46)
Copy-neutral
(N = 96455)
Duplication
(N = 70)

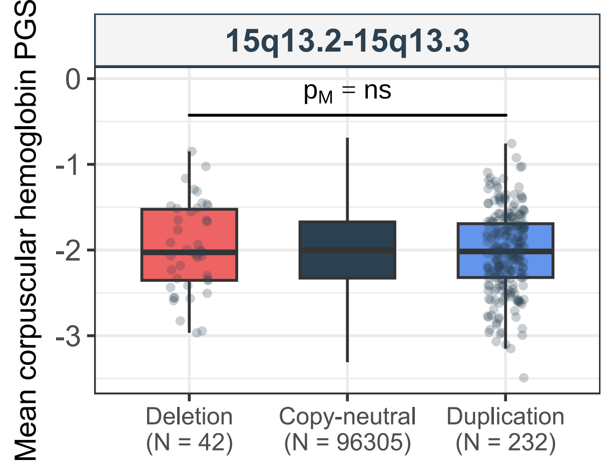
15q13.2-15q13.3
$p_M$ = ns
Mean corpuscular hemoglobin PGS
0
-1
-2
-3
Deletion
(N = 42)
Copy-neutral
(N = 96305)
Duplication
(N = 232)

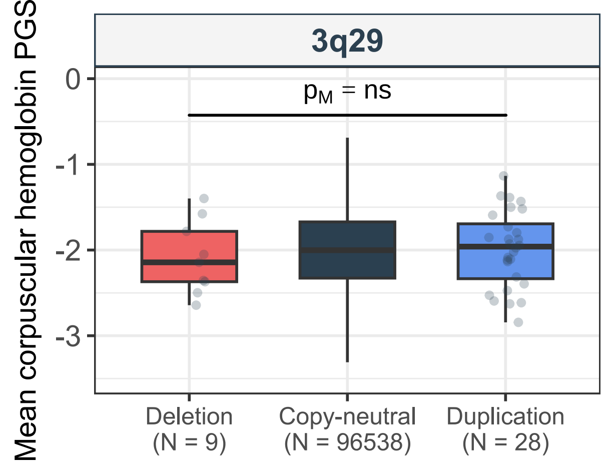
3q29
$p_M$ = ns
Mean corpuscular hemoglobin PGS
0
-1
-2
-3
Deletion
(N = 9)
Copy-neutral
(N = 96538)
Duplication
(N = 28)

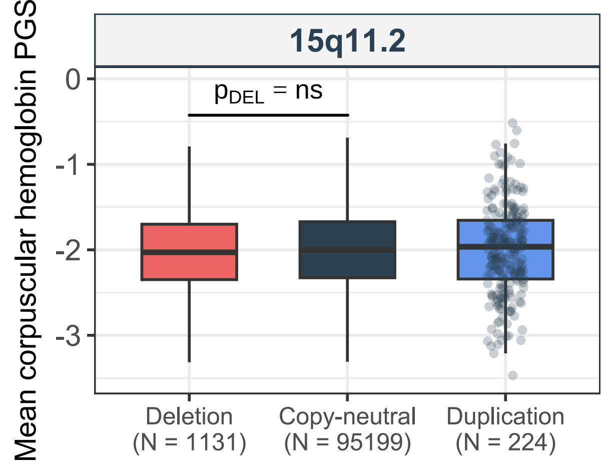
15q11.2
Mean corpuscular hemoglobin PGS
$p_{DEL}$ = ns
0
-1
-2
-3
Deletion
(N = 1131)
Copy-neutral
(N = 95199)
Duplication
(N = 224)

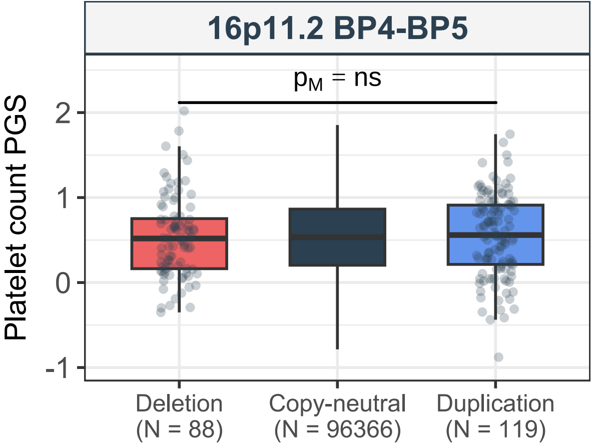

16p11.2 BP4-BP5
$p_M$ = ns
Platelet count PGS
2
1
0
-1
Deletion
(N = 88)
Copy-neutral
(N = 96366)
Duplication
(N = 119)

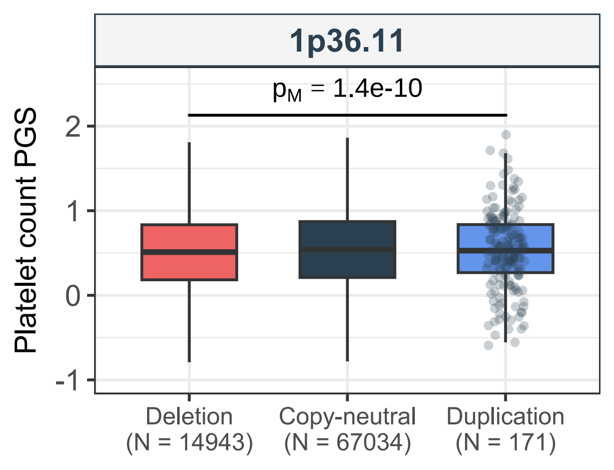

1p36.11
$p_M$ = 1.4e-10
Platelet count PGS
2
1
0
-1
Deletion
(N = 14943)
Copy-neutral
(N = 67034)
Duplication
(N = 171)

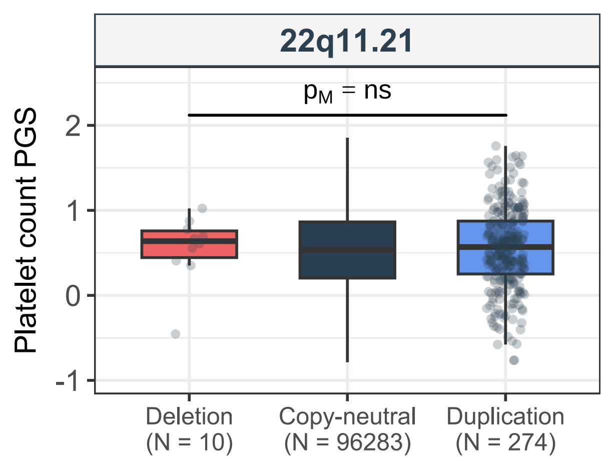
22q11.21
$p_M$ = ns
Platelet count PGS
2
1
0
-1
Deletion
(N = 10)
Copy-neutral
(N = 96283)
Duplication
(N = 274)

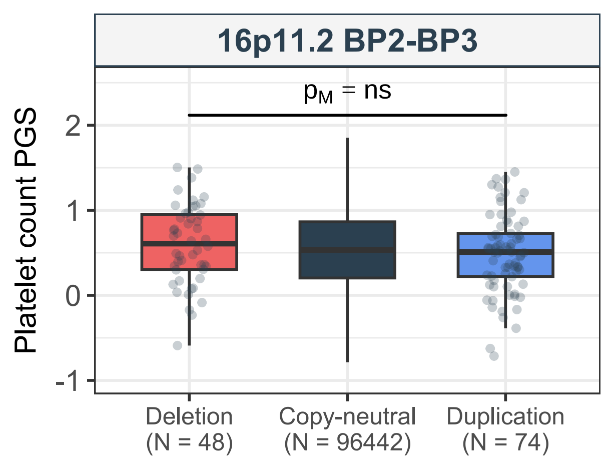

16p11.2 BP2-BP3
p_M = ns
Platelet count PGS
2
1
0
-1
Deletion
(N = 48)
Copy-neutral
(N = 96442)
Duplication
(N = 74)

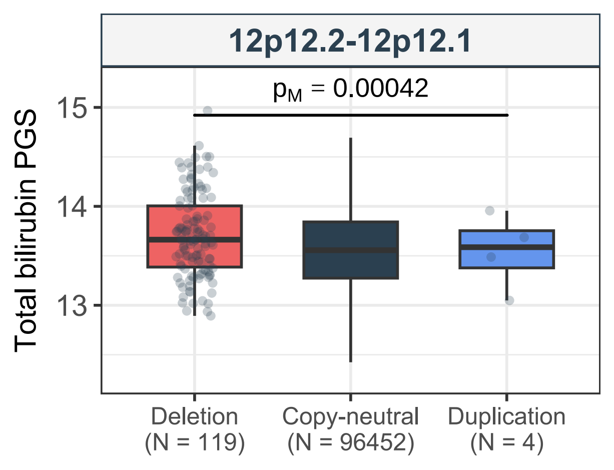
12p12.2-12p12.1
$p_M$ = 0.00042
Total bilirubin PGS
15
14
13
Deletion
(N = 119)
Copy-neutral
(N = 96452)
Duplication
(N = 4)

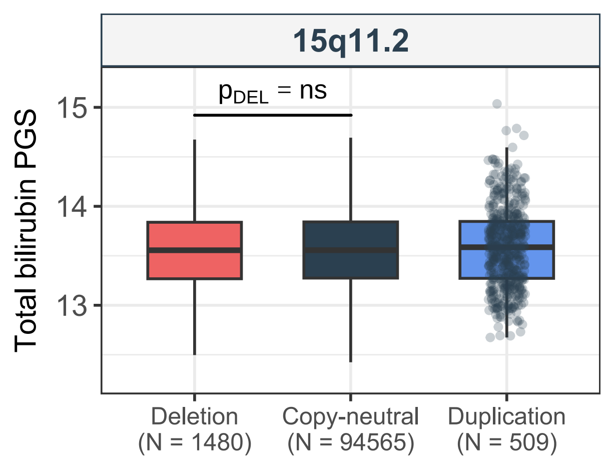
15q11.2
p_DEL = ns
Total bilirubin PGS
15
14
13
Deletion
(N = 1480)
Copy-neutral
(N = 94565)
Duplication
(N = 509)

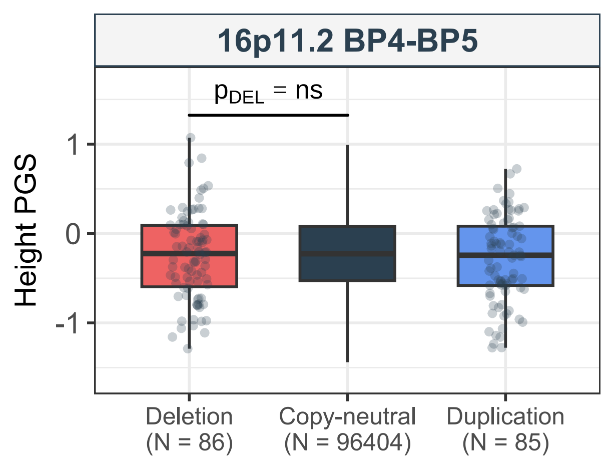

16p11.2 BP4-BP5
$p_{DEL}$ = ns
Height PGS
1
0
-1
Deletion
(N = 86)
Copy-neutral
(N = 96404)
Duplication
(N = 85)

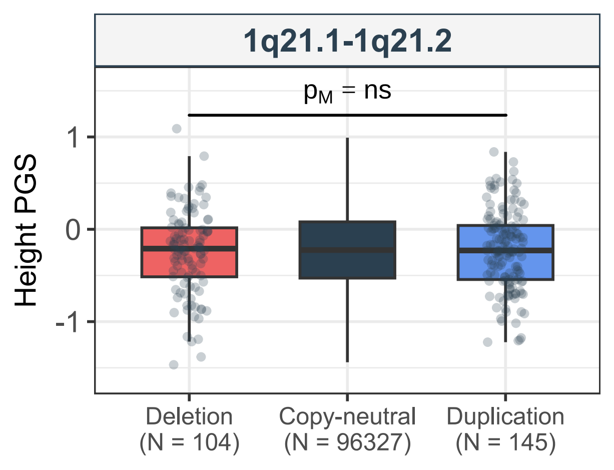
1q21.1-1q21.2
$p_M$ = ns
Height PGS
1
0
-1
Deletion
(N = 104)
Copy-neutral
(N = 96327)
Duplication
(N = 145)

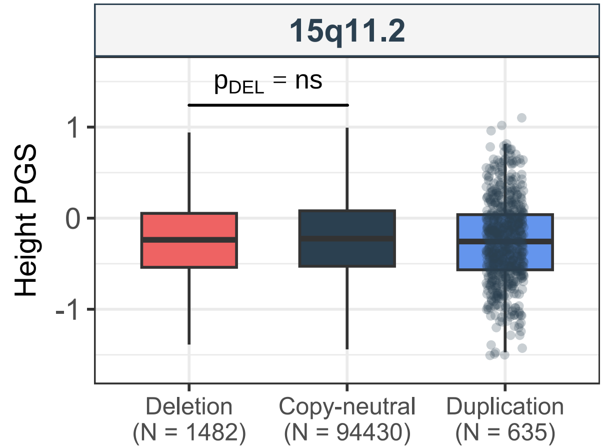
15q11.2
$p_{DEL}$ = ns
Height PGS
1
0
-1
Deletion
(N = 1482)
Copy-neutral
(N = 94430)
Duplication
(N = 635)

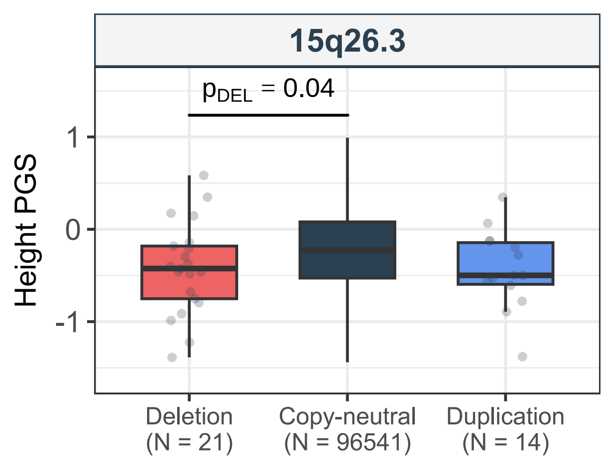
15q26.3
$p_{DEL}$ = 0.04
Height PGS
1
0
-1
Deletion
(N = 21)
Copy-neutral
(N = 96541)
Duplication
(N = 14)

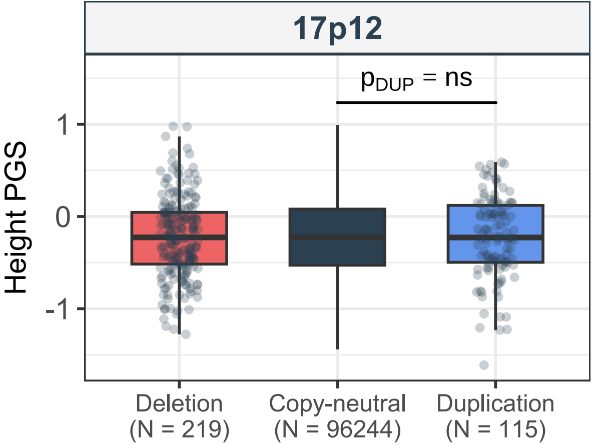

17p12
$p_{DUP}$ = ns
Height PGS
1
0
-1
Deletion
(N = 219)
Copy-neutral
(N = 96244)
Duplication
(N = 115)

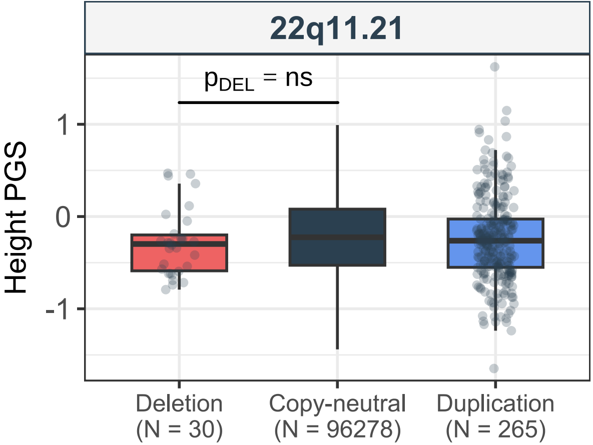

22q11.21
$p_{DEL}$ = ns
Height PGS
1
0
-1
Deletion
(N = 30)
Copy-neutral
(N = 96278)
Duplication
(N = 265)

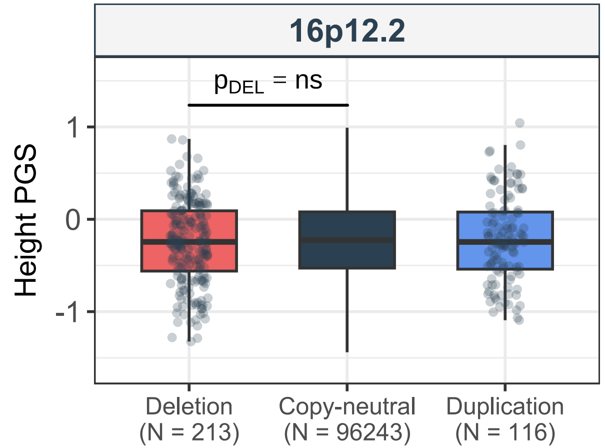
16p12.2
$p_{DEL}$ = ns
Height PGS
1
0
-1
Deletion
(N = 213)
Copy-neutral
(N = 96243)
Duplication
(N = 116)
